# Genetic Risk, Health-Associated Lifestyle, and Risk of Early-onset Total Cancer and Breast Cancer

**DOI:** 10.1101/2024.04.04.24305361

**Authors:** Yin Zhang, Sara Lindström, Peter Kraft, Yuxi Liu

**Author notes:** Corresponding Author: Yuxi Liu, PhD, MS, Yin Zhang, MD, MPH.

## Abstract

**Importance:** Early-onset cancer (diagnosed under 50 years of age) is associated with aggressive disease characteristics and its rising incidence is a global concern. The association between healthy lifestyle and early-onset cancer and whether it varies by common genetic variants is unknown.

**Objective:** To examine the associations between genetic risk, lifestyle, and risk of early-onset cancers.

**Design, Setting, and Participants:** We analyzed a prospective cohort of 66,308 white British participants who were under age 50 and free of cancer at baseline in the UK Biobank.

**Exposures:** Sex-specific composite total cancer polygenic risk scores (PRSs), a breast cancer-specific PRS, and sex-specific health-associated lifestyle scores (HLSs, which summarize smoking status, body mass index [males only], physical activity, alcohol consumption, and diet).

**Main Outcomes and Measures:** Hazard ratios (HRs) and 95% confidence intervals (CIs) for early-onset total and breast cancer.

**Results:** A total of 1,247 incident invasive early-onset cancer cases (female: 820, male: 427, breast: 386) were documented. In multivariable-adjusted analyses with 2-year latency, higher genetic risk (highest vs. lowest tertile of PRS) was associated with significantly increased risks of early-onset total cancer in females (HR, 95% CI: 1.85, 1.50-2.29) and males (1.94, 1.45-2.59) as well as early-onset breast cancer in females (3.06, 2.20-4.25). An unfavorable lifestyle (highest vs. lowest category of HLS) was associated with higher risk of total cancer and breast cancer in females across genetic risk categories; the association with total cancer was stronger in the highest genetic risk category than the lowest: HRs in females and men were 1.85 (1.02, 3.36), 3.27 (0.78, 13.72) in the highest genetic risk category and 1.15 (0.44, 2.98), 1.16 (0.39, 3.40) in the lowest.

**Conclusions and Relevance:** Both genetic and lifestyle factors were independently associated with early-onset total and breast cancer risk. Compared to those with low genetic risk, individuals with a high genetic risk may benefit more from adopting a healthy lifestyle in preventing early-onset cancer.

## Introduction

Cancers are caused by inherited variants and acquired mutations induced by environmental factors or result from unavoidable DNA replication errors. Modifiable lifestyle factors, such as smoking, physical activity, and diet, and their joint effect have been linked with the incidence of overall and major cancer types.^1,2^ In the United States, data from the Centers for Disease Control and Prevention and the National Cancer Institute suggested that over 40% of all incident cancers and cancer deaths were attributable to unhealthy lifestyle factors.^3^ Evidence from large population-based prospective studies suggest that a substantial number of cancer cases may be preventable through lifestyle modification.^1^ On the other hand, genome-wide association studies (GWAS) have identified thousands of susceptibility loci across major cancer types.^4^ Emerging studies have investigated the degree to which adherence to a healthy lifestyle may attenuate the risk of cancer across strata defined by common genetic variants, aggregated into cancer polygenic risk scores (PRS). These studies have focused on overall cancer^5^ and a few major cancer types.^6–14^

Early-onset cancer, defined as cancer diagnosed under age 50, represents a unique spectrum of malignancies^15,16^ and generally manifests with a more aggressive disease phenotype.^15,16^ The incidence of early-onset cancer has increased globally over the past decades, possibly supporting a basis in changing environmental hazards or interactions between hazardous environments and genetics.^15,17–19^ Mounting evidence has established close epidemiological and biological links between unhealthy lifestyle and early-onset cancer.^20^ In addition, cancer PRS tend to be more strongly associated with early-onset compared to late-onset cancer.^21^ However, no previous study has evaluated the magnitude to which adopting a healthy lifestyle may attenuate the impact of common genetic variants on early-onset cancer risk, highlighting a significant knowledge gap.

To address this important unanswered question, we conducted a large prospective cohort study to investigate the association between genetic risk, health-associated lifestyle, and risk of early-onset total cancer in both sexes, as well as early-onset breast cancer in females (which accounts for almost half of the early-onset cancer cases in our female study population).

### Study Population

The study was performed using data from the UK Biobank longitudinal cohort, the details of which have been described previously.^22–24^ In brief, the UK Biobank began between 2006 and 2010 when more than 500,000 participants aged 40 to 70 years from 22 assessment centers across England, Scotland, and Wales were enrolled. Information on demographics (age, sex, ethnicity), lifestyle and other health-related factors were collected via extensive baseline questionnaires, interviews, and physical measurements. Blood samples were collected at baseline and were used for genotyping. Informed consent was obtained from participants during the baseline assessment.

### Ascertainment of Analytic Population

We restricted the study population to self-reported white British participants. We excluded participants diagnosed with any cancer before or at the cohort baseline, aged over 50 at the cohort baseline, or without genotype data. We further excluded individuals with genetic sex discordance, second (or higher)-degree related individuals (kinship coefficients >0.088), and heterozygosity or call rate outliers based on genotyping data. Individuals who had withdrawn consent to participate, or with missingness in smoking, body mass index (BMI), physical activity, alcohol intake, and diet were further removed, resulting in 66,308 eligible participants (females: 34,383, males: 31,925) for the final analyses.

### Ascertainment of Genetic Risk

Genetic risk for early-onset total cancer was assessed using sex-specific composite PRSs. Briefly, we calculated composite PRSs as weighted sums of a spectrum of published cancer site-specific PRSs for European ancestry populations.^25–34^ These PRSs were selected based on their original training sample size, methods, test-set performance, possibility of overfit, and the availability of SNPs and weights. We used weights obtained from lasso regression by regressing early-onset total cancer on the PRSs of individual cancers (including lifestyle factors and a range of variables selected a priori [detailed in the Statistical Analysis section] as covariates, with a partial penalty on PRSs only).

We constructed sex-specific composite total cancer PRSs due to the difference in cancer spectra between females and males. Specifically in analyses focused on early-onset breast cancer among females, we constructed a breast cancer-specific PRS.^25^ The lists of published cancer site-specific PRSs included in the lasso regression and those ultimately selected for the development of sex-specific composite total cancer PRSs are summarized in eTables 1-2 in Supplement 1. The complete lists of SNPs included in the cancer site-specific PRSs are presented in eTables S1-S23 in Supplement 2. The early-onset cancer spectrum in females and males, showcasing cancers with published site-specific PRSs qualified for inclusion in the lasso regression, are summarized in eTable 3 in the Supplement 1.

PRSs for individual cancer types were calculated as weighted sums of the effect allele dosage for selected SNPs for each individual assuming an additive model based on the formula β_1_ × SNP*_i_*_,1_ + β_2_ × SNP*_i_*_,2_ + … + β*_j_* × SNP*_i_*_,*j*_ + … + β_n_ × SNP*_i_*_,n_, where SNP*_i_*_,*j*_ is the effect allele dosage for SNP *j* for individual *i*, β*_j_* is per-allele log odds ratio for SNP *j* for a specific cancer type, n is the total number of selected SNPs for the specific cancer type. The sex-specific composite total cancer PRSs were constructed based on the formula h_1_ × PRS_𝑖,1_ + h_2_ × PRS_𝑖,2_ + ⋯ + h_𝑘_ × PRS_𝑖,𝑘_, where PRS_𝑖,𝑘_ is the PRS for cancer type *k* for individual *i*, h*_k_* is the weight for cancer type *k* obtained from lasso regression.

### Ascertainment of Health-associated Lifestyle Scores

Sex-specific health-associated lifestyle scores (HLSs) were calculated based on a combination of baseline smoking status, BMI (males only), physical activity, alcohol consumption, and diet.^5–7^ BMI was not included as a component of female-specific HLS due to the widely recognized inverse association of BMI with the risk of early-onset/pre-menopausal breast cancer,^35^ coupled with the fact that early-onset total cancer in our female study population was predominantly driven by early-onset breast cancer. Healthy lifestyles were defined as no current smoking, normal BMI (18.5 to 25 kg/m^2^), adequate physical activity (exercised for at least 75 minutes of vigorous activity per week or 150 minutes of moderate activity per week or an equivalent combination), no alcohol consumption, and healthy diet (consumed an increased amount of fruits, vegetables, whole grains, and a reduced amount of red meats and processed meats).^5–7,36,37^

To streamline the delivery of the public health message in a simple way, participants received a score of 1 if they didn’t meet the criterion for a specific lifestyle factor or 0 otherwise.^5–7^ We then added up the sum across the lifestyle components resulting in a final unweighted HLS ranging from 0-4 (females) or 0-5 (males), with higher scores indicating an unhealthier lifestyle. More details of the sex-specific HLSs can be found in eTable 4 in Supplement 1.

### Ascertainment of Cancer and Death

Incident invasive cancer cases were ascertained via linkage to the National Health Service central registers and death registries in England, Wales, and Scotland. Cases were coded using the International Classification of Diseases (10^th^ Revision). Early-onset cancer cases were defined as those diagnosed under the age of 50 years.^17^ Deaths were ascertained through linkage to death registries.

### Statistical Analysis

The analyses of early-onset total cancer were conducted separately by sex, given the difference in cancer spectra between females and males. The analyses of early-onset breast cancer were restricted to females only. To minimize the potential impact of reverse causation, a latency period was introduced by excluding the first 2 years of follow-up.

Person-years of follow-up was calculated from the baseline until the date of diagnosis of any cancer (except non-melanoma skin cancer), death, loss to follow-up, or 50^th^ birthday, whichever occurred earliest. Descriptive analyses were performed to assess population characteristics across categories of PRS and HLS. Multivariable Cox proportional hazards models with age as the timescale were used to estimate hazard ratios (HRs) and 95% confidence intervals (CIs) of early-onset total cancer and early-onset breast cancer across PRS and HLS groups (PRS: high, intermediate, vs. low, based on tertiles; HLS: unhealthy, intermediate, vs. healthy, based on the predefined cutoffs detailed in eTable 4 in Supplement 1) in females and males, as well as for their joint associations by using the collapsed categories. HRs and 95% CIs per 1-SD increase in PRS and per 1-unit increase in HLS were also estimated. Moreover, stratified analysis was performed to assess the association between HLS and early-onset total cancer and breast cancer risk within each PRS stratum. The proportional hazards assumption was tested using the likelihood ratio test to compare models with and without product terms between exposures and log-transformed age. No violation of the assumption was detected.

Covariates were selected a priori. Multivariate analyses of early-onset total cancer were stratified by sex, and adjusted for the first 10 genetic principal components (continuous), genotyping batch (categorical), average total household income (less than 18,000, 18,000-30,999, 31,000-51,999, 52,000-100,000, greater than 100,000, prefer not to answer or do not know), education (college or university degree, some professional qualifications, secondary education, others, prefer not to answer or do not know), BMI (continuous, kg/m^2^; in analyses of females only), and family history of cancer (yes, no; family history of breast cancer was used in analyses of breast cancer). PRS and HLS were mutually adjusted.

Multivariable analyses of early-onset breast cancer were adjusted for the above-mentioned covariates, and were additionally adjusted for age at menarche (<12, 12-13, ≥14 years, prefer not to answer or do not know), parity (nulliparous, parous, prefer not to answer or do not know), age at first live birth (<25, 25-29, ≥30 years, prefer not to answer or do not know), oral contraceptive use (yes, no, prefer not to answer or do not know), menopausal status and hormone replacement therapy use (premenopausal, postmenopausal with hormone replacement therapy, postmenopausal without hormone replacement therapy, prefer not to answer or do not know), and history of mammograms (yes, no, prefer not to answer or do not know).

Data analyses were performed using R software (version 4.3.1, https://www.r-project.org/), PLINK (version 2.0, https://www.cog-genomics.org/plink/2.0/),^38^ and the Polygenic Score Catalog Calculator (version 2.0, https://github.com/PGScatalog/pgsc_calc).^39^ Tests were two-sided, with *P* values <0.05 indicating statistical significance.

## Results

### Population Characteristics

A total of 1,247 incident invasive early-onset cancer cases (820 in females, 427 in males) were documented among 66,308 eligible participants during 329,509 person-years of follow-up (females: 168,897, males: 160,612), including 386 cases of incident early-onset breast cancer in females. The top three most common incident invasive early-onset cancers documented were breast cancer, melanoma, and colorectal cancer in females, and melanoma, colorectal cancer, and prostate cancer in males. (eTable 3 in Supplement 1)

The distribution of age, HLS and its components (smoking status, BMI, physical activity, alcohol consumption, and diet), total household income, and education did not vary appreciably across the PRSs of interest (female-specific total cancer PRS, male-specific total cancer PRS, and breast cancer PRS). Further, we observe no difference in age at menarche, parity, age at first live birth, oral contraceptive use, menopausal status, and hormone replacement therapy use across the PRSs. In contrast, individuals with higher PRSs (indicating higher genetic risk) in both sexes tended to have family history of cancer, and females with higher PRSs were more likely to have undergone mammography. (Tables 1, and eTable 5 in Supplement 1)

**Table 1.**
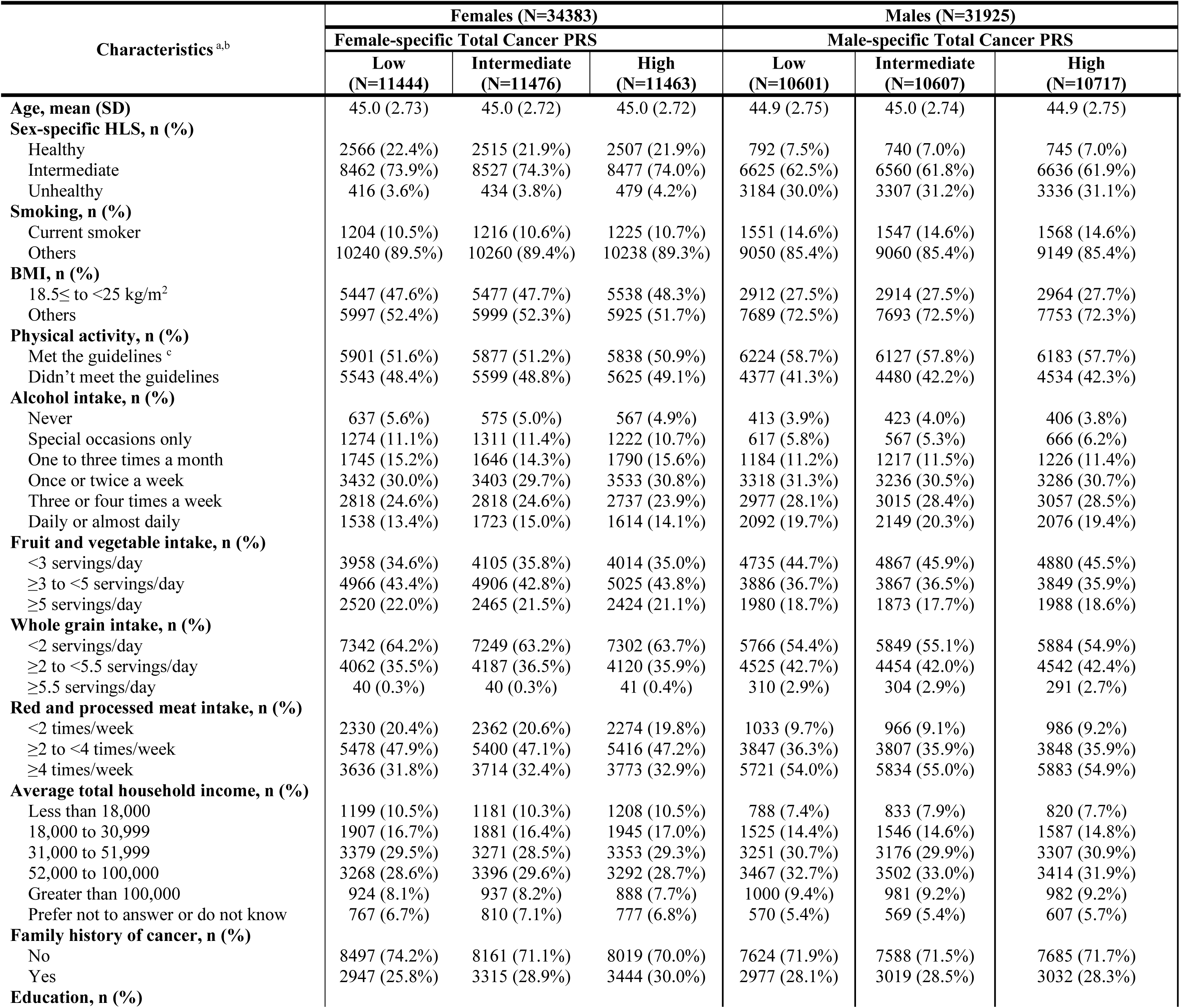
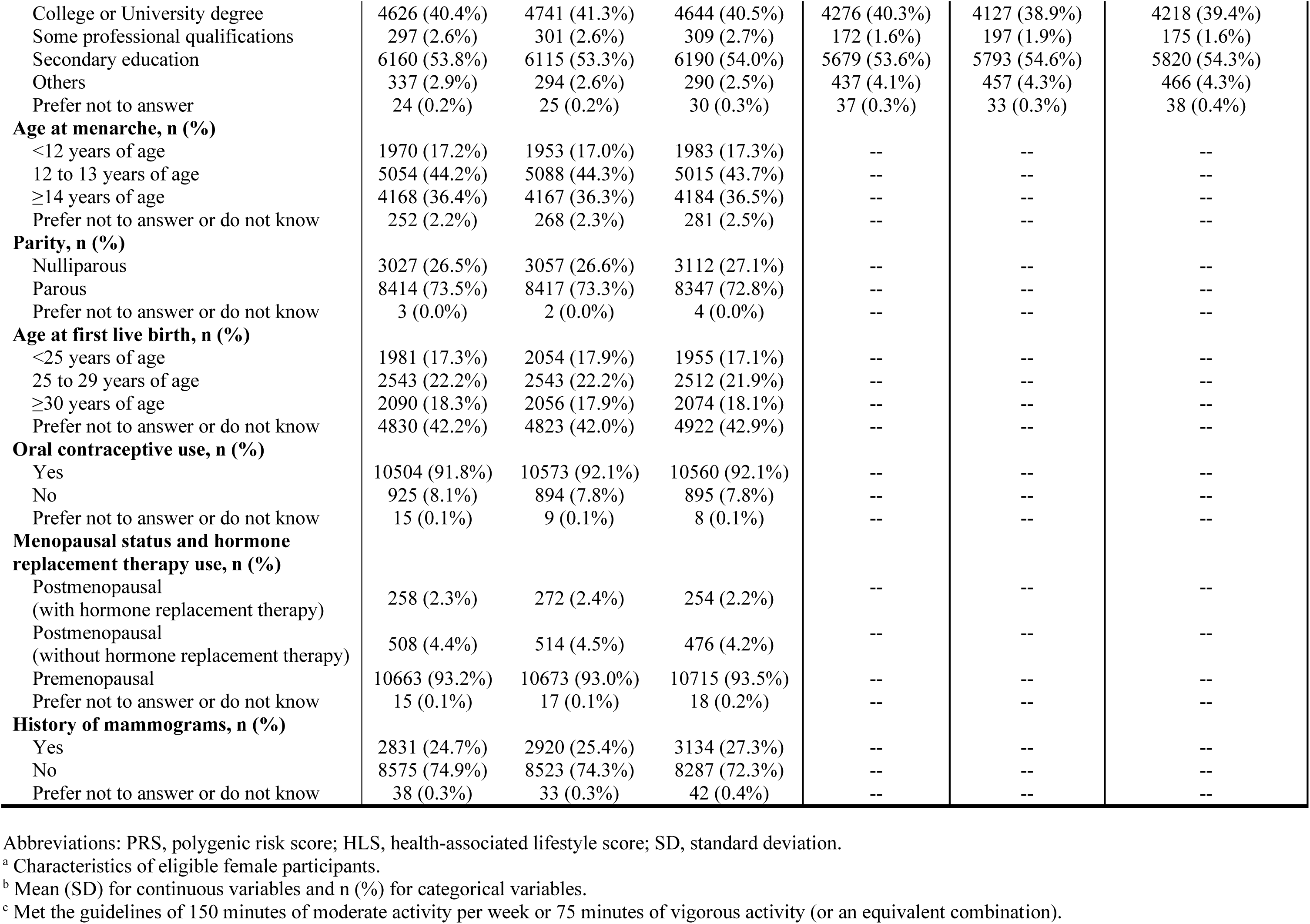
Characteristics of female and male participants at baseline according to female-specific and male-specific total cancer PRSs.

For both sexes, individuals with a higher HLS (indicating an unhealthier lifestyle) tended to have lower total household income and education levels and were more likely to have a family history of cancer. Among females, those with a higher HLS were less likely to undergo mammography or be premenopausal. Additionally, they were less likely to have experienced menarche between 12-13 years of age and to have their first birth before age 25 years. Moreover, they were more likely to use oral contraceptives. (eTables 6-7 in Supplement 1)

### PRS and the Risk of Early-onset Total and Breast Cancer

In multivariable-adjusted analyses with 2-year latency, higher genetic risk (highest vs. lowest tertile of PRS) was associated with significantly increased risks of early-onset total cancer in females (female-specific composite PRS HR=1.85; 95% CI, 1.50-2.29), early onset total cancer in males (male-specific composite PRS HR=1.94; 95% CI, 1.45-2.59), and early-onset breast cancer in females (breast-cancer PRS HR=3.06; 95% CI, 2.20-4.25). The HR (95% CI) per 1-SD increase in PRS was 1.29 (1.19, 1.40) for early-onset total cancer in females, 1.36 (1.22, 1.53) for early-onset total cancer in males, and 1.58 (1.41, 1.78) for early-onset breast cancer in females. (Figure 1)

**Figure 1.**
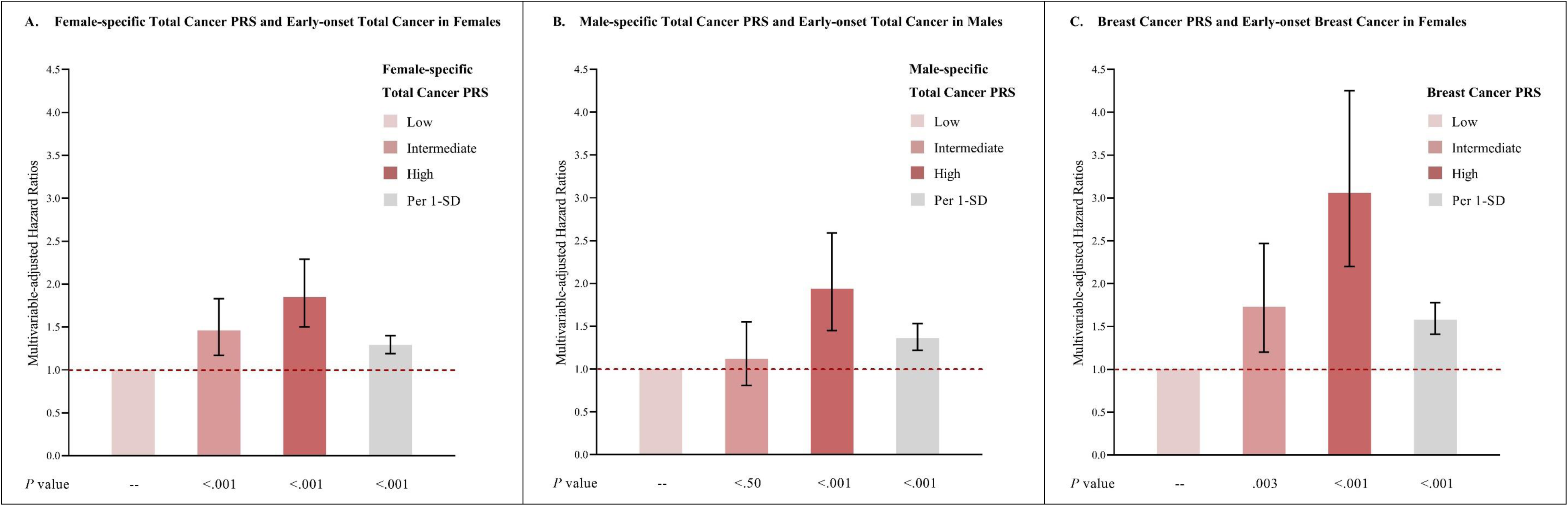
PRS and early-onset total and breast cancer risk. **A:** Multivariable-adjusted analysis of female-specific total cancer PRS and early-onset total cancer in females. **B**: Multivariable-adjusted analysis of male-specific total cancer PRS and early-onset total cancer in males. **C**: Multivariable-adjusted analysis of breast cancer PRS and early-onset breast cancer in females. Multivariate analyses of early-onset total cancer were stratified by sex, and adjust for the first 10 genetic principal components for ancestry, genotyping batch, average total household income, education, BMI (females only), and family history of cancer (family history of breast cancer was used in analyses of breast cancer), plus adjustment of HLS. Multivariate analyses of early-onset breast cancer were adjusted for above-mentioned covariates, and were additionally adjusted for age at menarche, parity, age at first live birth, oral contraceptive use, menopausal status and hormone replacement therapy use, and history of mammograms. Reference group: individuals with low PRS. Abbreviations: PRS, polygenic risk score; HLS, health-associated lifestyle score; SD, standard deviation; BMI, body mass index.

### HLS and the Risk of Early-onset Total and Breast Cancer

Adopting an unhealthy lifestyle (highest vs. lowest category of HLS) showed suggestive associations with increased risks in females of early-onset total cancer (female-specific HLS HR=1.49; 95% CI, 0.99-2.25), and early-onset breast cancer (HR=1.78; 95% CI, 0.97-3.24), but not for early-onset total cancer in males (male-specific HLS HR=1.14; 95% CI, 0.67-1.95). The associations among females were statistically significant when examining effect estimates on the scale of per 1-unit increase for early-onset total cancer (HR=1.12; 95%CI, 1.01-1.24) and early-onset breast cancer (HR=1.17; 95%CI, 1.02-1.35), but not in males for early-onset total cancer (HR=1.01; 95%CI, 0.90-1.14). (Figure 2)

**Figure 2:**
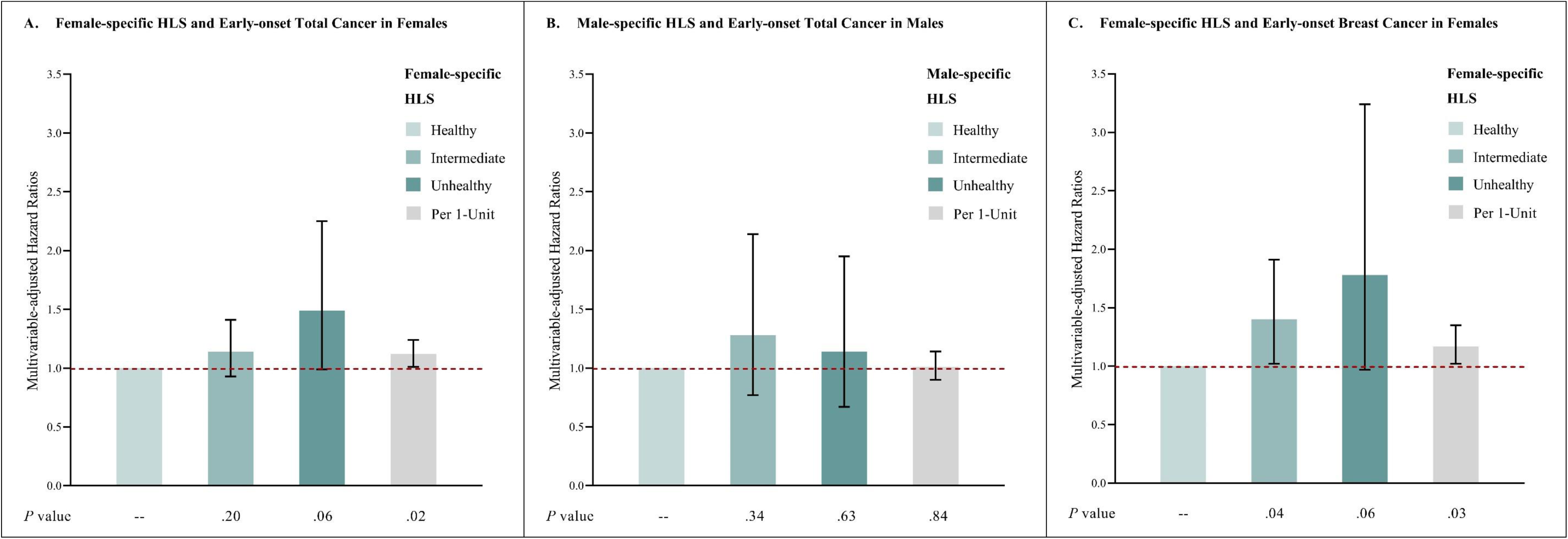
HLS and early-onset total and breast cancer risk. **A**: Multivariable-adjusted analysis of female-specific HLS and early-onset total cancer in females. **B**: Multivariable-adjusted analysis of male-specific HLS and early-onset total cancer in males. **C**: Multivariable-adjusted analysis of female-specific HLS and early-onset breast cancer in females. Multivariate analyses of early-onset total cancer were stratified by sex, and adjust for the first 10 genetic principal components for ancestry, genotyping batch, average total household income, education, BMI (females only), and family history of cancer (family history of breast cancer was used in analyses of breast cancer), plus adjustment of PRS. Multivariate analyses of early-onset breast cancer were adjusted for above-mentioned covariates, and were additionally adjusted for age at menarche, parity, age at first live birth, oral contraceptive use, menopausal status and hormone replacement therapy use, and history of mammograms. Reference group: individuals with healthy HLS. Abbreviations: PRS, polygenic risk score; HLS, health-associated lifestyle score; BMI, body mass index.

### HLS with Early-onset Total and Breast Cancer Risk, Stratified by PRS Category

The HRs (95% CI) of early-onset total cancer in females and males, and early-onset breast cancer in females associated with adopting an unfavorable lifestyle (highest vs. lowest category of HLS) were 1.85 (1.02, 3.36), 3.27 (0.78, 13.72), and 1.67 (0.71, 3.90), respectively, in those with high genetic risk; 1.25 (0.60, 2.57), 1.11 (0.42, 2.89), and 1.66 (0.54, 5.11), respectively, in those with intermediate genetic risk; and 1.15 (0.44, 2.98), 1.16 (0.39, 3.40), and 2.10 (0.57, 7.75), respectively, in those with low genetic risk. On the scale of per 1-unit increase in HLS, the HRs (95% CI) of early-onset total cancer in females and males, and early-onset breast cancer in females were 1.20 (1.03, 1.40), 1.06 (0.87, 1.29), and 1.22 (1.00, 1.48), respectively, in those with high genetic risk; 1.00 (0.84, 1.19), 1.01 (0.81, 1.26), and 1.14 (0.88, 1.48), respectively, in those with intermediate genetic risk; and 1.14 (0.93, 1.40), 0.98 (0.77, 1.25), and 1.07 (0.76, 1.51), respectively, in those with low genetic risk. The *P* value for the 2-df interaction test on the log HR scale was 0.39 for early-onset total cancer in females, 0.77 for early-onset total cancer in males, and 0.11 for early-onset breast cancer in females. (Figure 3)

**Figure 3:**
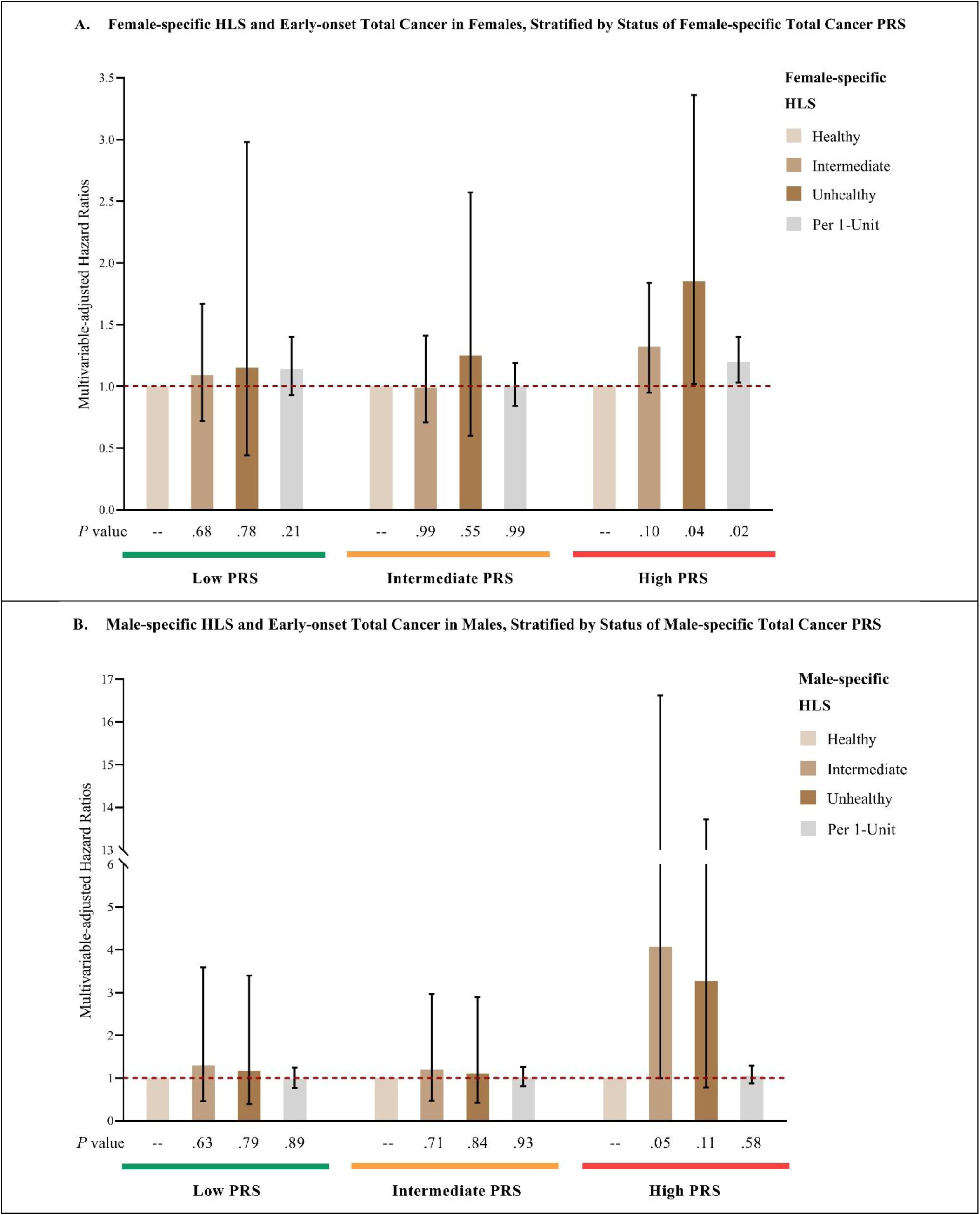
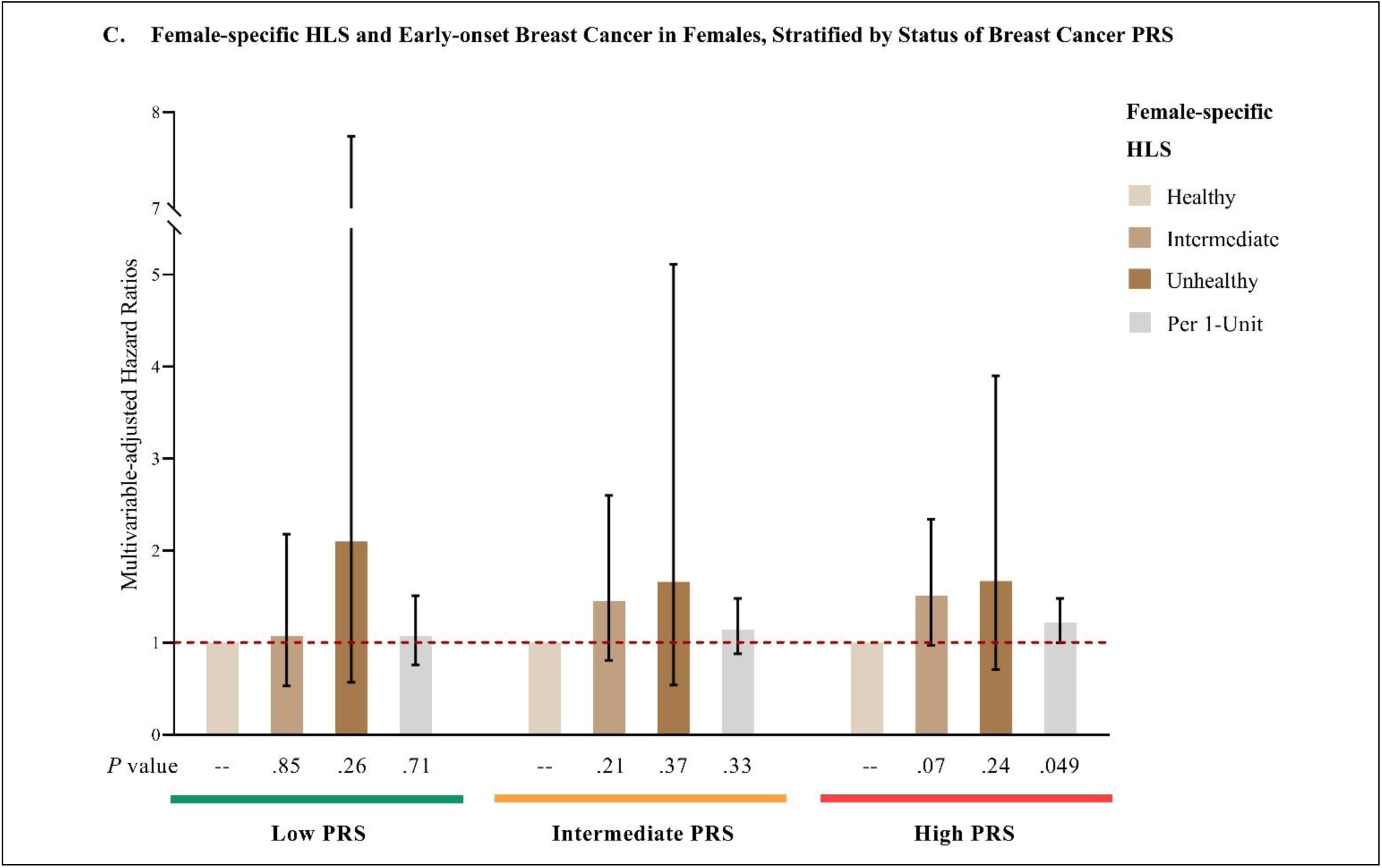
HLS and early-onset total and breast cancer risk, stratified by PRS category. **A**: Multivariable-adjusted analysis of female-specific HLS and early-onset total cancer in females, stratified by female-specific total cancer PRS. **B**: Multivariable-adjusted analysis of male-specific HLS and early-onset total cancer in males, stratified by male-specific total cancer PRS. **C**: Multivariable-adjusted analysis of female-specific HLS and early-onset breast cancer in females, stratified by breast cancer PRS. Multivariate analyses of early-onset total cancer were stratified by sex, and adjust for the first 10 genetic principal components for ancestry, genotyping batch, average total household income, education, BMI (females only), and family history of cancer (family history of breast cancer was used in analyses of breast cancer). Multivariate analyses of early-onset breast cancer were adjusted for above-mentioned covariates, and were additionally adjusted for age at menarche, parity, age at first live birth, oral contraceptive use, menopausal status and hormone replacement therapy use, and history of mammograms. Reference group: individuals with healthy HLS within each PRS stratum. Abbreviations: PRS, polygenic risk score; HLS, health-associated lifestyle score; BMI, body mass index.

### Joint Associations of PRS and HLS with Early-onset Total and Breast Cancer Risk

Compared to individuals with both low genetic risk (lowest tertile of PRS) and a favorable lifestyle (lowest category of HLS), those with both high genetic risk (highest tertile of PRS) and an unfavorable lifestyle (highest category of HLS) had increased risks of early-onset total cancer in females (HR=3.07; 95% CI, 1.64-5.78) and males (HR=2.18; 95% CI, 0.78-6.11), and significantly higher early-onset breast cancer risk in females (HR=4.11; 95% CI, 1.56-10.85). (Figure 4)

**Figure 4.**
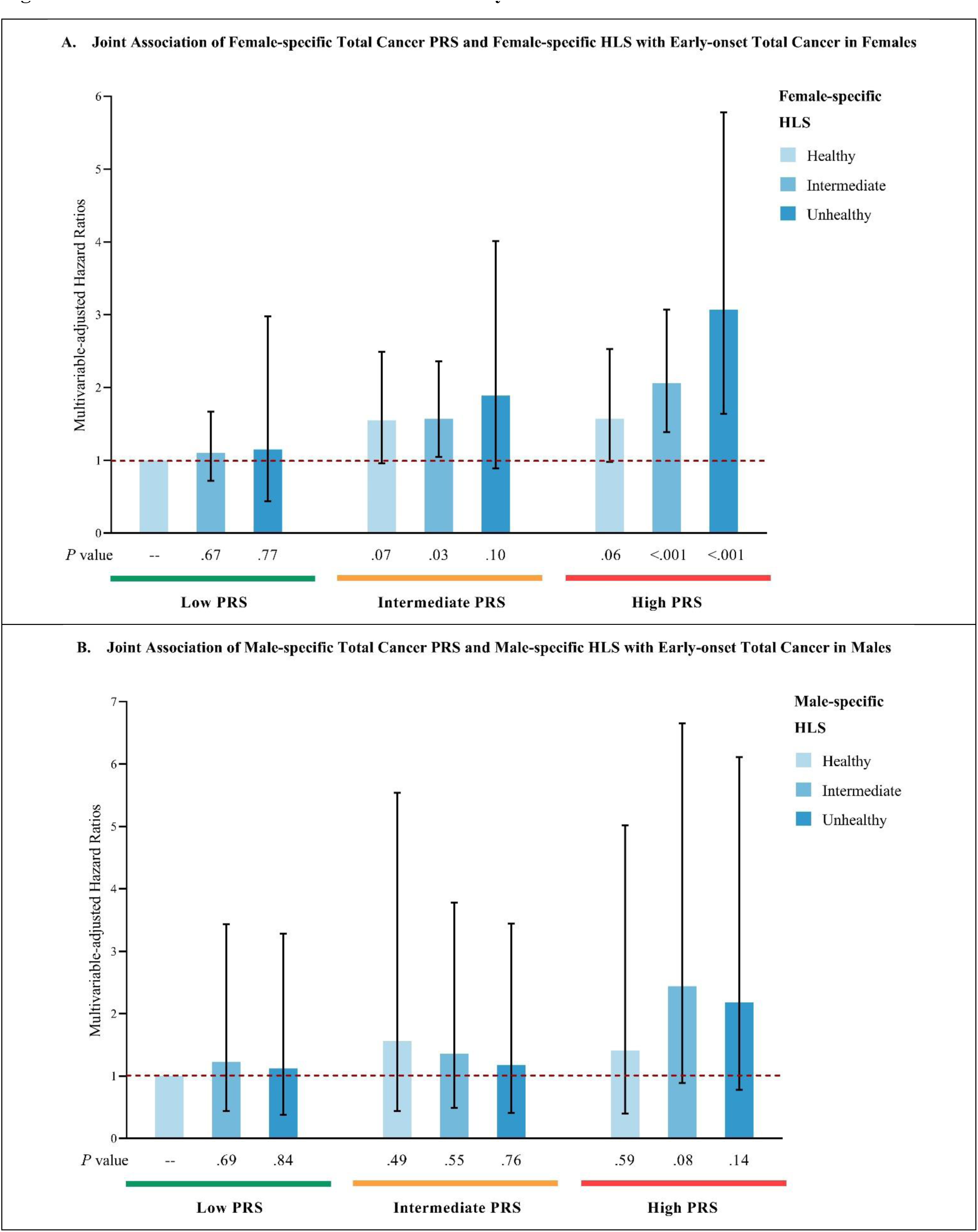
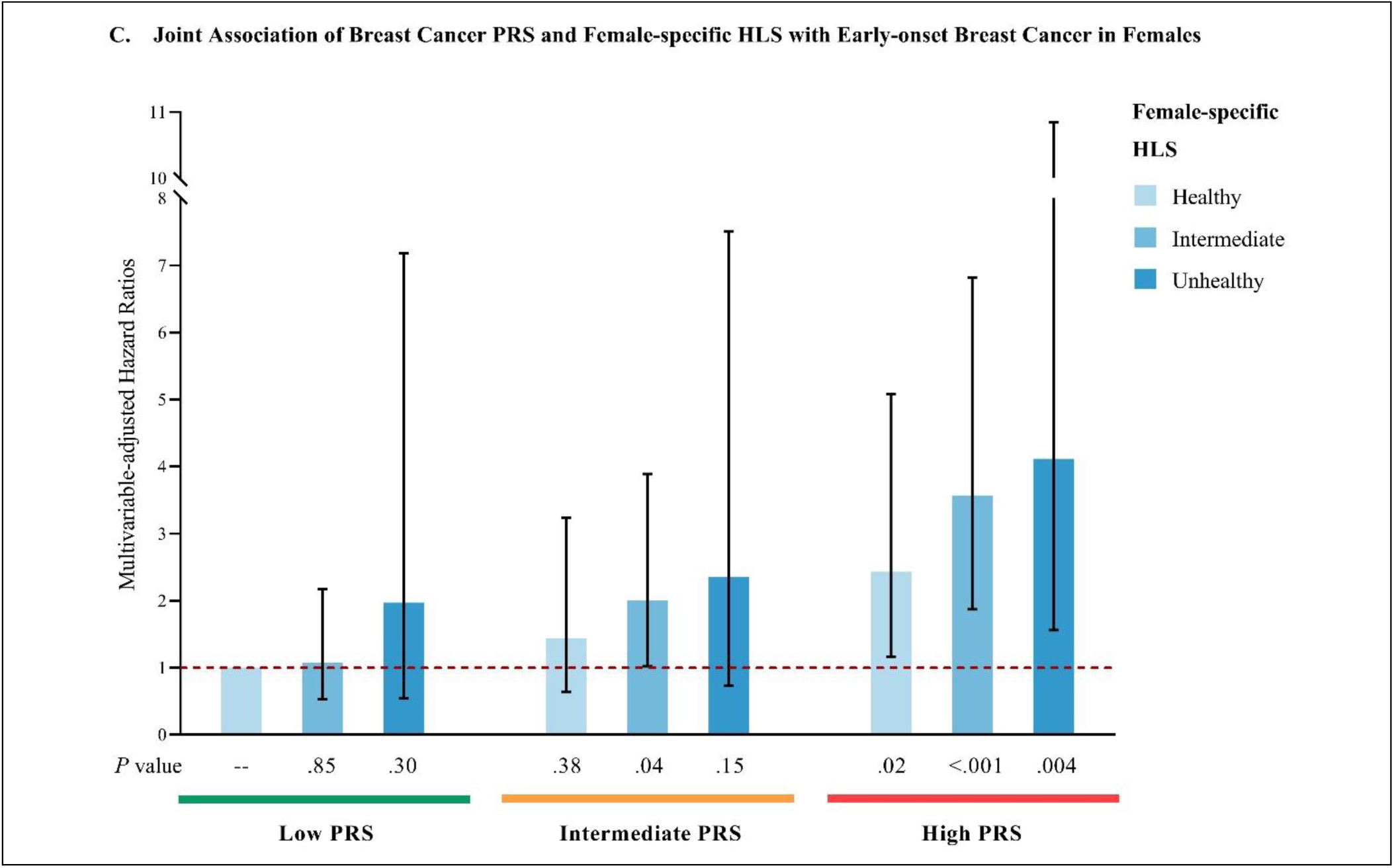
Joint associations of PRS and HLS with early-onset total and breast cancer risk. **A**: Multivariable-adjusted joint association analysis of female-specific total cancer PRS and female-specific HLS with early-onset total cancer in females. **B**: Multivariable-adjusted joint association analysis of male-specific total cancer PRS and male-specific HLS with early-onset total cancer in males. **C**: Multivariable-adjusted joint association analysis of breast cancer PRS and female-specific HLS with early-onset breast cancer in females. Multivariate analyses of early-onset total cancer were stratified by sex, and adjust for the first 10 genetic principal components for ancestry, genotyping batch, average total household income, education, BMI (females only), and family history of cancer (family history of breast cancer was used in analyses of breast cancer). Multivariate analyses of early-onset breast cancer were adjusted for above-mentioned covariates, and were additionally adjusted for age at menarche, parity, age at first live birth, oral contraceptive use, menopausal status and hormone replacement therapy use, and history of mammograms. Reference group: individuals with both low PRS and healthy HLS. Abbreviations: PRS, polygenic risk score; HLS, health-associated lifestyle score; BMI, body mass index.

## Discussion

Early-onset cancer is generally more aggressive compared to late-onset cancer with its rising incidence becoming a global concern. Deciphering the relationship between genetic risk, lifestyle modification, and risk of early-onset cancers may inform preventive strategies. In this large prospective cohort study among white British individuals, we present the first epidemiological evidence for this relationship for early-onset cancers.

Our results showed that genetic predisposition and lifestyle factors each demonstrated independent associations with the risk of early-onset total cancer and breast cancer. Stratified analyses by PRSs indicate that adopting a healthy lifestyle is likely beneficial for all individuals. Impressively, individuals with a high genetic risk may derive greater benefits from adopting a healthy lifestyle to prevent early-onset total cancer, compared to those with a low or intermediate genetic risk, although we were unable to detect statistically significant interaction between HLS and PRS mainly due to the limited number of early-onset cancer cases. These findings may inform preventive strategies for early-onset cancer in populations with varying genetic risk profiles.

To our knowledge, no previous study has investigated the extent to which adopting a healthy lifestyle could mitigate the impact of common genetic variants on the risk of early-onset cancers. A prior study of European ancestry population in the UK Biobank examined genetic risk and the benefits of adherence to a healthy lifestyle in relation to total cancer, irrespective of age at onset.^5^ Their findings demonstrated an additive interaction between genetic and lifestyle factors, indicating that individuals with a higher genetic risk may benefit more from lifestyle modification in relation to overall cancer risk in both sexes.^5^ Another study of invasive breast cancer among females of European ancestry in the UK Biobank reported that for premenopausal females, lifestyle intervention could have the greatest impact on those with a high genetic risk.^6^ Conversely, for postmenopausal females, lifestyle intervention might offer essentially similar benefits regardless of an individual’s genetic predisposition.^6^ The associations of genetic risk and lifestyle with risk of other major cancers, including cancers of thyroid, lung, stomach, pancreas, colorectum, ovarian, kidney, bladder, uterine, prostate, non-Hodgkin’s lymphoma, lymphocytic leukemia, and melanoma, have been reported.^8–14^ However, no such evidence has been reported for early-onset cancers.

### Strengths and Limitations

Our study has several strengths of note, including: 1) It provides the first epidemiological evidence addressing this important unanswered question in early-onset cancer prevention. 2) The prospective cohort study design, which minimized the potential for recall bias and selection bias. 3) The large sample size, with over 66,000 eligible participants involved in the final analytical population. 4) Ascertainment of cancer cases through linkage to national registries enhanced internal validity. 5) A wide spectrum of potential confounding factors was selected a priori and included as covariates in multivariable-adjusted analyses, which ensures relatively rigorous control for confounding. 6) Standardized protocols to assess heritable and lifestyle factors. 7) Novel approach for developing sex-specific composite total cancer PRSs to assess genetic risk of early-onset total cancer. 8) The concern of overfitting multicancer PRSs has been mitigated, given that 19 of the 23 site-specific cancer PRSs included did not incorporate UK Biobank data in their training samples. On average, the UK Biobank constitutes only 13% of the case data across these PRSs. 9) Applied 2-year latency to minimize the potential influence of reverse causation. 10) The analyses of early-onset total cancer were performed separately by sex to reflect the difference in cancer spectrum.

Our study also has several limitations: 1) The possibility for residual confounding cannot be completely ruled out due to the observational nature of study design. Making causal arguments should be approached with caution. 2) The aggregate analytic approach in analyses early-onset total cancer may potentially mask heterogeneity of effects across different cancer types, though it increases the power to detect the effects of interest. 3) The inability to explore heterogeneity across individual cancer types other than breast cancer further, due to power limitations. 4) The ethnic homogeneity of our analytic population (all white British) may limit generalizability of current findings to other groups. 5) Lifestyle factors were assessed based on a single measurement at baseline rather than through repeated measurements (which can better depict long-term trends), raising concern about the potential influence of random within-person variation. 6) We were not able to consider the age of 45 years^40^ as the cutoff in analyses of early-onset breast cancer due to the limited number of incident cases. We were unable to consider separate analysis by ER status because information on hormone-receptor subtype was unavailable in the UK Biobank.

Future large prospective investigations with repeated measurements and long-term follow-up are warranted to provide additional evidence on these associations in early-onset cancers in diverse populations and to examine potential heterogeneity across individual cancer types in the associations of interest.

### Conclusions

Both genetic and lifestyle factors were independently associated with risks of early-onset total cancer and breast cancer. Compared to those with low genetic risk, individuals with a high genetic risk may benefit more from adopting a healthy lifestyle in preventing early-onset cancer.

## Article Information

### Author Contributions

YZ, YL and PK have full access to all the data in the study and take responsibility for the integrity of the data and the accuracy of the data analysis. Concept and design: YZ, YL, and PK. Acquisition, analysis, or interpretation of data: All authors. Drafting of the manuscript: YZ and YL. Critical revision of the manuscript for important intellectual content: SL and PK. Statistical analysis: YZ and YL. Obtained funding: YZ, SL, and PK. Administrative, technical, or material support: SL and PK. Supervision: PK and YL.

### Disclaimer

The authors obtained access to the UK Biobank data through the approved project application 70925. The authors assume full responsibility for analyses and interpretation of these data. The contents of this publication are solely the responsibility of the authors and do not necessarily represent the official views of any agencies. The authors thank Dr. Kathryn L. Penney at the Harvard T. H. Chan School of Public Health and Harvard Medical School for her insightful comments and suggestions for this manuscript.

### Ethical Approval

The details of UK Biobank research ethics approval are elaborated at https://www.ukbiobank.ac.uk/learn-more-about-uk-biobank/about-us/ethics.

## Supporting information

eTables S1-S23

## Data Availability

The authors obtained access to the UK Biobank data through the approved project application 70925.

## Supplement 1 (Major Supplementary Tables)

**eTable 1.** List of published cancer site-specific PRSs included in the lasso regression in females, and list of cancer site-specific PRSs selected from the lasso regression for the development of the female-specific composite total cancer PRS.

| Cancer Types | Polygenic Score ID & Name (PGS catalog)<br>included in the lasso regression <sup>a</sup> | Number<br>of SNPs | Publication | PMID | Selected from the<br>lasso regression |
| --- | --- | --- | --- | --- | --- |
| Breast | <u>PGS000004 (PRS313_BC) <sup>b</sup></u> | 313 | <u>Mavaddat N et al. Am J Hum Genet (2018)</u> | 30554720 | Yes |
| Melanoma | <u>PGS000790 (CC_Melanoma_IV)</u> | 24 | <u>Kachuri L et al. Nat Commun (2020)</u> | 33247094 | No |
| Colorectum * | <u>PGS002252 (PRS_CRC)</u> | 141 | <u>Archambault AN et al. J Natl Cancer Inst (2022)</u><br><u>Huyghe JR et al. Nat Genet (2019)</u> | 35026030<br>30510241 | No |
| Non-Hodgkin lymphoma | <u>PGS000791 (CC_NHL_IV)</u> | 19 | <u>Kachuri L et al. Nat Commun (2020)</u> | 33247094 | No |
| Ovary | <u>PGS003394 (PRS_Stepwise)</u> | 36 | <u>Dareng EO et al. Eur J Hum Genet (2022)</u> | 35027648 | No |
| Brain * | <u>PGS000624 (PRSWEB_PHECODE191.11_GWAS-Catalog-r2019-05-03-X191.11_PT_UKB_20200608)</u> | 5 | <u>Fritsche LG et al. Am J Hum Genet (2020)</u> | 32991828 | No |
| Head and neck | <u>PGS000792 (CC_Oral_IV)</u> | 14 | <u>Kachuri L et al. Nat Commun (2020)</u> | 33247094 | No |
| Lung | <u>PGS000740 (PRS128_LC)</u> | 128 | <u>Hung RJ et al. Cancer Res (2021)</u> | 33472890 | Yes |
| Endometrium | <u>PGS000786 (CC_Endo_IV)</u> | 9 | <u>Kachuri L et al. Nat Commun (2020)</u> | 33247094 | No |
| Thyroid | <u>PGS000797 (CC_Thyroid_IV)</u> | 12 | <u>Kachuri L et al. Nat Commun (2020)</u> | 33247094 | No |
| Kidney | <u>PGS000787 (CC_Kidney_IV)</u> | 19 | <u>Kachuri L et al. Nat Commun (2020)</u> | 33247094 | No |
| Multiple myeloma * | <u>PGS000653 (PRSWEB_PHECODE204.4_GWAS-Catalog-r2019-05-03-X204.4_P_5e-08_UKB_20200608)</u> | 22 | <u>Fritsche LG et al. Am J Hum Genet (2020)</u> | 32991828 | Yes |
| Cervix | <u>PGS000784 (CC_Cervix_IV)</u> | 10 | <u>Kachuri L et al. Nat Commun (2020)</u> | 33247094 | No |
| Pancreas | <u>PGS000794 (CC_Pancreas_IV)</u> | 22 | <u>Kachuri L et al. Nat Commun (2020)</u> | 33247094 | No |
| Stomach | <u>PGS002299 (PRS3_gastric)</u> | 3 | <u>Choi J et al. Int J Cancer (2020)</u><br><u>Helgason et al. Nat Genet (2015)</u> | 32588423<br>26098866 | Yes |
| Lymphoid leukemia | <u>PGS000788 (CC_LL_IV)</u> | 75 | <u>Kachuri L et al. Nat Commun (2020)</u> | 33247094 | No |
| Esophagus | <u>PGS000364 (PRSWEB_PHECODE150_C15_LASSOSUM_MGI_20200608)</u> | 2001 | <u>Fritsche LG et al. Am J Hum Genet (2020)</u> | 32991828 | No |
| Hodgkin's lymphoma | <u>PGS000639 (PRSWEB_PHECODE201_GWAS-Catalog-r2019-05-03-X201_PT_MGI_20200608)</u> | 20 | <u>Fritsche LG et al. Am J Hum Genet (2020)</u> | 32991828 | No |
| Trachea * | <u>PGS000392 (PRSWEB_PHECODE165.1_GWAS-Catalog-r2019-05-03-X165.1_PT_UKB_20200608)</u> | 19 | <u>Fritsche LG et al. Am J Hum Genet (2020)</u> | 32991828 | No |

Abbreviations: PRS, polygenic risk score; GRCh37, Genome Reference Consortium Human Build 37.
<sup>a</sup> Published cancer site-specific PRSs included in the lasso regression were selected based on their training sample size, developing methods, test-set performance, possibility of overfit (i.e. training sample should not include UK Biobank), and the availability of SNPs and weights.
<sup>b</sup> PGS000004 (PRS313\_BC) was used specifically in analyses of breast cancer PRS and early-onset breast cancer among females.
\* UK Biobank was included in the training set.

**eTable 2.**
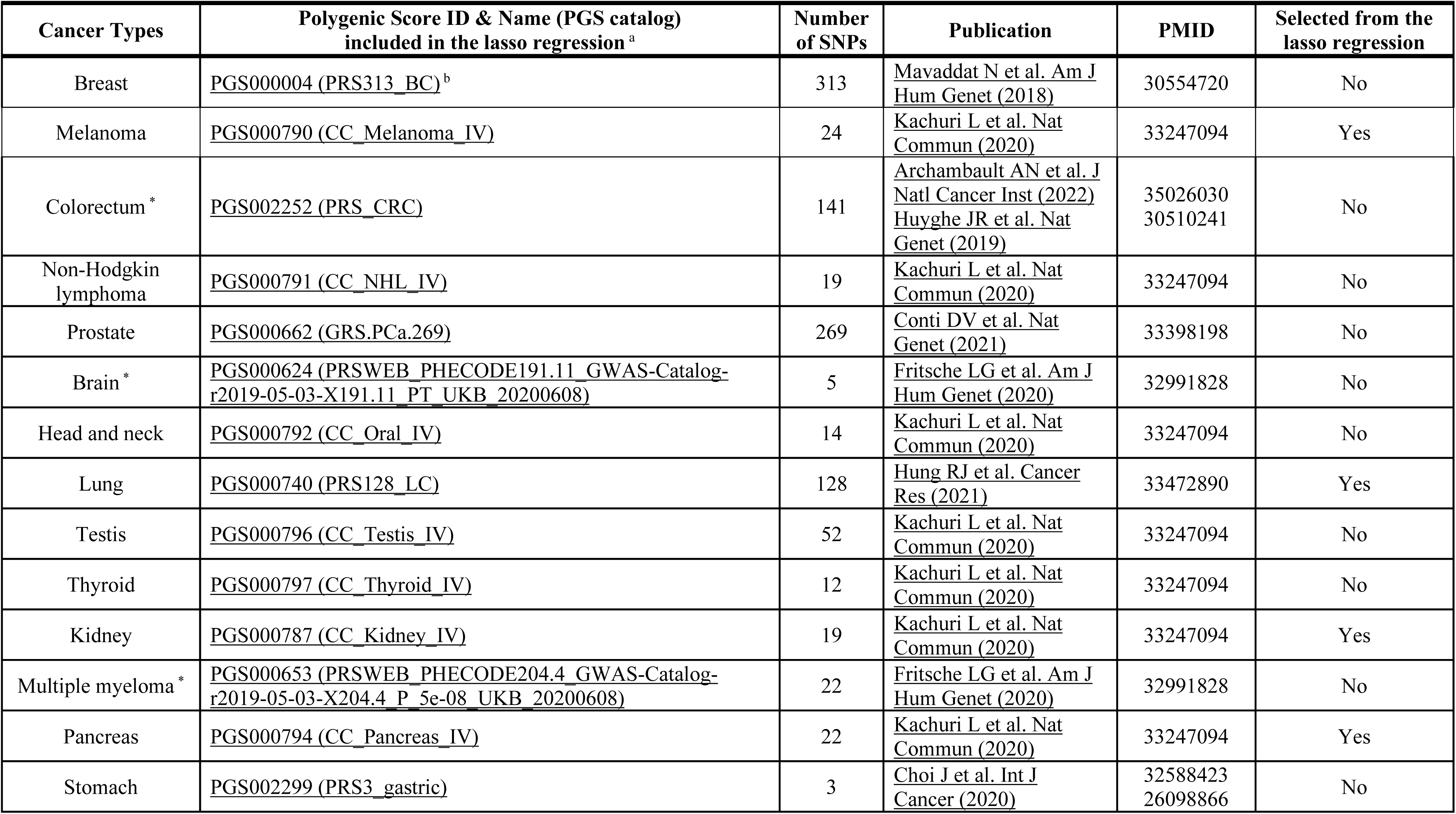
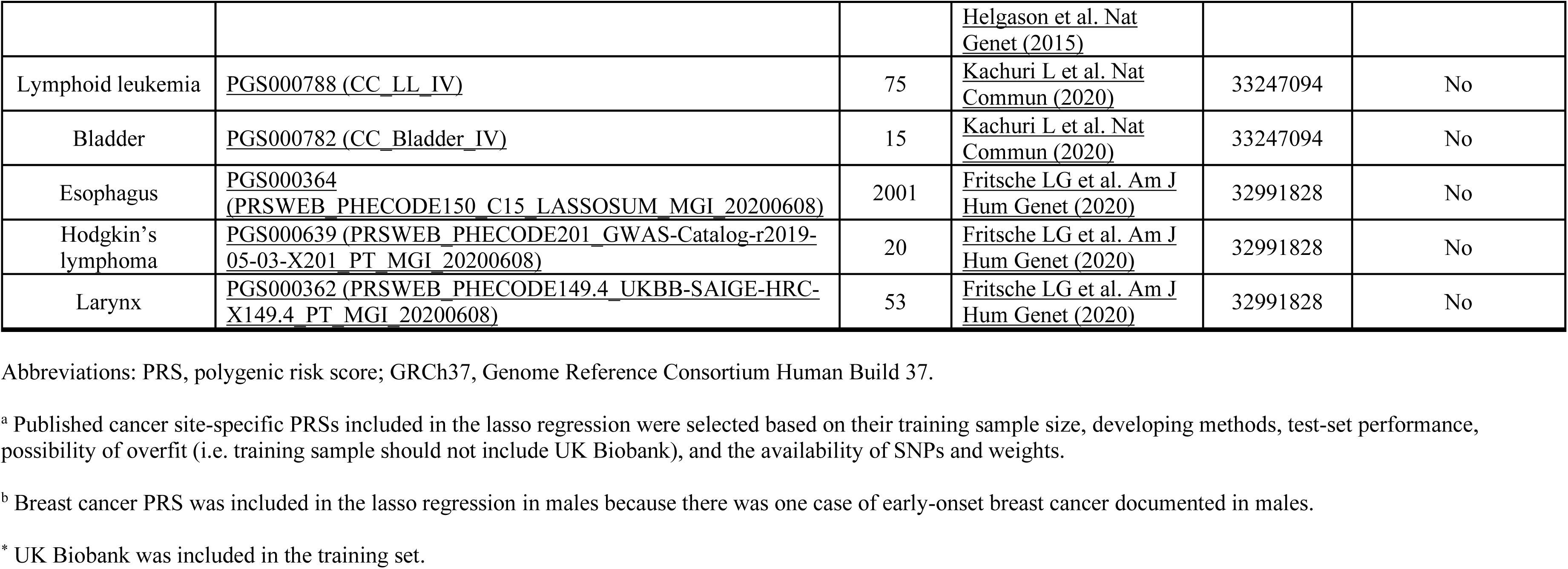
List of published cancer site-specific PRSs included in the lasso regression in males, and list of cancer site-specific PRSs selected from the lasso regression for the development of the male-specific composite total cancer PRS.

**eTable 3.** Summary of early-onset cancer spectrum in females and males, showcasing cancers with published site-specific PRSs qualified for inclusion in the lasso regression.

| **Cancer Types with Published Site-specific PRSs qualified for inclusion in the lasso regression** | **Number of Cases (jn females)** | **Number of Cases (in males)** | **Number of Cases (in the total study population)** |
| --- | --- | --- | --- |
| Breast | 386 | 1 | 387 |
| Melanoma | 60 | 36 | 96 |
| Colorectum | 23 | 28 | 51 |
| Brain | 11 | 19 | 30 |
| Non-Hodgkin lymphoma | 7 | 22 | 29 |
| Prostate | 0 | 28 | 28 |
| Ovary | 27 | 0 | 27 |
| Testis | 0 | 22 | 22 |
| Kidney | 7 | 9 | 16 |
| Head and neck | 7 | 9 | 16 |
| Lung | 8 | 7 | 15 |
| Endometrium | 12 | 0 | 12 |
| Thyroid | 9 | 3 | 12 |
| Multiple myeloma | 4 | 8 | 12 |
| Lymphoid leukemia | 4 | 5 | 9 |
| Cervix | 8 | 0 | 8 |
| Pancreas | 5 | 3 | 8 |
| Stomach | 2 | 5 | 7 |
| Bladder | 0 | 6 | 6 |
| Esophagus | 1 | 2 | 3 |
| Hodgkin's lymphoma | 2 | 1 | 3 |
| Larynx | 0 | 3 | 3 |
| Trachea | 1 | 0 | 1 |
| **Sum** | **584** | **217** | **801** |
| Others | 236 | 210 | 446 |
| **Total** | **820** | **427** | **1,247** |

**eTable 4.** Components of female-specific HLS and male-specific HLS.

| Components | Categories | Score assignments |
| --- | --- | --- |
| **Smoking status** | Current smoker<br>Others | **Smoking score**<br>1 for current smoker<br>0 for others |
| **BMI (males only) <sup>a</sup>** | 18.5≤ to <25 kg/m<sup>2</sup><br>Others | **BMI score**<br>0 for 18.5≤ to <25 kg/m<sup>2</sup><br>1 for others |
| **Physical activity** | Met the guidelines <sup>b</sup><br>Didn't meet the guidelines <sup>b</sup> | **Physical activity score**<br>0 for met the guidelines <sup>b</sup><br>1 for didn't meet the guidelines <sup>b</sup> |
| **Alcohol consumption** | Never<br>Others | **Alcohol intake score**<br>0 for never<br>1 for others |
| **Diet** | **Fruit and vegetable intake**<br><3 servings/day<br>≥3 to <5 servings/day<br>≥5 servings/day<br>*\*Amount per serving:*<br>*Fresh fruit: 1 piece*<br>*Dried fruit: 5 pieces*<br>*Cooked vegetables: 3 heaped tablespoons*<br>*Salad/raw vegetables: 3 heaped tablespoons*<br>**Whole grain intake**<br><2 servings/day<br>≥2 to <5.5 servings/day<br>≥5.5 servings/day<br>*\*Amount per serving:*<br>*Cereal: 1 bowl*<br>*Whole-meal bread: 1 slice*<br>*Whole-grain bread: 1 slice* | **Fruit and vegetable intake**<br>1 for <3 servings/day<br>0.5 for ≥3 to <5 servings/day<br>0 for ≥5 servings/day<br>**Whole grain intake**<br>1 for <2 servings/day<br>0.5 for ≥2 to <5.5 servings/day<br>0 for ≥5.5 servings/day<br>**Red and processed meat intake**<br>1 for ≥4 times/week<br>0.5 for ≥2 to <4 times/week<br>0 for <2 times/week<br>**Diet score**<br>(Fruit and vegetable intake score +<br>Whole grain intake score +<br>Red and processed meat intake score) / 3<br>1 for ≥0.5 |
| | *Brown bread: 1 slice*<br>*Red and processed meat intake*<br>$\geq 4$ times/week<br>$\geq 2$ to $< 4$ times/week<br>$< 2$ times/week | 0 for $< 0.5$ |
| **Total Score** |  |  |
| <u>**Female-specific HLS**</u><br>= *Smoking score* + *Physical activity score*<br>+ *Alcohol intake score* + *Diet score*<br>Ranging from 0 to 4, with higher scores indicating an unhealthier lifestyle.<br>Healthy: 0 to 1<br>Intermediate: 2 to 3<br>Unhealthy: 4 | <u>**Male-specific HLS**</u><br>= *Smoking score* + *BMI score* + *Physical activity score*<br>+ *Alcohol intake score* + *Diet score*<br>Ranging from 0 to 5, with higher scores indicating an unhealthier lifestyle.<br>Healthy: 0 to 1<br>Intermediate: 2 to 3<br>Unhealthy: 4 to 5 |  |

Abbreviations: HLS, health-associated lifestyle score; BMI, body mass index.
<sup>a</sup> BMI was not included as a component of female-specific HLS due to the widely recognized inverse association of BMI with the risk of early-onset/pre-menopausal breast cancer and the fact that early-onset total cancer in females was predominantly driven by early-onset breast cancer.
<sup>b</sup> Met the guidelines of 150 minutes of moderate activity per week or 75 minutes of vigorous activity (or an equivalent combination).

**eTable 5.** Characteristics of female participants (N=34383) at baseline according to breast cancer PRS.

| Characteristics<sup>a,b</sup> | Breast Cancer PRS |  |  |
| --- | --- | --- | --- |
|  | Low<br>(N=11428) | Intermediate<br>(N=11481) | High<br>(N=11474) |
| **Age, mean (SD)** | 45.0 (2.72) | 45.0 (2.73) | 45.0 (2.72) |
| **Female-specific HLS, n (%)** |  |  |  |
| Healthy | 2557 (22.4%) | 2504 (21.8%) | 2527 (22.0%) |
| Intermediate | 8468 (74.1%) | 8524 (74.2%) | 8474 (73.9%) |
| Unhealthy | 403 (3.5%) | 453 (3.9%) | 473 (4.1%) |
| **Smoking, n (%)** |  |  |  |
| Current smoker | 1201 (10.5%) | 1214 (10.6%) | 1230 (10.7%) |
| Others | 10227 (89.5%) | 10267 (89.4%) | 10244 (89.3%) |
| **BMI, n (%)** |  |  |  |
| 18.5≤ to <25 kg/m<sup>2</sup> | 5424 (47.5%) | 5503 (47.9%) | 5535 (48.2%) |
| Others | 6004 (52.5%) | 5978 (52.1%) | 5939 (51.8%) |
| **Physical activity, n (%)** |  |  |  |
| Met the guidelines<sup>c</sup> | 5875 (51.4%) | 5930 (51.7%) | 5811 (50.6%) |
| Didn't meet the guidelines | 5553 (48.6%) | 5551 (48.3%) | 5663 (49.4%) |
| **Alcohol intake, n (%)** |  |  |  |
| Never | 633 (5.5%) | 591 (5.1%) | 555 (4.8%) |
| Special occasions only | 1253 (11.0%) | 1322 (11.5%) | 1232 (10.7%) |
| One to three times a month | 1725 (15.1%) | 1687 (14.7%) | 1769 (15.4%) |
| Once or twice a week | 3418 (29.9%) | 3431 (29.9%) | 3519 (30.7%) |
| Three or four times a week | 2831 (24.8%) | 2793 (24.3%) | 2749 (24.0%) |
| Daily or almost daily | 1568 (13.7%) | 1657 (14.4%) | 1650 (14.4%) |
| **Fruit and vegetable intake, n (%)** |  |  |  |
| <3 servings/day | 3933 (34.4%) | 4157 (36.2%) | 3987 (34.7%) |
| ≥3 to <5 servings/day | 4959 (43.4%) | 4892 (42.6%) | 5046 (44.0%) |
| ≥5 servings/day | 2536 (22.2%) | 2432 (21.2%) | 2441 (21.3%) |
| **Whole grain intake, n (%)** |  |  |  |
| <2 servings/day | 7273 (63.6%) | 7343 (64.0%) | 7277 (63.4%) |
| ≥2 to <5.5 servings/day | 4114 (36.0%) | 4099 (35.7%) | 4156 (36.2%) |
| ≥5.5 servings/day | 41 (0.4%) | 39 (0.3%) | 41 (0.4%) |
| **Red and processed meat intake, n (%)** |  |  |  |
| <2 times/week | 2344 (20.5%) | 2310 (20.1%) | 2312 (20.2%) |
| ≥2 to <4 times/week | 5463 (47.8%) | 5437 (47.4%) | 5394 (47.0%) |
| ≥4 times/week | 3621 (31.7%) | 3734 (32.5%) | 3768 (32.8%) |
| **Average total household income, n (%)** |  |  |  |
| Less than 18,000 | 1177 (10.3%) | 1172 (10.2%) | 1239 (10.8%) |
| 18,000 to 30,999 | 1888 (16.5%) | 1922 (16.7%) | 1923 (16.8%) |
| 31,000 to 51,999 | 3361 (29.4%) | 3303 (28.8%) | 3339 (29.1%) |
| 52,000 to 100,000 | 3311 (29.0%) | 3361 (29.3%) | 3284 (28.6%) |
| Greater than 100,000 | 926 (8.1%) | 923 (8.0%) | 900 (7.8%) |
| Prefer not to answer or do not know | 765 (6.7%) | 800 (7.0%) | 789 (6.9%) |
| **Family history of breast cancer, n (%)** |  |  |  |
| No | 10583 (92.6%) | 10315 (89.8%) | 10092 (88.0%) |
| Yes | 845 (7.4%) | 1166 (10.2%) | 1382 (12.0%) |
| **Education, n (%)** |  |  |  |
| College or University degree | 4649 (40.7%) | 4702 (41.0%) | 4660 (40.6%) |
| Some professional qualifications | 293 (2.6%) | 305 (2.7%) | 309 (2.7%) |
| Secondary education | 6124 (53.6%) | 6148 (53.5%) | 6193 (54.0%) |
| Others | 336 (2.9%) | 302 (2.6%) | 283 (2.5%) |
| Prefer not to answer | 26 (0.2%) | 24 (0.2%) | 29 (0.3%) |
| **Age at menarche, n (%)** |  |  |  |
| <12 years of age | 1937 (16.9%) | 1983 (17.3%) | 1986 (17.3%) |
| 12 to 13 years of age | 5050 (44.2%) | 5060 (44.1%) | 5047 (44.0%) |
| ≥14 years of age | 4180 (36.6%) | 4172 (36.3%) | 4167 (36.3%) |
| Prefer not to answer or do not know | 261 (2.3%) | 266 (2.3%) | 274 (2.4%) |
| **Parity, n (%)** |  |  |  |
| Nulliparous | 3059 (26.8%) | 3026 (26.4%) | 3111 (27.1%) |
| Parous | 8366 (73.2%) | 8453 (73.6%) | 8359 (72.9%) |
| Prefer not to answer or do not know | 3 (0.0%) | 2 (0.0%) | 4 (0.0%) |
| **Age at first live birth, n (%)** |  |  |  |
| <25 years of age | 2004 (17.5%) | 2027 (17.7%) | 1959 (17.1%) |
| 25 to 29 years of age | 2508 (21.9%) | 2564 (22.3%) | 2526 (22.0%) |
| ≥30 years of age | 2048 (17.9%) | 2068 (18.0%) | 2104 (18.3%) |
| Prefer not to answer or do not know | 4868 (42.6%) | 4822 (42.0%) | 4885 (42.6%) |
| **Oral contraceptive use, n (%)** |  |  |  |
| Yes | 10473 (91.6%) | 10595 (92.3%) | 10569 (92.1%) |
| No | 939 (8.2%) | 878 (7.6%) | 897 (7.8%) |
| Prefer not to answer or do not know | 16 (0.1%) | 8 (0.1%) | 8 (0.1%) |
| **Menopausal status and hormone replacement therapy use, n (%)** |  |  |  |
| Postmenopausal (with hormone replacement therapy) | 266 (2.3%) | 264 (2.3%) | 254 (2.2%) |
| Postmenopausal (without hormone replacement therapy) | 504 (4.4%) | 512 (4.5%) | 482 (4.2%) |
| Premenopausal | 10641 (93.1%) | 10687 (93.1%) | 10723 (93.5%) |
| Prefer not to answer or do not know | 17 (0.1%) | 18 (0.2%) | 15 (0.1%) |
| **History of mammograms, n (%)** |  |  |  |
| Yes | 2799 (24.5%) | 2954 (25.7%) | 3132 (27.3%) |
| No | 8592 (75.2%) | 8490 (73.9%) | 8303 (72.4%) |
| Prefer not to answer or do not know | 37 (0.3%) | 37 (0.3%) | 39 (0.3%) |

Abbreviations: PRS, polygenic risk score; HLS, health-associated lifestyle score; SD, standard deviation.
<sup>a</sup> Characteristics of eligible female participants.
<sup>b</sup> Mean (SD) for continuous variables and n (%) for categorical variables.
<sup>c</sup> Met the guidelines of 150 minutes of moderate activity per week or 75 minutes of vigorous activity (or an equivalent combination).

**eTable 6.**
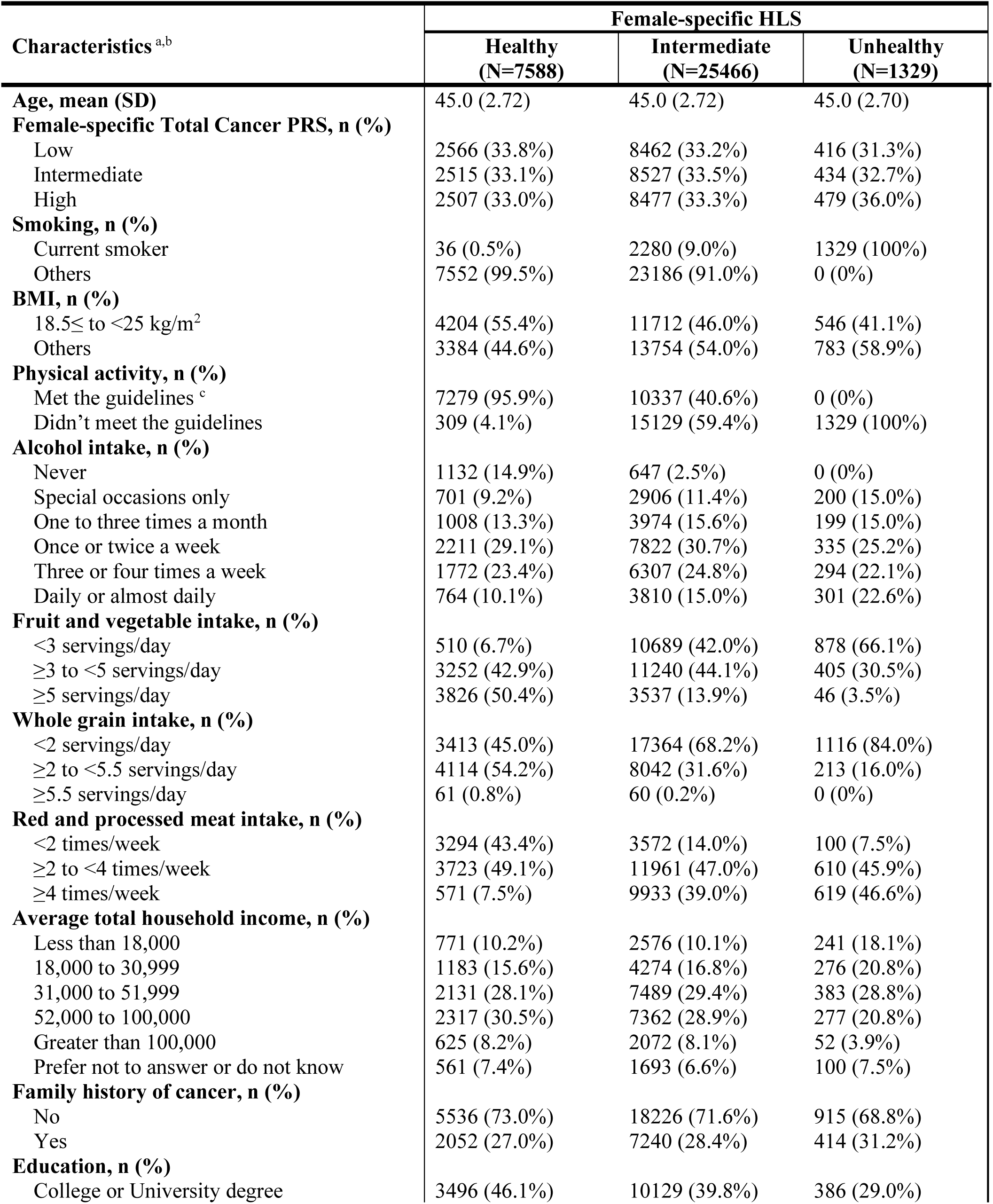
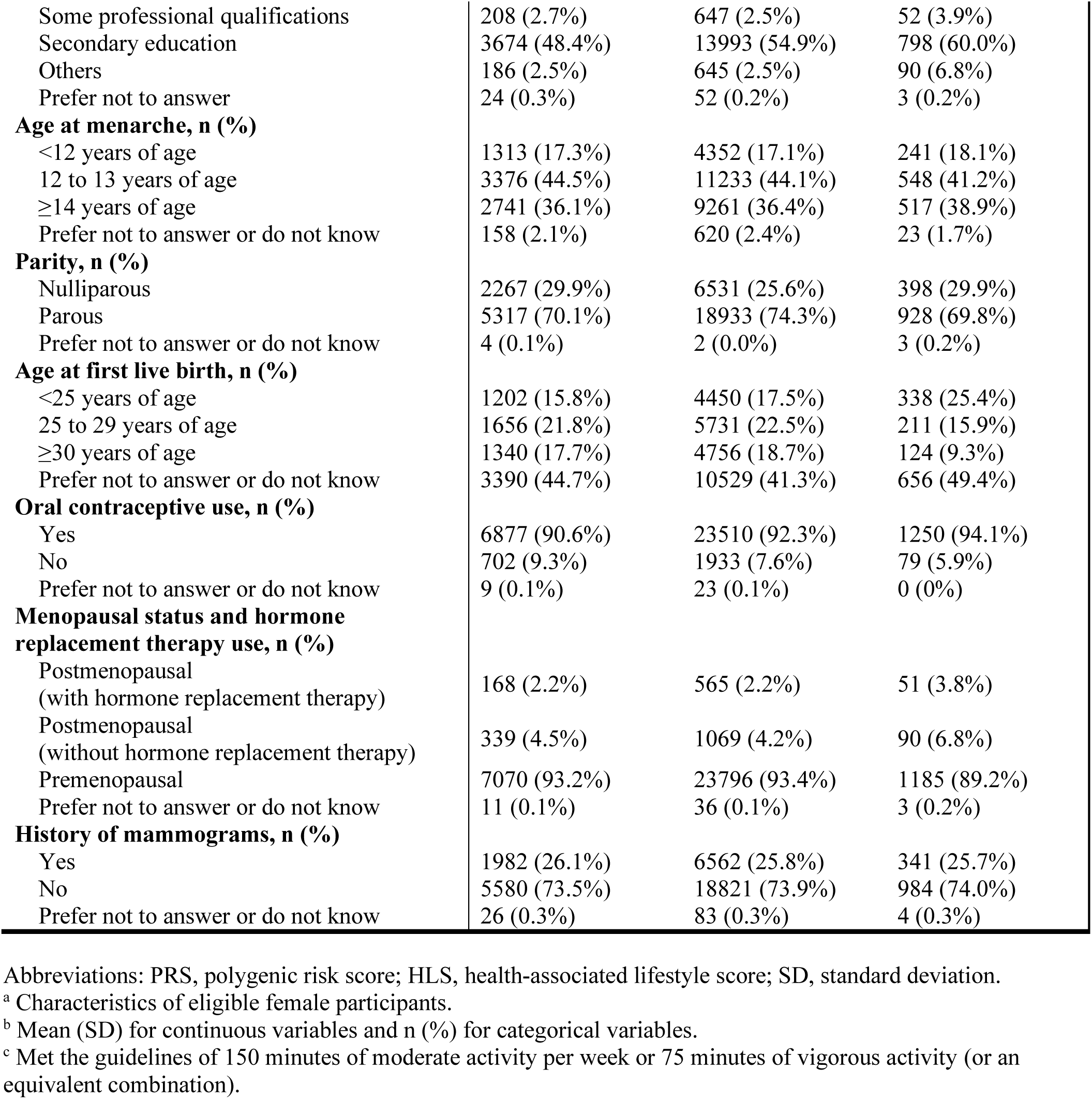
Characteristics of female participants (N=34383) at baseline according to female-specific HLS.

**eTable 7.**
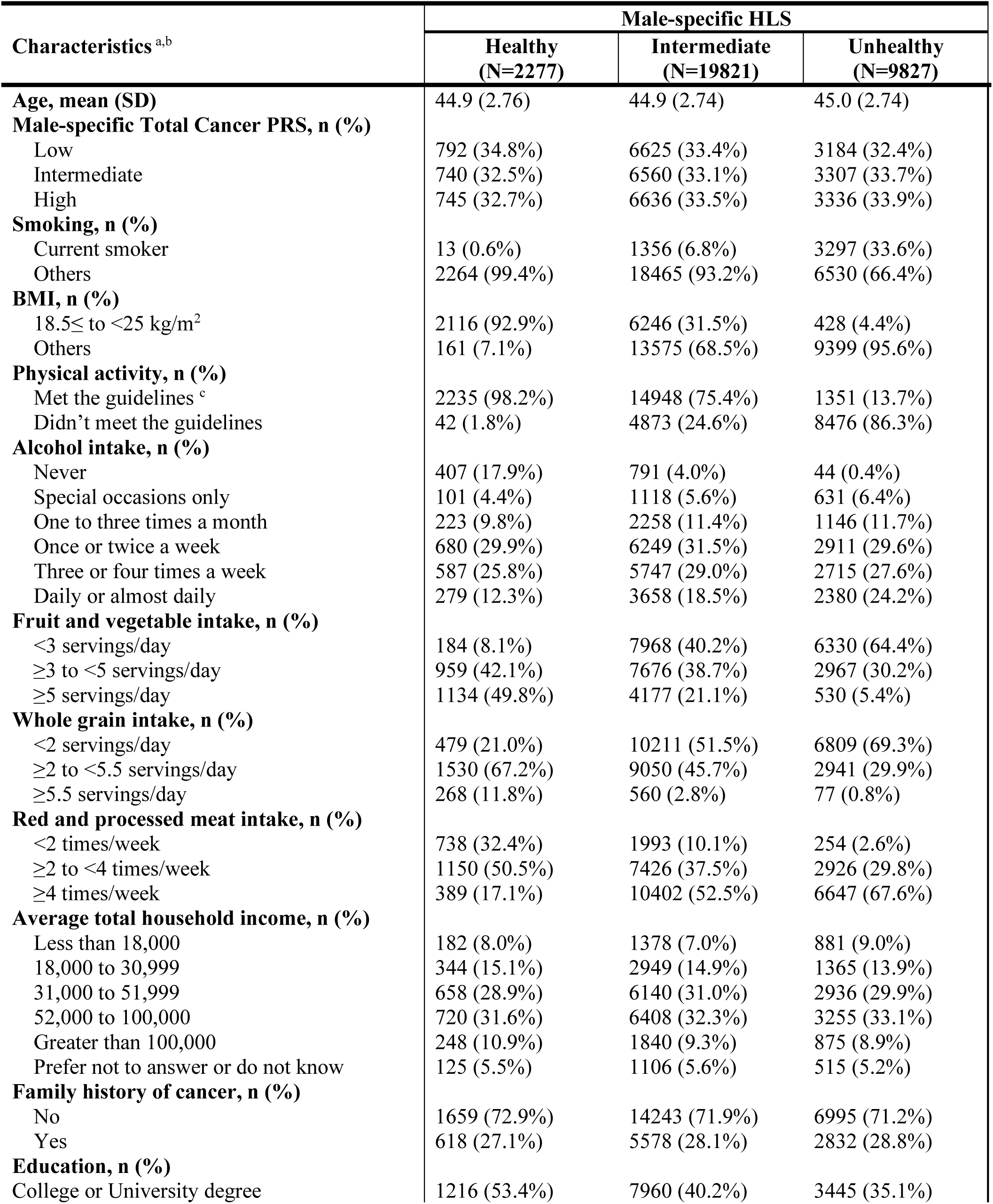
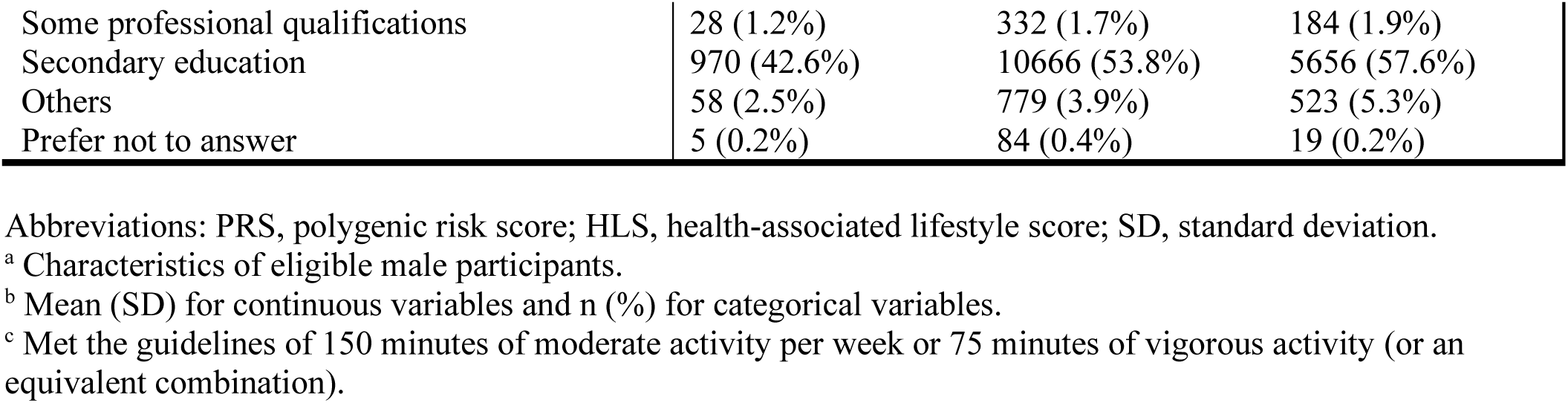
Characteristics of male participants (N=31925) at baseline according to male-specific HLS.

| Characteristics<sup>a,b</sup> | Male-specific HLS |  |  |
| --- | --- | --- | --- |
|  | Healthy<br>(N=2277) | Intermediate<br>(N=19821) | Unhealthy<br>(N=9827) |
| **Age, mean (SD)** | 44.9 (2.76) | 44.9 (2.74) | 45.0 (2.74) |
| **Male-specific Total Cancer PRS, n (%)** |  |  |  |
| Low | 792 (34.8%) | 6625 (33.4%) | 3184 (32.4%) |
| Intermediate | 740 (32.5%) | 6560 (33.1%) | 3307 (33.7%) |
| High | 745 (32.7%) | 6636 (33.5%) | 3336 (33.9%) |
| **Smoking, n (%)** |  |  |  |
| Current smoker | 13 (0.6%) | 1356 (6.8%) | 3297 (33.6%) |
| Others | 2264 (99.4%) | 18465 (93.2%) | 6530 (66.4%) |
| **BMI, n (%)** |  |  |  |
| 18.5≤ to <25 kg/m<sup>2</sup> | 2116 (92.9%) | 6246 (31.5%) | 428 (4.4%) |
| Others | 161 (7.1%) | 13575 (68.5%) | 9399 (95.6%) |
| **Physical activity, n (%)** |  |  |  |
| Met the guidelines<sup>c</sup> | 2235 (98.2%) | 14948 (75.4%) | 1351 (13.7%) |
| Didn't meet the guidelines | 42 (1.8%) | 4873 (24.6%) | 8476 (86.3%) |
| **Alcohol intake, n (%)** |  |  |  |
| Never | 407 (17.9%) | 791 (4.0%) | 44 (0.4%) |
| Special occasions only | 101 (4.4%) | 1118 (5.6%) | 631 (6.4%) |
| One to three times a month | 223 (9.8%) | 2258 (11.4%) | 1146 (11.7%) |
| Once or twice a week | 680 (29.9%) | 6249 (31.5%) | 2911 (29.6%) |
| Three or four times a week | 587 (25.8%) | 5747 (29.0%) | 2715 (27.6%) |
| Daily or almost daily | 279 (12.3%) | 3658 (18.5%) | 2380 (24.2%) |
| **Fruit and vegetable intake, n (%)** |  |  |  |
| <3 servings/day | 184 (8.1%) | 7968 (40.2%) | 6330 (64.4%) |
| ≥3 to <5 servings/day | 959 (42.1%) | 7676 (38.7%) | 2967 (30.2%) |
| ≥5 servings/day | 1134 (49.8%) | 4177 (21.1%) | 530 (5.4%) |
| **Whole grain intake, n (%)** |  |  |  |
| <2 servings/day | 479 (21.0%) | 10211 (51.5%) | 6809 (69.3%) |
| ≥2 to <5.5 servings/day | 1530 (67.2%) | 9050 (45.7%) | 2941 (29.9%) |
| ≥5.5 servings/day | 268 (11.8%) | 560 (2.8%) | 77 (0.8%) |
| **Red and processed meat intake, n (%)** |  |  |  |
| <2 times/week | 738 (32.4%) | 1993 (10.1%) | 254 (2.6%) |
| ≥2 to <4 times/week | 1150 (50.5%) | 7426 (37.5%) | 2926 (29.8%) |
| ≥4 times/week | 389 (17.1%) | 10402 (52.5%) | 6647 (67.6%) |
| **Average total household income, n (%)** |  |  |  |
| Less than 18,000 | 182 (8.0%) | 1378 (7.0%) | 881 (9.0%) |
| 18,000 to 30,999 | 344 (15.1%) | 2949 (14.9%) | 1365 (13.9%) |
| 31,000 to 51,999 | 658 (28.9%) | 6140 (31.0%) | 2936 (29.9%) |
| 52,000 to 100,000 | 720 (31.6%) | 6408 (32.3%) | 3255 (33.1%) |
| Greater than 100,000 | 248 (10.9%) | 1840 (9.3%) | 875 (8.9%) |
| Prefer not to answer or do not know | 125 (5.5%) | 1106 (5.6%) | 515 (5.2%) |
| **Family history of cancer, n (%)** |  |  |  |
| No | 1659 (72.9%) | 14243 (71.9%) | 6995 (71.2%) |
| Yes | 618 (27.1%) | 5578 (28.1%) | 2832 (28.8%) |
| **Education, n (%)** |  |  |  |
| College or University degree | 1216 (53.4%) | 7960 (40.2%) | 3445 (35.1%) |
| Some professional qualifications | 28 (1.2%) | 332 (1.7%) | 184 (1.9%) |
| Secondary education | 970 (42.6%) | 10666 (53.8%) | 5656 (57.6%) |
| Others | 58 (2.5%) | 779 (3.9%) | 523 (5.3%) |
| Prefer not to answer | 5 (0.2%) | 84 (0.4%) | 19 (0.2%) |

Abbreviations: PRS, polygenic risk score; HLS, health-associated lifestyle score; SD, standard deviation.
<sup>a</sup> Characteristics of eligible male participants.
<sup>b</sup> Mean (SD) for continuous variables and n (%) for categorical variables.
<sup>c</sup> Met the guidelines of 150 minutes of moderate activity per week or 75 minutes of vigorous activity (or an equivalent combination).

