## Supplementary material for "Genetic Risk, Health-Associated Lifestyle, and Risk of Early-onset Total Cancer and Breast Cancer": eTables S1-S23

### (Other Supplementary Tables: List of SNPs included in the cancer site-specific PRSs)

**eTable S1**
List of SNPs included in breast cancer PRS

**eTable S2**
List of SNPs included in melanoma PRS

**eTable S3**
List of SNPs included in colorectal cancer PRS

**eTable S4**
List of SNPs included in non-Hodgkin lymphoma PRS

**eTable S5**
List of SNPs included in ovarian cancer PRS

**eTable S6**
List of SNPs included in prostate cancer PRS

**eTable S7**
List of SNPs included in brain cancer PRS

**eTable S8**
List of SNPs included in head and neck cancer PRS

**eTable S9**
List of SNPs included in lung cancer PRS

**eTable S10**
List of SNPs included in endometrial cancer PRS

**eTable S11**
List of SNPs included in testicular cancer PRS

**eTable S12**
List of SNPs included in thyroid cancer PRS

**eTable S13**
List of SNPs included in kidney cancer PRS

**eTable S14**
List of SNPs included in multiple myeloma PRS

**eTable S15**
List of SNPs included in cervical cancer PRS

**eTable S16**
List of SNPs included in pancreatic cancer PRS

**eTable S17**
List of SNPs included in stomach cancer PRS

**eTable S18**
List of SNPs included in lymphoid leukemia PRS

**eTable S19**
List of SNPs included in bladder cancer PRS

**eTable S20**
List of SNPs included in Hodgkin's lymphoma PRS

**eTable S21**
List of SNPs included in larynx cancer PRS

**eTable S22**
List of SNPs included in tracheal cancer PRS

**eTable S23**
List of SNPs included in esophageal cancer PRS

**eTable S1. List of SNPs included in breast cancer PRS**

| Accession | Chr name | Chr position | Effect allele | Other allele | Effect weight | Include |
|---|---|---|---|---|---|---|
| PGS000004_hmPOS_GRCh37 | 1 | 100880328 | T | A | 0.0373 | NO |
| PGS000004_hmPOS_GRCh37 | 1 | 10566215 | G | A | -0.0586 | YES |
| PGS000004_hmPOS_GRCh37 | 1 | 110198129 | C | CAAA | 0.0458 | YES |
| PGS000004_hmPOS_GRCh37 | 1 | 114445880 | A | G | 0.0621 | YES |
| PGS000004_hmPOS_GRCh37 | 1 | 118141492 | C | A | 0.0452 | YES |
| PGS000004_hmPOS_GRCh37 | 1 | 120257110 | C | T | 0.0385 | YES |
| PGS000004_hmPOS_GRCh37 | 1 | 121280613 | G | A | 0.0881 | YES |
| PGS000004_hmPOS_GRCh37 | 1 | 121287994 | G | A | -0.0673 | YES |
| PGS000004_hmPOS_GRCh37 | 1 | 145604302 | CT | C | -0.0399 | YES |
| PGS000004_hmPOS_GRCh37 | 1 | 149906413 | C | T | 0.0548 | YES |
| PGS000004_hmPOS_GRCh37 | 1 | 155556971 | A | G | 0.0499 | YES |
| PGS000004_hmPOS_GRCh37 | 1 | 168171052 | C | CA | -0.068 | YES |
| PGS000004_hmPOS_GRCh37 | 1 | 172328767 | TA | T | -0.0435 | YES |
| PGS000004_hmPOS_GRCh37 | 1 | 18807339 | C | T | -0.0564 | YES |
| PGS000004_hmPOS_GRCh37 | 1 | 201437832 | T | C | 0.0917 | YES |
| PGS000004_hmPOS_GRCh37 | 1 | 202184600 | T | C | -0.0065 | YES |
| PGS000004_hmPOS_GRCh37 | 1 | 203770448 | A | T | 0.0498 | NO |
| PGS000004_hmPOS_GRCh37 | 1 | 204502514 | TTCTGAAACAGGG | T | -0.0321 | YES |
| PGS000004_hmPOS_GRCh37 | 1 | 208076291 | A | G | -0.0366 | YES |
| PGS000004_hmPOS_GRCh37 | 1 | 217053815 | G | T | 0.0417 | YES |
| PGS000004_hmPOS_GRCh37 | 1 | 217220574 | A | G | -0.044 | YES |
| PGS000004_hmPOS_GRCh37 | 1 | 220671050 | T | C | 0.0418 | YES |
| PGS000004_hmPOS_GRCh37 | 1 | 242034263 | G | A | 0.1428 | YES |
| PGS000004_hmPOS_GRCh37 | 1 | 41380440 | T | C | 0.0426 | YES |
| PGS000004_hmPOS_GRCh37 | 1 | 41389220 | C | T | 0.155 | YES |
| PGS000004_hmPOS_GRCh37 | 1 | 46670206 | T | TC | 0.0447 | YES |
| PGS000004_hmPOS_GRCh37 | 1 | 51467096 | C | CT | 0.0374 | YES |
| PGS000004_hmPOS_GRCh37 | 1 | 7917076 | A | G | -0.0409 | YES |
| PGS000004_hmPOS_GRCh37 | 1 | 88156923 | A | G | 0.0494 | YES |
| PGS000004_hmPOS_GRCh37 | 1 | 88428199 | A | C | -0.0387 | YES |
| PGS000004_hmPOS_GRCh37 | 2 | 10138983 | C | T | 0.0603 | YES |
| PGS000004_hmPOS_GRCh37 | 2 | 121058254 | G | A | -0.0334 | YES |
| PGS000004_hmPOS_GRCh37 | 2 | 121089731 | C | T | -0.0427 | YES |
| PGS000004_hmPOS_GRCh37 | 2 | 121159205 | A | G | -0.044 | YES |
| PGS000004_hmPOS_GRCh37 | 2 | 121246568 | C | T | 0.0992 | YES |
| PGS000004_hmPOS_GRCh37 | 2 | 172974566 | G | C | -0.0473 | NO |
| PGS000004_hmPOS_GRCh37 | 2 | 172974566 | G | C | -0.0473 | NO |
| PGS000004_hmPOS_GRCh37 | 2 | 174212910 | G | A | 0.0593 | YES |
| PGS000004_hmPOS_GRCh37 | 2 | 192381934 | T | C | 0.0316 | YES |
| PGS000004_hmPOS_GRCh37 | 2 | 19315675 | A | T | -0.0331 | NO |
| PGS000004_hmPOS_GRCh37 | 2 | 202204741 | C | T | -0.0492 | YES |
| PGS000004_hmPOS_GRCh37 | 2 | 217920769 | T | G | -0.1318 | YES |
| PGS000004_hmPOS_GRCh37 | 2 | 217955896 | G | GA | -0.2016 | YES |
| PGS000004_hmPOS_GRCh37 | 2 | 218292158 | G | C | -0.0757 | NO |
| PGS000004_hmPOS_GRCh37 | 2 | 218714845 | A | G | -0.0431 | YES |
| PGS000004_hmPOS_GRCh37 | 2 | 241388857 | A | C | -0.1232 | YES |

| | | | | | | |
|---|---|---|---|---|---|---|
| PGS000004_hmPOS_GRCh37 | 2 | 25129473 | G | A | -0.0427 | YES |
| PGS000004_hmPOS_GRCh37 | 2 | 29179452 | C | G | -0.0066 | NO |
| PGS000004_hmPOS_GRCh37 | 2 | 29615233 | C | T | -0.0427 | YES |
| PGS000004_hmPOS_GRCh37 | 2 | 39699510 | CT | C | -0.0402 | YES |
| PGS000004_hmPOS_GRCh37 | 2 | 70172587 | A | G | -0.0412 | YES |
| PGS000004_hmPOS_GRCh37 | 2 | 88358825 | C | G | 0.0473 | NO |
| PGS000004_hmPOS_GRCh37 | 3 | 141112859 | C | CTT | 0.0551 | YES |
| PGS000004_hmPOS_GRCh37 | 3 | 172285237 | A | G | 0.0422 | YES |
| PGS000004_hmPOS_GRCh37 | 3 | 189774456 | T | C | -0.0478 | YES |
| PGS000004_hmPOS_GRCh37 | 3 | 27353716 | A | C | 0.0748 | YES |
| PGS000004_hmPOS_GRCh37 | 3 | 27388664 | G | C | 0.0502 | NO |
| PGS000004_hmPOS_GRCh37 | 3 | 29294845 | T | C | -0.1281 | YES |
| PGS000004_hmPOS_GRCh37 | 3 | 30684907 | T | C | 0.0592 | YES |
| PGS000004_hmPOS_GRCh37 | 3 | 46888198 | C | T | -0.0806 | YES |
| PGS000004_hmPOS_GRCh37 | 3 | 4742251 | G | A | 0.0616 | YES |
| PGS000004_hmPOS_GRCh37 | 3 | 49709912 | CT | C | -0.0367 | YES |
| PGS000004_hmPOS_GRCh37 | 3 | 55970777 | AT | A | -0.1195 | YES |
| PGS000004_hmPOS_GRCh37 | 3 | 59373745 | T | C | -0.0394 | YES |
| PGS000004_hmPOS_GRCh37 | 3 | 63887449 | TTG | T | 0.0648 | NO |
| PGS000004_hmPOS_GRCh37 | 3 | 71620370 | G | T | -0.0374 | YES |
| PGS000004_hmPOS_GRCh37 | 3 | 87037543 | G | A | -0.0723 | YES |
| PGS000004_hmPOS_GRCh37 | 3 | 99403877 | A | G | -0.0376 | YES |
| PGS000004_hmPOS_GRCh37 | 4 | 106069013 | T | G | 0.0471 | YES |
| PGS000004_hmPOS_GRCh37 | 4 | 126752992 | AAT | A | -0.0377 | NO |
| PGS000004_hmPOS_GRCh37 | 4 | 143467195 | T | C | -0.0569 | YES |
| PGS000004_hmPOS_GRCh37 | 4 | 151218296 | C | CATATTT | 0.0388 | YES |
| PGS000004_hmPOS_GRCh37 | 4 | 175842495 | A | G | -0.0898 | YES |
| PGS000004_hmPOS_GRCh37 | 4 | 175847436 | A | C | 0.0348 | YES |
| PGS000004_hmPOS_GRCh37 | 4 | 187503758 | T | A | 0.0357 | NO |
| PGS000004_hmPOS_GRCh37 | 4 | 38784633 | T | G | 0.0489 | YES |
| PGS000004_hmPOS_GRCh37 | 4 | 84370124 | TA | TAA | -0.0464 | NO |
| PGS000004_hmPOS_GRCh37 | 4 | 89240476 | A | G | 0.0352 | YES |
| PGS000004_hmPOS_GRCh37 | 4 | 92594859 | T | TTCTTTC | -0.0407 | YES |
| PGS000004_hmPOS_GRCh37 | 5 | 104300273 | T | G | -0.0487 | YES |
| PGS000004_hmPOS_GRCh37 | 5 | 122478676 | A | C | -0.0386 | YES |
| PGS000004_hmPOS_GRCh37 | 5 | 122705244 | T | C | 0.0944 | YES |
| PGS000004_hmPOS_GRCh37 | 5 | 1279790 | T | C | 0.0617 | YES |
| PGS000004_hmPOS_GRCh37 | 5 | 1296255 | AG | A | -0.0549 | YES |
| PGS000004_hmPOS_GRCh37 | 5 | 131640536 | G | A | 0.0392 | YES |
| PGS000004_hmPOS_GRCh37 | 5 | 132407058 | T | C | -0.0388 | YES |
| PGS000004_hmPOS_GRCh37 | 5 | 1353077 | C | T | 0.1552 | YES |
| PGS000004_hmPOS_GRCh37 | 5 | 158244083 | T | C | -0.0677 | YES |
| PGS000004_hmPOS_GRCh37 | 5 | 16231194 | C | G | -0.0426 | NO |
| PGS000004_hmPOS_GRCh37 | 5 | 169591460 | C | T | 0.0412 | YES |
| PGS000004_hmPOS_GRCh37 | 5 | 173358154 | A | G | 0.0365 | YES |
| PGS000004_hmPOS_GRCh37 | 5 | 176134882 | C | T | 0.0363 | YES |
| PGS000004_hmPOS_GRCh37 | 5 | 2777029 | A | G | 0.0391 | YES |
| PGS000004_hmPOS_GRCh37 | 5 | 32579616 | T | TCA | 0.0363 | YES |
| PGS000004_hmPOS_GRCh37 | 5 | 345109 | C | T | 0.084 | YES |

| | | | | | | |
|---|---|---|---|---|---|---|
| PGS000004_hmPOS_GRCh37 | 5 | 44508264 | GT | G | -0.1177 | YES |
| PGS000004_hmPOS_GRCh37 | 5 | 44619502 | G | A | -0.1101 | YES |
| PGS000004_hmPOS_GRCh37 | 5 | 44649944 | T | C | 0.0492 | YES |
| PGS000004_hmPOS_GRCh37 | 5 | 44706498 | G | A | 0.0497 | YES |
| PGS000004_hmPOS_GRCh37 | 5 | 44853593 | C | G | -0.0336 | NO |
| PGS000004_hmPOS_GRCh37 | 5 | 52679539 | CA | C | 0.0571 | NO |
| PGS000004_hmPOS_GRCh37 | 5 | 55662540 | CT | C | -0.0458 | YES |
| PGS000004_hmPOS_GRCh37 | 5 | 55965167 | T | C | 0.0394 | YES |
| PGS000004_hmPOS_GRCh37 | 5 | 56023083 | G | T | 0.1366 | YES |
| PGS000004_hmPOS_GRCh37 | 5 | 56042972 | T | C | 0.0865 | YES |
| PGS000004_hmPOS_GRCh37 | 5 | 56045081 | C | T | -0.0564 | YES |
| PGS000004_hmPOS_GRCh37 | 5 | 58241712 | T | C | -0.0434 | YES |
| PGS000004_hmPOS_GRCh37 | 5 | 71965007 | A | G | -0.041 | YES |
| PGS000004_hmPOS_GRCh37 | 5 | 73234583 | C | T | -0.0363 | YES |
| PGS000004_hmPOS_GRCh37 | 5 | 77155397 | G | GT | -0.0408 | YES |
| PGS000004_hmPOS_GRCh37 | 5 | 79180995 | GA | G | 0.0328 | YES |
| PGS000004_hmPOS_GRCh37 | 5 | 81512947 | T | TA | -0.0598 | YES |
| PGS000004_hmPOS_GRCh37 | 5 | 90789470 | A | G | -0.0564 | YES |
| PGS000004_hmPOS_GRCh37 | 6 | 130341728 | CT | C | 0.0472 | YES |
| PGS000004_hmPOS_GRCh37 | 6 | 13713366 | C | G | -0.0553 | NO |
| PGS000004_hmPOS_GRCh37 | 6 | 149595505 | C | T | -0.0476 | YES |
| PGS000004_hmPOS_GRCh37 | 6 | 151949806 | C | A | 0.0703 | YES |
| PGS000004_hmPOS_GRCh37 | 6 | 151955914 | G | A | 0.1449 | YES |
| PGS000004_hmPOS_GRCh37 | 6 | 152022664 | C | CAAAAAAA | 0.0137 | YES |
| PGS000004_hmPOS_GRCh37 | 6 | 152023191 | A | G | 0.0626 | YES |
| PGS000004_hmPOS_GRCh37 | 6 | 152055978 | T | A | 0.074 | NO |
| PGS000004_hmPOS_GRCh37 | 6 | 152432902 | T | C | 0.0649 | YES |
| PGS000004_hmPOS_GRCh37 | 6 | 16399557 | T | C | -0.0373 | YES |
| PGS000004_hmPOS_GRCh37 | 6 | 169006947 | G | C | -0.0308 | NO |
| PGS000004_hmPOS_GRCh37 | 6 | 170332621 | C | T | 0.0373 | YES |
| PGS000004_hmPOS_GRCh37 | 6 | 18783140 | A | G | 0.0326 | YES |
| PGS000004_hmPOS_GRCh37 | 6 | 20537845 | C | CA | -0.0391 | YES |
| PGS000004_hmPOS_GRCh37 | 6 | 21923810 | C | T | -0.0321 | YES |
| PGS000004_hmPOS_GRCh37 | 6 | 27425644 | C | G | -0.0737 | NO |
| PGS000004_hmPOS_GRCh37 | 6 | 43227141 | A | G | -0.064 | YES |
| PGS000004_hmPOS_GRCh37 | 6 | 82263549 | A | AAT | 0.0477 | YES |
| PGS000004_hmPOS_GRCh37 | 6 | 85912194 | C | CAA | 0.0762 | YES |
| PGS000004_hmPOS_GRCh37 | 6 | 87803819 | C | T | 0.0383 | YES |
| PGS000004_hmPOS_GRCh37 | 7 | 101552440 | A | G | -0.0568 | YES |
| PGS000004_hmPOS_GRCh37 | 7 | 102481842 | C | T | 0.0418 | YES |
| PGS000004_hmPOS_GRCh37 | 7 | 130656911 | T | C | -0.0476 | YES |
| PGS000004_hmPOS_GRCh37 | 7 | 130674481 | A | G | 0.0416 | YES |
| PGS000004_hmPOS_GRCh37 | 7 | 139943702 | C | CT | 0.0582 | YES |
| PGS000004_hmPOS_GRCh37 | 7 | 144048902 | T | G | -0.0563 | YES |
| PGS000004_hmPOS_GRCh37 | 7 | 21940960 | G | A | -0.0467 | YES |
| PGS000004_hmPOS_GRCh37 | 7 | 25569548 | T | C | -0.0486 | YES |
| PGS000004_hmPOS_GRCh37 | 7 | 28869017 | A | G | -0.0572 | YES |
| PGS000004_hmPOS_GRCh37 | 7 | 55192256 | C | A | -0.0349 | YES |
| PGS000004_hmPOS_GRCh37 | 7 | 91459189 | ATT | A | 0.0452 | NO |

| | | | | | | |
|---|---|---|---|---|---|---|
| PGS000004_hmPOS_GRCh37 | 7 | 94113799 | C | T | 0.0449 | YES |
| PGS000004_hmPOS_GRCh37 | 7 | 98005235 | A | G | -0.0467 | YES |
| PGS000004_hmPOS_GRCh37 | 7 | 99948655 | G | T | 0.042 | YES |
| PGS000004_hmPOS_GRCh37 | 8 | 102483100 | C | T | 0.0593 | YES |
| PGS000004_hmPOS_GRCh37 | 8 | 106358620 | T | A | -0.0745 | NO |
| PGS000004_hmPOS_GRCh37 | 8 | 117209548 | G | A | -0.0417 | YES |
| PGS000004_hmPOS_GRCh37 | 8 | 120862186 | G | A | 0.0527 | YES |
| PGS000004_hmPOS_GRCh37 | 8 | 124563705 | C | T | 0.0477 | YES |
| PGS000004_hmPOS_GRCh37 | 8 | 124571581 | A | G | 0.034 | YES |
| PGS000004_hmPOS_GRCh37 | 8 | 124739913 | G | T | 0.0466 | YES |
| PGS000004_hmPOS_GRCh37 | 8 | 128213561 | CA | C | -0.043 | YES |
| PGS000004_hmPOS_GRCh37 | 8 | 128370949 | G | C | 0.0642 | NO |
| PGS000004_hmPOS_GRCh37 | 8 | 128372172 | G | A | 0.0597 | YES |
| PGS000004_hmPOS_GRCh37 | 8 | 129199566 | A | G | 0.0615 | YES |
| PGS000004_hmPOS_GRCh37 | 8 | 143669254 | G | A | -0.0346 | YES |
| PGS000004_hmPOS_GRCh37 | 8 | 170692 | C | T | 0.0477 | YES |
| PGS000004_hmPOS_GRCh37 | 8 | 17787610 | C | CT | -0.0377 | YES |
| PGS000004_hmPOS_GRCh37 | 8 | 23447496 | G | A | -0.0389 | YES |
| PGS000004_hmPOS_GRCh37 | 8 | 23663653 | A | C | 0.0335 | YES |
| PGS000004_hmPOS_GRCh37 | 8 | 29509616 | C | A | -0.0601 | YES |
| PGS000004_hmPOS_GRCh37 | 8 | 36858483 | G | A | -0.076 | YES |
| PGS000004_hmPOS_GRCh37 | 8 | 76230943 | G | A | 0.0755 | YES |
| PGS000004_hmPOS_GRCh37 | 8 | 76333056 | T | C | 0.1129 | YES |
| PGS000004_hmPOS_GRCh37 | 8 | 76378165 | T | G | -0.0391 | YES |
| PGS000004_hmPOS_GRCh37 | 9 | 110303808 | T | TAA | 0.0797 | YES |
| PGS000004_hmPOS_GRCh37 | 9 | 110837073 | G | A | 0.1158 | YES |
| PGS000004_hmPOS_GRCh37 | 9 | 110837176 | T | C | 0.0653 | YES |
| PGS000004_hmPOS_GRCh37 | 9 | 110849525 | T | G | 0.0153 | YES |
| PGS000004_hmPOS_GRCh37 | 9 | 110885479 | T | C | 0.0877 | YES |
| PGS000004_hmPOS_GRCh37 | 9 | 119313486 | G | A | -0.0462 | YES |
| PGS000004_hmPOS_GRCh37 | 9 | 129424719 | G | A | -0.0382 | YES |
| PGS000004_hmPOS_GRCh37 | 9 | 136146597 | T | C | 0.04 | YES |
| PGS000004_hmPOS_GRCh37 | 9 | 21964882 | C | CAAAA | 0.055 | YES |
| PGS000004_hmPOS_GRCh37 | 9 | 22041998 | G | C | 0.0289 | NO |
| PGS000004_hmPOS_GRCh37 | 9 | 36928288 | C | T | 0.0249 | YES |
| PGS000004_hmPOS_GRCh37 | 9 | 6880263 | G | A | 0.0348 | YES |
| PGS000004_hmPOS_GRCh37 | 9 | 87782211 | C | T | 0.0361 | YES |
| PGS000004_hmPOS_GRCh37 | 9 | 98362587 | C | T | 0.0576 | YES |
| PGS000004_hmPOS_GRCh37 | 10 | 114777670 | T | C | 0.0472 | YES |
| PGS000004_hmPOS_GRCh37 | 10 | 115128491 | C | T | -0.0592 | YES |
| PGS000004_hmPOS_GRCh37 | 10 | 123095209 | A | G | -0.0538 | YES |
| PGS000004_hmPOS_GRCh37 | 10 | 123340107 | G | A | 0.1508 | YES |
| PGS000004_hmPOS_GRCh37 | 10 | 123340431 | G | GC | -0.2408 | YES |
| PGS000004_hmPOS_GRCh37 | 10 | 123349324 | T | A | -0.2609 | NO |
| PGS000004_hmPOS_GRCh37 | 10 | 13892298 | A | G | 0.0371 | YES |
| PGS000004_hmPOS_GRCh37 | 10 | 22032942 | G | A | -0.058 | YES |
| PGS000004_hmPOS_GRCh37 | 10 | 22477776 | A | ACC | 0.1687 | YES |
| PGS000004_hmPOS_GRCh37 | 10 | 22861490 | C | A | 0.0875 | NO |
| PGS000004_hmPOS_GRCh37 | 10 | 38523626 | A | C | 0.0404 | YES |

| | | | | | | |
|---|---|---|---|---|---|---|
| PGS000004_hmPOS_GRCh37 | 10 | 5794652 | G | A | 0.047 | YES |
| PGS000004_hmPOS_GRCh37 | 10 | 64299890 | G | A | -0.1345 | YES |
| PGS000004_hmPOS_GRCh37 | 10 | 64819996 | T | G | 0.0472 | YES |
| PGS000004_hmPOS_GRCh37 | 10 | 71335574 | T | C | -0.0404 | YES |
| PGS000004_hmPOS_GRCh37 | 10 | 80851257 | T | G | -0.0805 | YES |
| PGS000004_hmPOS_GRCh37 | 10 | 80886726 | G | A | 0.0762 | YES |
| PGS000004_hmPOS_GRCh37 | 10 | 95292187 | C | CAA | -0.0512 | YES |
| PGS000004_hmPOS_GRCh37 | 11 | 103614438 | G | T | 0.0147 | YES |
| PGS000004_hmPOS_GRCh37 | 11 | 108267402 | CA | C | -0.0022 | YES |
| PGS000004_hmPOS_GRCh37 | 11 | 111696440 | C | T | -0.0396 | YES |
| PGS000004_hmPOS_GRCh37 | 11 | 116727936 | T | A | -0.0423 | NO |
| PGS000004_hmPOS_GRCh37 | 11 | 122966626 | G | A | -0.0383 | YES |
| PGS000004_hmPOS_GRCh37 | 11 | 129243417 | G | T | -0.0543 | YES |
| PGS000004_hmPOS_GRCh37 | 11 | 129461016 | G | A | 0.0453 | YES |
| PGS000004_hmPOS_GRCh37 | 11 | 18664241 | G | T | 0.0461 | YES |
| PGS000004_hmPOS_GRCh37 | 11 | 1895708 | A | C | -0.0762 | YES |
| PGS000004_hmPOS_GRCh37 | 11 | 42844441 | T | C | -0.0336 | YES |
| PGS000004_hmPOS_GRCh37 | 11 | 433617 | C | T | -0.0437 | YES |
| PGS000004_hmPOS_GRCh37 | 11 | 44368892 | A | G | 0.0374 | YES |
| PGS000004_hmPOS_GRCh37 | 11 | 46318032 | G | C | -0.0748 | NO |
| PGS000004_hmPOS_GRCh37 | 11 | 65553492 | A | C | 0.0425 | YES |
| PGS000004_hmPOS_GRCh37 | 11 | 65572431 | A | G | -0.0347 | YES |
| PGS000004_hmPOS_GRCh37 | 11 | 69328130 | T | A | -0.0423 | NO |
| PGS000004_hmPOS_GRCh37 | 11 | 69330983 | A | G | 0.1022 | YES |
| PGS000004_hmPOS_GRCh37 | 11 | 69331418 | T | C | 0.1782 | YES |
| PGS000004_hmPOS_GRCh37 | 11 | 803017 | G | A | 0.0457 | YES |
| PGS000004_hmPOS_GRCh37 | 12 | 103097887 | T | C | 0.0546 | YES |
| PGS000004_hmPOS_GRCh37 | 12 | 111600134 | T | G | -0.0442 | YES |
| PGS000004_hmPOS_GRCh37 | 12 | 115108136 | C | T | 0.0465 | YES |
| PGS000004_hmPOS_GRCh37 | 12 | 115796577 | G | A | -0.0428 | YES |
| PGS000004_hmPOS_GRCh37 | 12 | 115835836 | C | T | -0.0813 | YES |
| PGS000004_hmPOS_GRCh37 | 12 | 120832146 | T | C | 0.0516 | YES |
| PGS000004_hmPOS_GRCh37 | 12 | 14413931 | C | G | 0.0484 | NO |
| PGS000004_hmPOS_GRCh37 | 12 | 28149568 | T | C | -0.062 | YES |
| PGS000004_hmPOS_GRCh37 | 12 | 28174817 | T | C | -0.0856 | YES |
| PGS000004_hmPOS_GRCh37 | 12 | 28347382 | T | C | -0.0521 | YES |
| PGS000004_hmPOS_GRCh37 | 12 | 29140260 | A | G | 0.0647 | YES |
| PGS000004_hmPOS_GRCh37 | 12 | 293626 | G | A | 0.0401 | YES |
| PGS000004_hmPOS_GRCh37 | 12 | 57146069 | G | T | -0.0579 | YES |
| PGS000004_hmPOS_GRCh37 | 12 | 70798355 | T | A | 0.0469 | NO |
| PGS000004_hmPOS_GRCh37 | 12 | 83064195 | GA | G | 0.0671 | YES |
| PGS000004_hmPOS_GRCh37 | 12 | 85004551 | T | C | 0.0348 | YES |
| PGS000004_hmPOS_GRCh37 | 12 | 96027759 | G | A | -0.0867 | YES |
| PGS000004_hmPOS_GRCh37 | 13 | 32839990 | A | G | 0.0424 | YES |
| PGS000004_hmPOS_GRCh37 | 13 | 32972626 | T | A | 0.2687 | NO |
| PGS000004_hmPOS_GRCh37 | 13 | 43501356 | G | A | 0.0517 | YES |
| PGS000004_hmPOS_GRCh37 | 13 | 73806982 | C | T | 0.0345 | YES |
| PGS000004_hmPOS_GRCh37 | 13 | 73960952 | G | A | 0.0399 | YES |
| PGS000004_hmPOS_GRCh37 | 14 | 105213978 | G | T | 0.0399 | YES |

| | | | | | | |
|---|---|---|---|---|---|---|
| PGS000004_hmPOS_GRCh37 | 14 | 37128564 | A | C | -0.0733 | YES |
| PGS000004_hmPOS_GRCh37 | 14 | 37228504 | T | C | 0.039 | YES |
| PGS000004_hmPOS_GRCh37 | 14 | 68660428 | C | T | -0.0474 | YES |
| PGS000004_hmPOS_GRCh37 | 14 | 68979835 | C | T | -0.0911 | YES |
| PGS000004_hmPOS_GRCh37 | 14 | 91751788 | T | TC | 0.038 | YES |
| PGS000004_hmPOS_GRCh37 | 14 | 91841069 | G | A | 0.0513 | YES |
| PGS000004_hmPOS_GRCh37 | 14 | 93070286 | T | C | -0.0577 | YES |
| PGS000004_hmPOS_GRCh37 | 15 | 100905819 | C | A | -0.0608 | YES |
| PGS000004_hmPOS_GRCh37 | 15 | 46680811 | A | C | -0.1973 | YES |
| PGS000004_hmPOS_GRCh37 | 15 | 50694306 | G | A | -0.0417 | YES |
| PGS000004_hmPOS_GRCh37 | 15 | 66630569 | A | G | -0.0369 | YES |
| PGS000004_hmPOS_GRCh37 | 15 | 67457698 | G | A | 0.0782 | YES |
| PGS000004_hmPOS_GRCh37 | 15 | 75750383 | C | T | -0.0413 | YES |
| PGS000004_hmPOS_GRCh37 | 15 | 91512267 | T | G | -0.0589 | YES |
| PGS000004_hmPOS_GRCh37 | 16 | 10706580 | A | G | -0.074 | YES |
| PGS000004_hmPOS_GRCh37 | 16 | 23007047 | T | G | 0.1218 | YES |
| PGS000004_hmPOS_GRCh37 | 16 | 4008542 | C | CAAAAA | -0.0329 | YES |
| PGS000004_hmPOS_GRCh37 | 16 | 4106788 | A | C | -0.03 | YES |
| PGS000004_hmPOS_GRCh37 | 16 | 52538825 | A | C | 0.1147 | YES |
| PGS000004_hmPOS_GRCh37 | 16 | 52599188 | T | C | 0.107 | YES |
| PGS000004_hmPOS_GRCh37 | 16 | 53809123 | T | C | -0.0704 | YES |
| PGS000004_hmPOS_GRCh37 | 16 | 53861139 | T | C | -0.0338 | YES |
| PGS000004_hmPOS_GRCh37 | 16 | 53861592 | A | G | -0.0337 | YES |
| PGS000004_hmPOS_GRCh37 | 16 | 54682064 | A | G | 0.0477 | YES |
| PGS000004_hmPOS_GRCh37 | 16 | 6963972 | G | C | 0.0354 | NO |
| PGS000004_hmPOS_GRCh37 | 16 | 80648296 | G | A | 0.0839 | YES |
| PGS000004_hmPOS_GRCh37 | 16 | 85145977 | C | T | -0.0211 | YES |
| PGS000004_hmPOS_GRCh37 | 16 | 87086492 | C | T | -0.0469 | YES |
| PGS000004_hmPOS_GRCh37 | 17 | 29168077 | T | G | -0.0568 | YES |
| PGS000004_hmPOS_GRCh37 | 17 | 39251123 | C | T | 0.0799 | YES |
| PGS000004_hmPOS_GRCh37 | 17 | 40127060 | C | T | 0.0174 | YES |
| PGS000004_hmPOS_GRCh37 | 17 | 40485239 | T | G | -0.0571 | YES |
| PGS000004_hmPOS_GRCh37 | 17 | 40744470 | A | G | 0.2017 | YES |
| PGS000004_hmPOS_GRCh37 | 17 | 43212339 | CT | C | 0.0438 | YES |
| PGS000004_hmPOS_GRCh37 | 17 | 44283858 | A | G | -0.054 | YES |
| PGS000004_hmPOS_GRCh37 | 17 | 53209774 | C | A | -0.0793 | YES |
| PGS000004_hmPOS_GRCh37 | 17 | 77781725 | G | A | -0.0401 | YES |
| PGS000004_hmPOS_GRCh37 | 18 | 11696613 | T | C | -0.0381 | YES |
| PGS000004_hmPOS_GRCh37 | 18 | 20634253 | T | C | -0.0415 | YES |
| PGS000004_hmPOS_GRCh37 | 18 | 24125857 | C | T | 0.0346 | YES |
| PGS000004_hmPOS_GRCh37 | 18 | 24337424 | G | C | 0.0455 | NO |
| PGS000004_hmPOS_GRCh37 | 18 | 24518050 | A | AT | -0.0599 | YES |
| PGS000004_hmPOS_GRCh37 | 18 | 25407513 | G | C | 0.0399 | NO |
| PGS000004_hmPOS_GRCh37 | 18 | 29981526 | A | G | -0.1058 | YES |
| PGS000004_hmPOS_GRCh37 | 18 | 42411803 | C | G | -0.0877 | NO |
| PGS000004_hmPOS_GRCh37 | 18 | 42888797 | C | T | -0.0542 | YES |
| PGS000004_hmPOS_GRCh37 | 19 | 13249921 | T | G | 0.0956 | YES |
| PGS000004_hmPOS_GRCh37 | 19 | 17393925 | A | C | 0.0378 | YES |
| PGS000004_hmPOS_GRCh37 | 19 | 18569492 | T | C | -0.0719 | YES |

| | | | | | | |
|---|---|---|---|---|---|---|
| PGS000004_hmPOS_GRCh37 | 19 | 19517054 | CGGGCG | C | 0.0437 | YES |
| PGS000004_hmPOS_GRCh37 | 19 | 44283031 | C | T | 0.0619 | YES |
| PGS000004_hmPOS_GRCh37 | 19 | 46166073 | C | T | -0.036 | YES |
| PGS000004_hmPOS_GRCh37 | 19 | 55816678 | T | C | -0.0359 | YES |
| PGS000004_hmPOS_GRCh37 | 20 | 11379842 | C | T | 0.0844 | YES |
| PGS000004_hmPOS_GRCh37 | 20 | 41613706 | G | C | 0.0315 | NO |
| PGS000004_hmPOS_GRCh37 | 20 | 52296849 | A | G | 0.044 | YES |
| PGS000004_hmPOS_GRCh37 | 20 | 5948227 | A | G | 0.076 | YES |
| PGS000004_hmPOS_GRCh37 | 21 | 16364756 | G | T | 0.0646 | YES |
| PGS000004_hmPOS_GRCh37 | 21 | 16566350 | G | A | 0.0595 | YES |
| PGS000004_hmPOS_GRCh37 | 21 | 16574455 | A | C | -0.0707 | YES |
| PGS000004_hmPOS_GRCh37 | 21 | 47762932 | A | G | 0.0946 | YES |
| PGS000004_hmPOS_GRCh37 | 22 | 19766137 | T | C | -0.0367 | YES |
| PGS000004_hmPOS_GRCh37 | 22 | 29121087 | G | A | 0.1839 | YES |
| PGS000004_hmPOS_GRCh37 | 22 | 29135543 | A | G | 0.0654 | YES |
| PGS000004_hmPOS_GRCh37 | 22 | 29203724 | T | C | 0.1405 | YES |
| PGS000004_hmPOS_GRCh37 | 22 | 29551872 | G | A | -0.1716 | YES |
| PGS000004_hmPOS_GRCh37 | 22 | 38583315 | AAAAGAAAG | AAAAG | -0.0471 | NO |
| PGS000004_hmPOS_GRCh37 | 22 | 39343916 | A | T | 0.0407 | NO |
| PGS000004_hmPOS_GRCh37 | 22 | 40904707 | C | CT | 0.1148 | YES |
| PGS000004_hmPOS_GRCh37 | 22 | 43433100 | T | C | -0.06 | YES |
| PGS000004_hmPOS_GRCh37 | 22 | 45319953 | A | G | -0.0134 | YES |
| PGS000004_hmPOS_GRCh37 | 22 | 46283297 | A | G | 0.0736 | YES |

Abbreviations: SNP, Single nucleotide polymorphism; PRS, polygenic risk score.

Total number of variants: 313
Matched, n (%): 275 (87.86%)
Exclusions due to ambiguously matched, n (%): 30 (9.58%)
Exclusions due to unmatched, n (%): 8 (2.56%)

**eTable S2. List of SNPs included in melanoma PRS**

| Accession | Chr name | Chr position | Effect allele | Other allele | Effect weight | Include |
|---|---|---|---|---|---|---|
| PGS000790_hmPOS_GRCh37 | 1 | 150860471 | T | C | 6.48 | YES |
| PGS000790_hmPOS_GRCh37 | 2 | 38276549 | A | G | 6.13 | YES |
| PGS000790_hmPOS_GRCh37 | 2 | 202162811 | A | G | 6.13 | YES |
| PGS000790_hmPOS_GRCh37 | 5 | 1322087 | A | G | 5.54 | YES |
| PGS000790_hmPOS_GRCh37 | 5 | 33946571 | G | A | 9.64 | YES |
| PGS000790_hmPOS_GRCh37 | 6 | 421281 | A | G | 7.45 | YES |
| PGS000790_hmPOS_GRCh37 | 6 | 21163919 | C | T | 5.57 | YES |
| PGS000790_hmPOS_GRCh37 | 7 | 16984280 | T | C | 6.01 | YES |
| PGS000790_hmPOS_GRCh37 | 9 | 21803880 | C | A | 8.47 | YES |
| PGS000790_hmPOS_GRCh37 | 9 | 109060830 | T | C | 6.52 | YES |
| PGS000790_hmPOS_GRCh37 | 10 | 105668843 | G | A | 5.98 | YES |
| PGS000790_hmPOS_GRCh37 | 11 | 69367118 | A | C | 7.16 | YES |
| PGS000790_hmPOS_GRCh37 | 11 | 89011046 | A | G | 7.63 | YES |
| PGS000790_hmPOS_GRCh37 | 11 | 108175462 | G | A | 5.91 | YES |
| PGS000790_hmPOS_GRCh37 | 15 | 28335820 | A | G | 6.69 | YES |
| PGS000790_hmPOS_GRCh37 | 16 | 54114824 | A | G | 6.95 | YES |
| PGS000790_hmPOS_GRCh37 | 16 | 89726484 | A | G | 7.11 | YES |
| PGS000790_hmPOS_GRCh37 | 16 | 89986117 | T | C | 12.73 | YES |
| PGS000790_hmPOS_GRCh37 | 16 | 90029417 | G | A | 6.45 | YES |
| PGS000790_hmPOS_GRCh37 | 16 | 90066936 | A | C | 9.88 | YES |
| PGS000790_hmPOS_GRCh37 | 20 | 31806588 | C | T | 6.29 | YES |
| PGS000790_hmPOS_GRCh37 | 20 | 32665748 | A | G | 9.88 | YES |
| PGS000790_hmPOS_GRCh37 | 21 | 42746081 | A | G | 5.94 | YES |
| PGS000790_hmPOS_GRCh37 | 22 | 38563471 | C | T | 6.03 | YES |

Abbreviations: SNP, Single nucleotide polymorphism; PRS, polygenic risk score.

Total number of variants: 24
Matched, n (%): 24 (100%)
Exclusions due to ambiguously matched, n (%): 0 (0%)
Exclusions due to unmatched, n (%): 0 (0%)

**eTable S3. List of SNPs included in colorectal cancer PRS**

| Accession | Chr name | Chr position | Effect allele | Other allele | Effect weight | Include |
|---|---|---|---|---|---|---|
| PGS002252_hmPOS_GRCh37 | 1 | 22587728 | T | C | 0.0504 | YES |
| PGS002252_hmPOS_GRCh37 | 1 | 38455891 | G | C | 0.0523 | NO |
| PGS002252_hmPOS_GRCh37 | 1 | 55246035 | C | T | 0.0665 | YES |
| PGS002252_hmPOS_GRCh37 | 1 | 62673037 | C | T | 0.077 | YES |
| PGS002252_hmPOS_GRCh37 | 1 | 183002639 | A | G | 0.073 | YES |
| PGS002252_hmPOS_GRCh37 | 1 | 222112634 | G | A | 0.0877 | YES |
| PGS002252_hmPOS_GRCh37 | 2 | 48686695 | T | A | 0.0953 | NO |
| PGS002252_hmPOS_GRCh37 | 2 | 98275354 | G | A | 0.1133 | YES |
| PGS002252_hmPOS_GRCh37 | 2 | 159964552 | C | T | 0.0511 | YES |
| PGS002252_hmPOS_GRCh37 | 2 | 199612407 | C | T | 0.0535 | YES |
| PGS002252_hmPOS_GRCh37 | 2 | 199781586 | T | C | 0.0627 | YES |
| PGS002252_hmPOS_GRCh37 | 2 | 219191256 | T | C | 0.0613 | YES |
| PGS002252_hmPOS_GRCh37 | 3 | 40915239 | G | A | 0.0994 | YES |
| PGS002252_hmPOS_GRCh37 | 3 | 53088285 | G | T | 0.0677 | YES |
| PGS002252_hmPOS_GRCh37 | 3 | 66365163 | A | G | 0.0597 | YES |
| PGS002252_hmPOS_GRCh37 | 3 | 112903888 | A | G | 0.0474 | YES |
| PGS002252_hmPOS_GRCh37 | 3 | 112916918 | C | T | 0.077 | YES |
| PGS002252_hmPOS_GRCh37 | 3 | 112999560 | G | A | 0.1761 | YES |
| PGS002252_hmPOS_GRCh37 | 3 | 133701119 | A | G | 0.0597 | YES |
| PGS002252_hmPOS_GRCh37 | 3 | 133748789 | T | C | 0.0953 | YES |
| PGS002252_hmPOS_GRCh37 | 3 | 169517436 | C | T | 0.0453 | YES |
| PGS002252_hmPOS_GRCh37 | 4 | 94938618 | A | C | 0.052 | YES |
| PGS002252_hmPOS_GRCh37 | 4 | 106128760 | A | G | 0.0522 | YES |
| PGS002252_hmPOS_GRCh37 | 4 | 145659064 | C | T | 0.0842 | YES |
| PGS002252_hmPOS_GRCh37 | 5 | 1240204 | T | C | 0.1119 | YES |
| PGS002252_hmPOS_GRCh37 | 5 | 1296486 | G | A | 0.0865 | YES |
| PGS002252_hmPOS_GRCh37 | 5 | 40102443 | A | G | 0.0545 | YES |
| PGS002252_hmPOS_GRCh37 | 5 | 40280076 | A | G | 0.1013 | YES |
| PGS002252_hmPOS_GRCh37 | 5 | 98206082 | T | A | 0.5559 | NO |
| PGS002252_hmPOS_GRCh37 | 5 | 112097351 | G | A | 0.6286 | YES |
| PGS002252_hmPOS_GRCh37 | 5 | 125988175 | G | A | 0.0862 | YES |
| PGS002252_hmPOS_GRCh37 | 5 | 134467220 | C | T | 0.0693 | YES |
| PGS002252_hmPOS_GRCh37 | 6 | 12292772 | T | G | 0.0677 | YES |
| PGS002252_hmPOS_GRCh37 | 6 | 29809860 | A | G | 0.1133 | YES |
| PGS002252_hmPOS_GRCh37 | 6 | 30758466 | G | A | 0.0677 | YES |
| PGS002252_hmPOS_GRCh37 | 6 | 31010185 | C | T | 0.1545 | YES |
| PGS002252_hmPOS_GRCh37 | 6 | 31315512 | G | A | 0.0698 | YES |
| PGS002252_hmPOS_GRCh37 | 6 | 31449620 | C | T | 0.1118 | YES |
| PGS002252_hmPOS_GRCh37 | 6 | 32191339 | T | C | 0.1484 | YES |
| PGS002252_hmPOS_GRCh37 | 6 | 32593080 | G | A | 0.0889 | YES |
| PGS002252_hmPOS_GRCh37 | 6 | 35569562 | A | G | 0.0778 | YES |
| PGS002252_hmPOS_GRCh37 | 6 | 36623379 | A | G | 0.054 | YES |
| PGS002252_hmPOS_GRCh37 | 6 | 41702582 | C | T | 0.033 | YES |
| PGS002252_hmPOS_GRCh37 | 6 | 105970139 | C | A | 0.0322 | YES |
| PGS002252_hmPOS_GRCh37 | 6 | 55566108 | G | A | 0.068 | YES |

| | | | | | | |
|---|---|---|---|---|---|---|
| PGS002252_hmPOS_GRCh37 | 6 | 55712124 | C | T | 0.0724 | YES |
| PGS002252_hmPOS_GRCh37 | 7 | 45136423 | T | C | 0.065 | YES |
| PGS002252_hmPOS_GRCh37 | 7 | 46094089 | T | C | 0.0643 | YES |
| PGS002252_hmPOS_GRCh37 | 7 | 46926695 | C | T | 0.0583 | YES |
| PGS002252_hmPOS_GRCh37 | 7 | 47511161 | G | A | 0.077 | YES |
| PGS002252_hmPOS_GRCh37 | 8 | 117630683 | C | A | 0.2099 | YES |
| PGS002252_hmPOS_GRCh37 | 8 | 117632965 | G | C | 0.0677 | NO |
| PGS002252_hmPOS_GRCh37 | 8 | 117790914 | A | C | 0.1139 | YES |
| PGS002252_hmPOS_GRCh37 | 8 | 128413305 | G | T | 0.1052 | YES |
| PGS002252_hmPOS_GRCh37 | 8 | 128414892 | T | C | 0.0606 | YES |
| PGS002252_hmPOS_GRCh37 | 8 | 128571855 | G | T | 0.0608 | YES |
| PGS002252_hmPOS_GRCh37 | 9 | 22103183 | G | T | 0.0504 | YES |
| PGS002252_hmPOS_GRCh37 | 9 | 101679752 | T | G | 0.0818 | YES |
| PGS002252_hmPOS_GRCh37 | 9 | 113671403 | C | T | 0.0637 | YES |
| PGS002252_hmPOS_GRCh37 | 10 | 8663875 | C | T | 0.0462 | YES |
| PGS002252_hmPOS_GRCh37 | 10 | 8739580 | T | A | 0.1064 | NO |
| PGS002252_hmPOS_GRCh37 | 10 | 52648454 | C | T | 0.073 | YES |
| PGS002252_hmPOS_GRCh37 | 10 | 80819132 | G | A | 0.0765 | YES |
| PGS002252_hmPOS_GRCh37 | 10 | 81046265 | C | T | 0.0483 | YES |
| PGS002252_hmPOS_GRCh37 | 10 | 101315166 | G | A | 0.0514 | YES |
| PGS002252_hmPOS_GRCh37 | 10 | 101351704 | G | A | 0.0889 | YES |
| PGS002252_hmPOS_GRCh37 | 10 | 114288619 | C | T | 0.0975 | YES |
| PGS002252_hmPOS_GRCh37 | 10 | 114722621 | A | G | 0.0527 | YES |
| PGS002252_hmPOS_GRCh37 | 11 | 10286755 | C | A | 0.0953 | YES |
| PGS002252_hmPOS_GRCh37 | 11 | 61549025 | G | A | 0.0636 | YES |
| PGS002252_hmPOS_GRCh37 | 11 | 74280012 | G | T | 0.078 | YES |
| PGS002252_hmPOS_GRCh37 | 11 | 74409077 | C | T | 0.0603 | YES |
| PGS002252_hmPOS_GRCh37 | 11 | 74427921 | C | T | 0.1934 | YES |
| PGS002252_hmPOS_GRCh37 | 11 | 100717136 | G | A | 0.0774 | YES |
| PGS002252_hmPOS_GRCh37 | 11 | 101656397 | T | A | 0.0537 | NO |
| PGS002252_hmPOS_GRCh37 | 11 | 111156836 | T | C | 0.1122 | YES |
| PGS002252_hmPOS_GRCh37 | 12 | 4368607 | C | T | 0.089 | YES |
| PGS002252_hmPOS_GRCh37 | 12 | 4388271 | T | C | 0.1181 | YES |
| PGS002252_hmPOS_GRCh37 | 12 | 4400808 | T | C | 0.055 | YES |
| PGS002252_hmPOS_GRCh37 | 12 | 6406904 | C | T | 0.051 | YES |
| PGS002252_hmPOS_GRCh37 | 12 | 6421174 | T | A | 0.0597 | NO |
| PGS002252_hmPOS_GRCh37 | 12 | 12035649 | C | T | 0.0145 | YES |
| PGS002252_hmPOS_GRCh37 | 12 | 31594813 | T | C | 0.3646 | YES |
| PGS002252_hmPOS_GRCh37 | 12 | 43134191 | G | A | 0.053 | YES |
| PGS002252_hmPOS_GRCh37 | 12 | 51171090 | G | A | 0.0896 | YES |
| PGS002252_hmPOS_GRCh37 | 12 | 57533690 | A | C | 0.053 | YES |
| PGS002252_hmPOS_GRCh37 | 12 | 111973358 | G | A | 0.0737 | YES |
| PGS002252_hmPOS_GRCh37 | 12 | 115100714 | C | T | 0.0456 | YES |
| PGS002252_hmPOS_GRCh37 | 12 | 115890922 | C | T | 0.066 | YES |
| PGS002252_hmPOS_GRCh37 | 12 | 117763309 | A | C | 0.064 | YES |
| PGS002252_hmPOS_GRCh37 | 13 | 34092165 | C | T | 0.0468 | NO |
| PGS002252_hmPOS_GRCh37 | 13 | 37462010 | G | A | 0.0758 | YES |
| PGS002252_hmPOS_GRCh37 | 13 | 73649152 | A | G | 0.0505 | YES |

| | | | | | | |
|---|---|---|---|---|---|---|
| PGS002252_hmPOS_GRCh37 | 13 | 73791554 | C | T | 0.0982 | YES |
| PGS002252_hmPOS_GRCh37 | 13 | 73997961 | A | G | 0.0544 | YES |
| PGS002252_hmPOS_GRCh37 | 13 | 78609615 | C | T | 0.1044 | YES |
| PGS002252_hmPOS_GRCh37 | 13 | 111075881 | T | C | 0.0549 | YES |
| PGS002252_hmPOS_GRCh37 | 14 | 54369299 | A | G | 0.0407 | YES |
| PGS002252_hmPOS_GRCh37 | 14 | 54419106 | C | A | 0.0912 | YES |
| PGS002252_hmPOS_GRCh37 | 14 | 54445157 | G | A | 0.0465 | YES |
| PGS002252_hmPOS_GRCh37 | 14 | 59189361 | G | A | 0.0691 | YES |
| PGS002252_hmPOS_GRCh37 | 14 | 59208437 | A | G | 0.0454 | YES |
| PGS002252_hmPOS_GRCh37 | 15 | 32992836 | G | A | 0.0464 | YES |
| PGS002252_hmPOS_GRCh37 | 15 | 33010736 | A | G | 0.1248 | YES |
| PGS002252_hmPOS_GRCh37 | 15 | 33156386 | A | G | 0.0705 | YES |
| PGS002252_hmPOS_GRCh37 | 15 | 67007018 | C | G | 0.0476 | NO |
| PGS002252_hmPOS_GRCh37 | 15 | 67402824 | C | T | 0.0689 | YES |
| PGS002252_hmPOS_GRCh37 | 15 | 68060389 | G | T | 0.049 | YES |
| PGS002252_hmPOS_GRCh37 | 15 | 91172901 | T | C | 0.1044 | YES |
| PGS002252_hmPOS_GRCh37 | 16 | 68743939 | A | C | 0.055 | YES |
| PGS002252_hmPOS_GRCh37 | 16 | 80043258 | C | A | 0.0498 | YES |
| PGS002252_hmPOS_GRCh37 | 16 | 86252544 | A | G | 0.0472 | YES |
| PGS002252_hmPOS_GRCh37 | 16 | 86339315 | T | C | 0.0487 | YES |
| PGS002252_hmPOS_GRCh37 | 16 | 86703949 | T | C | 0.0481 | YES |
| PGS002252_hmPOS_GRCh37 | 17 | 809643 | G | A | 0.0514 | YES |
| PGS002252_hmPOS_GRCh37 | 17 | 814243 | A | T | 0.068 | NO |
| PGS002252_hmPOS_GRCh37 | 17 | 10707241 | A | G | 0.0748 | YES |
| PGS002252_hmPOS_GRCh37 | 17 | 70413253 | A | G | 0.0595 | YES |
| PGS002252_hmPOS_GRCh37 | 8 | 24766973 | A | G | 0.6995 | NO |
| PGS002252_hmPOS_GRCh37 | 2 | 101194093 | G | A | 0.0882 | NO |
| PGS002252_hmPOS_GRCh37 | 14 | 54705203 | A | T | 0.1606 | NO |
| PGS002252_hmPOS_GRCh37 | 2 | 53218607 | T | G | 0.1939 | NO |
| PGS002252_hmPOS_GRCh37 | 19 | 41871573 | A | G | 0.0441 | YES |
| PGS002252_hmPOS_GRCh37 | 12 | 8047901 | T | C | 0.0632 | NO |
| PGS002252_hmPOS_GRCh37 | 20 | 6376457 | G | C | 0.0795 | NO |
| PGS002252_hmPOS_GRCh37 | 20 | 6603622 | C | T | 0.0627 | YES |
| PGS002252_hmPOS_GRCh37 | 20 | 6699595 | G | T | 0.0819 | YES |
| PGS002252_hmPOS_GRCh37 | 20 | 6762221 | T | C | 0.0714 | YES |
| PGS002252_hmPOS_GRCh37 | 8 | 135019276 | G | A | 0.0874 | NO |
| PGS002252_hmPOS_GRCh37 | 20 | 33213196 | C | A | 0.045 | YES |
| PGS002252_hmPOS_GRCh37 | 20 | 42666475 | T | C | 0.0597 | YES |
| PGS002252_hmPOS_GRCh37 | 20 | 47340117 | A | G | 0.0719 | YES |
| PGS002252_hmPOS_GRCh37 | 20 | 48983697 | C | T | 0.0446 | YES |
| PGS002252_hmPOS_GRCh37 | 20 | 49055318 | C | T | 0.0547 | YES |
| PGS002252_hmPOS_GRCh37 | 20 | 49256285 | T | C | 0.062 | YES |
| PGS002252_hmPOS_GRCh37 | 20 | 57475191 | G | A | 0.077 | YES |
| PGS002252_hmPOS_GRCh37 | 20 | 60932414 | C | T | 0.1146 | YES |
| PGS002252_hmPOS_GRCh37 | 20 | 62308612 | T | G | 0.0593 | YES |

Abbreviations: SNP, Single nucleotide polymorphism; PRS, polygenic risk score.

Total number of variants: 141

Matched, n (%): 121 (85.82%)
Exclusions due to ambiguously matched, n (%): 10 (7.09%)
Exclusions due to unmatched, n (%): 10 (7.09%)

**eTable S4. List of SNPs included in non-Hodgkin lymphoma PRS**

| Accession | Chr name | Chr position | Effect allele | Other allele | Effect weight | Include |
|---|---|---|---|---|---|---|
| PGS000791_hmPOS_GRCh37 | 2 | 24694472 | T | C | 5.48 | YES |
| PGS000791_hmPOS_GRCh37 | 3 | 121800487 | A | C | 5.93 | YES |
| PGS000791_hmPOS_GRCh37 | 3 | 121817613 | A | G | 6.16 | YES |
| PGS000791_hmPOS_GRCh37 | 3 | 188299902 | G | A | 6.45 | YES |
| PGS000791_hmPOS_GRCh37 | 6 | 484453 | G | C | 9.49 | NO |
| PGS000791_hmPOS_GRCh37 | 6 | 29832526 | T | C | 5.75 | YES |
| PGS000791_hmPOS_GRCh37 | 6 | 31074030 | C | A | 6.61 | YES |
| PGS000791_hmPOS_GRCh37 | 6 | 31322790 | A | T | 6.33 | NO |
| PGS000791_hmPOS_GRCh37 | 6 | 32370587 | C | G | 7.86 | NO |
| PGS000791_hmPOS_GRCh37 | 6 | 32444544 | T | C | 21.23 | YES |
| PGS000791_hmPOS_GRCh37 | 6 | 32665420 | G | T | 13.24 | YES |
| PGS000791_hmPOS_GRCh37 | 6 | 32741868 | C | T | 9.41 | YES |
| PGS000791_hmPOS_GRCh37 | 6 | 32796751 | C | T | 7.52 | YES |
| PGS000791_hmPOS_GRCh37 | 8 | 129076573 | T | C | 7.13 | YES |
| PGS000791_hmPOS_GRCh37 | 8 | 129269466 | A | G | 6.62 | YES |
| PGS000791_hmPOS_GRCh37 | 11 | 58060192 | T | C | 5.5 | YES |
| PGS000791_hmPOS_GRCh37 | 11 | 118741842 | C | T | 9.15 | YES |
| PGS000791_hmPOS_GRCh37 | 11 | 128492739 | T | C | 6.53 | YES |
| PGS000791_hmPOS_GRCh37 | 18 | 60783211 | G | A | 6.14 | YES |

Abbreviations: SNP, Single nucleotide polymorphism; PRS, polygenic risk score.

Total number of variants: 19
Matched, n (%): 16 (84.21%)
Exclusions due to ambiguously matched, n (%): 3 (15.79%)
Exclusions due to unmatched, n (%): 0 (0%)

**eTable S5. List of SNPs included in ovarian cancer PRS**

| Accession | Chr name | Chr position | Effect allele | Other allele | Effect weight | Include |
|---|---|---|---|---|---|---|
| PGS003394_hmPOS_GRCh37 | 1 | 22468215 | T | C | 0.0808 | YES |
| PGS003394_hmPOS_GRCh37 | 1 | 38082122 | A | G | 0.0835 | YES |
| PGS003394_hmPOS_GRCh37 | 2 | 111818658 | C | A | 0.0642 | YES |
| PGS003394_hmPOS_GRCh37 | 2 | 111915946 | T | G | -0.0118 | YES |
| PGS003394_hmPOS_GRCh37 | 2 | 113973964 | C | T | 0.0518 | YES |
| PGS003394_hmPOS_GRCh37 | 2 | 113979364 | A | G | 0.0323 | YES |
| PGS003394_hmPOS_GRCh37 | 2 | 177037831 | G | C | -0.1005 | NO |
| PGS003394_hmPOS_GRCh37 | 3 | 156402487 | T | C | 0.3647 | YES |
| PGS003394_hmPOS_GRCh37 | 3 | 190531882 | A | G | -0.0668 | YES |
| PGS003394_hmPOS_GRCh37 | 4 | 70577859 | A | G | -0.0574 | YES |
| PGS003394_hmPOS_GRCh37 | 5 | 1279790 | T | C | 0.0351 | YES |
| PGS003394_hmPOS_GRCh37 | 5 | 1285974 | A | C | 0.059 | YES |
| PGS003394_hmPOS_GRCh37 | 5 | 1287194 | A | G | -0.0849 | YES |
| PGS003394_hmPOS_GRCh37 | 5 | 1295349 | G | A | -0.0797 | YES |
| PGS003394_hmPOS_GRCh37 | 5 | 54476556 | A | G | -0.0666 | YES |
| PGS003394_hmPOS_GRCh37 | 8 | 82653644 | G | A | 0.1294 | YES |
| PGS003394_hmPOS_GRCh37 | 8 | 128817883 | G | A | 0.076 | YES |
| PGS003394_hmPOS_GRCh37 | 8 | 129069820 | A | G | -0.0692 | YES |
| PGS003394_hmPOS_GRCh37 | 8 | 129217984 | C | G | 0.0485 | NO |
| PGS003394_hmPOS_GRCh37 | 8 | 129551633 | A | G | -0.1759 | YES |
| PGS003394_hmPOS_GRCh37 | 9 | 16914716 | A | G | -0.1392 | YES |
| PGS003394_hmPOS_GRCh37 | 9 | 16914835 | C | A | -0.101 | YES |
| PGS003394_hmPOS_GRCh37 | 9 | 19044489 | A | G | 0.0788 | YES |
| PGS003394_hmPOS_GRCh37 | 9 | 106912892 | A | G | 0.0582 | YES |
| PGS003394_hmPOS_GRCh37 | 9 | 136155000 | T | C | 0.0931 | YES |
| PGS003394_hmPOS_GRCh37 | 10 | 21821274 | A | G | 0.0799 | YES |
| PGS003394_hmPOS_GRCh37 | 10 | 112011084 | G | A | 0.0752 | YES |
| PGS003394_hmPOS_GRCh37 | 12 | 121415293 | A | G | -0.0593 | YES |
| PGS003394_hmPOS_GRCh37 | 15 | 91535329 | T | G | -0.0804 | YES |
| PGS003394_hmPOS_GRCh37 | 17 | 36100767 | G | A | -0.0582 | YES |
| PGS003394_hmPOS_GRCh37 | 17 | 44790203 | G | A | 0.1026 | YES |
| PGS003394_hmPOS_GRCh37 | 17 | 46472432 | G | C | 0.1182 | NO |
| PGS003394_hmPOS_GRCh37 | 18 | 21425852 | C | T | -0.0283 | YES |
| PGS003394_hmPOS_GRCh37 | 19 | 17390291 | C | T | 0.0797 | YES |
| PGS003394_hmPOS_GRCh37 | 19 | 17409380 | T | C | -0.0613 | YES |
| PGS003394_hmPOS_GRCh37 | 21 | 36080398 | C | T | -0.0599 | YES |

Abbreviations: SNP, Single nucleotide polymorphism; PRS, polygenic risk score.

Total number of variants: 36
Matched, n (%): 33 (91.67%)
Exclusions due to ambiguously matched, n (%): 3 (8.33%)
Exclusions due to unmatched, n (%): 0 (0%)

**eTable S6. List of SNPs included in prostate cancer PRS**

| Accession | Chr name | Chr position | Effect allele | Other allele | Effect weight | Include |
|---|---|---|---|---|---|---|
| PGS000662_hmPOS_GRCh37 | 1 | 5743196 | T | C | 0.102298257 | YES |
| PGS000662_hmPOS_GRCh37 | 1 | 10564675 | A | G | 0.042411273 | YES |
| PGS000662_hmPOS_GRCh37 | 1 | 16376831 | C | T | 0.055506528 | YES |
| PGS000662_hmPOS_GRCh37 | 1 | 46251655 | T | C | 0.07282201 | YES |
| PGS000662_hmPOS_GRCh37 | 1 | 88210716 | AT | A | 0.048255598 | NO |
| PGS000662_hmPOS_GRCh37 | 1 | 150772613 | C | T | 0.080240037 | YES |
| PGS000662_hmPOS_GRCh37 | 1 | 150954671 | A | G | 0.067047369 | YES |
| PGS000662_hmPOS_GRCh37 | 1 | 153923276 | T | C | 0.066274137 | YES |
| PGS000662_hmPOS_GRCh37 | 1 | 154980351 | T | C | 0.056375028 | YES |
| PGS000662_hmPOS_GRCh37 | 1 | 155118588 | C | T | 0.155225047 | YES |
| PGS000662_hmPOS_GRCh37 | 1 | 155690186 | A | C | 0.159117402 | YES |
| PGS000662_hmPOS_GRCh37 | 1 | 157119915 | C | G | 0.135841227 | NO |
| PGS000662_hmPOS_GRCh37 | 1 | 163295678 | A | C | 0.047902642 | YES |
| PGS000662_hmPOS_GRCh37 | 1 | 167135941 | T | A | 0.069514594 | NO |
| PGS000662_hmPOS_GRCh37 | 1 | 179897070 | A | C | 0.059516664 | YES |
| PGS000662_hmPOS_GRCh37 | 1 | 183032448 | CTAAG | C | 0.041323726 | NO |
| PGS000662_hmPOS_GRCh37 | 1 | 204030363 | TTTTG | T | 0.046586396 | NO |
| PGS000662_hmPOS_GRCh37 | 1 | 204518842 | A | C | 0.098400085 | YES |
| PGS000662_hmPOS_GRCh37 | 1 | 205739266 | C | T | 0.052895608 | YES |
| PGS000662_hmPOS_GRCh37 | 2 | 8598444 | T | G | 0.045271509 | YES |
| PGS000662_hmPOS_GRCh37 | 2 | 10094526 | G | A | 0.079052443 | YES |
| PGS000662_hmPOS_GRCh37 | 2 | 10781975 | T | C | 0.063322715 | YES |
| PGS000662_hmPOS_GRCh37 | 2 | 16016503 | C | A | 0.066285498 | YES |
| PGS000662_hmPOS_GRCh37 | 2 | 20878105 | G | A | 0.087466559 | YES |
| PGS000662_hmPOS_GRCh37 | 2 | 43064555 | C | G | 0.04556022 | NO |
| PGS000662_hmPOS_GRCh37 | 2 | 43637998 | A | G | 0.084466752 | YES |
| PGS000662_hmPOS_GRCh37 | 2 | 43851282 | C | G | 0.055212234 | NO |
| PGS000662_hmPOS_GRCh37 | 2 | 62752975 | G | A | 0.106870463 | YES |
| PGS000662_hmPOS_GRCh37 | 2 | 63277843 | G | C | 0.112596574 | NO |
| PGS000662_hmPOS_GRCh37 | 2 | 63938756 | G | A | 0.243643948 | YES |
| PGS000662_hmPOS_GRCh37 | 2 | 66652885 | T | C | 0.159911493 | YES |
| PGS000662_hmPOS_GRCh37 | 2 | 85767735 | C | T | 0.085899411 | YES |
| PGS000662_hmPOS_GRCh37 | 2 | 111861993 | A | T | 0.095280687 | NO |
| PGS000662_hmPOS_GRCh37 | 2 | 111893096 | T | G | 0.058380501 | YES |
| PGS000662_hmPOS_GRCh37 | 2 | 121103598 | T | C | 0.08789715 | YES |
| PGS000662_hmPOS_GRCh37 | 2 | 121373466 | G | A | 0.065252998 | YES |
| PGS000662_hmPOS_GRCh37 | 2 | 169012955 | C | T | 0.059835828 | YES |
| PGS000662_hmPOS_GRCh37 | 2 | 173319930 | C | T | 0.229445977 | YES |
| PGS000662_hmPOS_GRCh37 | 2 | 174234547 | C | T | 0.056961427 | YES |
| PGS000662_hmPOS_GRCh37 | 2 | 202126615 | G | A | 0.050616031 | YES |
| PGS000662_hmPOS_GRCh37 | 2 | 208118301 | C | T | 0.050644581 | YES |
| PGS000662_hmPOS_GRCh37 | 2 | 238411293 | C | T | 0.133853627 | YES |
| PGS000662_hmPOS_GRCh37 | 2 | 238443226 | G | A | 0.062186442 | YES |
| PGS000662_hmPOS_GRCh37 | 2 | 242135265 | A | G | 0.283885104 | YES |
| PGS000662_hmPOS_GRCh37 | 2 | 242139600 | G | A | 0.359062222 | YES |

| | | | | | | |
|---|---|---|---|---|---|---|
| PGS000662_hmPOS_GRCh37 | 2 | 242141719 | C | T | 0.053496477 | YES |
| PGS000662_hmPOS_GRCh37 | 2 | 242157241 | A | G | 0.11056242 | YES |
| PGS000662_hmPOS_GRCh37 | 3 | 18738940 | A | G | 0.059491057 | YES |
| PGS000662_hmPOS_GRCh37 | 3 | 23153062 | A | C | 0.06163615 | YES |
| PGS000662_hmPOS_GRCh37 | 3 | 49621719 | A | AT | 0.068887973 | NO |
| PGS000662_hmPOS_GRCh37 | 3 | 70796696 | T | C | 0.041720931 | YES |
| PGS000662_hmPOS_GRCh37 | 3 | 87144017 | G | A | 0.164627225 | YES |
| PGS000662_hmPOS_GRCh37 | 3 | 87175984 | C | T | 0.093723754 | YES |
| PGS000662_hmPOS_GRCh37 | 3 | 87399362 | A | G | 0.15607593 | YES |
| PGS000662_hmPOS_GRCh37 | 3 | 106962521 | G | C | 0.047017378 | NO |
| PGS000662_hmPOS_GRCh37 | 3 | 107193337 | C | T | 0.081504805 | YES |
| PGS000662_hmPOS_GRCh37 | 3 | 113300183 | A | T | 0.084309919 | NO |
| PGS000662_hmPOS_GRCh37 | 3 | 127898501 | C | A | 0.101362283 | YES |
| PGS000662_hmPOS_GRCh37 | 3 | 128213994 | G | A | 0.093750697 | YES |
| PGS000662_hmPOS_GRCh37 | 3 | 137562823 | A | G | 0.047975041 | YES |
| PGS000662_hmPOS_GRCh37 | 3 | 141147414 | C | T | 0.05209769 | YES |
| PGS000662_hmPOS_GRCh37 | 3 | 152011746 | C | CAT | 0.088057213 | NO |
| PGS000662_hmPOS_GRCh37 | 3 | 169482335 | T | C | 0.07375821 | YES |
| PGS000662_hmPOS_GRCh37 | 3 | 170074517 | G | C | 0.190119611 | NO |
| PGS000662_hmPOS_GRCh37 | 3 | 170083541 | C | CTTTTT | 0.10473957 | NO |
| PGS000662_hmPOS_GRCh37 | 3 | 172381777 | C | T | 0.0546219 | YES |
| PGS000662_hmPOS_GRCh37 | 4 | 74442349 | G | C | 0.204832836 | NO |
| PGS000662_hmPOS_GRCh37 | 4 | 74477135 | T | A | 0.07520493 | NO |
| PGS000662_hmPOS_GRCh37 | 4 | 95544718 | G | A | 0.077460586 | YES |
| PGS000662_hmPOS_GRCh37 | 4 | 106061534 | C | A | 0.097055643 | YES |
| PGS000662_hmPOS_GRCh37 | 4 | 106064754 | C | T | 0.079845258 | YES |
| PGS000662_hmPOS_GRCh37 | 4 | 140948835 | C | T | 0.060799244 | YES |
| PGS000662_hmPOS_GRCh37 | 4 | 146879237 | G | A | 0.064685356 | YES |
| PGS000662_hmPOS_GRCh37 | 4 | 152030340 | T | C | 0.085442782 | YES |
| PGS000662_hmPOS_GRCh37 | 5 | 1280028 | G | A | 0.135158159 | YES |
| PGS000662_hmPOS_GRCh37 | 5 | 1292118 | A | G | 0.172287838 | YES |
| PGS000662_hmPOS_GRCh37 | 5 | 1294086 | T | C | 0.072437187 | YES |
| PGS000662_hmPOS_GRCh37 | 5 | 1891174 | G | C | 0.138338763 | NO |
| PGS000662_hmPOS_GRCh37 | 5 | 14372363 | T | TGA | 0.056987101 | NO |
| PGS000662_hmPOS_GRCh37 | 5 | 37833419 | T | C | 0.038762919 | YES |
| PGS000662_hmPOS_GRCh37 | 5 | 44368506 | T | C | 0.039043082 | YES |
| PGS000662_hmPOS_GRCh37 | 5 | 56087910 | A | G | 0.051060896 | YES |
| PGS000662_hmPOS_GRCh37 | 5 | 133836209 | T | C | 0.077994327 | YES |
| PGS000662_hmPOS_GRCh37 | 5 | 169172133 | A | G | 0.275653809 | YES |
| PGS000662_hmPOS_GRCh37 | 5 | 172959030 | C | A | 0.042565359 | YES |
| PGS000662_hmPOS_GRCh37 | 5 | 177683905 | G | A | 0.104830802 | YES |
| PGS000662_hmPOS_GRCh37 | 5 | 177891551 | G | A | 0.050421302 | YES |
| PGS000662_hmPOS_GRCh37 | 6 | 1670985 | A | G | 0.044086511 | YES |
| PGS000662_hmPOS_GRCh37 | 6 | 11217897 | T | C | 0.066566318 | YES |
| PGS000662_hmPOS_GRCh37 | 6 | 21330689 | C | T | 0.089547244 | YES |
| PGS000662_hmPOS_GRCh37 | 6 | 21471490 | G | A | 0.058062324 | YES |
| PGS000662_hmPOS_GRCh37 | 6 | 21878849 | A | C | 0.043143107 | YES |
| PGS000662_hmPOS_GRCh37 | 6 | 26649831 | C | CT | 0.044483881 | NO |

| | | | | | | |
|---|---|---|---|---|---|---|
| PGS000662_hmPOS_GRCh37 | 6 | 30216712 | C | T | 0.045601728 | YES |
| PGS000662_hmPOS_GRCh37 | 6 | 32652620 | A | G | 0.049032422 | YES |
| PGS000662_hmPOS_GRCh37 | 6 | 34793124 | A | G | 0.04748673 | YES |
| PGS000662_hmPOS_GRCh37 | 6 | 41536587 | G | T | 0.10184104 | YES |
| PGS000662_hmPOS_GRCh37 | 6 | 43709785 | C | T | 0.041792627 | YES |
| PGS000662_hmPOS_GRCh37 | 6 | 76495882 | A | G | 0.077914375 | YES |
| PGS000662_hmPOS_GRCh37 | 6 | 109295293 | C | T | 0.073721053 | YES |
| PGS000662_hmPOS_GRCh37 | 6 | 117200434 | C | A | 0.146268108 | YES |
| PGS000662_hmPOS_GRCh37 | 6 | 134292718 | G | GTGTTGT | 0.049325055 | NO |
| PGS000662_hmPOS_GRCh37 | 6 | 153447516 | C | T | 0.072334334 | YES |
| PGS000662_hmPOS_GRCh37 | 6 | 160150279 | C | T | 0.065796318 | YES |
| PGS000662_hmPOS_GRCh37 | 6 | 160581544 | TG | T | 0.190781896 | NO |
| PGS000662_hmPOS_GRCh37 | 7 | 1928159 | T | C | 0.044010136 | YES |
| PGS000662_hmPOS_GRCh37 | 7 | 20414111 | T | TA | 0.048596315 | NO |
| PGS000662_hmPOS_GRCh37 | 7 | 20999211 | C | A | 0.088812729 | YES |
| PGS000662_hmPOS_GRCh37 | 7 | 21812043 | T | G | 0.049687897 | YES |
| PGS000662_hmPOS_GRCh37 | 7 | 27564862 | A | C | 0.06707809 | YES |
| PGS000662_hmPOS_GRCh37 | 7 | 27976563 | G | A | 0.111327016 | YES |
| PGS000662_hmPOS_GRCh37 | 7 | 40877473 | A | G | 0.080139298 | YES |
| PGS000662_hmPOS_GRCh37 | 7 | 47451918 | A | T | 0.051018785 | NO |
| PGS000662_hmPOS_GRCh37 | 7 | 92577760 | C | T | 0.056403933 | YES |
| PGS000662_hmPOS_GRCh37 | 7 | 97688440 | A | G | 0.101354017 | YES |
| PGS000662_hmPOS_GRCh37 | 8 | 8498803 | G | A | 0.061774284 | YES |
| PGS000662_hmPOS_GRCh37 | 8 | 11217455 | C | T | 0.048335618 | YES |
| PGS000662_hmPOS_GRCh37 | 8 | 23470785 | A | G | 0.072294359 | YES |
| PGS000662_hmPOS_GRCh37 | 8 | 23529521 | G | A | 0.169346695 | YES |
| PGS000662_hmPOS_GRCh37 | 8 | 25894202 | C | CCAAA | 0.077048439 | NO |
| PGS000662_hmPOS_GRCh37 | 8 | 26063165 | A | C | 0.060975403 | YES |
| PGS000662_hmPOS_GRCh37 | 8 | 38644914 | A | G | 0.045607893 | YES |
| PGS000662_hmPOS_GRCh37 | 8 | 108923107 | G | A | 0.058608641 | YES |
| PGS000662_hmPOS_GRCh37 | 8 | 127836926 | C | CAA | 0.034625563 | NO |
| PGS000662_hmPOS_GRCh37 | 8 | 127901649 | G | A | 0.089020363 | YES |
| PGS000662_hmPOS_GRCh37 | 8 | 127922200 | A | T | -0.149935287 | NO |
| PGS000662_hmPOS_GRCh37 | 8 | 128027954 | G | A | 0.18129333 | YES |
| PGS000662_hmPOS_GRCh37 | 8 | 128074815 | T | A | 0.735882073 | NO |
| PGS000662_hmPOS_GRCh37 | 8 | 128077146 | A | G | 0.637777711 | YES |
| PGS000662_hmPOS_GRCh37 | 8 | 128103969 | T | C | 0.374930958 | YES |
| PGS000662_hmPOS_GRCh37 | 8 | 128104117 | G | A | 0.819477616 | YES |
| PGS000662_hmPOS_GRCh37 | 8 | 128104218 | G | A | 0.097171134 | YES |
| PGS000662_hmPOS_GRCh37 | 8 | 128325355 | T | C | 0.056212284 | YES |
| PGS000662_hmPOS_GRCh37 | 8 | 128342866 | A | G | 0.114037591 | YES |
| PGS000662_hmPOS_GRCh37 | 8 | 128413305 | G | T | 0.159518618 | YES |
| PGS000662_hmPOS_GRCh37 | 8 | 128532137 | T | C | 0.349467374 | YES |
| PGS000662_hmPOS_GRCh37 | 8 | 128535543 | T | C | 0.162669308 | YES |
| PGS000662_hmPOS_GRCh37 | 8 | 128540776 | C | G | 0.178710296 | NO |
| PGS000662_hmPOS_GRCh37 | 9 | 18554773 | C | T | 0.051895692 | YES |
| PGS000662_hmPOS_GRCh37 | 9 | 19072246 | C | A | 0.058761512 | YES |
| PGS000662_hmPOS_GRCh37 | 9 | 22041998 | G | C | 0.075703052 | NO |

| | | | | | | |
|---|---|---|---|---|---|---|
| PGS000662_hmPOS_GRCh37 | 9 | 34049779 | T | A | 0.052259971 | NO |
| PGS000662_hmPOS_GRCh37 | 9 | 82090724 | T | TG | 0.078426239 | NO |
| PGS000662_hmPOS_GRCh37 | 9 | 109532734 | A | G | 0.052079327 | YES |
| PGS000662_hmPOS_GRCh37 | 9 | 110144887 | C | T | 0.072440511 | YES |
| PGS000662_hmPOS_GRCh37 | 9 | 110290217 | G | A | 0.26518417 | YES |
| PGS000662_hmPOS_GRCh37 | 9 | 130430116 | A | G | 0.04198732 | YES |
| PGS000662_hmPOS_GRCh37 | 9 | 132573536 | T | G | 0.059682543 | YES |
| PGS000662_hmPOS_GRCh37 | 10 | 838636 | T | C | 0.089270754 | YES |
| PGS000662_hmPOS_GRCh37 | 10 | 46104943 | A | C | 0.118993393 | YES |
| PGS000662_hmPOS_GRCh37 | 10 | 47599029 | T | C | 0.116991363 | YES |
| PGS000662_hmPOS_GRCh37 | 10 | 51549496 | T | C | 0.203974916 | YES |
| PGS000662_hmPOS_GRCh37 | 10 | 80236999 | C | A | 0.121628628 | YES |
| PGS000662_hmPOS_GRCh37 | 10 | 80835998 | C | T | 0.09074034 | YES |
| PGS000662_hmPOS_GRCh37 | 10 | 90195149 | C | T | 0.045668292 | YES |
| PGS000662_hmPOS_GRCh37 | 10 | 104428716 | C | T | 0.077375055 | YES |
| PGS000662_hmPOS_GRCh37 | 10 | 114711755 | T | C | 0.046793119 | YES |
| PGS000662_hmPOS_GRCh37 | 10 | 122794926 | A | G | 0.060375465 | YES |
| PGS000662_hmPOS_GRCh37 | 10 | 122834482 | C | T | 0.222648065 | YES |
| PGS000662_hmPOS_GRCh37 | 10 | 123054018 | T | A | 0.061850323 | NO |
| PGS000662_hmPOS_GRCh37 | 10 | 123185303 | A | G | 0.133130115 | YES |
| PGS000662_hmPOS_GRCh37 | 10 | 126697494 | C | G | 0.061056764 | NO |
| PGS000662_hmPOS_GRCh37 | 11 | 1507512 | T | C | 0.056092669 | YES |
| PGS000662_hmPOS_GRCh37 | 11 | 2234093 | T | C | 0.160859609 | YES |
| PGS000662_hmPOS_GRCh37 | 11 | 7547587 | A | G | 0.104991688 | YES |
| PGS000662_hmPOS_GRCh37 | 11 | 47428210 | T | TA | 0.051908161 | NO |
| PGS000662_hmPOS_GRCh37 | 11 | 58902679 | G | A | 0.128657643 | YES |
| PGS000662_hmPOS_GRCh37 | 11 | 61908440 | C | T | 0.058255547 | YES |
| PGS000662_hmPOS_GRCh37 | 11 | 66951965 | C | G | 0.130217912 | NO |
| PGS000662_hmPOS_GRCh37 | 11 | 68882926 | T | C | 0.051637519 | YES |
| PGS000662_hmPOS_GRCh37 | 11 | 68980788 | A | G | 0.092296194 | YES |
| PGS000662_hmPOS_GRCh37 | 11 | 69002342 | C | T | 0.240823718 | YES |
| PGS000662_hmPOS_GRCh37 | 11 | 69463273 | A | G | 0.16915579 | YES |
| PGS000662_hmPOS_GRCh37 | 11 | 76267331 | T | G | 0.059160904 | YES |
| PGS000662_hmPOS_GRCh37 | 11 | 102401661 | T | C | 0.07934903 | YES |
| PGS000662_hmPOS_GRCh37 | 11 | 108357137 | A | G | 0.144979413 | YES |
| PGS000662_hmPOS_GRCh37 | 11 | 113700547 | C | CA | 0.069578734 | NO |
| PGS000662_hmPOS_GRCh37 | 11 | 125054793 | T | C | 0.285947593 | YES |
| PGS000662_hmPOS_GRCh37 | 11 | 134266372 | G | A | 0.073446603 | YES |
| PGS000662_hmPOS_GRCh37 | 12 | 12871099 | T | G | 0.056424874 | YES |
| PGS000662_hmPOS_GRCh37 | 12 | 12877983 | A | G | 0.076447621 | YES |
| PGS000662_hmPOS_GRCh37 | 12 | 14416918 | G | A | 0.06097533 | YES |
| PGS000662_hmPOS_GRCh37 | 12 | 48419618 | A | C | 0.093020927 | YES |
| PGS000662_hmPOS_GRCh37 | 12 | 49672714 | G | A | 0.077256564 | YES |
| PGS000662_hmPOS_GRCh37 | 12 | 53308932 | A | C | 0.165492625 | YES |
| PGS000662_hmPOS_GRCh37 | 12 | 53329231 | T | C | 0.301244267 | YES |
| PGS000662_hmPOS_GRCh37 | 12 | 65012824 | T | C | 0.062609474 | YES |
| PGS000662_hmPOS_GRCh37 | 12 | 90156377 | A | G | 0.070819974 | YES |
| PGS000662_hmPOS_GRCh37 | 12 | 102446675 | G | T | 0.047788788 | YES |

| | | | | | | |
|---|---|---|---|---|---|---|
| PGS000662_hmPOS_GRCh37 | 12 | 114685571 | A | G | 0.073524623 | YES |
| PGS000662_hmPOS_GRCh37 | 12 | 133067989 | G | A | 0.062621956 | YES |
| PGS000662_hmPOS_GRCh37 | 13 | 51076440 | T | C | 0.056105531 | YES |
| PGS000662_hmPOS_GRCh37 | 13 | 73716861 | C | T | 0.102183238 | YES |
| PGS000662_hmPOS_GRCh37 | 13 | 73995877 | G | C | 0.093202538 | NO |
| PGS000662_hmPOS_GRCh37 | 13 | 110360784 | T | C | 0.424306385 | YES |
| PGS000662_hmPOS_GRCh37 | 14 | 23305649 | T | C | 0.049570527 | YES |
| PGS000662_hmPOS_GRCh37 | 14 | 37136194 | G | A | 0.06401016 | YES |
| PGS000662_hmPOS_GRCh37 | 14 | 38144592 | G | A | 0.046312065 | YES |
| PGS000662_hmPOS_GRCh37 | 14 | 53387109 | G | T | 0.088312271 | YES |
| PGS000662_hmPOS_GRCh37 | 14 | 61106699 | G | A | 0.074161166 | YES |
| PGS000662_hmPOS_GRCh37 | 14 | 64687926 | T | C | 0.067314721 | YES |
| PGS000662_hmPOS_GRCh37 | 14 | 68923908 | A | G | 0.057335106 | YES |
| PGS000662_hmPOS_GRCh37 | 14 | 69134264 | G | A | 0.052936267 | YES |
| PGS000662_hmPOS_GRCh37 | 14 | 70756333 | G | A | 0.048120875 | YES |
| PGS000662_hmPOS_GRCh37 | 15 | 40965044 | G | A | 0.066596828 | YES |
| PGS000662_hmPOS_GRCh37 | 15 | 56385868 | A | G | 0.178845155 | YES |
| PGS000662_hmPOS_GRCh37 | 15 | 66835704 | G | A | 0.062171559 | YES |
| PGS000662_hmPOS_GRCh37 | 15 | 66942093 | A | C | 0.093921349 | YES |
| PGS000662_hmPOS_GRCh37 | 15 | 70668824 | A | C | 0.048031342 | YES |
| PGS000662_hmPOS_GRCh37 | 16 | 54469331 | T | C | 0.04788069 | YES |
| PGS000662_hmPOS_GRCh37 | 16 | 54678305 | C | T | 0.060498718 | YES |
| PGS000662_hmPOS_GRCh37 | 16 | 57654576 | C | T | 0.142619205 | YES |
| PGS000662_hmPOS_GRCh37 | 16 | 79847632 | C | T | 0.08751587 | YES |
| PGS000662_hmPOS_GRCh37 | 16 | 82166181 | C | T | 0.044136641 | YES |
| PGS000662_hmPOS_GRCh37 | 17 | 618965 | C | T | 0.082495207 | YES |
| PGS000662_hmPOS_GRCh37 | 17 | 7571752 | G | T | 0.23290835 | YES |
| PGS000662_hmPOS_GRCh37 | 17 | 7803118 | C | T | 0.157114393 | YES |
| PGS000662_hmPOS_GRCh37 | 17 | 12585459 | A | G | 0.059475847 | YES |
| PGS000662_hmPOS_GRCh37 | 17 | 30092898 | G | A | 0.061679544 | YES |
| PGS000662_hmPOS_GRCh37 | 17 | 36047417 | A | G | 0.060811983 | YES |
| PGS000662_hmPOS_GRCh37 | 17 | 36074979 | G | A | 0.110163448 | YES |
| PGS000662_hmPOS_GRCh37 | 17 | 36103565 | A | G | 0.206472276 | YES |
| PGS000662_hmPOS_GRCh37 | 17 | 46805705 | T | C | 1.380384131 | YES |
| PGS000662_hmPOS_GRCh37 | 17 | 47380305 | T | C | 0.053949366 | YES |
| PGS000662_hmPOS_GRCh37 | 17 | 47398245 | T | C | 0.114231832 | YES |
| PGS000662_hmPOS_GRCh37 | 17 | 56426027 | T | C | 0.04254438 | YES |
| PGS000662_hmPOS_GRCh37 | 17 | 69117533 | G | GTTAT | 0.157548071 | NO |
| PGS000662_hmPOS_GRCh37 | 18 | 51771322 | C | T | 0.044999533 | YES |
| PGS000662_hmPOS_GRCh37 | 18 | 56745999 | G | C | 0.044548356 | NO |
| PGS000662_hmPOS_GRCh37 | 18 | 60961194 | CT | C | 0.051201137 | NO |
| PGS000662_hmPOS_GRCh37 | 18 | 73035513 | T | G | 0.039953327 | YES |
| PGS000662_hmPOS_GRCh37 | 18 | 76770820 | A | G | 0.08337745 | YES |
| PGS000662_hmPOS_GRCh37 | 19 | 17228554 | C | T | 0.054648648 | YES |
| PGS000662_hmPOS_GRCh37 | 19 | 32168343 | C | T | 0.079224515 | YES |
| PGS000662_hmPOS_GRCh37 | 19 | 38548094 | G | T | 0.067520462 | YES |
| PGS000662_hmPOS_GRCh37 | 19 | 38738130 | G | C | 0.091003863 | NO |
| PGS000662_hmPOS_GRCh37 | 19 | 41985931 | T | A | 0.086138393 | NO |

| | | | | | | |
|---|---|---|---|---|---|---|
| PGS000662_hmPOS_GRCh37 | 19 | 51345568 | G | C | 0.100157886 | NO |
| PGS000662_hmPOS_GRCh37 | 19 | 51361382 | G | A | 0.131928692 | YES |
| PGS000662_hmPOS_GRCh37 | 19 | 51362715 | T | G | 0.266769802 | YES |
| PGS000662_hmPOS_GRCh37 | 20 | 867324 | A | G | 0.04275786 | YES |
| PGS000662_hmPOS_GRCh37 | 20 | 31347513 | C | CA | 0.046670938 | NO |
| PGS000662_hmPOS_GRCh37 | 20 | 34006970 | C | T | 0.04968154 | YES |
| PGS000662_hmPOS_GRCh37 | 20 | 49548807 | T | C | 0.116771813 | YES |
| PGS000662_hmPOS_GRCh37 | 20 | 52464719 | C | T | 0.06358626 | YES |
| PGS000662_hmPOS_GRCh37 | 20 | 61001062 | TGGCAGTGGCAGC | T | 0.044630049 | NO |
| PGS000662_hmPOS_GRCh37 | 20 | 62229989 | A | G | 0.05314199 | YES |
| PGS000662_hmPOS_GRCh37 | 20 | 62307517 | A | G | 0.106016091 | YES |
| PGS000662_hmPOS_GRCh37 | 20 | 62374389 | C | T | 0.112172341 | YES |
| PGS000662_hmPOS_GRCh37 | 21 | 40296411 | C | T | 0.05185068 | YES |
| PGS000662_hmPOS_GRCh37 | 21 | 42866332 | A | G | 0.232397366 | YES |
| PGS000662_hmPOS_GRCh37 | 21 | 42882462 | C | T | 0.091599366 | YES |
| PGS000662_hmPOS_GRCh37 | 22 | 19749525 | G | A | 0.05750687 | YES |
| PGS000662_hmPOS_GRCh37 | 22 | 28888939 | A | G | 0.146472871 | YES |
| PGS000662_hmPOS_GRCh37 | 22 | 29091147 | C | A | 0.466208291 | YES |
| PGS000662_hmPOS_GRCh37 | 22 | 29091857 | A | AG | 0.390205621 | NO |
| PGS000662_hmPOS_GRCh37 | 22 | 39138332 | G | A | 0.153105391 | YES |
| PGS000662_hmPOS_GRCh37 | 22 | 40499107 | T | A | 0.072436521 | NO |
| PGS000662_hmPOS_GRCh37 | 22 | 43499741 | G | A | 0.154456689 | YES |
| PGS000662_hmPOS_GRCh37 | 22 | 43500212 | G | T | 0.120809585 | YES |
| PGS000662_hmPOS_GRCh37 | 22 | 45698149 | T | A | 0.047012421 | NO |
| PGS000662_hmPOS_GRCh37 | X | 9811095 | A | G | 0.05607936 | YES |
| PGS000662_hmPOS_GRCh37 | X | 11482634 | C | T | 0.059017309 | YES |
| PGS000662_hmPOS_GRCh37 | X | 30896320 | T | C | 0.032547584 | YES |
| PGS000662_hmPOS_GRCh37 | X | 51245277 | GA | G | 0.099701455 | NO |
| PGS000662_hmPOS_GRCh37 | X | 52695895 | G | A | 0.054267213 | YES |
| PGS000662_hmPOS_GRCh37 | X | 54454406 | A | G | 0.048769606 | YES |
| PGS000662_hmPOS_GRCh37 | X | 66825357 | T | C | 0.077895055 | YES |
| PGS000662_hmPOS_GRCh37 | X | 70139908 | A | G | 0.05554748 | YES |

Abbreviations: SNP, Single nucleotide polymorphism; PRS, polygenic risk score.

Total number of variants: 269
Matched, n (%): 219 (81.41%)
Exclusions due to ambiguously matched, n (%): 27 (10.04%)
Exclusions due to unmatched, n (%): 23 (8.55%)

**eTable S7. List of SNPs included in brain cancer PRS**

| Accession | Chr name | Chr position | Effect allele | Other allele | Effect weight | Include |
|---|---|---|---|---|---|---|
| PGS000624_hmPOS_GRCh37 | 5 | 1279790 | T | C | 0.476234179 | YES |
| PGS000624_hmPOS_GRCh37 | 7 | 54916280 | G | T | 0.488580015 | YES |
| PGS000624_hmPOS_GRCh37 | 9 | 22032152 | G | T | 0.31481074 | YES |
| PGS000624_hmPOS_GRCh37 | 17 | 7571752 | G | T | 0.966983846 | YES |
| PGS000624_hmPOS_GRCh37 | 20 | 62312299 | C | T | 0.392042088 | YES |

Abbreviations: SNP, Single nucleotide polymorphism; PRS, polygenic risk score.

Total number of variants: 5
Matched, n (%): 5 (0%)
Exclusions due to ambiguously matched, n (%): 0 (0%)
Exclusions due to unmatched, n (%): 0 (0%)

**eTable S8. List of SNPs included in head and neck cancer PRS**

| Accession | Chr name | Chr position | Effect allele | Other allele | Effect weight | Include |
|---|---|---|---|---|---|---|
| PGS000792_hmPOS_GRCh37 | 2 | 27855924 | G | A | 5.49 | YES |
| PGS000792_hmPOS_GRCh37 | 4 | 84374480 | C | T | 5.73 | YES |
| PGS000792_hmPOS_GRCh37 | 4 | 100239319 | G | A | 7.18 | YES |
| PGS000792_hmPOS_GRCh37 | 4 | 100341861 | G | A | 8.32 | YES |
| PGS000792_hmPOS_GRCh37 | 5 | 1336221 | G | A | 5.89 | YES |
| PGS000792_hmPOS_GRCh37 | 5 | 1343794 | C | T | 6.09 | YES |
| PGS000792_hmPOS_GRCh37 | 6 | 32587300 | G | A | 6.06 | YES |
| PGS000792_hmPOS_GRCh37 | 6 | 32620838 | C | G | 5.91 | NO |
| PGS000792_hmPOS_GRCh37 | 6 | 32633287 | T | C | 6.15 | YES |
| PGS000792_hmPOS_GRCh37 | 6 | 32635219 | A | G | 5.78 | YES |
| PGS000792_hmPOS_GRCh37 | 6 | 32636120 | C | T | 7.44 | YES |
| PGS000792_hmPOS_GRCh37 | 9 | 22060136 | A | G | 5.79 | YES |
| PGS000792_hmPOS_GRCh37 | 9 | 133952024 | G | A | 5.6 | YES |
| PGS000792_hmPOS_GRCh37 | 12 | 112521448 | A | G | 5.61 | YES |

Abbreviations: SNP, Single nucleotide polymorphism; PRS, polygenic risk score.

Total number of variants: 14
Matched, n (%): 13 (92.86%)
Exclusions due to ambiguously matched, n (%): 1 (7.14%)
Exclusions due to unmatched, n (%): 0 (0%)

**eTable S9. List of SNPs included in lung cancer PRS**

| Accession | Chr name | Chr position | Effect allele | Other allele | Effect weight | Include |
|---|---|---|---|---|---|---|
| PGS000740_hmPOS_GRCh37 | 1 | 77967507 | A | T | 0.131028262 | NO |
| PGS000740_hmPOS_GRCh37 | 3 | 189357199 | T | G | -0.061875404 | YES |
| PGS000740_hmPOS_GRCh37 | 5 | 1285974 | A | C | 0.1133286853070032 | YES |
| PGS000740_hmPOS_GRCh37 | 5 | 1286032 | A | G | 0.3293037471426003 | YES |
| PGS000740_hmPOS_GRCh37 | 5 | 1294086 | T | C | 0.131028262 | YES |
| PGS000740_hmPOS_GRCh37 | 5 | 1300025 | T | G | 0.076961041 | YES |
| PGS000740_hmPOS_GRCh37 | 5 | 1322087 | T | C | -0.139262067 | YES |
| PGS000740_hmPOS_GRCh37 | 5 | 1325767 | G | A | -0.094310679 | YES |
| PGS000740_hmPOS_GRCh37 | 6 | 28322296 | G | A | 0.067658648 | NO |
| PGS000740_hmPOS_GRCh37 | 6 | 31434111 | G | A | 0.1397619423751586 | YES |
| PGS000740_hmPOS_GRCh37 | 6 | 31472720 | T | G | 0.048790164 | NO |
| PGS000740_hmPOS_GRCh37 | 6 | 31628733 | T | C | 0.067658648 | YES |
| PGS000740_hmPOS_GRCh37 | 6 | 31678774 | A | G | -0.105360516 | YES |
| PGS000740_hmPOS_GRCh37 | 6 | 32590925 | A | G | 0.1133286853070032 | YES |
| PGS000740_hmPOS_GRCh37 | 6 | 32635629 | A | G | 0.058268908 | NO |
| PGS000740_hmPOS_GRCh37 | 6 | 167376466 | C | G | 0.067658648 | NO |
| PGS000740_hmPOS_GRCh37 | 8 | 27344719 | A | G | -0.139262067 | YES |
| PGS000740_hmPOS_GRCh37 | 8 | 32410110 | G | A | 0.067658648 | YES |
| PGS000740_hmPOS_GRCh37 | 9 | 21830157 | G | A | 0.086177696 | YES |
| PGS000740_hmPOS_GRCh37 | 9 | 136503819 | T | C | -0.040821995 | YES |
| PGS000740_hmPOS_GRCh37 | 10 | 105687632 | C | A | 0.067658648 | YES |
| PGS000740_hmPOS_GRCh37 | 11 | 118125625 | T | C | 0.067658648 | YES |
| PGS000740_hmPOS_GRCh37 | 12 | 998819 | C | G | -0.083381609 | NO |
| PGS000740_hmPOS_GRCh37 | 13 | 32972626 | T | A | 0.4700036292457356 | NO |
| PGS000740_hmPOS_GRCh37 | 15 | 43762196 | T | C | -0.072570693 | YES |
| PGS000740_hmPOS_GRCh37 | 15 | 47577451 | A | G | 0.067658648 | YES |
| PGS000740_hmPOS_GRCh37 | 15 | 49376624 | G | T | -0.083381609 | YES |
| PGS000740_hmPOS_GRCh37 | 15 | 78825917 | G | A | 0.1570037488096647 | YES |
| PGS000740_hmPOS_GRCh37 | 15 | 78857986 | G | C | 0.262364264 | NO |
| PGS000740_hmPOS_GRCh37 | 15 | 79089734 | C | T | 0.1222176327242491 | YES |
| PGS000740_hmPOS_GRCh37 | 17 | 28263980 | G | A | 0.029558802 | NO |
| PGS000740_hmPOS_GRCh37 | 17 | 72938100 | T | C | 0.039220713 | YES |
| PGS000740_hmPOS_GRCh37 | 19 | 41353107 | T | C | -0.127833372 | YES |
| PGS000740_hmPOS_GRCh37 | 20 | 62326579 | T | G | 0.076961041 | YES |
| PGS000740_hmPOS_GRCh37 | 22 | 29121087 | G | A | -0.510825624 | YES |
| PGS000740_hmPOS_GRCh37 | 1 | 78743005 | A | T | 0.029558802 | NO |
| PGS000740_hmPOS_GRCh37 | 1 | 154566225 | C | T | -0.051293294 | YES |
| PGS000740_hmPOS_GRCh37 | 1 | 168511081 | A | G | -0.051293294 | YES |
| PGS000740_hmPOS_GRCh37 | 2 | 38567201 | C | T | 0.039220713 | YES |
| PGS000740_hmPOS_GRCh37 | 2 | 45189737 | T | C | 0.048790164 | YES |
| PGS000740_hmPOS_GRCh37 | 2 | 67510377 | C | T | 0.048790164 | YES |
| PGS000740_hmPOS_GRCh37 | 2 | 119449740 | A | T | 0.1570037488096647 | NO |
| PGS000740_hmPOS_GRCh37 | 2 | 140398327 | C | T | -0.083381609 | YES |
| PGS000740_hmPOS_GRCh37 | 2 | 173983204 | A | G | -0.010050336 | NO |
| PGS000740_hmPOS_GRCh37 | 2 | 174075761 | G | A | -0.010050336 | YES |

| | | | | | | |
|---|---|---|---|---|---|---|
| PGS000740_hmPOS_GRCh37 | 4 | 67833774 | G | C | 0.058268908 | NO |
| PGS000740_hmPOS_GRCh37 | 4 | 164007992 | G | A | -0.020202707 | YES |
| PGS000740_hmPOS_GRCh37 | 5 | 1000156 | A | G | 0.086177696 | YES |
| PGS000740_hmPOS_GRCh37 | 5 | 1249816 | T | C | 0.067658648 | YES |
| PGS000740_hmPOS_GRCh37 | 5 | 1276873 | G | A | 0.019802627 | YES |
| PGS000740_hmPOS_GRCh37 | 5 | 1287194 | G | A | 0.058268908 | YES |
| PGS000740_hmPOS_GRCh37 | 5 | 1300269 | A | G | 0.048790164 | YES |
| PGS000740_hmPOS_GRCh37 | 5 | 90250631 | T | A | -0.040821995 | NO |
| PGS000740_hmPOS_GRCh37 | 5 | 150121458 | C | G | 0.131028262 | NO |
| PGS000740_hmPOS_GRCh37 | 6 | 410848 | A | G | -0.040821995 | YES |
| PGS000740_hmPOS_GRCh37 | 6 | 25885814 | T | C | 0 | YES |
| PGS000740_hmPOS_GRCh37 | 6 | 26198845 | C | G | 0 | NO |
| PGS000740_hmPOS_GRCh37 | 6 | 28217797 | T | G | -0.010050336 | NO |
| PGS000740_hmPOS_GRCh37 | 6 | 29223493 | C | G | -0.235722334 | NO |
| PGS000740_hmPOS_GRCh37 | 6 | 29477821 | T | G | 0.009950331 | YES |
| PGS000740_hmPOS_GRCh37 | 6 | 29606761 | A | G | 0 | YES |
| PGS000740_hmPOS_GRCh37 | 6 | 29759750 | A | G | -0.020202707 | YES |
| PGS000740_hmPOS_GRCh37 | 6 | 29922740 | G | A | -0.010050336 | YES |
| PGS000740_hmPOS_GRCh37 | 6 | 30074163 | A | G | 0 | YES |
| PGS000740_hmPOS_GRCh37 | 6 | 30864829 | C | T | 0 | YES |
| PGS000740_hmPOS_GRCh37 | 6 | 31002452 | G | A | -0.020202707 | YES |
| PGS000740_hmPOS_GRCh37 | 6 | 31067852 | G | A | 0.019802627 | YES |
| PGS000740_hmPOS_GRCh37 | 6 | 31081065 | C | T | 0.009950331 | YES |
| PGS000740_hmPOS_GRCh37 | 6 | 31117577 | G | C | 0 | NO |
| PGS000740_hmPOS_GRCh37 | 6 | 31321429 | A | T | -0.010050336 | NO |
| PGS000740_hmPOS_GRCh37 | 6 | 31322782 | C | G | -0.010050336 | NO |
| PGS000740_hmPOS_GRCh37 | 6 | 31324996 | G | C | 0.009950331 | NO |
| PGS000740_hmPOS_GRCh37 | 6 | 31412961 | C | T | 0.029558802 | YES |
| PGS000740_hmPOS_GRCh37 | 6 | 31840455 | A | G | 0 | YES |
| PGS000740_hmPOS_GRCh37 | 6 | 32335204 | A | G | 0 | YES |
| PGS000740_hmPOS_GRCh37 | 6 | 32609147 | T | A | -0.020202707 | NO |
| PGS000740_hmPOS_GRCh37 | 6 | 32757737 | G | A | -0.040821995 | YES |
| PGS000740_hmPOS_GRCh37 | 6 | 32783086 | G | A | 0.029558802 | YES |
| PGS000740_hmPOS_GRCh37 | 6 | 72384541 | A | G | 0.1570037488096647 | YES |
| PGS000740_hmPOS_GRCh37 | 6 | 117734267 | G | A | -0.010050336 | YES |
| PGS000740_hmPOS_GRCh37 | 7 | 130668618 | C | T | 0.019802627 | YES |
| PGS000740_hmPOS_GRCh37 | 8 | 27327841 | C | G | 0.029558802 | NO |
| PGS000740_hmPOS_GRCh37 | 8 | 27397087 | T | C | 0.029558802 | YES |
| PGS000740_hmPOS_GRCh37 | 8 | 144722420 | C | T | 0.039220713 | YES |
| PGS000740_hmPOS_GRCh37 | 9 | 6227418 | G | A | 0.076961041 | YES |
| PGS000740_hmPOS_GRCh37 | 9 | 17934120 | C | G | 0.019802627 | NO |
| PGS000740_hmPOS_GRCh37 | 9 | 21959751 | C | T | 0 | YES |
| PGS000740_hmPOS_GRCh37 | 9 | 22066572 | A | G | 0.1043600153242428 | YES |
| PGS000740_hmPOS_GRCh37 | 9 | 22083404 | C | T | 0 | YES |
| PGS000740_hmPOS_GRCh37 | 9 | 33427322 | C | A | -0.020202707 | YES |
| PGS000740_hmPOS_GRCh37 | 9 | 102587233 | T | C | 0.1906203596086497 | YES |
| PGS000740_hmPOS_GRCh37 | 9 | 136275229 | T | C | 0.086177696 | YES |
| PGS000740_hmPOS_GRCh37 | 10 | 102011702 | A | G | 0.058268908 | YES |

| | | | | | | |
|---|---|---|---|---|---|---|
| PGS000740_hmPOS_GRCh37 | 10 | 102672248 | T | A | 0.067658648 | NO |
| PGS000740_hmPOS_GRCh37 | 11 | 57250026 | T | C | -0.030459207 | YES |
| PGS000740_hmPOS_GRCh37 | 11 | 115998756 | A | G | -0.116533816 | YES |
| PGS000740_hmPOS_GRCh37 | 12 | 1036042 | T | C | -0.061875404 | YES |
| PGS000740_hmPOS_GRCh37 | 12 | 58831070 | A | G | 0 | YES |
| PGS000740_hmPOS_GRCh37 | 12 | 64913237 | T | G | 0.048790164 | YES |
| PGS000740_hmPOS_GRCh37 | 13 | 84281063 | T | A | 0.039220713 | NO |
| PGS000740_hmPOS_GRCh37 | 14 | 34013721 | A | G | -0.040821995 | YES |
| PGS000740_hmPOS_GRCh37 | 15 | 49253961 | C | A | -0.010050336 | YES |
| PGS000740_hmPOS_GRCh37 | 15 | 49615952 | G | C | 0.019802627 | NO |
| PGS000740_hmPOS_GRCh37 | 15 | 70431773 | T | G | 0.058268908 | YES |
| PGS000740_hmPOS_GRCh37 | 15 | 75055819 | T | C | 0.086177696 | YES |
| PGS000740_hmPOS_GRCh37 | 15 | 78880752 | A | G | 0.048790164 | YES |
| PGS000740_hmPOS_GRCh37 | 15 | 78909398 | T | C | 0.076961041 | YES |
| PGS000740_hmPOS_GRCh37 | 15 | 78981423 | G | A | -0.094310679 | YES |
| PGS000740_hmPOS_GRCh37 | 15 | 78987225 | A | G | 0.009950331 | YES |
| PGS000740_hmPOS_GRCh37 | 15 | 79058730 | G | A | 0 | YES |
| PGS000740_hmPOS_GRCh37 | 15 | 79065557 | G | A | -0.010050336 | YES |
| PGS000740_hmPOS_GRCh37 | 15 | 79075233 | C | T | 0 | YES |
| PGS000740_hmPOS_GRCh37 | 15 | 79110783 | T | C | 0 | YES |
| PGS000740_hmPOS_GRCh37 | 15 | 79198760 | G | T | -0.030459207 | YES |
| PGS000740_hmPOS_GRCh37 | 16 | 10740982 | C | G | -0.030459207 | NO |
| PGS000740_hmPOS_GRCh37 | 16 | 26980646 | C | T | 0.1988508587451651 | YES |
| PGS000740_hmPOS_GRCh37 | 17 | 70299958 | G | T | -0.030459207 | YES |
| PGS000740_hmPOS_GRCh37 | 18 | 20741135 | C | T | 0.039220713 | YES |
| PGS000740_hmPOS_GRCh37 | 19 | 41195170 | C | T | 0.009950331 | YES |
| PGS000740_hmPOS_GRCh37 | 19 | 41207206 | G | C | 0 | NO |
| PGS000740_hmPOS_GRCh37 | 19 | 41334199 | A | G | -0.072570693 | YES |
| PGS000740_hmPOS_GRCh37 | 19 | 41338988 | G | C | -0.010050336 | NO |
| PGS000740_hmPOS_GRCh37 | 19 | 41357457 | T | G | -0.15082289 | YES |
| PGS000740_hmPOS_GRCh37 | 19 | 41412192 | C | T | -0.105360516 | YES |
| PGS000740_hmPOS_GRCh37 | 19 | 54567858 | A | G | -0.010050336 | YES |
| PGS000740_hmPOS_GRCh37 | 20 | 61988382 | G | A | 0.086177696 | YES |
| PGS000740_hmPOS_GRCh37 | 20 | 62527305 | A | G | -0.094310679 | YES |

Abbreviations: SNP, Single nucleotide polymorphism; PRS, polygenic risk score.

Total number of variants: 128
Matched, n (%): 96 (75.00%)
Exclusions due to ambiguously matched, n (%): 23 (17.97%)
Exclusions due to unmatched, n (%): 9 (7.03%)

**eTable S10. List of SNPs included in endometrial cancer PRS**

| Accession | Chr name | Chr position | Effect allele | Other allele | Effect weight | Include |
|---|---|---|---|---|---|---|
| PGS000786_hmPOS_GRCh37 | 6 | 21649085 | G | A | 5.59 | YES |
| PGS000786_hmPOS_GRCh37 | 6 | 126010116 | A | G | 7.25 | YES |
| PGS000786_hmPOS_GRCh37 | 8 | 129599278 | C | G | 5.93 | NO |
| PGS000786_hmPOS_GRCh37 | 9 | 10260263 | T | C | 5.45 | YES |
| PGS000786_hmPOS_GRCh37 | 13 | 73811879 | A | T | 6.95 | NO |
| PGS000786_hmPOS_GRCh37 | 14 | 105243220 | A | G | 5.51 | YES |
| PGS000786_hmPOS_GRCh37 | 15 | 40322124 | C | T | 5.63 | YES |
| PGS000786_hmPOS_GRCh37 | 15 | 51537806 | C | T | 7.24 | YES |
| PGS000786_hmPOS_GRCh37 | 17 | 36103565 | A | G | 8.98 | YES |

Abbreviations: SNP, Single nucleotide polymorphism; PRS, polygenic risk score.

Total number of variants: 9
Matched, n (%): 7 (77.78%)
Exclusions due to ambiguously matched, n (%): 2 (22.2%)
Exclusions due to unmatched, n (%): 0 (0%)

**eTable S11. List of SNPs included in testicular cancer PRS**

| Accession | Chr name | Chr position | Effect allele | Other allele | Effect weight | Include |
|---|---|---|---|---|---|---|
| PGS000796_hmPOS_GRCh37 | 1 | 9713386 | T | C | 7.2 | YES |
| PGS000796_hmPOS_GRCh37 | 1 | 156169610 | G | A | 6.37 | YES |
| PGS000796_hmPOS_GRCh37 | 1 | 165873392 | C | T | 7.64 | YES |
| PGS000796_hmPOS_GRCh37 | 2 | 71572455 | C | T | 6.7 | YES |
| PGS000796_hmPOS_GRCh37 | 2 | 182377541 | T | C | 5.45 | YES |
| PGS000796_hmPOS_GRCh37 | 3 | 16625048 | A | G | 6.28 | YES |
| PGS000796_hmPOS_GRCh37 | 3 | 141818850 | C | T | 6.04 | YES |
| PGS000796_hmPOS_GRCh37 | 3 | 156300724 | C | T | 7.06 | YES |
| PGS000796_hmPOS_GRCh37 | 3 | 169756119 | T | C | 5.89 | YES |
| PGS000796_hmPOS_GRCh37 | 4 | 76520651 | G | A | 6.69 | YES |
| PGS000796_hmPOS_GRCh37 | 4 | 95224812 | T | G | 5.71 | YES |
| PGS000796_hmPOS_GRCh37 | 4 | 104054686 | A | G | 9.17 | YES |
| PGS000796_hmPOS_GRCh37 | 4 | 188921440 | A | G | 7.04 | YES |
| PGS000796_hmPOS_GRCh37 | 5 | 1286516 | A | C | 9.12 | YES |
| PGS000796_hmPOS_GRCh37 | 5 | 1308552 | A | G | 10.82 | YES |
| PGS000796_hmPOS_GRCh37 | 5 | 134366200 | T | C | 5.65 | YES |
| PGS000796_hmPOS_GRCh37 | 5 | 141681788 | A | G | 14.29 | YES |
| PGS000796_hmPOS_GRCh37 | 6 | 33542538 | G | A | 12.76 | YES |
| PGS000796_hmPOS_GRCh37 | 6 | 149972132 | G | A | 5.98 | YES |
| PGS000796_hmPOS_GRCh37 | 7 | 1968953 | C | T | 7.2 | YES |
| PGS000796_hmPOS_GRCh37 | 7 | 40920313 | G | C | 6.64 | NO |
| PGS000796_hmPOS_GRCh37 | 7 | 158501492 | T | C | 5.58 | YES |
| PGS000796_hmPOS_GRCh37 | 8 | 11611500 | C | G | 5.66 | NO |
| PGS000796_hmPOS_GRCh37 | 8 | 70976505 | C | T | 5.47 | YES |
| PGS000796_hmPOS_GRCh37 | 9 | 845516 | A | C | 14.14 | YES |
| PGS000796_hmPOS_GRCh37 | 9 | 878563 | G | A | 9.96 | YES |
| PGS000796_hmPOS_GRCh37 | 10 | 126274612 | G | C | 5.64 | NO |
| PGS000796_hmPOS_GRCh37 | 11 | 77997936 | T | C | 6.47 | YES |
| PGS000796_hmPOS_GRCh37 | 11 | 125071163 | G | A | 7.02 | YES |
| PGS000796_hmPOS_GRCh37 | 12 | 14653867 | C | T | 7.41 | YES |
| PGS000796_hmPOS_GRCh37 | 12 | 32141495 | A | G | 6.71 | YES |
| PGS000796_hmPOS_GRCh37 | 12 | 70563865 | A | G | 6.13 | YES |
| PGS000796_hmPOS_GRCh37 | 12 | 88953561 | A | C | 24.18 | YES |
| PGS000796_hmPOS_GRCh37 | 14 | 55880047 | G | A | 5.52 | YES |
| PGS000796_hmPOS_GRCh37 | 15 | 56038707 | A | G | 6.76 | YES |
| PGS000796_hmPOS_GRCh37 | 15 | 66821250 | G | A | 7.15 | YES |
| PGS000796_hmPOS_GRCh37 | 15 | 85605427 | G | T | 5.47 | YES |
| PGS000796_hmPOS_GRCh37 | 16 | 11920037 | A | G | 5.65 | YES |
| PGS000796_hmPOS_GRCh37 | 16 | 15530708 | T | G | 7.16 | NO |
| PGS000796_hmPOS_GRCh37 | 16 | 50142944 | G | A | 5.92 | YES |
| PGS000796_hmPOS_GRCh37 | 16 | 74670458 | C | T | 6.9 | YES |
| PGS000796_hmPOS_GRCh37 | 16 | 88549264 | G | C | 5.94 | NO |
| PGS000796_hmPOS_GRCh37 | 17 | 36101156 | C | T | 9.23 | YES |
| PGS000796_hmPOS_GRCh37 | 17 | 56632543 | T | G | 9.21 | YES |
| PGS000796_hmPOS_GRCh37 | 18 | 649311 | A | G | 6.24 | YES |

| | | | | | | |
|---|---|---|---|---|---|---|
| PGS000796_hmPOS_GRCh37 | 19 | 22267849 | T | G | 5.61 | YES |
| PGS000796_hmPOS_GRCh37 | 19 | 23205184 | G | A | 7.54 | YES |
| PGS000796_hmPOS_GRCh37 | 19 | 24050828 | T | A | 6.68 | NO |
| PGS000796_hmPOS_GRCh37 | 19 | 28257393 | G | A | 8.3 | YES |
| PGS000796_hmPOS_GRCh37 | 19 | 54284689 | T | G | 5.67 | YES |
| PGS000796_hmPOS_GRCh37 | 20 | 50708054 | T | A | 7 | NO |
| PGS000796_hmPOS_GRCh37 | 21 | 47690068 | T | C | 6.08 | YES |

Abbreviations: SNP, Single nucleotide polymorphism; PRS, polygenic risk score.

Total number of variants: 52
Matched, n (%): 45 (86.54%)
Exclusions due to ambiguously matched, n (%): 6 (11.54%)
Exclusions due to unmatched, n (%): 1 (1.92%)

**eTable S12. List of SNPs included in thyroid cancer PRS**

| Accession | Chr name | Chr position | Effect allele | Other allele | Effect weight | Include |
|---|---|---|---|---|---|---|
| PGS000797_hmPOS_GRCh37 | 1 | 233412561 | A | G | 6.6 | YES |
| PGS000797_hmPOS_GRCh37 | 2 | 218292158 | C | G | 10.23 | NO |
| PGS000797_hmPOS_GRCh37 | 3 | 169518455 | T | C | 5.56 | NO |
| PGS000797_hmPOS_GRCh37 | 5 | 111485904 | A | T | 6.3 | NO |
| PGS000797_hmPOS_GRCh37 | 8 | 32432796 | G | T | 8.53 | YES |
| PGS000797_hmPOS_GRCh37 | 9 | 4811553 | A | G | 5.63 | NO |
| PGS000797_hmPOS_GRCh37 | 9 | 100537802 | A | C | 16.12 | YES |
| PGS000797_hmPOS_GRCh37 | 10 | 105694301 | T | C | 6.56 | YES |
| PGS000797_hmPOS_GRCh37 | 14 | 36649246 | T | C | 9.01 | YES |
| PGS000797_hmPOS_GRCh37 | 14 | 36738361 | T | C | 8.29 | YES |
| PGS000797_hmPOS_GRCh37 | 15 | 67455630 | T | C | 5.85 | YES |
| PGS000797_hmPOS_GRCh37 | 15 | 67457485 | C | G | 5.93 | NO |

Abbreviations: SNP, Single nucleotide polymorphism; PRS, polygenic risk score.

Total number of variants: 12
Matched, n (%): 7 (58.33%)
Exclusions due to ambiguously matched, n (%): 3 (25.00%)
Exclusions due to unmatched, n (%): 2 (16.67%)

**eTable S13. List of SNPs included in kidney cancer PRS**

| Accession | Chr name | Chr position | Effect allele | Other allele | Effect weight | Include |
|---|---|---|---|---|---|---|
| PGS000787_hmPOS_GRCh37 | 1 | 50907438 | C | T | 6.29 | YES |
| PGS000787_hmPOS_GRCh37 | 1 | 165650787 | C | T | 5.59 | YES |
| PGS000787_hmPOS_GRCh37 | 2 | 46533376 | T | C | 7.9 | YES |
| PGS000787_hmPOS_GRCh37 | 2 | 46558208 | G | T | 5.84 | YES |
| PGS000787_hmPOS_GRCh37 | 2 | 46589295 | G | A | 6.34 | YES |
| PGS000787_hmPOS_GRCh37 | 2 | 145208193 | C | T | 5.89 | YES |
| PGS000787_hmPOS_GRCh37 | 3 | 40533243 | G | A | 5.57 | YES |
| PGS000787_hmPOS_GRCh37 | 3 | 169536637 | T | C | 5.75 | YES |
| PGS000787_hmPOS_GRCh37 | 3 | 172313367 | G | A | 5.49 | YES |
| PGS000787_hmPOS_GRCh37 | 4 | 101005318 | G | A | 5.86 | YES |
| PGS000787_hmPOS_GRCh37 | 8 | 22876739 | T | C | 5.82 | YES |
| PGS000787_hmPOS_GRCh37 | 10 | 105682296 | T | C | 5.49 | YES |
| PGS000787_hmPOS_GRCh37 | 11 | 69239741 | G | A | 9.65 | YES |
| PGS000787_hmPOS_GRCh37 | 11 | 108357137 | A | G | 6.36 | YES |
| PGS000787_hmPOS_GRCh37 | 12 | 26453283 | C | T | 8.21 | YES |
| PGS000787_hmPOS_GRCh37 | 12 | 125320850 | T | C | 6.99 | YES |
| PGS000787_hmPOS_GRCh37 | 14 | 73272730 | A | C | 6.55 | YES |
| PGS000787_hmPOS_GRCh37 | 14 | 73279420 | C | T | 10.19 | YES |
| PGS000787_hmPOS_GRCh37 | 22 | 47013535 | G | A | 5.58 | YES |

Abbreviations: SNP, Single nucleotide polymorphism; PRS, polygenic risk score.

Total number of variants: 19
Matched, n (%): 19 (100%)
Exclusions due to ambiguously matched, n (%): 0 (0%)
Exclusions due to unmatched, n (%): 0 (0%)

**eTable S14. List of SNPs included in multiple myeloma PRS**

| Accession | Chr name | Chr position | Effect allele | Other allele | Effect weight | Include |
|---|---|---|---|---|---|---|
| PGS000653_hmPOS_GRCh37 | 2 | 174808899 | T | C | 0.113328685 | YES |
| PGS000653_hmPOS_GRCh37 | 3 | 41786009 | C | T | 0.392042088 | YES |
| PGS000653_hmPOS_GRCh37 | 3 | 169492101 | C | T | 0.227932068 | YES |
| PGS000653_hmPOS_GRCh37 | 5 | 95242931 | T | C | 0.21511138 | YES |
| PGS000653_hmPOS_GRCh37 | 5 | 122743325 | T | A | 0.104360015 | NO |
| PGS000653_hmPOS_GRCh37 | 6 | 15244018 | G | C | 0.31481074 | NO |
| PGS000653_hmPOS_GRCh37 | 6 | 31107258 | T | C | 0.175632569 | YES |
| PGS000653_hmPOS_GRCh37 | 6 | 106667535 | G | T | 0.165514438 | YES |
| PGS000653_hmPOS_GRCh37 | 7 | 21938240 | C | A | 0.357674444 | YES |
| PGS000653_hmPOS_GRCh37 | 7 | 106291118 | C | T | 0.113328685 | YES |
| PGS000653_hmPOS_GRCh37 | 7 | 124583896 | T | C | 0.113328685 | YES |
| PGS000653_hmPOS_GRCh37 | 7 | 150950940 | A | G | 0.173953307 | YES |
| PGS000653_hmPOS_GRCh37 | 8 | 128222421 | C | T | 0.122217633 | YES |
| PGS000653_hmPOS_GRCh37 | 9 | 21991923 | C | T | 0.139761942 | YES |
| PGS000653_hmPOS_GRCh37 | 10 | 28856819 | G | A | 0.113328685 | YES |
| PGS000653_hmPOS_GRCh37 | 11 | 69462910 | G | A | 0.667829373 | YES |
| PGS000653_hmPOS_GRCh37 | 16 | 30700858 | C | T | 0.139761942 | YES |
| PGS000653_hmPOS_GRCh37 | 16 | 74664743 | T | C | 0.122217633 | YES |
| PGS000653_hmPOS_GRCh37 | 17 | 16820099 | A | G | 0.36 | YES |
| PGS000653_hmPOS_GRCh37 | 19 | 16438661 | T | A | 0.131028262 | NO |
| PGS000653_hmPOS_GRCh37 | 20 | 47355009 | C | T | 0.231111721 | YES |
| PGS000653_hmPOS_GRCh37 | 22 | 39542292 | A | G | 0.20538683 | YES |

Abbreviations: SNP, Single nucleotide polymorphism; PRS, polygenic risk score.

Total number of variants: 22
Matched, n (%): 19 (86.36%)
Exclusions due to ambiguously matched, n (%): 3 (13.64%)
Exclusions due to unmatched, n (%): 0 (0%)

**eTable S15. List of SNPs included in cervical cancer PRS**

| Accession | Chr name | Chr position | Effect allele | Other allele | Effect weight | Include |
|---|---|---|---|---|---|---|
| PGS000784_hmPOS_GRCh37 | 6 | 33063219 | C | T | 6.7 | YES |
| PGS000784_hmPOS_GRCh37 | 6 | 31480668 | G | A | 6.24 | YES |
| PGS000784_hmPOS_GRCh37 | 6 | 31331257 | G | A | 7.08 | YES |
| PGS000784_hmPOS_GRCh37 | 6 | 32594328 | C | T | 7.38 | YES |
| PGS000784_hmPOS_GRCh37 | 6 | 31347798 | G | A | 5.83 | YES |
| PGS000784_hmPOS_GRCh37 | 6 | 31540141 | A | C | 6.04 | YES |
| PGS000784_hmPOS_GRCh37 | 6 | 32595972 | G | A | 8.97 | YES |
| PGS000784_hmPOS_GRCh37 | 6 | 31382911 | A | G | 5.73 | YES |
| PGS000784_hmPOS_GRCh37 | 6 | 32311459 | G | A | 10.25 | YES |
| PGS000784_hmPOS_GRCh37 | 6 | 32584581 | C | T | 6.24 | YES |

Abbreviations: SNP, Single nucleotide polymorphism; PRS, polygenic risk score.

Total number of variants: 10
Matched, n (%): 10 (100%)
Exclusions due to ambiguously matched, n (%): 0 (0%)
Exclusions due to unmatched, n (%): 0 (0%)

**eTable S16. List of SNPs included in pancreatic cancer PRS**

| Accession | Chr name | Chr position | Effect allele | Other allele | Effect weight | Include |
|---|---|---|---|---|---|---|
| PGS000794_hmPOS_GRCh37 | 1 | 894573 | G | A | 7.46 | YES |
| PGS000794_hmPOS_GRCh37 | 1 | 199985368 | A | T | 7.88 | NO |
| PGS000794_hmPOS_GRCh37 | 1 | 200007432 | A | G | 8.06 | YES |
| PGS000794_hmPOS_GRCh37 | 2 | 67639769 | G | T | 5.91 | YES |
| PGS000794_hmPOS_GRCh37 | 3 | 189508471 | G | A | 5.58 | YES |
| PGS000794_hmPOS_GRCh37 | 5 | 1294086 | C | T | 7.79 | YES |
| PGS000794_hmPOS_GRCh37 | 5 | 1295373 | C | T | 5.64 | YES |
| PGS000794_hmPOS_GRCh37 | 5 | 1322087 | T | C | 8.31 | YES |
| PGS000794_hmPOS_GRCh37 | 7 | 40866663 | C | A | 5.71 | YES |
| PGS000794_hmPOS_GRCh37 | 7 | 47488569 | A | T | 5.48 | NO |
| PGS000794_hmPOS_GRCh37 | 7 | 130680521 | T | C | 7.48 | YES |
| PGS000794_hmPOS_GRCh37 | 8 | 76470404 | A | G | 6.18 | YES |
| PGS000794_hmPOS_GRCh37 | 8 | 128719884 | T | A | 6.08 | NO |
| PGS000794_hmPOS_GRCh37 | 9 | 106797388 | C | T | 5.54 | YES |
| PGS000794_hmPOS_GRCh37 | 9 | 136149229 | C | T | 10.73 | YES |
| PGS000794_hmPOS_GRCh37 | 13 | 28493997 | A | G | 7.53 | YES |
| PGS000794_hmPOS_GRCh37 | 13 | 73916628 | C | T | 9.79 | YES |
| PGS000794_hmPOS_GRCh37 | 16 | 75263661 | A | G | 6.77 | YES |
| PGS000794_hmPOS_GRCh37 | 17 | 36078510 | G | A | 5.68 | YES |
| PGS000794_hmPOS_GRCh37 | 17 | 70401476 | T | C | 7.75 | YES |
| PGS000794_hmPOS_GRCh37 | 18 | 56878274 | C | T | 5.53 | YES |
| PGS000794_hmPOS_GRCh37 | 22 | 29300306 | T | C | 5.7 | YES |

Abbreviations: SNP, Single nucleotide polymorphism; PRS, polygenic risk score.

Total number of variants: 22
Matched, n (%): 19 (86.36%)
Exclusions due to ambiguously matched, n (%): 3 (13.64%)
Exclusions due to unmatched, n (%): 0 (0%)

**eTable S17. List of SNPs included in stomach cancer PRS**

| Accession | Chr name | Chr position | Effect allele | Other allele | Effect weight | Include |
|---|---|---|---|---|---|---|
| PGS002299_hmPOS_GRCh37 | 1 | 155184975 | G | A | 0.235722334 | YES |
| PGS002299_hmPOS_GRCh37 | 5 | 40685795 | T | C | 0.210721031 | YES |
| PGS002299_hmPOS_GRCh37 | 8 | 143761931 | T | C | 0.19062036 | YES |

Abbreviations: SNP, Single nucleotide polymorphism; PRS, polygenic risk score.

Total number of variants: 3
Matched, n (%): 3 (100%)
Exclusions due to ambiguously matched, n (%): 0 (0%)
Exclusions due to unmatched, n (%): 0 (0%)

**eTable S18. List of SNPs included in lymphoid leukemia PRS**

| Accession | Chr name | Chr position | Effect allele | Other allele | Effect weight | Include |
|---|---|---|---|---|---|---|
| PGS000788_hmPOS_GRCh37 | 1 | 23943735 | C | A | 9.88 | YES |
| PGS000788_hmPOS_GRCh37 | 1 | 228880296 | G | A | 6.46 | YES |
| PGS000788_hmPOS_GRCh37 | 2 | 37596089 | T | C | 5.64 | YES |
| PGS000788_hmPOS_GRCh37 | 2 | 111600519 | C | T | 9.93 | YES |
| PGS000788_hmPOS_GRCh37 | 2 | 111616619 | C | T | 9.96 | YES |
| PGS000788_hmPOS_GRCh37 | 2 | 111797458 | G | A | 10.95 | YES |
| PGS000788_hmPOS_GRCh37 | 2 | 111927379 | G | A | 10.39 | YES |
| PGS000788_hmPOS_GRCh37 | 2 | 146124454 | A | C | 5.53 | YES |
| PGS000788_hmPOS_GRCh37 | 2 | 202023949 | A | G | 6.57 | YES |
| PGS000788_hmPOS_GRCh37 | 2 | 231091223 | G | T | 12.22 | YES |
| PGS000788_hmPOS_GRCh37 | 2 | 242294913 | G | A | 6.81 | YES |
| PGS000788_hmPOS_GRCh37 | 3 | 27777779 | T | C | 6.67 | YES |
| PGS000788_hmPOS_GRCh37 | 3 | 169492101 | C | T | 6.8 | YES |
| PGS000788_hmPOS_GRCh37 | 3 | 188115682 | C | A | 5.59 | YES |
| PGS000788_hmPOS_GRCh37 | 3 | 189401776 | G | C | 5.75 | NO |
| PGS000788_hmPOS_GRCh37 | 4 | 102741002 | C | T | 6.42 | YES |
| PGS000788_hmPOS_GRCh37 | 4 | 109016824 | A | C | 6.24 | YES |
| PGS000788_hmPOS_GRCh37 | 4 | 114683844 | C | G | 5.93 | NO |
| PGS000788_hmPOS_GRCh37 | 4 | 185254772 | T | C | 5.51 | YES |
| PGS000788_hmPOS_GRCh37 | 5 | 1279790 | T | C | 6.45 | YES |
| PGS000788_hmPOS_GRCh37 | 5 | 1285974 | A | C | 6.19 | YES |
| PGS000788_hmPOS_GRCh37 | 6 | 411064 | G | A | 12.56 | YES |
| PGS000788_hmPOS_GRCh37 | 6 | 442357 | G | A | 5.82 | YES |
| PGS000788_hmPOS_GRCh37 | 6 | 2969278 | T | C | 5.7 | YES |
| PGS000788_hmPOS_GRCh37 | 6 | 32257566 | A | G | 5.49 | YES |
| PGS000788_hmPOS_GRCh37 | 6 | 32578127 | G | A | 9.21 | YES |
| PGS000788_hmPOS_GRCh37 | 6 | 32611641 | G | A | 5.84 | YES |
| PGS000788_hmPOS_GRCh37 | 6 | 32626272 | A | C | 6.34 | YES |
| PGS000788_hmPOS_GRCh37 | 6 | 33546930 | C | T | 8.37 | YES |
| PGS000788_hmPOS_GRCh37 | 6 | 34616322 | C | G | 5.61 | NO |
| PGS000788_hmPOS_GRCh37 | 6 | 154478440 | C | A | 6.41 | YES |
| PGS000788_hmPOS_GRCh37 | 7 | 50418949 | T | C | 5.63 | YES |
| PGS000788_hmPOS_GRCh37 | 7 | 50477144 | A | G | 15.83 | YES |
| PGS000788_hmPOS_GRCh37 | 7 | 50546925 | C | T | 6.66 | YES |
| PGS000788_hmPOS_GRCh37 | 7 | 50567493 | A | G | 5.47 | YES |
| PGS000788_hmPOS_GRCh37 | 7 | 50576903 | A | G | 6.56 | YES |
| PGS000788_hmPOS_GRCh37 | 7 | 124462661 | C | T | 5.81 | YES |
| PGS000788_hmPOS_GRCh37 | 8 | 103578874 | G | T | 5.94 | YES |
| PGS000788_hmPOS_GRCh37 | 8 | 128192981 | G | A | 7.81 | YES |
| PGS000788_hmPOS_GRCh37 | 8 | 130194104 | A | C | 5.89 | YES |
| PGS000788_hmPOS_GRCh37 | 9 | 21970916 | A | G | 10.91 | YES |
| PGS000788_hmPOS_GRCh37 | 9 | 21984661 | G | T | 6.65 | YES |
| PGS000788_hmPOS_GRCh37 | 9 | 22044122 | C | T | 8.04 | YES |
| PGS000788_hmPOS_GRCh37 | 9 | 22060833 | A | G | 5.47 | YES |
| PGS000788_hmPOS_GRCh37 | 9 | 22206987 | C | T | 6.24 | YES |

| | | | | | | |
|---|---|---|---|---|---|---|
| PGS000788_hmPOS_GRCh37 | 10 | 8104208 | A | C | 7.64 | YES |
| PGS000788_hmPOS_GRCh37 | 10 | 22341574 | G | C | 5.48 | NO |
| PGS000788_hmPOS_GRCh37 | 10 | 22856279 | C | T | 6.88 | YES |
| PGS000788_hmPOS_GRCh37 | 10 | 63752159 | G | T | 13.87 | YES |
| PGS000788_hmPOS_GRCh37 | 10 | 63773039 | A | G | 6.75 | YES |
| PGS000788_hmPOS_GRCh37 | 10 | 90759724 | G | A | 8.96 | YES |
| PGS000788_hmPOS_GRCh37 | 10 | 126292655 | C | T | 6.87 | YES |
| PGS000788_hmPOS_GRCh37 | 11 | 2321650 | A | G | 6.56 | YES |
| PGS000788_hmPOS_GRCh37 | 11 | 113517203 | T | C | 6.68 | YES |
| PGS000788_hmPOS_GRCh37 | 11 | 123361397 | A | G | 16.62 | YES |
| PGS000788_hmPOS_GRCh37 | 11 | 123368333 | G | A | 8.1 | YES |
| PGS000788_hmPOS_GRCh37 | 12 | 96612762 | T | A | 5.76 | NO |
| PGS000788_hmPOS_GRCh37 | 12 | 113380008 | G | A | 5.58 | YES |
| PGS000788_hmPOS_GRCh37 | 12 | 117002658 | A | G | 5.69 | YES |
| PGS000788_hmPOS_GRCh37 | 14 | 23589349 | A | G | 9.48 | YES |
| PGS000788_hmPOS_GRCh37 | 14 | 92697912 | T | A | 5.82 | NO |
| PGS000788_hmPOS_GRCh37 | 15 | 40397936 | T | G | 6.17 | YES |
| PGS000788_hmPOS_GRCh37 | 15 | 56340896 | A | G | 6.43 | YES |
| PGS000788_hmPOS_GRCh37 | 15 | 56777691 | G | A | 8.73 | YES |
| PGS000788_hmPOS_GRCh37 | 15 | 70018990 | A | G | 12.07 | YES |
| PGS000788_hmPOS_GRCh37 | 16 | 85928621 | A | T | 11.1 | NO |
| PGS000788_hmPOS_GRCh37 | 16 | 85975659 | T | C | 6 | YES |
| PGS000788_hmPOS_GRCh37 | 18 | 47843534 | A | G | 5.53 | YES |
| PGS000788_hmPOS_GRCh37 | 18 | 57628926 | A | G | 5.83 | YES |
| PGS000788_hmPOS_GRCh37 | 18 | 60793549 | G | A | 6.99 | YES |
| PGS000788_hmPOS_GRCh37 | 18 | 60793921 | C | T | 7.18 | YES |
| PGS000788_hmPOS_GRCh37 | 19 | 4069119 | A | G | 5.46 | YES |
| PGS000788_hmPOS_GRCh37 | 19 | 47176752 | C | A | 5.52 | YES |
| PGS000788_hmPOS_GRCh37 | 19 | 47207654 | A | G | 5.89 | YES |
| PGS000788_hmPOS_GRCh37 | 22 | 50971266 | T | C | 5.95 | YES |

Abbreviations: SNP, Single nucleotide polymorphism; PRS, polygenic risk score.

Total number of variants: 75
Matched, n (%): 68 (90.67%)
Exclusions due to ambiguously matched, n (%): 7 (9.33%)
Exclusions due to unmatched, n (%): 0 (0%)

**eTable S19. List of SNPs included in bladder cancer PRS**

| Accession | Chr name | Chr position | Effect allele | Other allele | Effect weight | Include |
|---|---|---|---|---|---|---|
| PGS000782_hmPOS_GRCh37 | 2 | 234565283 | A | C | 6.25 | YES |
| PGS000782_hmPOS_GRCh37 | 3 | 169492101 | C | T | 5.86 | YES |
| PGS000782_hmPOS_GRCh37 | 3 | 189645933 | A | G | 6.71 | YES |
| PGS000782_hmPOS_GRCh37 | 4 | 1734239 | T | C | 10.3 | YES |
| PGS000782_hmPOS_GRCh37 | 5 | 1322087 | C | T | 6.59 | YES |
| PGS000782_hmPOS_GRCh37 | 8 | 18272881 | A | G | 6.6 | YES |
| PGS000782_hmPOS_GRCh37 | 8 | 128718068 | T | G | 12.92 | YES |
| PGS000782_hmPOS_GRCh37 | 8 | 143761931 | T | C | 7.9 | YES |
| PGS000782_hmPOS_GRCh37 | 11 | 1874072 | A | G | 5.49 | YES |
| PGS000782_hmPOS_GRCh37 | 13 | 113659108 | A | G | 6.28 | YES |
| PGS000782_hmPOS_GRCh37 | 18 | 43309911 | T | C | 6.51 | YES |
| PGS000782_hmPOS_GRCh37 | 19 | 30296853 | C | T | 6.78 | YES |
| PGS000782_hmPOS_GRCh37 | 20 | 10961935 | A | C | 6.75 | YES |
| PGS000782_hmPOS_GRCh37 | 20 | 10988099 | A | G | 6.69 | YES |
| PGS000782_hmPOS_GRCh37 | 22 | 39332623 | T | C | 6.83 | YES |

Abbreviations: SNP, Single nucleotide polymorphism; PRS, polygenic risk score.

Total number of variants: 15
Matched, n (%): 15 (100%)
Exclusions due to ambiguously matched, n (%): 0 (0%)
Exclusions due to unmatched, n (%): 0 (0%)

**eTable S20. List of SNPs included in Hodgkin's lymphoma PRS**

| Accession | Chr name | Chr position | Effect allele | Other allele | Effect weight | Include |
|---|---|---|---|---|---|---|
| PGS000639_hmPOS_GRCh37 | 1 | 114377568 | A | G | 0.19062036 | YES |
| PGS000639_hmPOS_GRCh37 | 2 | 61066666 | G | A | 0.21511138 | YES |
| PGS000639_hmPOS_GRCh37 | 3 | 27764623 | G | A | 0.231111721 | YES |
| PGS000639_hmPOS_GRCh37 | 3 | 187954414 | A | C | 0.262364264 | YES |
| PGS000639_hmPOS_GRCh37 | 5 | 131998413 | A | G | 0.463734016 | YES |
| PGS000639_hmPOS_GRCh37 | 6 | 32428285 | C | T | 0.494696242 | YES |
| PGS000639_hmPOS_GRCh37 | 6 | 33684313 | A | G | 0.254642218 | YES |
| PGS000639_hmPOS_GRCh37 | 6 | 128288536 | C | T | 0.182321557 | YES |
| PGS000639_hmPOS_GRCh37 | 6 | 135415004 | C | T | 0.19062036 | YES |
| PGS000639_hmPOS_GRCh37 | 6 | 135626348 | G | T | 0.207014169 | YES |
| PGS000639_hmPOS_GRCh37 | 6 | 137981584 | T | C | 0.173953307 | YES |
| PGS000639_hmPOS_GRCh37 | 8 | 129075832 | C | T | 0.182321557 | YES |
| PGS000639_hmPOS_GRCh37 | 8 | 129192271 | C | T | 0.262364264 | YES |
| PGS000639_hmPOS_GRCh37 | 10 | 8093034 | G | T | 0.329303747 | YES |
| PGS000639_hmPOS_GRCh37 | 10 | 8101927 | T | C | 0.329303747 | YES |
| PGS000639_hmPOS_GRCh37 | 11 | 111249611 | A | G | 0.173953307 | YES |
| PGS000639_hmPOS_GRCh37 | 13 | 115059729 | C | T | 0.329303747 | YES |
| PGS000639_hmPOS_GRCh37 | 16 | 11198938 | A | G | 0.21511138 | YES |
| PGS000639_hmPOS_GRCh37 | 16 | 30174926 | T | C | 0.148420005 | YES |
| PGS000639_hmPOS_GRCh37 | 20 | 44702120 | T | C | 0.139761942 | YES |

Abbreviations: SNP, Single nucleotide polymorphism; PRS, polygenic risk score.

Total number of variants: 20
Matched, n (%): 20 (100%)
Exclusions due to ambiguously matched, n (%): 0 (0%)
Exclusions due to unmatched, n (%): 0 (0%)

**eTable S21. List of SNPs included in larynx cancer PRS**

| Accession | Chr name | Chr position | Effect allele | Other allele | Effect weight | Include |
|---|---|---|---|---|---|---|
| PGS000362_hmPOS_GRCh37 | 1 | 10859513 | G | A | -0.444 | YES |
| PGS000362_hmPOS_GRCh37 | 1 | 32334427 | G | A | 1.85 | YES |
| PGS000362_hmPOS_GRCh37 | 1 | 67500797 | C | T | 2.29 | YES |
| PGS000362_hmPOS_GRCh37 | 1 | 94529355 | G | A | 2.11 | YES |
| PGS000362_hmPOS_GRCh37 | 1 | 98159558 | A | G | 0.39 | YES |
| PGS000362_hmPOS_GRCh37 | 1 | 157522111 | G | C | 2.43 | NO |
| PGS000362_hmPOS_GRCh37 | 2 | 38463396 | G | C | 1.52 | NO |
| PGS000362_hmPOS_GRCh37 | 2 | 61931907 | C | T | 0.506 | YES |
| PGS000362_hmPOS_GRCh37 | 2 | 142878715 | A | C | 1.5 | YES |
| PGS000362_hmPOS_GRCh37 | 3 | 19238171 | T | G | 0.888 | YES |
| PGS000362_hmPOS_GRCh37 | 3 | 195277690 | T | C | 1.18 | YES |
| PGS000362_hmPOS_GRCh37 | 4 | 8092688 | T | C | 1.5 | YES |
| PGS000362_hmPOS_GRCh37 | 4 | 117595443 | T | A | 1.24 | NO |
| PGS000362_hmPOS_GRCh37 | 4 | 146510243 | G | T | 1.57 | YES |
| PGS000362_hmPOS_GRCh37 | 5 | 14483164 | A | G | 0.435 | YES |
| PGS000362_hmPOS_GRCh37 | 5 | 31507079 | A | G | 1.47 | YES |
| PGS000362_hmPOS_GRCh37 | 6 | 23104359 | T | C | 1.12 | YES |
| PGS000362_hmPOS_GRCh37 | 6 | 44498186 | C | T | 0.398 | YES |
| PGS000362_hmPOS_GRCh37 | 6 | 52133731 | G | A | 1.56 | YES |
| PGS000362_hmPOS_GRCh37 | 6 | 74145883 | T | C | 1.49 | YES |
| PGS000362_hmPOS_GRCh37 | 6 | 103690786 | C | T | -0.599 | YES |
| PGS000362_hmPOS_GRCh37 | 6 | 126441138 | G | A | 0.752 | YES |
| PGS000362_hmPOS_GRCh37 | 7 | 47597180 | G | T | 0.852 | YES |
| PGS000362_hmPOS_GRCh37 | 7 | 54131046 | T | C | 0.507 | YES |
| PGS000362_hmPOS_GRCh37 | 7 | 113297775 | C | G | 0.579 | NO |
| PGS000362_hmPOS_GRCh37 | 8 | 14509228 | G | T | 0.894 | YES |
| PGS000362_hmPOS_GRCh37 | 8 | 16925030 | T | C | 1.5 | YES |
| PGS000362_hmPOS_GRCh37 | 8 | 30395392 | A | G | 1.07 | YES |
| PGS000362_hmPOS_GRCh37 | 9 | 122465539 | C | A | 1.11 | YES |
| PGS000362_hmPOS_GRCh37 | 10 | 17347299 | A | T | 0.403 | NO |
| PGS000362_hmPOS_GRCh37 | 10 | 29819017 | T | C | 1.9 | YES |
| PGS000362_hmPOS_GRCh37 | 10 | 116317908 | G | A | -1.06 | YES |
| PGS000362_hmPOS_GRCh37 | 10 | 133519852 | C | G | 1.21 | NO |
| PGS000362_hmPOS_GRCh37 | 11 | 4481261 | T | A | -0.387 | NO |
| PGS000362_hmPOS_GRCh37 | 11 | 61942541 | T | G | 0.454 | YES |
| PGS000362_hmPOS_GRCh37 | 11 | 92966435 | A | G | 0.421 | YES |
| PGS000362_hmPOS_GRCh37 | 11 | 106368253 | T | C | 1.82 | YES |
| PGS000362_hmPOS_GRCh37 | 11 | 125823353 | T | C | 0.5 | YES |
| PGS000362_hmPOS_GRCh37 | 12 | 4945348 | T | C | 0.978 | YES |
| PGS000362_hmPOS_GRCh37 | 13 | 24008773 | A | G | 0.431 | YES |
| PGS000362_hmPOS_GRCh37 | 13 | 32933022 | T | G | 0.397 | YES |
| PGS000362_hmPOS_GRCh37 | 14 | 100032934 | T | C | 1.58 | YES |
| PGS000362_hmPOS_GRCh37 | 15 | 94110918 | A | G | -0.71 | YES |
| PGS000362_hmPOS_GRCh37 | 16 | 393948 | T | C | 0.411 | YES |
| PGS000362_hmPOS_GRCh37 | 16 | 12249506 | A | G | 0.778 | YES |

| | | | | | | |
|---|---|---|---|---|---|---|
| PGS000362_hmPOS_GRCh37 | 16 | 82771075 | A | G | 0.475 | YES |
| PGS000362_hmPOS_GRCh37 | 17 | 78514637 | G | A | 0.551 | YES |
| PGS000362_hmPOS_GRCh37 | 18 | 65333614 | G | A | -0.535 | YES |
| PGS000362_hmPOS_GRCh37 | 19 | 53404512 | C | T | 2.33 | YES |
| PGS000362_hmPOS_GRCh37 | 20 | 4682314 | T | C | 2.35 | YES |
| PGS000362_hmPOS_GRCh37 | 20 | 18955117 | C | G | 1.81 | NO |
| PGS000362_hmPOS_GRCh37 | 20 | 58450616 | A | G | 1.64 | YES |
| PGS000362_hmPOS_GRCh37 | 21 | 21853777 | T | C | -0.42 | YES |

Abbreviations: SNP, Single nucleotide polymorphism; PRS, polygenic risk score.

Total number of variants: 53
Matched, n (%): 45 (84.91%)
Exclusions due to ambiguously matched, n (%): 8 (15.09%)
Exclusions due to unmatched, n (%): 0 (0%)

**eTable S22. List of SNPs included in tracheal cancer PRS**

| Accession | Chr name | Chr position | Effect allele | Other allele | Effect weight | Include |
|---|---|---|---|---|---|---|
| PGS000392_hmPOS_GRCh37 | 1 | 77967507 | A | T | 0.127764345 | NO |
| PGS000392_hmPOS_GRCh37 | 3 | 189357199 | G | T | 0.110739354 | YES |
| PGS000392_hmPOS_GRCh37 | 5 | 1285974 | A | C | 0.222222087 | YES |
| PGS000392_hmPOS_GRCh37 | 5 | 1320247 | G | A | 0.141287474 | YES |
| PGS000392_hmPOS_GRCh37 | 6 | 26285867 | C | T | 0.075844863 | YES |
| PGS000392_hmPOS_GRCh37 | 6 | 29781049 | T | G | 0.095199628 | YES |
| PGS000392_hmPOS_GRCh37 | 6 | 31434111 | G | A | 0.223154591 | YES |
| PGS000392_hmPOS_GRCh37 | 8 | 27344719 | G | A | 0.141137604 | YES |
| PGS000392_hmPOS_GRCh37 | 8 | 32410110 | G | A | 0.124309867 | YES |
| PGS000392_hmPOS_GRCh37 | 9 | 21830157 | G | A | 0.15454663 | YES |
| PGS000392_hmPOS_GRCh37 | 9 | 22052068 | G | A | 0.165877847 | YES |
| PGS000392_hmPOS_GRCh37 | 10 | 105687632 | C | A | 0.150557805 | YES |
| PGS000392_hmPOS_GRCh37 | 11 | 118125625 | T | C | 0.102138631 | YES |
| PGS000392_hmPOS_GRCh37 | 12 | 998819 | G | C | 0.145612966 | NO |
| PGS000392_hmPOS_GRCh37 | 13 | 32972626 | T | A | 0.472187431 | NO |
| PGS000392_hmPOS_GRCh37 | 15 | 49376624 | T | G | 0.155057925 | YES |
| PGS000392_hmPOS_GRCh37 | 15 | 78857986 | G | C | 0.260378679 | NO |
| PGS000392_hmPOS_GRCh37 | 15 | 79185180 | G | A | 0.091838889 | YES |
| PGS000392_hmPOS_GRCh37 | 19 | 41353107 | C | T | 0.122792512 | YES |

Abbreviations: SNP, Single nucleotide polymorphism; PRS, polygenic risk score.

Total number of variants: 19
Matched, n (%): 15 (78.95%)
Exclusions due to ambiguously matched, n (%): 4 (21.05%)
Exclusions due to unmatched, n (%): 0 (0%)

**eTable S23. List of SNPs included in esophageal cancer PRS**

| Accession | Chr name | Chr position | Effect allele | Other allele | Effect weight | Include |
|---|---|---|---|---|---|---|
| PGS000364_hmPOS_GRCh37 | 1 | 11410483 | G | T | 0.004512395 | YES |
| PGS000364_hmPOS_GRCh37 | 1 | 11411071 | T | G | 0.004049102 | YES |
| PGS000364_hmPOS_GRCh37 | 1 | 11411432 | A | G | 0.004203736 | YES |
| PGS000364_hmPOS_GRCh37 | 1 | 15718366 | G | T | 0.000846096 | YES |
| PGS000364_hmPOS_GRCh37 | 1 | 20903408 | C | T | 0.011614661 | YES |
| PGS000364_hmPOS_GRCh37 | 1 | 20973655 | G | T | 0.001973367 | YES |
| PGS000364_hmPOS_GRCh37 | 1 | 30504582 | A | G | 0.000574702 | YES |
| PGS000364_hmPOS_GRCh37 | 1 | 31568123 | C | T | 0.001405238 | YES |
| PGS000364_hmPOS_GRCh37 | 1 | 31568962 | C | T | 0.002386453 | YES |
| PGS000364_hmPOS_GRCh37 | 1 | 31569772 | T | C | 0.001036376 | YES |
| PGS000364_hmPOS_GRCh37 | 1 | 31572571 | C | T | 0.000261527 | YES |
| PGS000364_hmPOS_GRCh37 | 1 | 31573531 | C | G | 0.000341948 | NO |
| PGS000364_hmPOS_GRCh37 | 1 | 31573640 | T | C | 0.00011493 | YES |
| PGS000364_hmPOS_GRCh37 | 1 | 31573885 | T | C | 0.000129741 | YES |
| PGS000364_hmPOS_GRCh37 | 1 | 31574031 | T | C | 0.000134305 | YES |
| PGS000364_hmPOS_GRCh37 | 1 | 31574982 | T | G | 1.85513E-05 | YES |
| PGS000364_hmPOS_GRCh37 | 1 | 34249144 | G | C | 0.001651071 | NO |
| PGS000364_hmPOS_GRCh37 | 1 | 34249160 | G | C | 0.001608022 | NO |
| PGS000364_hmPOS_GRCh37 | 1 | 34250018 | C | T | 0.005243709 | YES |
| PGS000364_hmPOS_GRCh37 | 1 | 34251481 | C | T | 0.010521591 | YES |
| PGS000364_hmPOS_GRCh37 | 1 | 34251754 | C | T | 0.01035266 | YES |
| PGS000364_hmPOS_GRCh37 | 1 | 34252019 | G | A | 0.009523849 | YES |
| PGS000364_hmPOS_GRCh37 | 1 | 34253041 | C | G | 0.004966635 | NO |
| PGS000364_hmPOS_GRCh37 | 1 | 34253141 | T | G | 0.004337845 | YES |
| PGS000364_hmPOS_GRCh37 | 1 | 38749491 | G | C | -0.003992458 | NO |
| PGS000364_hmPOS_GRCh37 | 1 | 43464391 | G | A | 0.000212649 | YES |
| PGS000364_hmPOS_GRCh37 | 1 | 43464635 | C | T | 0.00019099 | YES |
| PGS000364_hmPOS_GRCh37 | 1 | 43464697 | G | T | 6.16799E-05 | YES |
| PGS000364_hmPOS_GRCh37 | 1 | 44855878 | A | G | 0.000156893 | YES |
| PGS000364_hmPOS_GRCh37 | 1 | 44855942 | A | G | 0.013681187 | YES |
| PGS000364_hmPOS_GRCh37 | 1 | 58787119 | T | C | 0.005382391 | YES |
| PGS000364_hmPOS_GRCh37 | 1 | 58789343 | A | G | 0.005409216 | YES |
| PGS000364_hmPOS_GRCh37 | 1 | 58793895 | A | G | 0.004613325 | YES |
| PGS000364_hmPOS_GRCh37 | 1 | 71023538 | T | C | 0.0030005 | YES |
| PGS000364_hmPOS_GRCh37 | 1 | 71037431 | T | C | 0.004318786 | YES |
| PGS000364_hmPOS_GRCh37 | 1 | 71175583 | C | G | 0.003960897 | NO |
| PGS000364_hmPOS_GRCh37 | 1 | 71184053 | A | G | 0.002569862 | YES |
| PGS000364_hmPOS_GRCh37 | 1 | 71256068 | C | T | 6.40692E-05 | YES |
| PGS000364_hmPOS_GRCh37 | 1 | 71702864 | C | T | 0.002703838 | YES |
| PGS000364_hmPOS_GRCh37 | 1 | 71909921 | T | A | 0.01326197 | NO |
| PGS000364_hmPOS_GRCh37 | 1 | 71996378 | T | C | 0.002458081 | YES |
| PGS000364_hmPOS_GRCh37 | 1 | 72986136 | T | G | 0.000347144 | YES |
| PGS000364_hmPOS_GRCh37 | 1 | 73006137 | T | C | 0.029545529 | YES |
| PGS000364_hmPOS_GRCh37 | 1 | 73008966 | C | T | 0.024821385 | YES |
| PGS000364_hmPOS_GRCh37 | 1 | 81534528 | T | C | 0.003214837 | YES |
| PGS000364_hmPOS_GRCh37 | 1 | 81577225 | A | T | 0.00276969 | NO |
| PGS000364_hmPOS_GRCh37 | 1 | 81613271 | C | A | 0.002112725 | YES |
| PGS000364_hmPOS_GRCh37 | 1 | 92178335 | G | C | -0.001303759 | NO |
| PGS000364_hmPOS_GRCh37 | 1 | 92180390 | G | C | -0.001103526 | NO |
| PGS000364_hmPOS_GRCh37 | 1 | 95777223 | T | C | 7.360127040279e-6 | YES |
| PGS000364_hmPOS_GRCh37 | 1 | 95780019 | G | A | 0.000255565 | YES |
| PGS000364_hmPOS_GRCh37 | 1 | 102537880 | A | G | 0.003260179 | YES |
| PGS000364_hmPOS_GRCh37 | 1 | 102538360 | G | T | 0.004620788 | YES |
| PGS000364_hmPOS_GRCh37 | 1 | 102561485 | G | C | 0.00578853 | NO |
| PGS000364_hmPOS_GRCh37 | 1 | 102566184 | C | A | 0.005825664 | YES |
| PGS000364_hmPOS_GRCh37 | 1 | 102569709 | G | A | 0.005865617 | YES |
| PGS000364_hmPOS_GRCh37 | 1 | 102590737 | T | C | 0.007014334 | YES |
| PGS000364_hmPOS_GRCh37 | 1 | 102595955 | T | C | 0.007266251 | YES |
| PGS000364_hmPOS_GRCh37 | 1 | 102597251 | C | T | 0.00731557 | YES |
| PGS000364_hmPOS_GRCh37 | 1 | 102597496 | A | C | 0.006490954 | YES |

| | | | | | | |
|---|---|---|---|---|---|---|
| PGS000364_hmPOS_GRCh37 | 1 | 102598108 | T | A | 0.00742713 | NO |
| PGS000364_hmPOS_GRCh37 | 1 | 102598469 | G | C | 0.007408903 | NO |
| PGS000364_hmPOS_GRCh37 | 1 | 102598488 | T | C | 0.011718703 | YES |
| PGS000364_hmPOS_GRCh37 | 1 | 102599506 | G | T | 0.007360117 | YES |
| PGS000364_hmPOS_GRCh37 | 1 | 102599540 | T | G | 0.006465627 | YES |
| PGS000364_hmPOS_GRCh37 | 1 | 102600822 | G | A | 0.007414083 | YES |
| PGS000364_hmPOS_GRCh37 | 1 | 102605045 | A | G | 0.007718607 | YES |
| PGS000364_hmPOS_GRCh37 | 1 | 102605420 | A | G | 0.008571526 | YES |
| PGS000364_hmPOS_GRCh37 | 1 | 102607447 | T | C | 0.008697671 | YES |
| PGS000364_hmPOS_GRCh37 | 1 | 102607563 | T | C | 0.008051858 | YES |
| PGS000364_hmPOS_GRCh37 | 1 | 102607815 | G | T | 0.008554174 | YES |
| PGS000364_hmPOS_GRCh37 | 1 | 102607883 | A | C | 0.008638247 | YES |
| PGS000364_hmPOS_GRCh37 | 1 | 102610378 | T | G | 0.008419604 | YES |
| PGS000364_hmPOS_GRCh37 | 1 | 102610446 | G | A | 0.0077388 | YES |
| PGS000364_hmPOS_GRCh37 | 1 | 102610524 | C | A | 0.00834689 | YES |
| PGS000364_hmPOS_GRCh37 | 1 | 102611292 | G | A | 0.008476011 | YES |
| PGS000364_hmPOS_GRCh37 | 1 | 102612004 | T | A | 0.008485075 | NO |
| PGS000364_hmPOS_GRCh37 | 1 | 102613797 | A | G | 0.009462913 | YES |
| PGS000364_hmPOS_GRCh37 | 1 | 102621502 | G | A | 0.013622128 | YES |
| PGS000364_hmPOS_GRCh37 | 1 | 102626414 | G | A | 0.013448457 | YES |
| PGS000364_hmPOS_GRCh37 | 1 | 102629663 | C | T | 0.013253639 | YES |
| PGS000364_hmPOS_GRCh37 | 1 | 107488481 | C | T | 0.005512904 | YES |
| PGS000364_hmPOS_GRCh37 | 1 | 107488772 | A | G | 0.007049696 | YES |
| PGS000364_hmPOS_GRCh37 | 1 | 107489766 | C | T | 0.010311184 | YES |
| PGS000364_hmPOS_GRCh37 | 1 | 107668721 | A | T | 0.003098843 | NO |
| PGS000364_hmPOS_GRCh37 | 1 | 116711962 | T | G | 0.001762362 | YES |
| PGS000364_hmPOS_GRCh37 | 1 | 116712641 | A | G | 0.001350477 | YES |
| PGS000364_hmPOS_GRCh37 | 1 | 116716836 | T | C | 0.002877892 | YES |
| PGS000364_hmPOS_GRCh37 | 1 | 116716977 | A | T | 0.002748799 | NO |
| PGS000364_hmPOS_GRCh37 | 1 | 116717419 | A | G | 0.003157179 | YES |
| PGS000364_hmPOS_GRCh37 | 1 | 116719908 | A | G | 0.001967906 | YES |
| PGS000364_hmPOS_GRCh37 | 1 | 116720866 | C | A | 0.001750223 | YES |
| PGS000364_hmPOS_GRCh37 | 1 | 116721566 | G | A | 0.001830361 | YES |
| PGS000364_hmPOS_GRCh37 | 1 | 116722687 | A | G | 0.00123975 | YES |
| PGS000364_hmPOS_GRCh37 | 1 | 116753658 | A | G | 0.005674348 | YES |
| PGS000364_hmPOS_GRCh37 | 1 | 116769998 | A | G | 0.007459768 | YES |
| PGS000364_hmPOS_GRCh37 | 1 | 116778750 | C | T | 0.007262814 | YES |
| PGS000364_hmPOS_GRCh37 | 1 | 116780234 | A | G | 0.007636175 | YES |
| PGS000364_hmPOS_GRCh37 | 1 | 116787096 | C | T | 0.006924097 | YES |
| PGS000364_hmPOS_GRCh37 | 1 | 161680868 | C | T | 0.003275618 | YES |
| PGS000364_hmPOS_GRCh37 | 1 | 164573681 | C | A | -0.006400019 | YES |
| PGS000364_hmPOS_GRCh37 | 1 | 164575121 | A | G | -0.0056701 | YES |
| PGS000364_hmPOS_GRCh37 | 1 | 164575854 | T | C | -0.006003085 | YES |
| PGS000364_hmPOS_GRCh37 | 1 | 165690515 | A | T | 0.001987041 | NO |
| PGS000364_hmPOS_GRCh37 | 1 | 175276630 | C | T | -0.00192677 | YES |
| PGS000364_hmPOS_GRCh37 | 1 | 182476185 | T | C | 0.006522026 | YES |
| PGS000364_hmPOS_GRCh37 | 1 | 188764506 | C | T | 0.010859532 | YES |
| PGS000364_hmPOS_GRCh37 | 1 | 203402315 | T | C | -0.00979253 | YES |
| PGS000364_hmPOS_GRCh37 | 1 | 205281255 | T | G | -0.001887755 | YES |
| PGS000364_hmPOS_GRCh37 | 1 | 212329535 | T | C | 0.000918981 | YES |
| PGS000364_hmPOS_GRCh37 | 1 | 212333314 | T | G | 0.000290392 | YES |
| PGS000364_hmPOS_GRCh37 | 1 | 218055182 | C | T | 0.00948692 | YES |
| PGS000364_hmPOS_GRCh37 | 1 | 238131997 | C | T | 0.005264892 | YES |
| PGS000364_hmPOS_GRCh37 | 1 | 238344901 | C | A | -0.005052302 | YES |
| PGS000364_hmPOS_GRCh37 | 1 | 238345361 | T | C | -0.005048836 | YES |
| PGS000364_hmPOS_GRCh37 | 1 | 238351005 | G | A | -0.005025405 | YES |
| PGS000364_hmPOS_GRCh37 | 1 | 238351334 | A | G | -0.004796863 | YES |
| PGS000364_hmPOS_GRCh37 | 1 | 238352551 | A | C | -0.004767355 | YES |
| PGS000364_hmPOS_GRCh37 | 1 | 238359520 | A | G | -0.004160043 | YES |
| PGS000364_hmPOS_GRCh37 | 1 | 238361422 | G | C | -0.004911916 | NO |
| PGS000364_hmPOS_GRCh37 | 1 | 238363110 | A | C | -0.004894841 | YES |
| PGS000364_hmPOS_GRCh37 | 1 | 238363373 | G | A | -0.003707959 | YES |
| PGS000364_hmPOS_GRCh37 | 1 | 238364867 | G | C | -0.000564641 | NO |

| | | | | | | |
|---|---|---|---|---|---|---|
| PGS000364_hmPOS_GRCh37 | 1 | 238740359 | A | G | 0.010099318 | YES |
| PGS000364_hmPOS_GRCh37 | 1 | 238742904 | G | A | 0.008039003 | YES |
| PGS000364_hmPOS_GRCh37 | 1 | 241571995 | C | T | 0.021924302 | YES |
| PGS000364_hmPOS_GRCh37 | 1 | 245481598 | T | C | 0.008965311 | YES |
| PGS000364_hmPOS_GRCh37 | 1 | 247184302 | A | G | 0.007554718 | YES |
| PGS000364_hmPOS_GRCh37 | 2 | 4652574 | G | C | 0.003305219 | NO |
| PGS000364_hmPOS_GRCh37 | 2 | 7693201 | C | T | 0.009943001 | YES |
| PGS000364_hmPOS_GRCh37 | 2 | 7693398 | C | G | 0.010193728 | NO |
| PGS000364_hmPOS_GRCh37 | 2 | 7693578 | A | G | 0.010123312 | NO |
| PGS000364_hmPOS_GRCh37 | 2 | 7693796 | G | T | 0.009998622 | YES |
| PGS000364_hmPOS_GRCh37 | 2 | 7694032 | C | G | 0.009820861 | NO |
| PGS000364_hmPOS_GRCh37 | 2 | 7695163 | A | C | 0.005896068 | YES |
| PGS000364_hmPOS_GRCh37 | 2 | 7695234 | G | T | 0.004350586 | YES |
| PGS000364_hmPOS_GRCh37 | 2 | 7695856 | A | G | 0.004605871 | YES |
| PGS000364_hmPOS_GRCh37 | 2 | 7696479 | C | T | 0.004263361 | YES |
| PGS000364_hmPOS_GRCh37 | 2 | 7697371 | G | A | 0.005293258 | YES |
| PGS000364_hmPOS_GRCh37 | 2 | 7697461 | A | T | 0.004263159 | NO |
| PGS000364_hmPOS_GRCh37 | 2 | 7698069 | A | T | 0.004213653 | NO |
| PGS000364_hmPOS_GRCh37 | 2 | 7700390 | G | A | 0.004447495 | YES |
| PGS000364_hmPOS_GRCh37 | 2 | 7700861 | A | G | 0.004447703 | YES |
| PGS000364_hmPOS_GRCh37 | 2 | 7701025 | T | C | 0.004219652 | YES |
| PGS000364_hmPOS_GRCh37 | 2 | 7701498 | C | T | 0.004550183 | YES |
| PGS000364_hmPOS_GRCh37 | 2 | 7701637 | T | A | 0.004656633 | NO |
| PGS000364_hmPOS_GRCh37 | 2 | 7702036 | T | C | 0.004374062 | YES |
| PGS000364_hmPOS_GRCh37 | 2 | 7702949 | A | T | 0.004821377 | NO |
| PGS000364_hmPOS_GRCh37 | 2 | 7703442 | G | C | 0.005607125 | NO |
| PGS000364_hmPOS_GRCh37 | 2 | 7704185 | T | C | 0.006196806 | YES |
| PGS000364_hmPOS_GRCh37 | 2 | 7704998 | G | A | 0.006460348 | YES |
| PGS000364_hmPOS_GRCh37 | 2 | 7705237 | T | C | 0.006195912 | YES |
| PGS000364_hmPOS_GRCh37 | 2 | 7705812 | T | C | 0.006197324 | YES |
| PGS000364_hmPOS_GRCh37 | 2 | 7705841 | A | T | 0.006579045 | NO |
| PGS000364_hmPOS_GRCh37 | 2 | 7706063 | C | T | 0.006152595 | YES |
| PGS000364_hmPOS_GRCh37 | 2 | 7706296 | G | A | 0.006261134 | YES |
| PGS000364_hmPOS_GRCh37 | 2 | 7706355 | C | A | 0.006465182 | YES |
| PGS000364_hmPOS_GRCh37 | 2 | 7707691 | A | G | 0.00646538 | YES |
| PGS000364_hmPOS_GRCh37 | 2 | 7708677 | G | T | 0.006465528 | YES |
| PGS000364_hmPOS_GRCh37 | 2 | 7708781 | C | T | 0.006207595 | YES |
| PGS000364_hmPOS_GRCh37 | 2 | 7709715 | A | G | 0.006483176 | YES |
| PGS000364_hmPOS_GRCh37 | 2 | 7709746 | T | C | 0.006207783 | YES |
| PGS000364_hmPOS_GRCh37 | 2 | 7710141 | T | C | 0.006473529 | YES |
| PGS000364_hmPOS_GRCh37 | 2 | 7710274 | A | G | 0.006346993 | YES |
| PGS000364_hmPOS_GRCh37 | 2 | 7710698 | A | G | 0.006789025 | YES |
| PGS000364_hmPOS_GRCh37 | 2 | 7710836 | T | C | 0.006347863 | YES |
| PGS000364_hmPOS_GRCh37 | 2 | 7711220 | T | C | 0.006261134 | YES |
| PGS000364_hmPOS_GRCh37 | 2 | 7713284 | C | T | 0.006279539 | YES |
| PGS000364_hmPOS_GRCh37 | 2 | 7714640 | T | C | 0.006317347 | YES |
| PGS000364_hmPOS_GRCh37 | 2 | 7714913 | C | T | 0.005142471 | YES |
| PGS000364_hmPOS_GRCh37 | 2 | 7717019 | A | G | 0.013365382 | YES |
| PGS000364_hmPOS_GRCh37 | 2 | 7717192 | C | A | 0.006271187 | YES |
| PGS000364_hmPOS_GRCh37 | 2 | 7718791 | A | G | 0.006402898 | YES |
| PGS000364_hmPOS_GRCh37 | 2 | 7720688 | G | A | 0.006665413 | YES |
| PGS000364_hmPOS_GRCh37 | 2 | 7721162 | A | G | 0.006509925 | YES |
| PGS000364_hmPOS_GRCh37 | 2 | 7722245 | C | A | 0.006051231 | YES |
| PGS000364_hmPOS_GRCh37 | 2 | 7723328 | G | A | 0.006595334 | YES |
| PGS000364_hmPOS_GRCh37 | 2 | 7723685 | T | C | 0.006858343 | YES |
| PGS000364_hmPOS_GRCh37 | 2 | 7724374 | G | C | 0.006690166 | NO |
| PGS000364_hmPOS_GRCh37 | 2 | 7724739 | T | C | 0.006995198 | YES |
| PGS000364_hmPOS_GRCh37 | 2 | 7728010 | T | A | 0.002807057 | NO |
| PGS000364_hmPOS_GRCh37 | 2 | 7729461 | T | G | 0.005937868 | YES |
| PGS000364_hmPOS_GRCh37 | 2 | 7731894 | A | G | 0.006026376 | YES |
| PGS000364_hmPOS_GRCh37 | 2 | 7732198 | C | T | 0.003071328 | YES |
| PGS000364_hmPOS_GRCh37 | 2 | 7732257 | A | G | 0.005785858 | YES |
| PGS000364_hmPOS_GRCh37 | 2 | 7732403 | A | G | 0.003128202 | YES |

| | | | | | | |
|---|---|---|---|---|---|---|
| PGS000364_hmPOS_GRCh37 | 2 | 7732825 | T | A | 0.003049534 | NO |
| PGS000364_hmPOS_GRCh37 | 2 | 7733033 | A | G | 0.003059368 | YES |
| PGS000364_hmPOS_GRCh37 | 2 | 7733287 | G | A | 0.003127316 | YES |
| PGS000364_hmPOS_GRCh37 | 2 | 7734831 | C | T | 0.003154817 | YES |
| PGS000364_hmPOS_GRCh37 | 2 | 7735110 | T | A | 0.00314793 | NO |
| PGS000364_hmPOS_GRCh37 | 2 | 7737783 | C | T | 0.003581448 | YES |
| PGS000364_hmPOS_GRCh37 | 2 | 7738378 | T | C | 0.003657849 | YES |
| PGS000364_hmPOS_GRCh37 | 2 | 7738514 | G | A | 0.003679537 | YES |
| PGS000364_hmPOS_GRCh37 | 2 | 7739207 | G | T | 0.003816296 | YES |
| PGS000364_hmPOS_GRCh37 | 2 | 7739957 | C | T | 0.00576209 | YES |
| PGS000364_hmPOS_GRCh37 | 2 | 7741423 | T | C | 0.002755521 | YES |
| PGS000364_hmPOS_GRCh37 | 2 | 7741817 | G | A | 0.002719923 | YES |
| PGS000364_hmPOS_GRCh37 | 2 | 7742585 | G | T | 0.002675154 | YES |
| PGS000364_hmPOS_GRCh37 | 2 | 7742949 | G | A | 0.002493228 | YES |
| PGS000364_hmPOS_GRCh37 | 2 | 7743134 | A | G | 0.002686616 | YES |
| PGS000364_hmPOS_GRCh37 | 2 | 7744151 | A | C | 0.00272173 | YES |
| PGS000364_hmPOS_GRCh37 | 2 | 7745033 | C | T | 0.002760353 | YES |
| PGS000364_hmPOS_GRCh37 | 2 | 7745192 | C | G | 0.002760119 | NO |
| PGS000364_hmPOS_GRCh37 | 2 | 7746242 | T | C | 0.002779355 | YES |
| PGS000364_hmPOS_GRCh37 | 2 | 7746617 | A | G | 0.002779329 | YES |
| PGS000364_hmPOS_GRCh37 | 2 | 7746969 | T | C | 0.002779016 | YES |
| PGS000364_hmPOS_GRCh37 | 2 | 7747191 | A | G | 0.002778703 | YES |
| PGS000364_hmPOS_GRCh37 | 2 | 7747471 | C | T | 0.002778651 | YES |
| PGS000364_hmPOS_GRCh37 | 2 | 7748030 | G | A | 0.002005894 | YES |
| PGS000364_hmPOS_GRCh37 | 2 | 7748374 | A | G | 0.002022602 | YES |
| PGS000364_hmPOS_GRCh37 | 2 | 7748757 | T | C | 0.001709006 | YES |
| PGS000364_hmPOS_GRCh37 | 2 | 7749131 | C | T | 0.001890583 | YES |
| PGS000364_hmPOS_GRCh37 | 2 | 7749562 | T | C | 0.001891122 | YES |
| PGS000364_hmPOS_GRCh37 | 2 | 7749808 | A | T | 0.001809617 | NO |
| PGS000364_hmPOS_GRCh37 | 2 | 7750549 | G | A | 0.002068546 | YES |
| PGS000364_hmPOS_GRCh37 | 2 | 7750601 | A | G | 0.002068546 | YES |
| PGS000364_hmPOS_GRCh37 | 2 | 7750776 | T | C | 0.002068546 | YES |
| PGS000364_hmPOS_GRCh37 | 2 | 7751128 | T | C | 0.001406004 | YES |
| PGS000364_hmPOS_GRCh37 | 2 | 7751300 | A | G | 0.002073081 | YES |
| PGS000364_hmPOS_GRCh37 | 2 | 7751342 | G | A | 0.002072479 | YES |
| PGS000364_hmPOS_GRCh37 | 2 | 7751706 | A | T | 0.000719831 | NO |
| PGS000364_hmPOS_GRCh37 | 2 | 7751782 | A | G | 0.001645638 | YES |
| PGS000364_hmPOS_GRCh37 | 2 | 7751874 | A | T | 0.001640425 | NO |
| PGS000364_hmPOS_GRCh37 | 2 | 7751923 | C | T | 0.001640425 | YES |
| PGS000364_hmPOS_GRCh37 | 2 | 7752234 | G | C | 0.001640985 | NO |
| PGS000364_hmPOS_GRCh37 | 2 | 7752255 | A | G | 0.001640985 | YES |
| PGS000364_hmPOS_GRCh37 | 2 | 7752313 | C | T | 0.001640985 | YES |
| PGS000364_hmPOS_GRCh37 | 2 | 7752411 | G | A | 0.001640985 | YES |
| PGS000364_hmPOS_GRCh37 | 2 | 7752472 | A | G | 0.001608365 | YES |
| PGS000364_hmPOS_GRCh37 | 2 | 7752587 | C | T | 0.001608237 | YES |
| PGS000364_hmPOS_GRCh37 | 2 | 7753327 | G | T | 0.001607616 | YES |
| PGS000364_hmPOS_GRCh37 | 2 | 7753354 | T | C | 0.001556208 | YES |
| PGS000364_hmPOS_GRCh37 | 2 | 7753456 | G | T | 0.0016304 | YES |
| PGS000364_hmPOS_GRCh37 | 2 | 7753616 | C | T | 0.001625437 | YES |
| PGS000364_hmPOS_GRCh37 | 2 | 7753854 | C | G | 0.001588011 | NO |
| PGS000364_hmPOS_GRCh37 | 2 | 7753880 | G | A | 0.00159461 | YES |
| PGS000364_hmPOS_GRCh37 | 2 | 7753964 | G | A | 0.001588865 | YES |
| PGS000364_hmPOS_GRCh37 | 2 | 7754174 | C | T | 0.001572599 | YES |
| PGS000364_hmPOS_GRCh37 | 2 | 7754215 | C | T | 0.001547658 | YES |
| PGS000364_hmPOS_GRCh37 | 2 | 7754417 | C | T | 0.001562602 | YES |
| PGS000364_hmPOS_GRCh37 | 2 | 7754458 | A | G | 0.001562602 | YES |
| PGS000364_hmPOS_GRCh37 | 2 | 7754657 | G | T | 0.001817462 | YES |
| PGS000364_hmPOS_GRCh37 | 2 | 7754699 | G | A | 0.001562432 | YES |
| PGS000364_hmPOS_GRCh37 | 2 | 7754842 | G | C | 0.001600553 | NO |
| PGS000364_hmPOS_GRCh37 | 2 | 7755076 | A | G | 0.001600981 | YES |
| PGS000364_hmPOS_GRCh37 | 2 | 7755388 | T | C | 0.00162939 | YES |
| PGS000364_hmPOS_GRCh37 | 2 | 7755632 | G | A | 0.001589143 | YES |
| PGS000364_hmPOS_GRCh37 | 2 | 7755670 | A | G | 0.001589143 | YES |

| | | | | | | |
|---|---|---|---|---|---|---|
| PGS000364_hmPOS_GRCh37 | 2 | 7755837 | T | C | 0.001714845 | YES |
| PGS000364_hmPOS_GRCh37 | 2 | 7756035 | T | C | 0.001698452 | YES |
| PGS000364_hmPOS_GRCh37 | 2 | 7756042 | G | A | 0.001713908 | YES |
| PGS000364_hmPOS_GRCh37 | 2 | 7757047 | C | T | 0.00194779 | YES |
| PGS000364_hmPOS_GRCh37 | 2 | 7758261 | C | G | 0.002397785 | NO |
| PGS000364_hmPOS_GRCh37 | 2 | 7758980 | G | C | 0.002494569 | NO |
| PGS000364_hmPOS_GRCh37 | 2 | 7759149 | G | A | 0.002464678 | YES |
| PGS000364_hmPOS_GRCh37 | 2 | 7759930 | T | C | 0.002404879 | YES |
| PGS000364_hmPOS_GRCh37 | 2 | 7762018 | C | T | 0.002108113 | YES |
| PGS000364_hmPOS_GRCh37 | 2 | 7762704 | A | G | 0.002368377 | YES |
| PGS000364_hmPOS_GRCh37 | 2 | 7763768 | A | C | 0.002428594 | YES |
| PGS000364_hmPOS_GRCh37 | 2 | 7767116 | A | G | 0.002755443 | YES |
| PGS000364_hmPOS_GRCh37 | 2 | 7771210 | A | G | 0.001040915 | YES |
| PGS000364_hmPOS_GRCh37 | 2 | 7775304 | C | A | 0.002019436 | YES |
| PGS000364_hmPOS_GRCh37 | 2 | 7783425 | T | C | 0.001831384 | YES |
| PGS000364_hmPOS_GRCh37 | 2 | 7797021 | T | C | 0.012672146 | YES |
| PGS000364_hmPOS_GRCh37 | 2 | 7800453 | T | C | 0.01286635 | YES |
| PGS000364_hmPOS_GRCh37 | 2 | 15310579 | A | G | 0.001227214 | YES |
| PGS000364_hmPOS_GRCh37 | 2 | 15310606 | A | G | 0.001282563 | YES |
| PGS000364_hmPOS_GRCh37 | 2 | 15310820 | T | C | 0.001433149 | YES |
| PGS000364_hmPOS_GRCh37 | 2 | 15311885 | A | G | 0.006699624 | YES |
| PGS000364_hmPOS_GRCh37 | 2 | 15313341 | T | C | 0.005117098 | YES |
| PGS000364_hmPOS_GRCh37 | 2 | 15313634 | T | C | 0.005103948 | YES |
| PGS000364_hmPOS_GRCh37 | 2 | 15314180 | T | C | 0.005097617 | YES |
| PGS000364_hmPOS_GRCh37 | 2 | 15323165 | C | A | 0.005397383 | YES |
| PGS000364_hmPOS_GRCh37 | 2 | 15332877 | C | T | 0.005709275 | YES |
| PGS000364_hmPOS_GRCh37 | 2 | 15345668 | T | C | 0.002015583 | YES |
| PGS000364_hmPOS_GRCh37 | 2 | 15346445 | A | G | 0.002435847 | YES |
| PGS000364_hmPOS_GRCh37 | 2 | 15347328 | C | T | 0.00528058 | YES |
| PGS000364_hmPOS_GRCh37 | 2 | 18281737 | A | G | 0.014833307 | YES |
| PGS000364_hmPOS_GRCh37 | 2 | 21319066 | T | G | 0.00479631 | YES |
| PGS000364_hmPOS_GRCh37 | 2 | 21384070 | C | A | 0.01014993 | YES |
| PGS000364_hmPOS_GRCh37 | 2 | 21391669 | T | C | 0.004659726 | YES |
| PGS000364_hmPOS_GRCh37 | 2 | 40614266 | C | T | 0.002315342 | YES |
| PGS000364_hmPOS_GRCh37 | 2 | 40615007 | A | G | 0.001170397 | YES |
| PGS000364_hmPOS_GRCh37 | 2 | 40615384 | C | T | 0.001023633 | YES |
| PGS000364_hmPOS_GRCh37 | 2 | 40615386 | G | C | 0.001012256 | NO |
| PGS000364_hmPOS_GRCh37 | 2 | 40615444 | G | C | 0.001100731 | NO |
| PGS000364_hmPOS_GRCh37 | 2 | 40615896 | T | A | 0.001133692 | NO |
| PGS000364_hmPOS_GRCh37 | 2 | 40616346 | T | C | 0.001024916 | YES |
| PGS000364_hmPOS_GRCh37 | 2 | 40617126 | G | T | 0.00442258 | YES |
| PGS000364_hmPOS_GRCh37 | 2 | 40618588 | C | T | 0.003338439 | YES |
| PGS000364_hmPOS_GRCh37 | 2 | 40619230 | G | A | 0.004367355 | YES |
| PGS000364_hmPOS_GRCh37 | 2 | 40619660 | G | T | 0.000694379 | YES |
| PGS000364_hmPOS_GRCh37 | 2 | 40621139 | C | A | 0.010509834 | YES |
| PGS000364_hmPOS_GRCh37 | 2 | 40683536 | C | A | 0.008100922 | YES |
| PGS000364_hmPOS_GRCh37 | 2 | 46337172 | A | C | -0.001841039 | YES |
| PGS000364_hmPOS_GRCh37 | 2 | 59096806 | A | G | 0.009115244 | YES |
| PGS000364_hmPOS_GRCh37 | 2 | 59097683 | C | A | 0.001940722 | YES |
| PGS000364_hmPOS_GRCh37 | 2 | 59100050 | A | G | 0.01196856 | YES |
| PGS000364_hmPOS_GRCh37 | 2 | 59708647 | T | G | 0.001727984 | YES |
| PGS000364_hmPOS_GRCh37 | 2 | 59735942 | G | C | 0.017373782 | NO |
| PGS000364_hmPOS_GRCh37 | 2 | 59774394 | A | G | 0.013144624 | YES |
| PGS000364_hmPOS_GRCh37 | 2 | 60106395 | G | A | 0.010231273 | YES |
| PGS000364_hmPOS_GRCh37 | 2 | 60777498 | C | G | 0.00323238 | NO |
| PGS000364_hmPOS_GRCh37 | 2 | 64154797 | T | A | 0.000292838 | NO |
| PGS000364_hmPOS_GRCh37 | 2 | 64197149 | A | C | 0.006732026 | YES |
| PGS000364_hmPOS_GRCh37 | 2 | 64200508 | T | C | 0.00676139 | YES |
| PGS000364_hmPOS_GRCh37 | 2 | 70885377 | A | G | 0.003729068 | YES |
| PGS000364_hmPOS_GRCh37 | 2 | 71827555 | T | C | 0.009899249 | YES |
| PGS000364_hmPOS_GRCh37 | 2 | 76571471 | A | G | 0.00320289 | YES |
| PGS000364_hmPOS_GRCh37 | 2 | 76579521 | C | T | 0.002957806 | YES |
| PGS000364_hmPOS_GRCh37 | 2 | 76584023 | T | A | 0.001306548 | NO |

| | | | | | | |
|---|---|---|---|---|---|---|
| PGS000364_hmPOS_GRCh37 | 2 | 76585748 | C | T | 0.001141693 | YES |
| PGS000364_hmPOS_GRCh37 | 2 | 76587733 | C | T | 0.002354091 | YES |
| PGS000364_hmPOS_GRCh37 | 2 | 76602785 | C | A | 0.001856846 | YES |
| PGS000364_hmPOS_GRCh37 | 2 | 76606274 | G | A | 0.00262785 | YES |
| PGS000364_hmPOS_GRCh37 | 2 | 76612231 | A | C | 0.004071843 | YES |
| PGS000364_hmPOS_GRCh37 | 2 | 76613163 | T | A | 0.004731437 | NO |
| PGS000364_hmPOS_GRCh37 | 2 | 76614533 | G | C | 0.003099228 | NO |
| PGS000364_hmPOS_GRCh37 | 2 | 76615798 | C | T | 0.003116291 | YES |
| PGS000364_hmPOS_GRCh37 | 2 | 102677765 | A | G | -0.001998963 | YES |
| PGS000364_hmPOS_GRCh37 | 2 | 111447499 | A | G | -0.000239318 | YES |
| PGS000364_hmPOS_GRCh37 | 2 | 120021014 | A | G | 0.016429925 | YES |
| PGS000364_hmPOS_GRCh37 | 2 | 120088893 | A | G | 0.0138631 | YES |
| PGS000364_hmPOS_GRCh37 | 2 | 120096766 | C | T | 0.010070881 | YES |
| PGS000364_hmPOS_GRCh37 | 2 | 120116915 | A | G | -0.005844654 | YES |
| PGS000364_hmPOS_GRCh37 | 2 | 120135008 | C | G | 0.007808283 | NO |
| PGS000364_hmPOS_GRCh37 | 2 | 120147188 | G | A | 0.014951175 | YES |
| PGS000364_hmPOS_GRCh37 | 2 | 120152982 | G | A | -0.008055765 | YES |
| PGS000364_hmPOS_GRCh37 | 2 | 125280649 | T | C | 0.003801786 | NO |
| PGS000364_hmPOS_GRCh37 | 2 | 125281910 | T | C | 0.004480917 | YES |
| PGS000364_hmPOS_GRCh37 | 2 | 125284632 | G | A | 0.008969528 | YES |
| PGS000364_hmPOS_GRCh37 | 2 | 127910410 | G | A | 0.002489031 | YES |
| PGS000364_hmPOS_GRCh37 | 2 | 127926992 | A | G | 0.010915671 | YES |
| PGS000364_hmPOS_GRCh37 | 2 | 145295331 | C | T | 0.001837032 | YES |
| PGS000364_hmPOS_GRCh37 | 2 | 150279882 | T | C | -0.003953662 | YES |
| PGS000364_hmPOS_GRCh37 | 2 | 150313726 | G | A | -0.003179962 | YES |
| PGS000364_hmPOS_GRCh37 | 2 | 150407582 | T | C | -0.006137298 | YES |
| PGS000364_hmPOS_GRCh37 | 2 | 150410565 | G | A | -0.005817733 | YES |
| PGS000364_hmPOS_GRCh37 | 2 | 151002345 | C | T | 0.00595805 | YES |
| PGS000364_hmPOS_GRCh37 | 2 | 162361899 | G | A | 0.015540191 | YES |
| PGS000364_hmPOS_GRCh37 | 2 | 162497467 | T | C | 0.015944992 | YES |
| PGS000364_hmPOS_GRCh37 | 2 | 169110394 | T | C | 0.004775314 | YES |
| PGS000364_hmPOS_GRCh37 | 2 | 169117025 | T | G | 0.005330936 | YES |
| PGS000364_hmPOS_GRCh37 | 2 | 169119609 | G | C | 0.005252157 | NO |
| PGS000364_hmPOS_GRCh37 | 2 | 169120541 | T | A | 0.005332998 | NO |
| PGS000364_hmPOS_GRCh37 | 2 | 169129145 | T | G | 0.00353374 | YES |
| PGS000364_hmPOS_GRCh37 | 2 | 169133796 | G | A | 0.010267597 | YES |
| PGS000364_hmPOS_GRCh37 | 2 | 169143035 | A | G | 0.002391628 | YES |
| PGS000364_hmPOS_GRCh37 | 2 | 169150494 | A | G | 0.00395637 | YES |
| PGS000364_hmPOS_GRCh37 | 2 | 169150881 | A | G | 0.00396498 | YES |
| PGS000364_hmPOS_GRCh37 | 2 | 169154841 | A | G | 0.010842683 | YES |
| PGS000364_hmPOS_GRCh37 | 2 | 169155595 | G | T | 0.003716913 | YES |
| PGS000364_hmPOS_GRCh37 | 2 | 169166282 | C | T | 0.017150025 | YES |
| PGS000364_hmPOS_GRCh37 | 2 | 173219252 | T | C | -0.021622276 | YES |
| PGS000364_hmPOS_GRCh37 | 2 | 173219523 | A | G | -0.022621957 | YES |
| PGS000364_hmPOS_GRCh37 | 2 | 173219858 | T | C | -0.023303082 | YES |
| PGS000364_hmPOS_GRCh37 | 2 | 173220116 | C | T | -0.021490162 | YES |
| PGS000364_hmPOS_GRCh37 | 2 | 174235452 | G | T | 0.00209114 | YES |
| PGS000364_hmPOS_GRCh37 | 2 | 174241632 | A | C | 0.009710141 | YES |
| PGS000364_hmPOS_GRCh37 | 2 | 174242984 | T | G | 0.001319822 | YES |
| PGS000364_hmPOS_GRCh37 | 2 | 174275519 | G | A | 0.000320605 | YES |
| PGS000364_hmPOS_GRCh37 | 2 | 174275650 | T | G | 0.000142823 | YES |
| PGS000364_hmPOS_GRCh37 | 2 | 174276685 | T | C | 0.000300578 | YES |
| PGS000364_hmPOS_GRCh37 | 2 | 178605665 | G | T | 0.003623767 | YES |
| PGS000364_hmPOS_GRCh37 | 2 | 178613409 | G | A | 0.00381919 | YES |
| PGS000364_hmPOS_GRCh37 | 2 | 178625226 | A | G | 0.005492947 | YES |
| PGS000364_hmPOS_GRCh37 | 2 | 178627440 | T | C | 0.004332607 | YES |
| PGS000364_hmPOS_GRCh37 | 2 | 188637405 | G | A | 0.001628788 | YES |
| PGS000364_hmPOS_GRCh37 | 2 | 193428110 | T | A | -6.7107E-05 | NO |
| PGS000364_hmPOS_GRCh37 | 2 | 206981677 | T | A | 0.00012208 | NO |
| PGS000364_hmPOS_GRCh37 | 2 | 207004923 | G | A | 0.004065378 | YES |
| PGS000364_hmPOS_GRCh37 | 2 | 207007930 | T | C | 0.004074671 | YES |
| PGS000364_hmPOS_GRCh37 | 2 | 207011836 | T | C | 0.004036036 | YES |
| PGS000364_hmPOS_GRCh37 | 2 | 207017586 | A | G | 0.004019873 | YES |

| | | | | | | |
|---|---|---|---|---|---|---|
| PGS000364_hmPOS_GRCh37 | 2 | 207019118 | A | G | 0.00402445 | YES |
| PGS000364_hmPOS_GRCh37 | 2 | 207020304 | A | G | 0.003975969 | YES |
| PGS000364_hmPOS_GRCh37 | 2 | 207027903 | T | C | 0.000949081 | YES |
| PGS000364_hmPOS_GRCh37 | 2 | 217494921 | A | G | 0.006062619 | YES |
| PGS000364_hmPOS_GRCh37 | 2 | 236685678 | C | G | -0.003023879 | NO |
| PGS000364_hmPOS_GRCh37 | 2 | 240496929 | A | G | 0.002260005 | YES |
| PGS000364_hmPOS_GRCh37 | 2 | 240609925 | C | T | 0.002071808 | YES |
| PGS000364_hmPOS_GRCh37 | 3 | 2608424 | A | G | 0.008811903 | YES |
| PGS000364_hmPOS_GRCh37 | 3 | 3048016 | T | C | -0.000729894 | YES |
| PGS000364_hmPOS_GRCh37 | 3 | 3868538 | G | C | 0.004205898 | NO |
| PGS000364_hmPOS_GRCh37 | 3 | 3920408 | A | G | 0.021194843 | YES |
| PGS000364_hmPOS_GRCh37 | 3 | 4056290 | A | C | 0.028881788 | YES |
| PGS000364_hmPOS_GRCh37 | 3 | 4058564 | T | C | 0.029099701 | YES |
| PGS000364_hmPOS_GRCh37 | 3 | 4065339 | T | C | 0.02728389 | YES |
| PGS000364_hmPOS_GRCh37 | 3 | 4066089 | C | A | 0.028993441 | YES |
| PGS000364_hmPOS_GRCh37 | 3 | 4066168 | G | T | 0.025150923 | YES |
| PGS000364_hmPOS_GRCh37 | 3 | 4078391 | G | A | 0.020996176 | YES |
| PGS000364_hmPOS_GRCh37 | 3 | 5337049 | T | C | -0.001485292 | YES |
| PGS000364_hmPOS_GRCh37 | 3 | 5337140 | T | C | -0.003569776 | YES |
| PGS000364_hmPOS_GRCh37 | 3 | 5337538 | T | C | -0.001353165 | YES |
| PGS000364_hmPOS_GRCh37 | 3 | 5340118 | T | C | -0.000555711 | YES |
| PGS000364_hmPOS_GRCh37 | 3 | 6732150 | C | T | 0.006212596 | YES |
| PGS000364_hmPOS_GRCh37 | 3 | 6756286 | A | C | 0.013913875 | YES |
| PGS000364_hmPOS_GRCh37 | 3 | 6758905 | G | A | 0.011292461 | YES |
| PGS000364_hmPOS_GRCh37 | 3 | 12468410 | A | G | -0.001678159 | NO |
| PGS000364_hmPOS_GRCh37 | 3 | 12622623 | C | G | 0.003165191 | NO |
| PGS000364_hmPOS_GRCh37 | 3 | 21034193 | G | A | 0.004310178 | YES |
| PGS000364_hmPOS_GRCh37 | 3 | 22587535 | G | T | -7.7593E-05 | YES |
| PGS000364_hmPOS_GRCh37 | 3 | 22587564 | G | A | -7.96724E-05 | YES |
| PGS000364_hmPOS_GRCh37 | 3 | 22587653 | G | A | -0.000104914 | YES |
| PGS000364_hmPOS_GRCh37 | 3 | 22587743 | C | T | -9.22303E-05 | YES |
| PGS000364_hmPOS_GRCh37 | 3 | 22587894 | G | A | -8.80804E-05 | YES |
| PGS000364_hmPOS_GRCh37 | 3 | 22588115 | T | A | -6.68081E-05 | NO |
| PGS000364_hmPOS_GRCh37 | 3 | 25751549 | A | G | 0.001526981 | YES |
| PGS000364_hmPOS_GRCh37 | 3 | 31869036 | G | A | 0.000991923 | YES |
| PGS000364_hmPOS_GRCh37 | 3 | 38374375 | A | G | 0.016845388 | YES |
| PGS000364_hmPOS_GRCh37 | 3 | 38379604 | G | A | 0.019657824 | YES |
| PGS000364_hmPOS_GRCh37 | 3 | 38390273 | C | T | 0.016288684 | YES |
| PGS000364_hmPOS_GRCh37 | 3 | 38422897 | A | G | 0.001591641 | YES |
| PGS000364_hmPOS_GRCh37 | 3 | 38422907 | A | G | 0.001078588 | YES |
| PGS000364_hmPOS_GRCh37 | 3 | 38425746 | A | G | 0.000138806 | YES |
| PGS000364_hmPOS_GRCh37 | 3 | 38425808 | T | C | 7.88738E-05 | YES |
| PGS000364_hmPOS_GRCh37 | 3 | 38441543 | A | C | 0.000825561 | YES |
| PGS000364_hmPOS_GRCh37 | 3 | 38447793 | C | T | 0.003142964 | YES |
| PGS000364_hmPOS_GRCh37 | 3 | 38457783 | C | T | 0.003093011 | YES |
| PGS000364_hmPOS_GRCh37 | 3 | 38464688 | T | C | 0.001068653 | YES |
| PGS000364_hmPOS_GRCh37 | 3 | 38474109 | G | A | 0.002938168 | YES |
| PGS000364_hmPOS_GRCh37 | 3 | 38486696 | G | T | 0.002974868 | YES |
| PGS000364_hmPOS_GRCh37 | 3 | 38492856 | G | A | 0.000566064 | YES |
| PGS000364_hmPOS_GRCh37 | 3 | 38492866 | T | C | 0.000518659 | YES |
| PGS000364_hmPOS_GRCh37 | 3 | 38497081 | C | A | 0.002588565 | YES |
| PGS000364_hmPOS_GRCh37 | 3 | 38500647 | G | C | 0.00112584 | NO |
| PGS000364_hmPOS_GRCh37 | 3 | 38504388 | G | A | 0.003068042 | YES |
| PGS000364_hmPOS_GRCh37 | 3 | 42742507 | A | G | 0.001942828 | YES |
| PGS000364_hmPOS_GRCh37 | 3 | 42872590 | A | G | 0.000402763 | YES |
| PGS000364_hmPOS_GRCh37 | 3 | 42877394 | T | C | 0.000217769 | YES |
| PGS000364_hmPOS_GRCh37 | 3 | 42877413 | G | A | 0.001885646 | YES |
| PGS000364_hmPOS_GRCh37 | 3 | 42877711 | G | A | 0.000363367 | YES |
| PGS000364_hmPOS_GRCh37 | 3 | 42890731 | C | T | 0.001848703 | YES |
| PGS000364_hmPOS_GRCh37 | 3 | 42896515 | A | T | 9.22637E-05 | NO |
| PGS000364_hmPOS_GRCh37 | 3 | 42896639 | G | A | 7.95726E-05 | YES |
| PGS000364_hmPOS_GRCh37 | 3 | 42905465 | T | C | 0.001019262 | YES |
| PGS000364_hmPOS_GRCh37 | 3 | 42913610 | C | T | 0.006216231 | YES |

| | | | | | | |
|---|---|---|---|---|---|---|
| PGS000364_hmPOS_GRCh37 | 3 | 42914876 | T | G | 0.000851364 | YES |
| PGS000364_hmPOS_GRCh37 | 3 | 42917906 | C | T | 0.007449067 | YES |
| PGS000364_hmPOS_GRCh37 | 3 | 42918297 | T | C | 0.007032192 | YES |
| PGS000364_hmPOS_GRCh37 | 3 | 42920235 | T | C | 0.006582823 | YES |
| PGS000364_hmPOS_GRCh37 | 3 | 42921398 | A | G | 0.006827102 | YES |
| PGS000364_hmPOS_GRCh37 | 3 | 42921627 | A | C | 0.006404753 | YES |
| PGS000364_hmPOS_GRCh37 | 3 | 42921649 | C | T | 0.006813121 | YES |
| PGS000364_hmPOS_GRCh37 | 3 | 42922020 | A | G | 0.005970835 | YES |
| PGS000364_hmPOS_GRCh37 | 3 | 42922177 | G | C | 0.00530186 | NO |
| PGS000364_hmPOS_GRCh37 | 3 | 42922697 | T | G | 0.005498148 | YES |
| PGS000364_hmPOS_GRCh37 | 3 | 42923776 | T | G | 0.006290917 | YES |
| PGS000364_hmPOS_GRCh37 | 3 | 42923964 | C | T | 0.006274383 | YES |
| PGS000364_hmPOS_GRCh37 | 3 | 42924625 | C | A | 0.004698168 | YES |
| PGS000364_hmPOS_GRCh37 | 3 | 42924642 | G | A | 0.005033192 | YES |
| PGS000364_hmPOS_GRCh37 | 3 | 42924683 | G | C | 0.005067689 | NO |
| PGS000364_hmPOS_GRCh37 | 3 | 45672209 | T | G | 0.001015166 | YES |
| PGS000364_hmPOS_GRCh37 | 3 | 57204419 | G | A | -0.016635689 | YES |
| PGS000364_hmPOS_GRCh37 | 3 | 57270466 | C | T | -5.42028E-05 | YES |
| PGS000364_hmPOS_GRCh37 | 3 | 57314594 | G | A | -0.007944354 | YES |
| PGS000364_hmPOS_GRCh37 | 3 | 70857372 | A | G | 0.004362466 | YES |
| PGS000364_hmPOS_GRCh37 | 3 | 80800297 | C | T | 0.00158336 | YES |
| PGS000364_hmPOS_GRCh37 | 3 | 103845815 | T | G | 0.007229024 | YES |
| PGS000364_hmPOS_GRCh37 | 3 | 103854473 | A | G | 0.004555268 | YES |
| PGS000364_hmPOS_GRCh37 | 3 | 103886834 | T | C | 0.010290032 | YES |
| PGS000364_hmPOS_GRCh37 | 3 | 106010357 | T | G | 0.009994427 | YES |
| PGS000364_hmPOS_GRCh37 | 3 | 107502947 | C | T | -0.003469653 | YES |
| PGS000364_hmPOS_GRCh37 | 3 | 107503063 | A | G | -0.003489771 | YES |
| PGS000364_hmPOS_GRCh37 | 3 | 107504143 | T | C | -0.003773491 | YES |
| PGS000364_hmPOS_GRCh37 | 3 | 107507776 | C | T | -0.003439766 | YES |
| PGS000364_hmPOS_GRCh37 | 3 | 107515310 | T | G | -0.003044923 | YES |
| PGS000364_hmPOS_GRCh37 | 3 | 107515483 | T | G | -0.003044132 | YES |
| PGS000364_hmPOS_GRCh37 | 3 | 107520406 | G | A | 0.002536762 | YES |
| PGS000364_hmPOS_GRCh37 | 3 | 127438887 | C | T | 0.003927729 | YES |
| PGS000364_hmPOS_GRCh37 | 3 | 127440083 | C | A | 0.003945228 | YES |
| PGS000364_hmPOS_GRCh37 | 3 | 127444862 | T | C | 0.003431459 | YES |
| PGS000364_hmPOS_GRCh37 | 3 | 127445412 | T | C | 0.001401884 | YES |
| PGS000364_hmPOS_GRCh37 | 3 | 127446371 | C | T | 0.00276407 | YES |
| PGS000364_hmPOS_GRCh37 | 3 | 127453530 | A | G | 0.003638112 | YES |
| PGS000364_hmPOS_GRCh37 | 3 | 127465159 | A | G | 0.000253271 | YES |
| PGS000364_hmPOS_GRCh37 | 3 | 127465870 | A | G | 0.000729894 | YES |
| PGS000364_hmPOS_GRCh37 | 3 | 130362816 | A | G | 0.013129977 | YES |
| PGS000364_hmPOS_GRCh37 | 3 | 130363448 | T | C | 0.013229944 | YES |
| PGS000364_hmPOS_GRCh37 | 3 | 130368069 | G | A | 0.014681339 | YES |
| PGS000364_hmPOS_GRCh37 | 3 | 130392340 | G | C | 0.019325753 | NO |
| PGS000364_hmPOS_GRCh37 | 3 | 130446362 | T | C | 0.021706453 | YES |
| PGS000364_hmPOS_GRCh37 | 3 | 130478554 | A | G | 0.000404517 | YES |
| PGS000364_hmPOS_GRCh37 | 3 | 142016969 | T | C | 0.000949772 | YES |
| PGS000364_hmPOS_GRCh37 | 3 | 142879648 | T | C | 0.014539405 | YES |
| PGS000364_hmPOS_GRCh37 | 3 | 145063253 | C | T | 0.000309677 | YES |
| PGS000364_hmPOS_GRCh37 | 3 | 145871154 | C | T | 0.002298442 | YES |
| PGS000364_hmPOS_GRCh37 | 3 | 147410783 | T | C | 0.009823449 | YES |
| PGS000364_hmPOS_GRCh37 | 3 | 147412482 | T | C | 0.010059866 | YES |
| PGS000364_hmPOS_GRCh37 | 3 | 147412572 | A | G | 0.009949157 | YES |
| PGS000364_hmPOS_GRCh37 | 3 | 147553218 | C | T | 0.00431994 | YES |
| PGS000364_hmPOS_GRCh37 | 3 | 148894364 | T | A | -0.000140554 | NO |
| PGS000364_hmPOS_GRCh37 | 3 | 148915721 | G | T | -0.003710891 | YES |
| PGS000364_hmPOS_GRCh37 | 3 | 148921843 | A | G | -0.002586419 | YES |
| PGS000364_hmPOS_GRCh37 | 3 | 148942394 | C | T | -0.007663154 | YES |
| PGS000364_hmPOS_GRCh37 | 3 | 162343579 | T | C | 0.000821669 | NO |
| PGS000364_hmPOS_GRCh37 | 3 | 162375501 | A | T | 0.001034351 | NO |
| PGS000364_hmPOS_GRCh37 | 3 | 162377127 | C | T | 0.001034351 | YES |
| PGS000364_hmPOS_GRCh37 | 3 | 162377525 | T | C | 0.001034798 | YES |
| PGS000364_hmPOS_GRCh37 | 3 | 166673264 | T | C | 0.000138672 | YES |

| | | | | | | |
|---|---|---|---|---|---|---|
| PGS000364_hmPOS_GRCh37 | 3 | 175542154 | G | A | 0.005446553 | YES |
| PGS000364_hmPOS_GRCh37 | 3 | 186915592 | A | G | 0.00533753 | YES |
| PGS000364_hmPOS_GRCh37 | 3 | 186941603 | G | A | 0.009132343 | YES |
| PGS000364_hmPOS_GRCh37 | 3 | 189455911 | G | C | 0.002915686 | NO |
| PGS000364_hmPOS_GRCh37 | 3 | 195562342 | C | T | 0.009286117 | YES |
| PGS000364_hmPOS_GRCh37 | 3 | 196166769 | A | C | -0.00182654 | YES |
| PGS000364_hmPOS_GRCh37 | 3 | 196232298 | G | C | -0.0031544 | NO |
| PGS000364_hmPOS_GRCh37 | 3 | 196233795 | A | G | -0.000470715 | YES |
| PGS000364_hmPOS_GRCh37 | 3 | 196238028 | G | A | -0.000694913 | YES |
| PGS000364_hmPOS_GRCh37 | 3 | 196252167 | G | A | -1.35E-06 | YES |
| PGS000364_hmPOS_GRCh37 | 3 | 196253722 | G | A | -0.001336563 | YES |
| PGS000364_hmPOS_GRCh37 | 3 | 196254442 | A | G | -0.001246299 | YES |
| PGS000364_hmPOS_GRCh37 | 3 | 196254741 | A | G | -0.001122382 | YES |
| PGS000364_hmPOS_GRCh37 | 3 | 196254970 | C | A | -0.001218677 | YES |
| PGS000364_hmPOS_GRCh37 | 3 | 196255010 | T | C | -0.001266417 | YES |
| PGS000364_hmPOS_GRCh37 | 3 | 196255042 | G | C | -0.00124257 | NO |
| PGS000364_hmPOS_GRCh37 | 3 | 196255183 | G | A | -0.002552574 | YES |
| PGS000364_hmPOS_GRCh37 | 3 | 196255252 | A | C | -0.002472917 | YES |
| PGS000364_hmPOS_GRCh37 | 3 | 196255383 | A | G | -0.003787436 | YES |
| PGS000364_hmPOS_GRCh37 | 3 | 196255734 | C | G | -0.003431546 | NO |
| PGS000364_hmPOS_GRCh37 | 3 | 196255862 | G | C | -3.44019E-05 | NO |
| PGS000364_hmPOS_GRCh37 | 3 | 196255870 | C | G | -0.001323068 | NO |
| PGS000364_hmPOS_GRCh37 | 3 | 196255954 | C | G | -0.001949399 | NO |
| PGS000364_hmPOS_GRCh37 | 3 | 196256871 | C | G | 0.000796854 | NO |
| PGS000364_hmPOS_GRCh37 | 3 | 196257452 | G | A | -0.001049418 | YES |
| PGS000364_hmPOS_GRCh37 | 3 | 196259144 | T | C | -0.002917207 | YES |
| PGS000364_hmPOS_GRCh37 | 3 | 196260352 | A | G | -0.001921374 | YES |
| PGS000364_hmPOS_GRCh37 | 3 | 196260587 | T | G | -0.001632142 | YES |
| PGS000364_hmPOS_GRCh37 | 3 | 196261452 | G | A | -0.001214524 | YES |
| PGS000364_hmPOS_GRCh37 | 3 | 196261460 | C | A | -0.001212779 | YES |
| PGS000364_hmPOS_GRCh37 | 3 | 196261613 | C | T | -0.001209992 | YES |
| PGS000364_hmPOS_GRCh37 | 3 | 196261782 | A | G | -0.000917224 | YES |
| PGS000364_hmPOS_GRCh37 | 3 | 196261842 | A | G | -0.001212759 | YES |
| PGS000364_hmPOS_GRCh37 | 3 | 196262048 | C | T | -0.001196218 | YES |
| PGS000364_hmPOS_GRCh37 | 3 | 196262526 | G | A | -0.001688134 | YES |
| PGS000364_hmPOS_GRCh37 | 3 | 196262697 | T | C | -0.001196278 | YES |
| PGS000364_hmPOS_GRCh37 | 3 | 196262959 | A | G | -0.00236879 | YES |
| PGS000364_hmPOS_GRCh37 | 3 | 196264256 | A | G | -0.000876859 | YES |
| PGS000364_hmPOS_GRCh37 | 3 | 196264286 | G | A | -0.001663789 | YES |
| PGS000364_hmPOS_GRCh37 | 3 | 196264713 | C | T | -0.00034082 | YES |
| PGS000364_hmPOS_GRCh37 | 3 | 196264893 | G | T | -0.001227797 | YES |
| PGS000364_hmPOS_GRCh37 | 3 | 196265174 | G | T | -0.000997783 | YES |
| PGS000364_hmPOS_GRCh37 | 3 | 196265996 | T | C | -0.001382692 | YES |
| PGS000364_hmPOS_GRCh37 | 3 | 196266051 | A | G | -0.001721826 | YES |
| PGS000364_hmPOS_GRCh37 | 3 | 196266606 | A | G | -0.002267988 | YES |
| PGS000364_hmPOS_GRCh37 | 3 | 196269890 | C | T | -0.008475848 | YES |
| PGS000364_hmPOS_GRCh37 | 3 | 196271307 | C | A | -0.00876747 | YES |
| PGS000364_hmPOS_GRCh37 | 3 | 196272079 | A | G | -0.002205699 | YES |
| PGS000364_hmPOS_GRCh37 | 3 | 196274965 | C | A | -0.00930506 | YES |
| PGS000364_hmPOS_GRCh37 | 3 | 196276600 | G | C | -0.009109302 | NO |
| PGS000364_hmPOS_GRCh37 | 3 | 196278176 | C | A | -0.009585709 | YES |
| PGS000364_hmPOS_GRCh37 | 3 | 196278366 | T | C | -0.009532911 | YES |
| PGS000364_hmPOS_GRCh37 | 3 | 196280233 | G | A | -0.00279753 | YES |
| PGS000364_hmPOS_GRCh37 | 3 | 196280386 | G | A | -0.010658848 | YES |
| PGS000364_hmPOS_GRCh37 | 3 | 196280669 | G | A | -0.009360059 | YES |
| PGS000364_hmPOS_GRCh37 | 3 | 196281208 | C | T | -0.008873822 | YES |
| PGS000364_hmPOS_GRCh37 | 3 | 196283353 | C | G | -0.009239817 | NO |
| PGS000364_hmPOS_GRCh37 | 3 | 196283444 | A | G | -0.008714299 | YES |
| PGS000364_hmPOS_GRCh37 | 3 | 196283477 | C | A | -0.008683517 | YES |
| PGS000364_hmPOS_GRCh37 | 3 | 196284063 | A | C | -0.002050163 | YES |
| PGS000364_hmPOS_GRCh37 | 3 | 196286146 | C | G | -0.001728093 | NO |
| PGS000364_hmPOS_GRCh37 | 3 | 196286536 | A | G | -0.002022189 | YES |
| PGS000364_hmPOS_GRCh37 | 3 | 196288777 | A | G | -0.008048956 | YES |

| | | | | | | |
|---|---|---|---|---|---|---|
| PGS000364_hmPOS_GRCh37 | 3 | 196288847 | C | T | -0.008382025 | YES |
| PGS000364_hmPOS_GRCh37 | 3 | 196289014 | G | C | -0.001672355 | NO |
| PGS000364_hmPOS_GRCh37 | 3 | 196289387 | T | C | -0.001542614 | YES |
| PGS000364_hmPOS_GRCh37 | 3 | 196292385 | G | A | -0.012229717 | YES |
| PGS000364_hmPOS_GRCh37 | 3 | 196292987 | G | A | -0.008194284 | YES |
| PGS000364_hmPOS_GRCh37 | 3 | 196293516 | C | T | -0.007070052 | YES |
| PGS000364_hmPOS_GRCh37 | 3 | 196294755 | A | G | -0.003734556 | YES |
| PGS000364_hmPOS_GRCh37 | 3 | 196295163 | G | A | -0.003662726 | YES |
| PGS000364_hmPOS_GRCh37 | 3 | 196296320 | T | C | 0.002268036 | YES |
| PGS000364_hmPOS_GRCh37 | 4 | 1889648 | C | T | -0.00898492 | YES |
| PGS000364_hmPOS_GRCh37 | 4 | 2159978 | G | C | 0.004750519 | NO |
| PGS000364_hmPOS_GRCh37 | 4 | 2181864 | T | C | 0.006804361 | YES |
| PGS000364_hmPOS_GRCh37 | 4 | 2211314 | A | G | 0.00341886 | YES |
| PGS000364_hmPOS_GRCh37 | 4 | 3902018 | T | G | 0.003429886 | YES |
| PGS000364_hmPOS_GRCh37 | 4 | 3903945 | G | A | 0.005042835 | YES |
| PGS000364_hmPOS_GRCh37 | 4 | 3906926 | C | G | 0.004189622 | NO |
| PGS000364_hmPOS_GRCh37 | 4 | 3909117 | A | C | 0.001923338 | YES |
| PGS000364_hmPOS_GRCh37 | 4 | 3911370 | A | G | 0.001031314 | YES |
| PGS000364_hmPOS_GRCh37 | 4 | 3914204 | C | A | 0.000156387 | YES |
| PGS000364_hmPOS_GRCh37 | 4 | 16478955 | A | G | 0.003105067 | YES |
| PGS000364_hmPOS_GRCh37 | 4 | 17298632 | A | C | 0.000979216 | YES |
| PGS000364_hmPOS_GRCh37 | 4 | 17299793 | G | A | 0.000721458 | YES |
| PGS000364_hmPOS_GRCh37 | 4 | 17302823 | C | G | 0.000163096 | NO |
| PGS000364_hmPOS_GRCh37 | 4 | 17303802 | C | T | 0.001202 | YES |
| PGS000364_hmPOS_GRCh37 | 4 | 17305195 | C | T | 0.001744458 | YES |
| PGS000364_hmPOS_GRCh37 | 4 | 17305208 | C | T | 0.001142822 | YES |
| PGS000364_hmPOS_GRCh37 | 4 | 17306702 | G | T | 0.000860588 | YES |
| PGS000364_hmPOS_GRCh37 | 4 | 24458145 | C | T | 0.00613632 | YES |
| PGS000364_hmPOS_GRCh37 | 4 | 24462075 | A | G | 0.005220977 | YES |
| PGS000364_hmPOS_GRCh37 | 4 | 24462248 | C | G | 0.006687342 | NO |
| PGS000364_hmPOS_GRCh37 | 4 | 31703249 | T | C | 0.00437057 | YES |
| PGS000364_hmPOS_GRCh37 | 4 | 40338079 | A | G | 0.024768585 | YES |
| PGS000364_hmPOS_GRCh37 | 4 | 59396970 | T | C | 0.004314379 | YES |
| PGS000364_hmPOS_GRCh37 | 4 | 73854746 | A | G | 0.005597586 | YES |
| PGS000364_hmPOS_GRCh37 | 4 | 82474592 | T | C | 0.007569317 | YES |
| PGS000364_hmPOS_GRCh37 | 4 | 91311465 | A | G | 0.009393752 | YES |
| PGS000364_hmPOS_GRCh37 | 4 | 95019067 | A | G | 7.8591E-05 | YES |
| PGS000364_hmPOS_GRCh37 | 4 | 101460676 | A | T | -0.007154023 | NO |
| PGS000364_hmPOS_GRCh37 | 4 | 101463177 | G | A | -0.004713072 | YES |
| PGS000364_hmPOS_GRCh37 | 4 | 101507954 | G | A | -0.000291752 | YES |
| PGS000364_hmPOS_GRCh37 | 4 | 101507968 | A | G | -0.000220816 | YES |
| PGS000364_hmPOS_GRCh37 | 4 | 101508001 | A | G | -0.001020602 | YES |
| PGS000364_hmPOS_GRCh37 | 4 | 101508721 | A | C | -0.002796013 | YES |
| PGS000364_hmPOS_GRCh37 | 4 | 101508727 | A | G | -0.001577306 | YES |
| PGS000364_hmPOS_GRCh37 | 4 | 101510000 | G | A | -0.002603427 | YES |
| PGS000364_hmPOS_GRCh37 | 4 | 109659741 | A | G | 0.002502775 | YES |
| PGS000364_hmPOS_GRCh37 | 4 | 109685983 | A | G | 0.005131332 | YES |
| PGS000364_hmPOS_GRCh37 | 4 | 109690334 | T | C | 0.004197156 | YES |
| PGS000364_hmPOS_GRCh37 | 4 | 109694619 | A | C | 0.002535435 | YES |
| PGS000364_hmPOS_GRCh37 | 4 | 115381722 | T | C | 0.003846519 | YES |
| PGS000364_hmPOS_GRCh37 | 4 | 122157569 | C | T | 0.000276715 | YES |
| PGS000364_hmPOS_GRCh37 | 4 | 124302098 | A | G | 0.001420308 | YES |
| PGS000364_hmPOS_GRCh37 | 4 | 128656054 | A | G | 0.002232434 | YES |
| PGS000364_hmPOS_GRCh37 | 4 | 128959213 | A | G | 0.010032209 | YES |
| PGS000364_hmPOS_GRCh37 | 4 | 128967824 | C | T | 0.013536533 | YES |
| PGS000364_hmPOS_GRCh37 | 4 | 128993780 | T | C | 0.010977221 | YES |
| PGS000364_hmPOS_GRCh37 | 4 | 129022604 | G | A | 0.010189766 | YES |
| PGS000364_hmPOS_GRCh37 | 4 | 129024273 | T | C | 0.012786266 | YES |
| PGS000364_hmPOS_GRCh37 | 4 | 129034787 | T | C | 0.012139802 | YES |
| PGS000364_hmPOS_GRCh37 | 4 | 129036153 | T | C | 0.012120484 | YES |
| PGS000364_hmPOS_GRCh37 | 4 | 129047537 | A | G | 0.012261125 | YES |
| PGS000364_hmPOS_GRCh37 | 4 | 129047722 | C | A | 0.012270507 | YES |
| PGS000364_hmPOS_GRCh37 | 4 | 129055474 | G | A | 0.012206155 | YES |

| | | | | | | |
|---|---|---|---|---|---|---|
| PGS000364_hmPOS_GRCh37 | 4 | 129084813 | C | T | 0.010750275 | YES |
| PGS000364_hmPOS_GRCh37 | 4 | 129115454 | C | T | 0.009507701 | YES |
| PGS000364_hmPOS_GRCh37 | 4 | 129125483 | G | A | 0.010015315 | YES |
| PGS000364_hmPOS_GRCh37 | 4 | 129132929 | T | C | 0.008713789 | YES |
| PGS000364_hmPOS_GRCh37 | 4 | 142811893 | T | C | -0.003679446 | YES |
| PGS000364_hmPOS_GRCh37 | 4 | 142811946 | A | C | -0.003702495 | YES |
| PGS000364_hmPOS_GRCh37 | 4 | 142812073 | T | C | -0.003805697 | YES |
| PGS000364_hmPOS_GRCh37 | 4 | 142814856 | G | A | -0.00199894 | YES |
| PGS000364_hmPOS_GRCh37 | 4 | 142815639 | A | G | -0.001726673 | YES |
| PGS000364_hmPOS_GRCh37 | 4 | 142816579 | G | C | -0.003478808 | NO |
| PGS000364_hmPOS_GRCh37 | 4 | 142836961 | G | A | -0.001158024 | YES |
| PGS000364_hmPOS_GRCh37 | 4 | 143243556 | T | C | -0.00226488 | YES |
| PGS000364_hmPOS_GRCh37 | 4 | 143244840 | C | T | -0.01755302 | YES |
| PGS000364_hmPOS_GRCh37 | 4 | 143255255 | A | G | -7.19572E-05 | YES |
| PGS000364_hmPOS_GRCh37 | 4 | 143257940 | T | C | -0.001868063 | YES |
| PGS000364_hmPOS_GRCh37 | 4 | 143264762 | C | G | -5.0509E-05 | NO |
| PGS000364_hmPOS_GRCh37 | 4 | 143265405 | A | G | -0.000306844 | YES |
| PGS000364_hmPOS_GRCh37 | 4 | 143265572 | A | G | -0.000299957 | YES |
| PGS000364_hmPOS_GRCh37 | 4 | 143265791 | T | C | -0.000330721 | YES |
| PGS000364_hmPOS_GRCh37 | 4 | 143268348 | A | G | -0.002917607 | YES |
| PGS000364_hmPOS_GRCh37 | 4 | 143268528 | A | T | -0.002863752 | NO |
| PGS000364_hmPOS_GRCh37 | 4 | 143269745 | A | T | -0.002740453 | NO |
| PGS000364_hmPOS_GRCh37 | 4 | 143277523 | A | G | -0.003316033 | YES |
| PGS000364_hmPOS_GRCh37 | 4 | 143786032 | A | G | 0.013655211 | YES |
| PGS000364_hmPOS_GRCh37 | 4 | 146562236 | T | C | 0.002131489 | YES |
| PGS000364_hmPOS_GRCh37 | 4 | 146562933 | T | C | 0.001302395 | YES |
| PGS000364_hmPOS_GRCh37 | 4 | 153662207 | A | G | 0.005063134 | YES |
| PGS000364_hmPOS_GRCh37 | 4 | 177958179 | T | C | 0.001516595 | YES |
| PGS000364_hmPOS_GRCh37 | 4 | 181580491 | T | C | 0.005199363 | YES |
| PGS000364_hmPOS_GRCh37 | 4 | 187406118 | C | G | 0.001315374 | NO |
| PGS000364_hmPOS_GRCh37 | 4 | 189991096 | C | G | 1.1176E-05 | NO |
| PGS000364_hmPOS_GRCh37 | 5 | 1124703 | T | C | 0.00845713 | YES |
| PGS000364_hmPOS_GRCh37 | 5 | 1125033 | T | C | 0.008620539 | YES |
| PGS000364_hmPOS_GRCh37 | 5 | 3573929 | T | A | 0.006259944 | NO |
| PGS000364_hmPOS_GRCh37 | 5 | 13910597 | C | G | -0.010136154 | NO |
| PGS000364_hmPOS_GRCh37 | 5 | 25346330 | C | G | 0.001075647 | NO |
| PGS000364_hmPOS_GRCh37 | 5 | 37680949 | A | G | -0.001197578 | YES |
| PGS000364_hmPOS_GRCh37 | 5 | 53214068 | T | C | 0.010299046 | YES |
| PGS000364_hmPOS_GRCh37 | 5 | 66431562 | G | C | 3.84495E-05 | NO |
| PGS000364_hmPOS_GRCh37 | 5 | 66807910 | A | C | -0.000357461 | YES |
| PGS000364_hmPOS_GRCh37 | 5 | 66814587 | A | G | -0.004433778 | YES |
| PGS000364_hmPOS_GRCh37 | 5 | 66814842 | C | T | -0.000152076 | YES |
| PGS000364_hmPOS_GRCh37 | 5 | 72515051 | C | T | 0.00034511 | YES |
| PGS000364_hmPOS_GRCh37 | 5 | 72518148 | C | T | 0.002864916 | YES |
| PGS000364_hmPOS_GRCh37 | 5 | 72518452 | G | A | 0.001411493 | YES |
| PGS000364_hmPOS_GRCh37 | 5 | 72519678 | G | A | 0.001440871 | YES |
| PGS000364_hmPOS_GRCh37 | 5 | 72520347 | T | A | 0.002512711 | NO |
| PGS000364_hmPOS_GRCh37 | 5 | 72520535 | T | C | 0.001889864 | YES |
| PGS000364_hmPOS_GRCh37 | 5 | 72521202 | G | A | 0.001527699 | YES |
| PGS000364_hmPOS_GRCh37 | 5 | 72521622 | G | C | 0.001514085 | NO |
| PGS000364_hmPOS_GRCh37 | 5 | 72521854 | T | C | 0.001594952 | YES |
| PGS000364_hmPOS_GRCh37 | 5 | 72522437 | T | C | 0.001488566 | YES |
| PGS000364_hmPOS_GRCh37 | 5 | 72523428 | C | T | 0.000876613 | YES |
| PGS000364_hmPOS_GRCh37 | 5 | 72524578 | C | T | 0.001333043 | YES |
| PGS000364_hmPOS_GRCh37 | 5 | 72525986 | A | G | 0.001569237 | YES |
| PGS000364_hmPOS_GRCh37 | 5 | 72527849 | C | T | 0.000241488 | YES |
| PGS000364_hmPOS_GRCh37 | 5 | 72528922 | C | A | 0.000337712 | YES |
| PGS000364_hmPOS_GRCh37 | 5 | 72529325 | T | C | 0.000415951 | YES |
| PGS000364_hmPOS_GRCh37 | 5 | 72531994 | C | A | 0.000986358 | YES |
| PGS000364_hmPOS_GRCh37 | 5 | 72539850 | A | C | 0.00543699 | YES |
| PGS000364_hmPOS_GRCh37 | 5 | 72541598 | C | A | 0.004241398 | YES |
| PGS000364_hmPOS_GRCh37 | 5 | 72541687 | A | G | 0.005683243 | YES |
| PGS000364_hmPOS_GRCh37 | 5 | 72542050 | A | C | 0.004333185 | YES |

| | | | | | | |
|---|---|---|---|---|---|---|
| PGS000364_hmPOS_GRCh37 | 5 | 72543056 | G | A | 0.004706068 | YES |
| PGS000364_hmPOS_GRCh37 | 5 | 72543213 | T | G | 0.004783909 | YES |
| PGS000364_hmPOS_GRCh37 | 5 | 72543998 | T | C | 0.004526791 | YES |
| PGS000364_hmPOS_GRCh37 | 5 | 72544390 | C | T | 0.004164896 | YES |
| PGS000364_hmPOS_GRCh37 | 5 | 72545909 | C | T | 0.004303747 | YES |
| PGS000364_hmPOS_GRCh37 | 5 | 72550088 | G | T | 0.00464898 | YES |
| PGS000364_hmPOS_GRCh37 | 5 | 72551134 | G | A | 0.004679727 | YES |
| PGS000364_hmPOS_GRCh37 | 5 | 72559339 | G | T | 0.012734895 | YES |
| PGS000364_hmPOS_GRCh37 | 5 | 89867373 | G | A | 0.004688253 | YES |
| PGS000364_hmPOS_GRCh37 | 5 | 89889650 | C | T | 0.009919211 | YES |
| PGS000364_hmPOS_GRCh37 | 5 | 89977598 | A | G | 3.99701E-05 | YES |
| PGS000364_hmPOS_GRCh37 | 5 | 105896673 | T | A | -0.002939266 | NO |
| PGS000364_hmPOS_GRCh37 | 5 | 105899335 | A | G | -0.001265405 | YES |
| PGS000364_hmPOS_GRCh37 | 5 | 105902493 | G | A | -0.002742473 | YES |
| PGS000364_hmPOS_GRCh37 | 5 | 105904156 | C | T | -0.003384866 | YES |
| PGS000364_hmPOS_GRCh37 | 5 | 105905893 | A | G | -0.003036638 | YES |
| PGS000364_hmPOS_GRCh37 | 5 | 105909464 | A | G | -0.002261677 | YES |
| PGS000364_hmPOS_GRCh37 | 5 | 105910403 | G | C | -0.003355608 | NO |
| PGS000364_hmPOS_GRCh37 | 5 | 105911088 | T | A | -0.003390127 | NO |
| PGS000364_hmPOS_GRCh37 | 5 | 105916636 | T | A | -0.002350719 | NO |
| PGS000364_hmPOS_GRCh37 | 5 | 105920735 | T | G | -0.004108002 | YES |
| PGS000364_hmPOS_GRCh37 | 5 | 105948256 | G | C | 0.001858118 | NO |
| PGS000364_hmPOS_GRCh37 | 5 | 106005120 | G | A | -0.000701322 | YES |
| PGS000364_hmPOS_GRCh37 | 5 | 106006952 | C | G | -0.000897075 | NO |
| PGS000364_hmPOS_GRCh37 | 5 | 106009952 | G | T | -0.001244969 | YES |
| PGS000364_hmPOS_GRCh37 | 5 | 106010168 | C | T | -0.001285526 | YES |
| PGS000364_hmPOS_GRCh37 | 5 | 106016806 | C | G | -0.000380611 | NO |
| PGS000364_hmPOS_GRCh37 | 5 | 117692172 | C | T | 0.000959667 | YES |
| PGS000364_hmPOS_GRCh37 | 5 | 117697578 | G | A | 0.000111617 | YES |
| PGS000364_hmPOS_GRCh37 | 5 | 117701098 | G | C | 0.010263914 | NO |
| PGS000364_hmPOS_GRCh37 | 5 | 117701468 | A | G | 0.002074076 | YES |
| PGS000364_hmPOS_GRCh37 | 5 | 117709592 | A | G | 0.000786231 | YES |
| PGS000364_hmPOS_GRCh37 | 5 | 119675960 | T | G | -0.00080712 | YES |
| PGS000364_hmPOS_GRCh37 | 5 | 119781866 | G | T | -0.000524633 | YES |
| PGS000364_hmPOS_GRCh37 | 5 | 119789295 | G | A | -0.001473303 | YES |
| PGS000364_hmPOS_GRCh37 | 5 | 119791574 | T | G | -0.001107663 | YES |
| PGS000364_hmPOS_GRCh37 | 5 | 119801534 | G | C | -0.004371563 | NO |
| PGS000364_hmPOS_GRCh37 | 5 | 119857153 | T | C | -0.003012964 | YES |
| PGS000364_hmPOS_GRCh37 | 5 | 119862127 | G | A | -0.000642625 | YES |
| PGS000364_hmPOS_GRCh37 | 5 | 119864882 | T | A | -0.000429768 | NO |
| PGS000364_hmPOS_GRCh37 | 5 | 121756884 | T | C | 0.002626605 | YES |
| PGS000364_hmPOS_GRCh37 | 5 | 124679571 | G | A | 0.001502522 | YES |
| PGS000364_hmPOS_GRCh37 | 5 | 124725647 | T | C | 0.01136001 | YES |
| PGS000364_hmPOS_GRCh37 | 5 | 124854331 | C | T | 0.006906936 | YES |
| PGS000364_hmPOS_GRCh37 | 5 | 124900867 | A | C | 0.000226131 | YES |
| PGS000364_hmPOS_GRCh37 | 5 | 124914586 | A | C | 0.011592178 | YES |
| PGS000364_hmPOS_GRCh37 | 5 | 124941524 | C | T | 0.012935174 | YES |
| PGS000364_hmPOS_GRCh37 | 5 | 124943054 | T | G | 0.006154137 | YES |
| PGS000364_hmPOS_GRCh37 | 5 | 124960510 | T | C | 0.014505994 | YES |
| PGS000364_hmPOS_GRCh37 | 5 | 125055586 | T | C | 0.005598772 | YES |
| PGS000364_hmPOS_GRCh37 | 5 | 125121440 | C | T | 0.002100993 | YES |
| PGS000364_hmPOS_GRCh37 | 5 | 148223138 | C | A | 0.008104573 | YES |
| PGS000364_hmPOS_GRCh37 | 5 | 152180918 | T | C | 0.004665845 | YES |
| PGS000364_hmPOS_GRCh37 | 5 | 152207669 | C | A | 0.001074883 | YES |
| PGS000364_hmPOS_GRCh37 | 5 | 152210177 | A | G | 0.000780161 | YES |
| PGS000364_hmPOS_GRCh37 | 5 | 162387260 | G | A | 0.006634154 | YES |
| PGS000364_hmPOS_GRCh37 | 5 | 163550616 | T | A | 0.000680683 | NO |
| PGS000364_hmPOS_GRCh37 | 5 | 163550678 | C | G | 0.00029575 | NO |
| PGS000364_hmPOS_GRCh37 | 5 | 171635750 | C | T | 0.001095064 | YES |
| PGS000364_hmPOS_GRCh37 | 5 | 171636059 | C | T | 0.002036413 | YES |
| PGS000364_hmPOS_GRCh37 | 5 | 171637347 | C | G | 0.002071808 | NO |
| PGS000364_hmPOS_GRCh37 | 5 | 171664504 | C | T | 0.006006998 | YES |
| PGS000364_hmPOS_GRCh37 | 5 | 171672429 | C | T | 0.005069581 | YES |

| | | | | | | |
|---|---|---|---|---|---|---|
| PGS000364_hmPOS_GRCh37 | 5 | 171683884 | C | T | 0.006160259 | YES |
| PGS000364_hmPOS_GRCh37 | 5 | 171685829 | G | A | 0.006163627 | YES |
| PGS000364_hmPOS_GRCh37 | 5 | 171709497 | C | G | 0.007481378 | NO |
| PGS000364_hmPOS_GRCh37 | 5 | 172872410 | G | A | 0.006708635 | YES |
| PGS000364_hmPOS_GRCh37 | 5 | 172872881 | G | A | 0.007329792 | YES |
| PGS000364_hmPOS_GRCh37 | 5 | 172880042 | T | C | 0.007248177 | YES |
| PGS000364_hmPOS_GRCh37 | 5 | 172881712 | C | T | 0.007038286 | YES |
| PGS000364_hmPOS_GRCh37 | 5 | 172884244 | G | C | 0.007056802 | NO |
| PGS000364_hmPOS_GRCh37 | 5 | 172886986 | C | G | 0.00580999 | NO |
| PGS000364_hmPOS_GRCh37 | 5 | 173206446 | G | A | 0.000734302 | YES |
| PGS000364_hmPOS_GRCh37 | 5 | 177926531 | C | A | 0.000591055 | YES |
| PGS000364_hmPOS_GRCh37 | 5 | 177928404 | C | T | 0.000369069 | YES |
| PGS000364_hmPOS_GRCh37 | 6 | 1671997 | T | C | 0.004777113 | YES |
| PGS000364_hmPOS_GRCh37 | 6 | 1675928 | C | T | 0.004357649 | YES |
| PGS000364_hmPOS_GRCh37 | 6 | 1682704 | C | T | 0.003588834 | YES |
| PGS000364_hmPOS_GRCh37 | 6 | 1683287 | T | C | 0.003497896 | YES |
| PGS000364_hmPOS_GRCh37 | 6 | 3441938 | A | T | 0.001093197 | NO |
| PGS000364_hmPOS_GRCh37 | 6 | 3742857 | G | A | 0.003159626 | YES |
| PGS000364_hmPOS_GRCh37 | 6 | 19569186 | A | G | 0.014651283 | YES |
| PGS000364_hmPOS_GRCh37 | 6 | 19601623 | T | C | 0.014244222 | YES |
| PGS000364_hmPOS_GRCh37 | 6 | 19611195 | A | T | 0.013543506 | NO |
| PGS000364_hmPOS_GRCh37 | 6 | 24449382 | C | G | 0.001722808 | NO |
| PGS000364_hmPOS_GRCh37 | 6 | 24449405 | C | T | 0.001631239 | YES |
| PGS000364_hmPOS_GRCh37 | 6 | 41656820 | T | C | 0.000976516 | YES |
| PGS000364_hmPOS_GRCh37 | 6 | 43655832 | G | C | 0.001130269 | NO |
| PGS000364_hmPOS_GRCh37 | 6 | 49771994 | A | C | -0.002299764 | YES |
| PGS000364_hmPOS_GRCh37 | 6 | 51239904 | C | G | 0.010095234 | NO |
| PGS000364_hmPOS_GRCh37 | 6 | 51279138 | T | G | 0.00693884 | YES |
| PGS000364_hmPOS_GRCh37 | 6 | 51297423 | A | G | 0.001176633 | YES |
| PGS000364_hmPOS_GRCh37 | 6 | 57125194 | G | A | 0.005962068 | YES |
| PGS000364_hmPOS_GRCh37 | 6 | 87805210 | T | A | 0.000274121 | NO |
| PGS000364_hmPOS_GRCh37 | 6 | 87805592 | T | C | 0.000255548 | YES |
| PGS000364_hmPOS_GRCh37 | 6 | 87805890 | G | T | 0.000243009 | YES |
| PGS000364_hmPOS_GRCh37 | 6 | 87807180 | G | A | 0.001197318 | YES |
| PGS000364_hmPOS_GRCh37 | 6 | 87808430 | T | A | 0.000229728 | NO |
| PGS000364_hmPOS_GRCh37 | 6 | 88830565 | G | T | -1.36279E-05 | YES |
| PGS000364_hmPOS_GRCh37 | 6 | 97461972 | A | G | 0.00272315 | YES |
| PGS000364_hmPOS_GRCh37 | 6 | 105003599 | A | G | 0.01472284 | YES |
| PGS000364_hmPOS_GRCh37 | 6 | 105207689 | G | A | 0.01412758 | YES |
| PGS000364_hmPOS_GRCh37 | 6 | 105336393 | A | G | 0.012148552 | YES |
| PGS000364_hmPOS_GRCh37 | 6 | 105349842 | A | C | 0.009773943 | YES |
| PGS000364_hmPOS_GRCh37 | 6 | 105446339 | T | A | 0.005208141 | NO |
| PGS000364_hmPOS_GRCh37 | 6 | 105462351 | T | C | 0.00656456 | YES |
| PGS000364_hmPOS_GRCh37 | 6 | 105625409 | T | C | 0.001080471 | YES |
| PGS000364_hmPOS_GRCh37 | 6 | 112306397 | G | T | 0.001671749 | YES |
| PGS000364_hmPOS_GRCh37 | 6 | 112306744 | A | T | 0.002504791 | NO |
| PGS000364_hmPOS_GRCh37 | 6 | 112308562 | G | A | 0.00406012 | NO |
| PGS000364_hmPOS_GRCh37 | 6 | 112321699 | A | G | 0.003179404 | YES |
| PGS000364_hmPOS_GRCh37 | 6 | 112326344 | T | C | 0.003143047 | YES |
| PGS000364_hmPOS_GRCh37 | 6 | 112332250 | A | T | 0.00318415 | NO |
| PGS000364_hmPOS_GRCh37 | 6 | 112332529 | G | C | 0.003225345 | NO |
| PGS000364_hmPOS_GRCh37 | 6 | 123729699 | C | T | -2.7024E-05 | YES |
| PGS000364_hmPOS_GRCh37 | 6 | 125612350 | A | G | 0.000815339 | YES |
| PGS000364_hmPOS_GRCh37 | 6 | 136387111 | G | A | 0.002484738 | YES |
| PGS000364_hmPOS_GRCh37 | 6 | 136594187 | T | C | 0.007356564 | YES |
| PGS000364_hmPOS_GRCh37 | 6 | 138879771 | T | C | 0.020633992 | YES |
| PGS000364_hmPOS_GRCh37 | 6 | 144089607 | A | G | 2.84263E-05 | YES |
| PGS000364_hmPOS_GRCh37 | 6 | 158036144 | A | G | -0.008162692 | YES |
| PGS000364_hmPOS_GRCh37 | 6 | 158036684 | C | T | -0.007781472 | YES |
| PGS000364_hmPOS_GRCh37 | 6 | 158036933 | G | A | -0.006396654 | YES |
| PGS000364_hmPOS_GRCh37 | 6 | 158038751 | A | G | -0.005718743 | YES |
| PGS000364_hmPOS_GRCh37 | 6 | 158038975 | G | A | -0.005740375 | YES |
| PGS000364_hmPOS_GRCh37 | 6 | 158040365 | G | A | -0.00706209 | YES |

| | | | | | | |
|---|---|---|---|---|---|---|
| PGS000364_hmPOS_GRCh37 | 6 | 158040465 | G | A | -0.006108448 | YES |
| PGS000364_hmPOS_GRCh37 | 6 | 158040969 | G | C | -0.006909926 | NO |
| PGS000364_hmPOS_GRCh37 | 6 | 158041022 | T | G | -0.006571396 | YES |
| PGS000364_hmPOS_GRCh37 | 6 | 158041511 | T | G | -0.004031708 | YES |
| PGS000364_hmPOS_GRCh37 | 6 | 158042765 | A | G | -0.003792019 | YES |
| PGS000364_hmPOS_GRCh37 | 6 | 158043637 | G | C | -0.006299873 | NO |
| PGS000364_hmPOS_GRCh37 | 6 | 158043779 | C | T | -0.005920537 | YES |
| PGS000364_hmPOS_GRCh37 | 6 | 158053181 | A | G | -0.002566718 | YES |
| PGS000364_hmPOS_GRCh37 | 6 | 158053235 | T | C | -0.000938303 | YES |
| PGS000364_hmPOS_GRCh37 | 6 | 158132487 | T | A | 0.000938916 | NO |
| PGS000364_hmPOS_GRCh37 | 6 | 160808901 | T | G | 0.003409741 | YES |
| PGS000364_hmPOS_GRCh37 | 6 | 160822675 | G | A | 0.003341966 | YES |
| PGS000364_hmPOS_GRCh37 | 6 | 160833664 | T | C | 0.002312257 | YES |
| PGS000364_hmPOS_GRCh37 | 6 | 160834515 | C | T | 0.00203945 | YES |
| PGS000364_hmPOS_GRCh37 | 6 | 160835192 | A | C | 0.002398152 | YES |
| PGS000364_hmPOS_GRCh37 | 6 | 161502445 | T | C | -0.000722235 | YES |
| PGS000364_hmPOS_GRCh37 | 6 | 162935587 | T | C | 0.018734717 | YES |
| PGS000364_hmPOS_GRCh37 | 6 | 162952921 | A | G | 0.019262416 | YES |
| PGS000364_hmPOS_GRCh37 | 6 | 163402433 | T | C | -0.01048161 | YES |
| PGS000364_hmPOS_GRCh37 | 6 | 165969237 | C | A | 0.002499889 | YES |
| PGS000364_hmPOS_GRCh37 | 6 | 166196371 | C | T | 0.006900854 | YES |
| PGS000364_hmPOS_GRCh37 | 6 | 167576698 | C | A | 0.000518641 | YES |
| PGS000364_hmPOS_GRCh37 | 6 | 167647068 | A | G | -0.0010594 | YES |
| PGS000364_hmPOS_GRCh37 | 7 | 809825 | A | G | 0.006721119 | YES |
| PGS000364_hmPOS_GRCh37 | 7 | 4250040 | T | C | 0.012264536 | YES |
| PGS000364_hmPOS_GRCh37 | 7 | 5623731 | T | C | 0.000797078 | YES |
| PGS000364_hmPOS_GRCh37 | 7 | 5642631 | A | G | 4.64204E-05 | YES |
| PGS000364_hmPOS_GRCh37 | 7 | 5644243 | A | G | 0.001146133 | YES |
| PGS000364_hmPOS_GRCh37 | 7 | 14931662 | T | G | -0.00094555 | YES |
| PGS000364_hmPOS_GRCh37 | 7 | 15481653 | C | G | -0.002049333 | NO |
| PGS000364_hmPOS_GRCh37 | 7 | 15482665 | G | C | -0.002656024 | NO |
| PGS000364_hmPOS_GRCh37 | 7 | 15484961 | T | A | -0.00267139 | NO |
| PGS000364_hmPOS_GRCh37 | 7 | 15485123 | T | C | -0.002177859 | YES |
| PGS000364_hmPOS_GRCh37 | 7 | 15493547 | C | T | -0.00349395 | YES |
| PGS000364_hmPOS_GRCh37 | 7 | 17887004 | C | G | 0.001157389 | NO |
| PGS000364_hmPOS_GRCh37 | 7 | 19254640 | C | T | -0.002090328 | YES |
| PGS000364_hmPOS_GRCh37 | 7 | 19254715 | A | G | -0.002561941 | YES |
| PGS000364_hmPOS_GRCh37 | 7 | 19254726 | A | G | -0.005775797 | YES |
| PGS000364_hmPOS_GRCh37 | 7 | 19257738 | C | G | -0.001880309 | NO |
| PGS000364_hmPOS_GRCh37 | 7 | 19258451 | G | A | -0.001616766 | YES |
| PGS000364_hmPOS_GRCh37 | 7 | 19260180 | A | T | -0.001643074 | NO |
| PGS000364_hmPOS_GRCh37 | 7 | 19261413 | C | A | -0.001525692 | YES |
| PGS000364_hmPOS_GRCh37 | 7 | 19261523 | A | T | -0.000478721 | NO |
| PGS000364_hmPOS_GRCh37 | 7 | 19261584 | A | G | -0.00160019 | YES |
| PGS000364_hmPOS_GRCh37 | 7 | 19262091 | T | C | -0.000751519 | YES |
| PGS000364_hmPOS_GRCh37 | 7 | 19262783 | C | A | -0.006330536 | YES |
| PGS000364_hmPOS_GRCh37 | 7 | 19264504 | T | C | -0.005995766 | YES |
| PGS000364_hmPOS_GRCh37 | 7 | 19264888 | A | G | -0.002329003 | YES |
| PGS000364_hmPOS_GRCh37 | 7 | 19265301 | G | T | -0.002626351 | YES |
| PGS000364_hmPOS_GRCh37 | 7 | 19267296 | A | T | -0.003019369 | NO |
| PGS000364_hmPOS_GRCh37 | 7 | 19267342 | G | C | -0.003302683 | NO |
| PGS000364_hmPOS_GRCh37 | 7 | 19269017 | G | A | -0.001656725 | YES |
| PGS000364_hmPOS_GRCh37 | 7 | 19270317 | G | T | -0.002350864 | YES |
| PGS000364_hmPOS_GRCh37 | 7 | 19271417 | C | T | -0.001730628 | YES |
| PGS000364_hmPOS_GRCh37 | 7 | 19271726 | C | A | -0.005410896 | YES |
| PGS000364_hmPOS_GRCh37 | 7 | 19274183 | C | T | -0.007309493 | YES |
| PGS000364_hmPOS_GRCh37 | 7 | 19274665 | C | T | -0.001533365 | YES |
| PGS000364_hmPOS_GRCh37 | 7 | 19463507 | C | T | -0.001963768 | YES |
| PGS000364_hmPOS_GRCh37 | 7 | 22497005 | G | A | 0.000584002 | YES |
| PGS000364_hmPOS_GRCh37 | 7 | 28408943 | C | T | 0.004215619 | YES |
| PGS000364_hmPOS_GRCh37 | 7 | 31006942 | G | A | -0.001542487 | YES |
| PGS000364_hmPOS_GRCh37 | 7 | 31136610 | A | C | 0.001551539 | YES |
| PGS000364_hmPOS_GRCh37 | 7 | 43473208 | G | C | 0.005347253 | NO |

| | | | | | | |
|---|---|---|---|---|---|---|
| PGS000364_hmPOS_GRCh37 | 7 | 43474087 | G | A | 0.008057458 | YES |
| PGS000364_hmPOS_GRCh37 | 7 | 43474488 | A | G | 0.008056349 | YES |
| PGS000364_hmPOS_GRCh37 | 7 | 44792994 | T | C | 0.003309297 | YES |
| PGS000364_hmPOS_GRCh37 | 7 | 69238281 | A | G | -0.005315069 | YES |
| PGS000364_hmPOS_GRCh37 | 7 | 69252380 | A | C | -0.000141125 | YES |
| PGS000364_hmPOS_GRCh37 | 7 | 69257350 | T | C | -0.000281855 | YES |
| PGS000364_hmPOS_GRCh37 | 7 | 71127751 | G | A | -0.000524383 | YES |
| PGS000364_hmPOS_GRCh37 | 7 | 71134745 | A | G | -0.004201309 | YES |
| PGS000364_hmPOS_GRCh37 | 7 | 71153323 | G | C | -0.004262521 | NO |
| PGS000364_hmPOS_GRCh37 | 7 | 71153438 | T | C | -0.003132214 | YES |
| PGS000364_hmPOS_GRCh37 | 7 | 71155695 | A | G | -0.002345652 | YES |
| PGS000364_hmPOS_GRCh37 | 7 | 71157097 | C | T | -0.004095097 | YES |
| PGS000364_hmPOS_GRCh37 | 7 | 71160137 | G | A | -0.001420993 | YES |
| PGS000364_hmPOS_GRCh37 | 7 | 71178550 | A | C | -0.001949966 | YES |
| PGS000364_hmPOS_GRCh37 | 7 | 71181184 | A | G | -0.001921893 | YES |
| PGS000364_hmPOS_GRCh37 | 7 | 85506052 | A | T | 0.00126407 | NO |
| PGS000364_hmPOS_GRCh37 | 7 | 85589868 | C | T | 0.009531718 | YES |
| PGS000364_hmPOS_GRCh37 | 7 | 87836113 | A | C | 0.004952083 | YES |
| PGS000364_hmPOS_GRCh37 | 7 | 87844303 | G | A | 0.004751543 | YES |
| PGS000364_hmPOS_GRCh37 | 7 | 87844535 | C | T | 0.004773846 | YES |
| PGS000364_hmPOS_GRCh37 | 7 | 87845378 | T | C | 0.004805859 | YES |
| PGS000364_hmPOS_GRCh37 | 7 | 87978996 | A | G | 0.001834007 | YES |
| PGS000364_hmPOS_GRCh37 | 7 | 87980922 | A | G | 0.002066811 | YES |
| PGS000364_hmPOS_GRCh37 | 7 | 90741500 | G | A | 0.001233388 | YES |
| PGS000364_hmPOS_GRCh37 | 7 | 90746088 | C | A | 0.000269073 | YES |
| PGS000364_hmPOS_GRCh37 | 7 | 90917290 | A | G | 0.001318026 | YES |
| PGS000364_hmPOS_GRCh37 | 7 | 96896364 | C | T | 0.003088228 | YES |
| PGS000364_hmPOS_GRCh37 | 7 | 96896639 | A | G | 0.003123667 | YES |
| PGS000364_hmPOS_GRCh37 | 7 | 96900736 | A | G | 0.002212164 | YES |
| PGS000364_hmPOS_GRCh37 | 7 | 96909380 | G | A | 0.000493607 | YES |
| PGS000364_hmPOS_GRCh37 | 7 | 96911814 | T | G | 0.001506121 | YES |
| PGS000364_hmPOS_GRCh37 | 7 | 96913812 | T | G | 0.002912991 | YES |
| PGS000364_hmPOS_GRCh37 | 7 | 96913813 | C | T | 0.002954018 | YES |
| PGS000364_hmPOS_GRCh37 | 7 | 96916407 | C | T | 0.003166945 | YES |
| PGS000364_hmPOS_GRCh37 | 7 | 96922808 | C | G | 0.002731912 | NO |
| PGS000364_hmPOS_GRCh37 | 7 | 96923734 | A | G | 0.003678747 | YES |
| PGS000364_hmPOS_GRCh37 | 7 | 101696140 | G | T | 0.00021714 | YES |
| PGS000364_hmPOS_GRCh37 | 7 | 101698583 | T | C | 0.000511761 | YES |
| PGS000364_hmPOS_GRCh37 | 7 | 103778499 | T | C | 0.003316376 | YES |
| PGS000364_hmPOS_GRCh37 | 7 | 103790082 | G | A | 0.003533059 | YES |
| PGS000364_hmPOS_GRCh37 | 7 | 104223762 | G | A | 0.000171536 | YES |
| PGS000364_hmPOS_GRCh37 | 7 | 104223995 | C | T | 0.000184809 | YES |
| PGS000364_hmPOS_GRCh37 | 7 | 104224704 | G | A | 0.000194058 | YES |
| PGS000364_hmPOS_GRCh37 | 7 | 104225738 | G | A | 0.000256918 | YES |
| PGS000364_hmPOS_GRCh37 | 7 | 104227319 | T | A | 0.000192583 | NO |
| PGS000364_hmPOS_GRCh37 | 7 | 104230525 | G | A | 0.000153945 | YES |
| PGS000364_hmPOS_GRCh37 | 7 | 104231069 | G | A | 6.08505E-05 | YES |
| PGS000364_hmPOS_GRCh37 | 7 | 104233143 | G | C | 1.68548E-05 | NO |
| PGS000364_hmPOS_GRCh37 | 7 | 104233243 | C | A | 0.000117759 | YES |
| PGS000364_hmPOS_GRCh37 | 7 | 104235361 | G | T | 0.000169155 | YES |
| PGS000364_hmPOS_GRCh37 | 7 | 104236193 | G | A | 0.000159168 | YES |
| PGS000364_hmPOS_GRCh37 | 7 | 104236907 | T | G | 0.000280996 | YES |
| PGS000364_hmPOS_GRCh37 | 7 | 104243848 | C | G | 0.000184504 | NO |
| PGS000364_hmPOS_GRCh37 | 7 | 104254219 | C | A | 1.50436E-05 | YES |
| PGS000364_hmPOS_GRCh37 | 7 | 104256175 | C | A | 3.5013E-05 | YES |
| PGS000364_hmPOS_GRCh37 | 7 | 104256681 | G | A | 3.24369E-05 | YES |
| PGS000364_hmPOS_GRCh37 | 7 | 104270608 | T | A | 0.003206337 | NO |
| PGS000364_hmPOS_GRCh37 | 7 | 120921328 | G | A | -0.001051175 | YES |
| PGS000364_hmPOS_GRCh37 | 7 | 130800666 | G | A | 0.005717618 | YES |
| PGS000364_hmPOS_GRCh37 | 7 | 142098460 | T | G | 0.000989681 | YES |
| PGS000364_hmPOS_GRCh37 | 7 | 142099013 | T | C | 0.00152016 | YES |
| PGS000364_hmPOS_GRCh37 | 7 | 142099069 | A | C | 0.000949791 | YES |
| PGS000364_hmPOS_GRCh37 | 7 | 142099988 | G | A | 0.00095102 | YES |

| | | | | | | |
|---|---|---|---|---|---|---|
| PGS000364_hmPOS_GRCh37 | 7 | 142100535 | A | C | 9.82687E-05 | YES |
| PGS000364_hmPOS_GRCh37 | 7 | 142100784 | C | G | 0.001135691 | NO |
| PGS000364_hmPOS_GRCh37 | 7 | 142327259 | A | C | 0.008608466 | YES |
| PGS000364_hmPOS_GRCh37 | 7 | 148222226 | C | T | -0.000479147 | YES |
| PGS000364_hmPOS_GRCh37 | 7 | 148226296 | T | C | -1.59162E-05 | YES |
| PGS000364_hmPOS_GRCh37 | 7 | 148227242 | C | T | -0.000194651 | YES |
| PGS000364_hmPOS_GRCh37 | 7 | 148227838 | A | G | -3.46827E-05 | YES |
| PGS000364_hmPOS_GRCh37 | 7 | 148227866 | G | C | -5.95403E-05 | NO |
| PGS000364_hmPOS_GRCh37 | 7 | 148227961 | T | A | -0.000194092 | NO |
| PGS000364_hmPOS_GRCh37 | 7 | 148228101 | A | C | -2.06107E-05 | YES |
| PGS000364_hmPOS_GRCh37 | 7 | 148232643 | T | C | -7.67282E-05 | YES |
| PGS000364_hmPOS_GRCh37 | 7 | 148233113 | A | G | -0.000595472 | YES |
| PGS000364_hmPOS_GRCh37 | 7 | 148233221 | C | G | -0.000224717 | NO |
| PGS000364_hmPOS_GRCh37 | 7 | 148233242 | G | T | -0.000603154 | YES |
| PGS000364_hmPOS_GRCh37 | 7 | 148237744 | C | G | -0.000116118 | NO |
| PGS000364_hmPOS_GRCh37 | 7 | 148238713 | T | C | -0.000910926 | YES |
| PGS000364_hmPOS_GRCh37 | 7 | 148239599 | C | T | -0.000580714 | YES |
| PGS000364_hmPOS_GRCh37 | 7 | 154936358 | T | C | 0.001046569 | YES |
| PGS000364_hmPOS_GRCh37 | 7 | 154936498 | T | C | 0.011122681 | YES |
| PGS000364_hmPOS_GRCh37 | 7 | 154938461 | T | C | 0.001682776 | YES |
| PGS000364_hmPOS_GRCh37 | 7 | 154941695 | T | G | 0.002843475 | YES |
| PGS000364_hmPOS_GRCh37 | 7 | 154942356 | A | G | 0.001841039 | YES |
| PGS000364_hmPOS_GRCh37 | 7 | 154945975 | G | A | 0.002054707 | YES |
| PGS000364_hmPOS_GRCh37 | 7 | 155271237 | T | A | 0.011885861 | NO |
| PGS000364_hmPOS_GRCh37 | 7 | 156491132 | T | C | 0.024405766 | YES |
| PGS000364_hmPOS_GRCh37 | 7 | 156496219 | A | G | 0.015947347 | YES |
| PGS000364_hmPOS_GRCh37 | 7 | 156542640 | T | C | 0.020821118 | YES |
| PGS000364_hmPOS_GRCh37 | 7 | 156553970 | T | C | 0.024623763 | YES |
| PGS000364_hmPOS_GRCh37 | 7 | 156568235 | A | G | 0.013247404 | YES |
| PGS000364_hmPOS_GRCh37 | 7 | 156635091 | T | C | 0.02198672 | YES |
| PGS000364_hmPOS_GRCh37 | 8 | 5796901 | T | A | 0.010324372 | NO |
| PGS000364_hmPOS_GRCh37 | 8 | 5808499 | C | G | 0.011977788 | NO |
| PGS000364_hmPOS_GRCh37 | 8 | 6013715 | T | C | -0.001127362 | YES |
| PGS000364_hmPOS_GRCh37 | 8 | 18075298 | G | C | 0.001782436 | NO |
| PGS000364_hmPOS_GRCh37 | 8 | 18082384 | A | G | 0.008249384 | YES |
| PGS000364_hmPOS_GRCh37 | 8 | 18087430 | T | C | 0.004033129 | YES |
| PGS000364_hmPOS_GRCh37 | 8 | 25242721 | C | G | 0.01023425 | NO |
| PGS000364_hmPOS_GRCh37 | 8 | 26555395 | T | A | -2.12369E-05 | NO |
| PGS000364_hmPOS_GRCh37 | 8 | 26566229 | G | C | -0.000553499 | NO |
| PGS000364_hmPOS_GRCh37 | 8 | 74972392 | G | A | 0.022615767 | YES |
| PGS000364_hmPOS_GRCh37 | 8 | 94408811 | G | A | 0.006797133 | YES |
| PGS000364_hmPOS_GRCh37 | 8 | 117166281 | C | T | 0.002816127 | YES |
| PGS000364_hmPOS_GRCh37 | 8 | 119258160 | C | G | 0.003459313 | NO |
| PGS000364_hmPOS_GRCh37 | 8 | 125685942 | C | G | -0.020248991 | NO |
| PGS000364_hmPOS_GRCh37 | 8 | 125686407 | T | G | -0.007576784 | YES |
| PGS000364_hmPOS_GRCh37 | 8 | 125687170 | T | C | -0.020323032 | YES |
| PGS000364_hmPOS_GRCh37 | 8 | 125687498 | C | T | -0.019164782 | YES |
| PGS000364_hmPOS_GRCh37 | 8 | 125689659 | C | T | -0.020275728 | YES |
| PGS000364_hmPOS_GRCh37 | 8 | 125691531 | C | T | -0.020314503 | YES |
| PGS000364_hmPOS_GRCh37 | 8 | 125691816 | C | T | -0.021723838 | YES |
| PGS000364_hmPOS_GRCh37 | 8 | 125696512 | A | G | -0.020732889 | YES |
| PGS000364_hmPOS_GRCh37 | 8 | 125697808 | A | T | -0.020267309 | NO |
| PGS000364_hmPOS_GRCh37 | 8 | 125700089 | A | G | -0.020613197 | YES |
| PGS000364_hmPOS_GRCh37 | 8 | 125700925 | A | G | -0.002110791 | YES |
| PGS000364_hmPOS_GRCh37 | 8 | 125701953 | G | C | -0.021530503 | NO |
| PGS000364_hmPOS_GRCh37 | 8 | 127698883 | T | C | -0.007164453 | YES |
| PGS000364_hmPOS_GRCh37 | 8 | 127701948 | A | G | -0.006407244 | YES |
| PGS000364_hmPOS_GRCh37 | 8 | 127702342 | A | G | -0.006400117 | YES |
| PGS000364_hmPOS_GRCh37 | 8 | 127705842 | A | G | -0.008367464 | YES |
| PGS000364_hmPOS_GRCh37 | 8 | 134365641 | G | T | 0.007704621 | YES |
| PGS000364_hmPOS_GRCh37 | 8 | 139580731 | G | A | 0.001801848 | YES |
| PGS000364_hmPOS_GRCh37 | 8 | 141544459 | C | G | -0.001720909 | NO |
| PGS000364_hmPOS_GRCh37 | 8 | 142297429 | T | C | 0.018766937 | YES |

| | | | | | | |
|---|---|---|---|---|---|---|
| PGS000364_hmPOS_GRCh37 | 8 | 142338289 | T | C | 0.010191228 | YES |
| PGS000364_hmPOS_GRCh37 | 8 | 142370862 | A | G | 0.001631841 | YES |
| PGS000364_hmPOS_GRCh37 | 8 | 142370872 | G | C | 0.001631411 | NO |
| PGS000364_hmPOS_GRCh37 | 9 | 620438 | G | A | 0.008147877 | YES |
| PGS000364_hmPOS_GRCh37 | 9 | 2226092 | T | C | 0.006727061 | YES |
| PGS000364_hmPOS_GRCh37 | 9 | 4984549 | T | G | -0.00584461 | YES |
| PGS000364_hmPOS_GRCh37 | 9 | 4985879 | G | A | -0.004991199 | YES |
| PGS000364_hmPOS_GRCh37 | 9 | 4991975 | T | C | -0.014375758 | YES |
| PGS000364_hmPOS_GRCh37 | 9 | 4992030 | C | T | -0.014880658 | YES |
| PGS000364_hmPOS_GRCh37 | 9 | 4994052 | C | T | -0.013699947 | YES |
| PGS000364_hmPOS_GRCh37 | 9 | 4994387 | T | G | -0.014218221 | YES |
| PGS000364_hmPOS_GRCh37 | 9 | 4997224 | T | C | -0.013254983 | YES |
| PGS000364_hmPOS_GRCh37 | 9 | 5002188 | A | C | -0.001956069 | YES |
| PGS000364_hmPOS_GRCh37 | 9 | 5003093 | A | G | -0.015653218 | YES |
| PGS000364_hmPOS_GRCh37 | 9 | 5010471 | A | C | -0.006027107 | YES |
| PGS000364_hmPOS_GRCh37 | 9 | 5021529 | T | C | -0.003962651 | YES |
| PGS000364_hmPOS_GRCh37 | 9 | 5021738 | A | G | -0.017194527 | YES |
| PGS000364_hmPOS_GRCh37 | 9 | 5043007 | T | C | -0.004988685 | YES |
| PGS000364_hmPOS_GRCh37 | 9 | 5063199 | A | G | -0.007397396 | YES |
| PGS000364_hmPOS_GRCh37 | 9 | 5065642 | G | A | -0.024060282 | YES |
| PGS000364_hmPOS_GRCh37 | 9 | 5070905 | T | C | -0.001130862 | YES |
| PGS000364_hmPOS_GRCh37 | 9 | 5072846 | A | G | -0.002786033 | YES |
| PGS000364_hmPOS_GRCh37 | 9 | 5090641 | A | G | -0.000685809 | YES |
| PGS000364_hmPOS_GRCh37 | 9 | 5093646 | T | C | -0.001671597 | YES |
| PGS000364_hmPOS_GRCh37 | 9 | 5102190 | G | C | -0.000126303 | NO |
| PGS000364_hmPOS_GRCh37 | 9 | 5102910 | C | G | -0.001025946 | NO |
| PGS000364_hmPOS_GRCh37 | 9 | 5103942 | T | C | -0.001004579 | YES |
| PGS000364_hmPOS_GRCh37 | 9 | 5104574 | T | G | -0.001845449 | YES |
| PGS000364_hmPOS_GRCh37 | 9 | 5104640 | A | G | -0.000838364 | YES |
| PGS000364_hmPOS_GRCh37 | 9 | 5105140 | A | G | -0.001782568 | YES |
| PGS000364_hmPOS_GRCh37 | 9 | 5105645 | T | C | -0.000182355 | YES |
| PGS000364_hmPOS_GRCh37 | 9 | 5105651 | C | G | -0.000257603 | NO |
| PGS000364_hmPOS_GRCh37 | 9 | 5106023 | T | A | -0.000957245 | NO |
| PGS000364_hmPOS_GRCh37 | 9 | 5109485 | T | G | -0.001229305 | YES |
| PGS000364_hmPOS_GRCh37 | 9 | 5180604 | T | C | -0.000592087 | YES |
| PGS000364_hmPOS_GRCh37 | 9 | 13050293 | A | G | 0.00107647 | YES |
| PGS000364_hmPOS_GRCh37 | 9 | 13051154 | C | G | 0.00311458 | NO |
| PGS000364_hmPOS_GRCh37 | 9 | 13051193 | C | G | 0.002151697 | NO |
| PGS000364_hmPOS_GRCh37 | 9 | 13051238 | T | C | 0.002299644 | YES |
| PGS000364_hmPOS_GRCh37 | 9 | 13051712 | T | C | 0.000426139 | YES |
| PGS000364_hmPOS_GRCh37 | 9 | 22829912 | C | T | 0.003026108 | YES |
| PGS000364_hmPOS_GRCh37 | 9 | 22949466 | T | C | 0.000308312 | YES |
| PGS000364_hmPOS_GRCh37 | 9 | 22961293 | T | C | 0.00682079 | YES |
| PGS000364_hmPOS_GRCh37 | 9 | 22961577 | G | T | 0.006949275 | YES |
| PGS000364_hmPOS_GRCh37 | 9 | 23705736 | T | G | -0.004559969 | YES |
| PGS000364_hmPOS_GRCh37 | 9 | 27745826 | A | G | 0.001659986 | YES |
| PGS000364_hmPOS_GRCh37 | 9 | 38631465 | T | C | 0.000165171 | YES |
| PGS000364_hmPOS_GRCh37 | 9 | 38632373 | A | T | 0.00023604 | NO |
| PGS000364_hmPOS_GRCh37 | 9 | 38633280 | T | C | 0.000603807 | YES |
| PGS000364_hmPOS_GRCh37 | 9 | 38633358 | C | G | 0.000395452 | NO |
| PGS000364_hmPOS_GRCh37 | 9 | 38633384 | T | C | 0.000958302 | YES |
| PGS000364_hmPOS_GRCh37 | 9 | 38634798 | C | T | 0.000371948 | YES |
| PGS000364_hmPOS_GRCh37 | 9 | 38636168 | A | G | 5.44513E-05 | YES |
| PGS000364_hmPOS_GRCh37 | 9 | 38636183 | T | C | 0.000267632 | YES |
| PGS000364_hmPOS_GRCh37 | 9 | 38636216 | T | C | 6.06183E-05 | YES |
| PGS000364_hmPOS_GRCh37 | 9 | 38636226 | G | C | 0.000117625 | NO |
| PGS000364_hmPOS_GRCh37 | 9 | 38636331 | T | A | 0.003556251 | NO |
| PGS000364_hmPOS_GRCh37 | 9 | 38636463 | G | A | 0.003267681 | YES |
| PGS000364_hmPOS_GRCh37 | 9 | 71281591 | A | C | 0.005654291 | YES |
| PGS000364_hmPOS_GRCh37 | 9 | 78431574 | G | A | -6.59282E-05 | YES |
| PGS000364_hmPOS_GRCh37 | 9 | 78432428 | A | G | -0.000508618 | YES |
| PGS000364_hmPOS_GRCh37 | 9 | 78433674 | T | G | -0.000520035 | YES |
| PGS000364_hmPOS_GRCh37 | 9 | 78434101 | G | A | -0.000696165 | YES |

| | | | | | | |
|---|---|---|---|---|---|---|
| PGS000364_hmPOS_GRCh37 | 9 | 78435049 | C | G | -0.000598533 | NO |
| PGS000364_hmPOS_GRCh37 | 9 | 78444576 | G | A | -0.000662512 | YES |
| PGS000364_hmPOS_GRCh37 | 9 | 78453563 | T | C | -0.00078924 | YES |
| PGS000364_hmPOS_GRCh37 | 9 | 78453738 | C | G | -0.000762974 | NO |
| PGS000364_hmPOS_GRCh37 | 9 | 78458756 | T | C | -0.001651028 | YES |
| PGS000364_hmPOS_GRCh37 | 9 | 78460723 | C | T | -0.001184072 | YES |
| PGS000364_hmPOS_GRCh37 | 9 | 78461648 | C | A | -0.001146827 | YES |
| PGS000364_hmPOS_GRCh37 | 9 | 82032456 | T | C | 0.003012151 | YES |
| PGS000364_hmPOS_GRCh37 | 9 | 82041391 | G | A | 0.00021486 | YES |
| PGS000364_hmPOS_GRCh37 | 9 | 82046669 | T | G | 0.000502285 | YES |
| PGS000364_hmPOS_GRCh37 | 9 | 82367951 | T | A | 0.002354333 | NO |
| PGS000364_hmPOS_GRCh37 | 9 | 82387889 | G | A | 0.010136442 | YES |
| PGS000364_hmPOS_GRCh37 | 9 | 82428904 | A | G | 0.006870416 | NO |
| PGS000364_hmPOS_GRCh37 | 9 | 91048200 | A | G | 0.001020097 | YES |
| PGS000364_hmPOS_GRCh37 | 9 | 104199126 | A | G | 0.001178546 | YES |
| PGS000364_hmPOS_GRCh37 | 9 | 104226305 | G | C | 0.002070211 | NO |
| PGS000364_hmPOS_GRCh37 | 9 | 107291731 | A | G | 0.011158086 | YES |
| PGS000364_hmPOS_GRCh37 | 9 | 107291735 | A | T | 0.01117967 | NO |
| PGS000364_hmPOS_GRCh37 | 9 | 107292001 | C | T | 0.024158587 | YES |
| PGS000364_hmPOS_GRCh37 | 9 | 107292315 | C | T | 0.024579606 | YES |
| PGS000364_hmPOS_GRCh37 | 9 | 107292449 | T | C | 0.024640855 | YES |
| PGS000364_hmPOS_GRCh37 | 9 | 107292576 | G | A | 0.024613145 | YES |
| PGS000364_hmPOS_GRCh37 | 9 | 107292848 | A | G | 0.024597749 | YES |
| PGS000364_hmPOS_GRCh37 | 9 | 107292997 | A | C | 0.024571311 | YES |
| PGS000364_hmPOS_GRCh37 | 9 | 107293126 | A | G | 0.024693114 | YES |
| PGS000364_hmPOS_GRCh37 | 9 | 107293318 | C | T | 0.009564999 | YES |
| PGS000364_hmPOS_GRCh37 | 9 | 107293424 | C | T | 0.009580842 | YES |
| PGS000364_hmPOS_GRCh37 | 9 | 107293507 | A | G | 0.012979241 | YES |
| PGS000364_hmPOS_GRCh37 | 9 | 107294438 | T | C | 0.009932141 | YES |
| PGS000364_hmPOS_GRCh37 | 9 | 107294722 | G | T | 0.014810934 | YES |
| PGS000364_hmPOS_GRCh37 | 9 | 107297371 | A | C | 0.026395892 | YES |
| PGS000364_hmPOS_GRCh37 | 9 | 107337368 | G | A | 0.003638836 | YES |
| PGS000364_hmPOS_GRCh37 | 9 | 107337792 | C | T | 0.003470122 | YES |
| PGS000364_hmPOS_GRCh37 | 9 | 107338202 | C | A | 0.003409132 | YES |
| PGS000364_hmPOS_GRCh37 | 9 | 107339052 | A | T | 0.003255118 | NO |
| PGS000364_hmPOS_GRCh37 | 9 | 107340851 | A | G | 0.00342648 | YES |
| PGS000364_hmPOS_GRCh37 | 9 | 107340964 | C | T | 0.00342648 | YES |
| PGS000364_hmPOS_GRCh37 | 9 | 107341056 | C | T | 0.00367325 | YES |
| PGS000364_hmPOS_GRCh37 | 9 | 107341313 | T | C | 0.00342296 | YES |
| PGS000364_hmPOS_GRCh37 | 9 | 107341366 | G | A | 0.00342296 | YES |
| PGS000364_hmPOS_GRCh37 | 9 | 107341372 | A | C | 0.00342296 | YES |
| PGS000364_hmPOS_GRCh37 | 9 | 107341636 | A | G | 0.003426335 | YES |
| PGS000364_hmPOS_GRCh37 | 9 | 107341901 | A | T | 0.003426335 | NO |
| PGS000364_hmPOS_GRCh37 | 9 | 107341978 | G | A | 0.003426335 | YES |
| PGS000364_hmPOS_GRCh37 | 9 | 107342109 | G | A | 0.003425549 | YES |
| PGS000364_hmPOS_GRCh37 | 9 | 107342198 | G | A | 0.003425549 | YES |
| PGS000364_hmPOS_GRCh37 | 9 | 107342240 | A | C | 0.003425869 | YES |
| PGS000364_hmPOS_GRCh37 | 9 | 107342262 | C | A | 0.00342424 | YES |
| PGS000364_hmPOS_GRCh37 | 9 | 107342361 | C | A | 0.003505059 | YES |
| PGS000364_hmPOS_GRCh37 | 9 | 107343079 | T | C | 0.003426015 | YES |
| PGS000364_hmPOS_GRCh37 | 9 | 107344490 | A | G | 0.003423716 | YES |
| PGS000364_hmPOS_GRCh37 | 9 | 107344619 | C | T | 0.002882799 | YES |
| PGS000364_hmPOS_GRCh37 | 9 | 107344866 | T | C | 0.003322434 | YES |
| PGS000364_hmPOS_GRCh37 | 9 | 107344927 | G | A | 0.002936534 | YES |
| PGS000364_hmPOS_GRCh37 | 9 | 107345069 | A | G | 0.00285049 | YES |
| PGS000364_hmPOS_GRCh37 | 9 | 107345119 | T | C | 0.00285049 | YES |
| PGS000364_hmPOS_GRCh37 | 9 | 107345349 | T | C | 0.00285049 | YES |
| PGS000364_hmPOS_GRCh37 | 9 | 107347210 | A | C | 0.003324322 | YES |
| PGS000364_hmPOS_GRCh37 | 9 | 107348346 | A | T | 0.002851388 | NO |
| PGS000364_hmPOS_GRCh37 | 9 | 107348744 | A | G | 0.002851388 | YES |
| PGS000364_hmPOS_GRCh37 | 9 | 107349598 | A | G | 0.002851388 | YES |
| PGS000364_hmPOS_GRCh37 | 9 | 107349640 | G | A | 0.001323906 | YES |
| PGS000364_hmPOS_GRCh37 | 9 | 107349732 | C | A | 0.002849646 | YES |

| | | | | | | |
|---|---|---|---|---|---|---|
| PGS000364_hmPOS_GRCh37 | 9 | 107350187 | G | A | 0.003731121 | YES |
| PGS000364_hmPOS_GRCh37 | 9 | 107350211 | C | T | 0.003257832 | YES |
| PGS000364_hmPOS_GRCh37 | 9 | 107350418 | G | A | 0.001718116 | YES |
| PGS000364_hmPOS_GRCh37 | 9 | 107350474 | A | C | 0.001871911 | YES |
| PGS000364_hmPOS_GRCh37 | 9 | 107350544 | G | A | 0.001719491 | YES |
| PGS000364_hmPOS_GRCh37 | 9 | 107350630 | G | A | 0.002197256 | YES |
| PGS000364_hmPOS_GRCh37 | 9 | 107350761 | T | C | 0.001756344 | YES |
| PGS000364_hmPOS_GRCh37 | 9 | 107354089 | A | G | 0.001719883 | YES |
| PGS000364_hmPOS_GRCh37 | 9 | 107354655 | G | T | 0.001719578 | YES |
| PGS000364_hmPOS_GRCh37 | 9 | 107354870 | G | T | 0.001670515 | YES |
| PGS000364_hmPOS_GRCh37 | 9 | 107355655 | G | C | 0.002194538 | NO |
| PGS000364_hmPOS_GRCh37 | 9 | 107356155 | A | C | 0.001155601 | YES |
| PGS000364_hmPOS_GRCh37 | 9 | 107356689 | C | T | 0.001798553 | YES |
| PGS000364_hmPOS_GRCh37 | 9 | 107357501 | A | C | 0.001718225 | YES |
| PGS000364_hmPOS_GRCh37 | 9 | 107358185 | A | G | 0.000433469 | YES |
| PGS000364_hmPOS_GRCh37 | 9 | 107358222 | C | A | 0.001722699 | YES |
| PGS000364_hmPOS_GRCh37 | 9 | 107358376 | G | C | 0.002190264 | NO |
| PGS000364_hmPOS_GRCh37 | 9 | 107358806 | T | C | 0.001721498 | YES |
| PGS000364_hmPOS_GRCh37 | 9 | 107358983 | A | T | 0.002190264 | NO |
| PGS000364_hmPOS_GRCh37 | 9 | 107359427 | G | C | 0.001721476 | NO |
| PGS000364_hmPOS_GRCh37 | 9 | 107359547 | G | A | 0.002203286 | YES |
| PGS000364_hmPOS_GRCh37 | 9 | 107359802 | C | A | 0.001755686 | YES |
| PGS000364_hmPOS_GRCh37 | 9 | 107360014 | G | A | 0.001721498 | YES |
| PGS000364_hmPOS_GRCh37 | 9 | 107360171 | A | G | 0.001721498 | YES |
| PGS000364_hmPOS_GRCh37 | 9 | 107360373 | T | C | 0.002203357 | YES |
| PGS000364_hmPOS_GRCh37 | 9 | 107360396 | G | A | 0.001721498 | YES |
| PGS000364_hmPOS_GRCh37 | 9 | 107360826 | G | A | 0.000451091 | YES |
| PGS000364_hmPOS_GRCh37 | 9 | 107360851 | C | T | 0.001483258 | YES |
| PGS000364_hmPOS_GRCh37 | 9 | 107361129 | T | C | 0.00205653 | YES |
| PGS000364_hmPOS_GRCh37 | 9 | 107361599 | C | G | 0.003384779 | NO |
| PGS000364_hmPOS_GRCh37 | 9 | 107361710 | A | G | 0.001550266 | YES |
| PGS000364_hmPOS_GRCh37 | 9 | 107362004 | G | A | 0.001911881 | YES |
| PGS000364_hmPOS_GRCh37 | 9 | 107362114 | C | A | 0.001236185 | YES |
| PGS000364_hmPOS_GRCh37 | 9 | 107362302 | A | C | 0.001910845 | YES |
| PGS000364_hmPOS_GRCh37 | 9 | 107362735 | C | T | 0.001885758 | YES |
| PGS000364_hmPOS_GRCh37 | 9 | 107362820 | G | A | 0.00188845 | YES |
| PGS000364_hmPOS_GRCh37 | 9 | 107363266 | A | C | 0.003776828 | YES |
| PGS000364_hmPOS_GRCh37 | 9 | 107363738 | C | T | 0.003994639 | YES |
| PGS000364_hmPOS_GRCh37 | 9 | 107363777 | C | T | 0.003733851 | YES |
| PGS000364_hmPOS_GRCh37 | 9 | 107363792 | A | G | 0.004028513 | YES |
| PGS000364_hmPOS_GRCh37 | 9 | 107363808 | C | A | 0.003706493 | YES |
| PGS000364_hmPOS_GRCh37 | 9 | 107363850 | A | G | 0.002504542 | YES |
| PGS000364_hmPOS_GRCh37 | 9 | 107364027 | C | T | 0.002160851 | YES |
| PGS000364_hmPOS_GRCh37 | 9 | 107364421 | A | G | 0.001885848 | YES |
| PGS000364_hmPOS_GRCh37 | 9 | 107364924 | C | T | 0.001881677 | YES |
| PGS000364_hmPOS_GRCh37 | 9 | 107365325 | A | C | 0.00216353 | YES |
| PGS000364_hmPOS_GRCh37 | 9 | 107366471 | A | G | 0.001877016 | YES |
| PGS000364_hmPOS_GRCh37 | 9 | 107366884 | G | C | 0.002161415 | NO |
| PGS000364_hmPOS_GRCh37 | 9 | 107367008 | C | T | 0.001876837 | YES |
| PGS000364_hmPOS_GRCh37 | 9 | 107367841 | T | A | 0.000252346 | NO |
| PGS000364_hmPOS_GRCh37 | 9 | 107368257 | G | A | 0.001716023 | YES |
| PGS000364_hmPOS_GRCh37 | 9 | 107368368 | T | C | 0.001713385 | YES |
| PGS000364_hmPOS_GRCh37 | 9 | 107368682 | G | A | 0.001713058 | YES |
| PGS000364_hmPOS_GRCh37 | 9 | 107369564 | C | T | 0.000193668 | YES |
| PGS000364_hmPOS_GRCh37 | 9 | 107370792 | A | G | 0.000236706 | YES |
| PGS000364_hmPOS_GRCh37 | 9 | 107371359 | T | C | 0.000251251 | YES |
| PGS000364_hmPOS_GRCh37 | 9 | 107372721 | A | G | 0.000937268 | YES |
| PGS000364_hmPOS_GRCh37 | 9 | 107374371 | A | C | 0.000266808 | YES |
| PGS000364_hmPOS_GRCh37 | 9 | 107378964 | G | C | 0.00025488 | NO |
| PGS000364_hmPOS_GRCh37 | 9 | 107380215 | A | T | 0.00025488 | NO |
| PGS000364_hmPOS_GRCh37 | 9 | 107395750 | G | C | 0.001577903 | NO |
| PGS000364_hmPOS_GRCh37 | 9 | 107395947 | G | C | 0.003920377 | NO |
| PGS000364_hmPOS_GRCh37 | 9 | 107398743 | G | A | 0.004757614 | YES |

| | | | | | | |
|---|---|---|---|---|---|---|
| PGS000364_hmPOS_GRCh37 | 9 | 107401447 | C | T | 0.004421406 | YES |
| PGS000364_hmPOS_GRCh37 | 9 | 107402481 | C | T | 0.00442517 | YES |
| PGS000364_hmPOS_GRCh37 | 9 | 107409898 | A | G | 0.000769325 | YES |
| PGS000364_hmPOS_GRCh37 | 9 | 107424198 | A | G | 0.003849522 | YES |
| PGS000364_hmPOS_GRCh37 | 9 | 107430962 | A | T | 0.004535587 | NO |
| PGS000364_hmPOS_GRCh37 | 9 | 109218627 | T | C | 0.014888049 | YES |
| PGS000364_hmPOS_GRCh37 | 9 | 109218799 | C | T | 0.015509279 | YES |
| PGS000364_hmPOS_GRCh37 | 9 | 109243327 | C | A | 0.009330084 | YES |
| PGS000364_hmPOS_GRCh37 | 9 | 109244902 | C | T | 0.010244863 | YES |
| PGS000364_hmPOS_GRCh37 | 9 | 109254458 | C | T | 0.004131829 | YES |
| PGS000364_hmPOS_GRCh37 | 9 | 116820024 | A | G | -0.002827682 | YES |
| PGS000364_hmPOS_GRCh37 | 9 | 131575934 | A | G | 0.002324245 | YES |
| PGS000364_hmPOS_GRCh37 | 9 | 131599032 | A | G | 0.003491006 | YES |
| PGS000364_hmPOS_GRCh37 | 9 | 131620132 | G | A | 0.008668483 | YES |
| PGS000364_hmPOS_GRCh37 | 9 | 131639867 | G | C | 0.010488197 | NO |
| PGS000364_hmPOS_GRCh37 | 9 | 131726683 | G | C | 0.009190778 | NO |
| PGS000364_hmPOS_GRCh37 | 9 | 135908157 | T | A | 0.000112721 | NO |
| PGS000364_hmPOS_GRCh37 | 9 | 135909790 | G | T | 0.004012726 | YES |
| PGS000364_hmPOS_GRCh37 | 9 | 135909850 | C | A | 0.003864371 | YES |
| PGS000364_hmPOS_GRCh37 | 9 | 135910328 | C | T | 0.004018392 | YES |
| PGS000364_hmPOS_GRCh37 | 9 | 135912230 | C | T | 0.004009477 | YES |
| PGS000364_hmPOS_GRCh37 | 9 | 135913740 | G | C | 0.004000383 | NO |
| PGS000364_hmPOS_GRCh37 | 9 | 135914998 | C | T | 0.000977596 | YES |
| PGS000364_hmPOS_GRCh37 | 9 | 135925115 | C | T | 0.001047486 | YES |
| PGS000364_hmPOS_GRCh37 | 9 | 135926017 | G | A | 0.00074877 | YES |
| PGS000364_hmPOS_GRCh37 | 9 | 135926993 | G | A | 0.001270768 | YES |
| PGS000364_hmPOS_GRCh37 | 9 | 135929200 | C | A | 0.002653701 | YES |
| PGS000364_hmPOS_GRCh37 | 10 | 618486 | C | T | 0.000945876 | YES |
| PGS000364_hmPOS_GRCh37 | 10 | 625356 | A | G | 0.004404308 | YES |
| PGS000364_hmPOS_GRCh37 | 10 | 1340851 | G | C | 0.001439205 | NO |
| PGS000364_hmPOS_GRCh37 | 10 | 3485839 | A | G | 0.004645892 | YES |
| PGS000364_hmPOS_GRCh37 | 10 | 3494886 | T | G | 0.009731989 | YES |
| PGS000364_hmPOS_GRCh37 | 10 | 4670108 | T | C | 0.00192869 | YES |
| PGS000364_hmPOS_GRCh37 | 10 | 4670216 | G | C | 0.001288408 | NO |
| PGS000364_hmPOS_GRCh37 | 10 | 4672693 | T | C | 0.001520055 | YES |
| PGS000364_hmPOS_GRCh37 | 10 | 4672895 | T | C | 0.001847209 | YES |
| PGS000364_hmPOS_GRCh37 | 10 | 4673718 | G | A | 0.00094486 | YES |
| PGS000364_hmPOS_GRCh37 | 10 | 4674439 | C | A | 0.001362142 | YES |
| PGS000364_hmPOS_GRCh37 | 10 | 9972004 | T | C | 0.004282946 | YES |
| PGS000364_hmPOS_GRCh37 | 10 | 10027790 | T | C | 0.002961515 | YES |
| PGS000364_hmPOS_GRCh37 | 10 | 20179266 | G | A | 0.002028918 | YES |
| PGS000364_hmPOS_GRCh37 | 10 | 20180184 | C | A | 0.003164773 | YES |
| PGS000364_hmPOS_GRCh37 | 10 | 20202914 | T | A | 0.003127233 | NO |
| PGS000364_hmPOS_GRCh37 | 10 | 20203534 | T | C | 0.002885214 | YES |
| PGS000364_hmPOS_GRCh37 | 10 | 20206184 | A | G | 0.006283648 | YES |
| PGS000364_hmPOS_GRCh37 | 10 | 31740828 | T | C | 0.001690631 | YES |
| PGS000364_hmPOS_GRCh37 | 10 | 31755375 | T | C | 0.003804797 | YES |
| PGS000364_hmPOS_GRCh37 | 10 | 31769469 | G | A | 0.003835147 | YES |
| PGS000364_hmPOS_GRCh37 | 10 | 31820655 | C | A | 0.004583283 | YES |
| PGS000364_hmPOS_GRCh37 | 10 | 31823443 | A | G | 0.004473249 | YES |
| PGS000364_hmPOS_GRCh37 | 10 | 31841318 | C | T | 0.003545341 | YES |
| PGS000364_hmPOS_GRCh37 | 10 | 33854166 | T | C | -0.003979807 | YES |
| PGS000364_hmPOS_GRCh37 | 10 | 33856151 | A | G | -0.000197687 | YES |
| PGS000364_hmPOS_GRCh37 | 10 | 59290336 | A | G | 0.006506593 | YES |
| PGS000364_hmPOS_GRCh37 | 10 | 60145079 | G | A | 0.011374575 | YES |
| PGS000364_hmPOS_GRCh37 | 10 | 60147270 | A | G | 0.007018813 | YES |
| PGS000364_hmPOS_GRCh37 | 10 | 60147784 | A | G | 0.006885833 | YES |
| PGS000364_hmPOS_GRCh37 | 10 | 60148692 | G | A | 0.006754995 | YES |
| PGS000364_hmPOS_GRCh37 | 10 | 60151026 | G | A | 0.006513557 | YES |
| PGS000364_hmPOS_GRCh37 | 10 | 60156584 | A | G | 0.006020852 | YES |
| PGS000364_hmPOS_GRCh37 | 10 | 60157339 | T | C | 0.005963829 | YES |
| PGS000364_hmPOS_GRCh37 | 10 | 60161172 | A | G | 0.007943364 | YES |
| PGS000364_hmPOS_GRCh37 | 10 | 60161640 | G | A | 0.008166484 | YES |

| | | | | | | |
|---|---|---|---|---|---|---|
| PGS000364_hmPOS_GRCh37 | 10 | 60167882 | T | C | 0.001368374 | YES |
| PGS000364_hmPOS_GRCh37 | 10 | 60168003 | A | G | 0.000199434 | YES |
| PGS000364_hmPOS_GRCh37 | 10 | 60169880 | T | G | 0.004605443 | YES |
| PGS000364_hmPOS_GRCh37 | 10 | 60219560 | A | G | -0.007245987 | YES |
| PGS000364_hmPOS_GRCh37 | 10 | 60588885 | C | A | 0.002954394 | YES |
| PGS000364_hmPOS_GRCh37 | 10 | 62572565 | A | G | 0.002259241 | YES |
| PGS000364_hmPOS_GRCh37 | 10 | 64671764 | A | G | -0.007353113 | YES |
| PGS000364_hmPOS_GRCh37 | 10 | 64743728 | T | C | -0.002512661 | YES |
| PGS000364_hmPOS_GRCh37 | 10 | 64745058 | A | T | -0.005441522 | NO |
| PGS000364_hmPOS_GRCh37 | 10 | 64745059 | A | T | -0.005442058 | NO |
| PGS000364_hmPOS_GRCh37 | 10 | 64745589 | C | T | -0.005930858 | YES |
| PGS000364_hmPOS_GRCh37 | 10 | 64753309 | A | T | -0.004275905 | NO |
| PGS000364_hmPOS_GRCh37 | 10 | 64754145 | G | T | -0.005472189 | YES |
| PGS000364_hmPOS_GRCh37 | 10 | 64757342 | C | T | -0.002318791 | YES |
| PGS000364_hmPOS_GRCh37 | 10 | 67786957 | C | T | 0.015625872 | YES |
| PGS000364_hmPOS_GRCh37 | 10 | 67794066 | C | T | 0.014488255 | YES |
| PGS000364_hmPOS_GRCh37 | 10 | 67797918 | T | C | 0.011300528 | YES |
| PGS000364_hmPOS_GRCh37 | 10 | 67804202 | A | C | 0.011068127 | YES |
| PGS000364_hmPOS_GRCh37 | 10 | 67806009 | T | G | 0.011404333 | YES |
| PGS000364_hmPOS_GRCh37 | 10 | 67806210 | C | T | 0.009358671 | YES |
| PGS000364_hmPOS_GRCh37 | 10 | 67806844 | A | T | 0.010637079 | NO |
| PGS000364_hmPOS_GRCh37 | 10 | 67807756 | A | T | 0.010583107 | NO |
| PGS000364_hmPOS_GRCh37 | 10 | 67809091 | C | T | 0.010542804 | YES |
| PGS000364_hmPOS_GRCh37 | 10 | 67809536 | G | A | 0.010489876 | YES |
| PGS000364_hmPOS_GRCh37 | 10 | 67809646 | A | G | 0.010474481 | YES |
| PGS000364_hmPOS_GRCh37 | 10 | 67810031 | G | A | 0.009133771 | YES |
| PGS000364_hmPOS_GRCh37 | 10 | 67810251 | C | T | 0.009837251 | YES |
| PGS000364_hmPOS_GRCh37 | 10 | 67810327 | T | C | 0.009823992 | YES |
| PGS000364_hmPOS_GRCh37 | 10 | 67810493 | G | A | 0.009832625 | YES |
| PGS000364_hmPOS_GRCh37 | 10 | 67812643 | T | C | 0.010994217 | YES |
| PGS000364_hmPOS_GRCh37 | 10 | 67813956 | C | A | 0.009902462 | YES |
| PGS000364_hmPOS_GRCh37 | 10 | 67814548 | C | A | 0.009521948 | YES |
| PGS000364_hmPOS_GRCh37 | 10 | 67815753 | T | C | 0.009518841 | YES |
| PGS000364_hmPOS_GRCh37 | 10 | 67815927 | C | T | 0.009570785 | YES |
| PGS000364_hmPOS_GRCh37 | 10 | 67816170 | A | G | 0.009777486 | YES |
| PGS000364_hmPOS_GRCh37 | 10 | 67818765 | G | A | 0.009422381 | YES |
| PGS000364_hmPOS_GRCh37 | 10 | 67818958 | C | T | 0.00954454 | YES |
| PGS000364_hmPOS_GRCh37 | 10 | 67820224 | T | A | 0.009434282 | NO |
| PGS000364_hmPOS_GRCh37 | 10 | 67820766 | G | A | 0.009376909 | YES |
| PGS000364_hmPOS_GRCh37 | 10 | 67821207 | A | G | 0.009380804 | YES |
| PGS000364_hmPOS_GRCh37 | 10 | 67821634 | C | A | 0.009374416 | YES |
| PGS000364_hmPOS_GRCh37 | 10 | 67822758 | A | T | 0.009300635 | NO |
| PGS000364_hmPOS_GRCh37 | 10 | 67822926 | T | C | 0.009390483 | YES |
| PGS000364_hmPOS_GRCh37 | 10 | 67826749 | A | G | 0.009359244 | YES |
| PGS000364_hmPOS_GRCh37 | 10 | 67831067 | C | G | 0.009479379 | NO |
| PGS000364_hmPOS_GRCh37 | 10 | 67832274 | T | C | 0.009601317 | YES |
| PGS000364_hmPOS_GRCh37 | 10 | 67833198 | G | A | 0.007909799 | YES |
| PGS000364_hmPOS_GRCh37 | 10 | 67835039 | C | A | 0.007998414 | YES |
| PGS000364_hmPOS_GRCh37 | 10 | 67835117 | C | T | 0.008184812 | YES |
| PGS000364_hmPOS_GRCh37 | 10 | 67835356 | C | T | 0.008178721 | YES |
| PGS000364_hmPOS_GRCh37 | 10 | 67835750 | A | T | 0.008001406 | NO |
| PGS000364_hmPOS_GRCh37 | 10 | 67837187 | A | C | 0.008079661 | YES |
| PGS000364_hmPOS_GRCh37 | 10 | 67838194 | C | T | 0.008017932 | YES |
| PGS000364_hmPOS_GRCh37 | 10 | 67838899 | G | A | 0.007970569 | YES |
| PGS000364_hmPOS_GRCh37 | 10 | 67852050 | C | T | 0.000156236 | YES |
| PGS000364_hmPOS_GRCh37 | 10 | 67853279 | G | A | 0.002292699 | YES |
| PGS000364_hmPOS_GRCh37 | 10 | 67853610 | C | A | 0.002529409 | YES |
| PGS000364_hmPOS_GRCh37 | 10 | 67854376 | G | A | 0.002103086 | YES |
| PGS000364_hmPOS_GRCh37 | 10 | 67855521 | C | T | 0.002070813 | YES |
| PGS000364_hmPOS_GRCh37 | 10 | 67863801 | G | A | 0.002101481 | YES |
| PGS000364_hmPOS_GRCh37 | 10 | 67864181 | C | T | 0.00178965 | YES |
| PGS000364_hmPOS_GRCh37 | 10 | 67871841 | A | G | 0.000915773 | YES |
| PGS000364_hmPOS_GRCh37 | 10 | 67873775 | A | G | 0.000786792 | YES |

| | | | | | | |
|---|---|---|---|---|---|---|
| PGS000364_hmPOS_GRCh37 | 10 | 67876471 | A | G | 0.000758974 | YES |
| PGS000364_hmPOS_GRCh37 | 10 | 67876596 | A | G | 0.000779676 | YES |
| PGS000364_hmPOS_GRCh37 | 10 | 77971481 | G | A | 0.000416637 | YES |
| PGS000364_hmPOS_GRCh37 | 10 | 78116099 | T | G | 0.002646686 | YES |
| PGS000364_hmPOS_GRCh37 | 10 | 82241838 | A | G | 0.001970863 | YES |
| PGS000364_hmPOS_GRCh37 | 10 | 87767732 | T | A | 0.01104214 | NO |
| PGS000364_hmPOS_GRCh37 | 10 | 90449236 | A | G | 0.001706153 | YES |
| PGS000364_hmPOS_GRCh37 | 10 | 108388923 | C | T | 0.002882401 | YES |
| PGS000364_hmPOS_GRCh37 | 10 | 112225713 | C | T | 0.004883423 | YES |
| PGS000364_hmPOS_GRCh37 | 10 | 122296627 | T | A | -0.002576989 | NO |
| PGS000364_hmPOS_GRCh37 | 10 | 122311938 | G | T | -0.00141452 | YES |
| PGS000364_hmPOS_GRCh37 | 10 | 122311974 | A | T | -0.001067753 | NO |
| PGS000364_hmPOS_GRCh37 | 10 | 122325454 | G | A | 0.003360585 | YES |
| PGS000364_hmPOS_GRCh37 | 10 | 122339342 | A | G | -0.003166583 | YES |
| PGS000364_hmPOS_GRCh37 | 10 | 122339357 | C | T | -0.002380162 | YES |
| PGS000364_hmPOS_GRCh37 | 10 | 123547144 | C | T | 0.023290963 | YES |
| PGS000364_hmPOS_GRCh37 | 10 | 123692726 | A | T | 4.30125E-05 | NO |
| PGS000364_hmPOS_GRCh37 | 10 | 129439607 | T | G | 0.001186627 | YES |
| PGS000364_hmPOS_GRCh37 | 10 | 132134542 | G | C | 0.002473511 | NO |
| PGS000364_hmPOS_GRCh37 | 11 | 890866 | A | G | 0.003540354 | YES |
| PGS000364_hmPOS_GRCh37 | 11 | 11215989 | C | T | 0.004051628 | YES |
| PGS000364_hmPOS_GRCh37 | 11 | 17589129 | A | C | 0.004158855 | YES |
| PGS000364_hmPOS_GRCh37 | 11 | 17597822 | T | C | 0.002423018 | YES |
| PGS000364_hmPOS_GRCh37 | 11 | 17599093 | C | T | 2.93932258838003e-7 | YES |
| PGS000364_hmPOS_GRCh37 | 11 | 17657888 | T | C | 0.002317512 | YES |
| PGS000364_hmPOS_GRCh37 | 11 | 18193107 | T | C | 0.000310108 | YES |
| PGS000364_hmPOS_GRCh37 | 11 | 18210849 | T | C | 0.000594349 | YES |
| PGS000364_hmPOS_GRCh37 | 11 | 36432830 | G | T | 0.002314209 | YES |
| PGS000364_hmPOS_GRCh37 | 11 | 36434542 | A | G | 0.00031839 | YES |
| PGS000364_hmPOS_GRCh37 | 11 | 36437164 | T | C | 0.000416813 | YES |
| PGS000364_hmPOS_GRCh37 | 11 | 36437868 | G | T | 0.006490954 | YES |
| PGS000364_hmPOS_GRCh37 | 11 | 37397873 | G | A | 0.013021324 | YES |
| PGS000364_hmPOS_GRCh37 | 11 | 40143344 | A | G | 3.88296E-05 | YES |
| PGS000364_hmPOS_GRCh37 | 11 | 40143958 | C | T | 0.001665043 | YES |
| PGS000364_hmPOS_GRCh37 | 11 | 40146249 | A | G | 0.004065151 | YES |
| PGS000364_hmPOS_GRCh37 | 11 | 40146979 | A | C | 0.002208067 | YES |
| PGS000364_hmPOS_GRCh37 | 11 | 40147836 | G | A | 0.000317058 | YES |
| PGS000364_hmPOS_GRCh37 | 11 | 60515110 | G | C | 0.002054569 | NO |
| PGS000364_hmPOS_GRCh37 | 11 | 60515320 | G | A | 0.001022156 | YES |
| PGS000364_hmPOS_GRCh37 | 11 | 70386369 | G | A | 0.023070916 | YES |
| PGS000364_hmPOS_GRCh37 | 11 | 70387342 | C | G | 0.007739648 | NO |
| PGS000364_hmPOS_GRCh37 | 11 | 70390284 | G | A | 0.007604408 | YES |
| PGS000364_hmPOS_GRCh37 | 11 | 70390462 | A | G | 0.008016401 | YES |
| PGS000364_hmPOS_GRCh37 | 11 | 70390719 | G | A | 0.007784219 | YES |
| PGS000364_hmPOS_GRCh37 | 11 | 78425906 | T | C | 0.000856069 | YES |
| PGS000364_hmPOS_GRCh37 | 11 | 83055365 | A | G | -0.003529562 | YES |
| PGS000364_hmPOS_GRCh37 | 11 | 84663249 | C | T | 0.000551216 | YES |
| PGS000364_hmPOS_GRCh37 | 11 | 84695711 | C | T | 0.004788359 | YES |
| PGS000364_hmPOS_GRCh37 | 11 | 84736164 | A | G | 0.008575508 | YES |
| PGS000364_hmPOS_GRCh37 | 11 | 84782539 | G | A | 0.001711554 | YES |
| PGS000364_hmPOS_GRCh37 | 11 | 84811390 | G | A | 0.00317246 | YES |
| PGS000364_hmPOS_GRCh37 | 11 | 84845925 | A | G | 0.006999928 | YES |
| PGS000364_hmPOS_GRCh37 | 11 | 84856250 | T | C | 0.006903595 | YES |
| PGS000364_hmPOS_GRCh37 | 11 | 84861343 | T | C | 0.002424295 | YES |
| PGS000364_hmPOS_GRCh37 | 11 | 84872234 | G | T | 0.017908856 | YES |
| PGS000364_hmPOS_GRCh37 | 11 | 84878532 | C | T | 0.008370636 | YES |
| PGS000364_hmPOS_GRCh37 | 11 | 84883931 | C | T | 0.009670522 | YES |
| PGS000364_hmPOS_GRCh37 | 11 | 84899827 | C | T | 0.002571498 | YES |
| PGS000364_hmPOS_GRCh37 | 11 | 84903637 | T | C | 0.005081453 | YES |
| PGS000364_hmPOS_GRCh37 | 11 | 84904742 | C | T | 0.002001867 | YES |
| PGS000364_hmPOS_GRCh37 | 11 | 84904818 | T | G | 0.002452823 | YES |
| PGS000364_hmPOS_GRCh37 | 11 | 85057399 | T | A | 0.004506751 | NO |
| PGS000364_hmPOS_GRCh37 | 11 | 85122449 | A | G | 0.000814569 | YES |

| | | | | | | |
|---|---|---|---|---|---|---|
| PGS000364_hmPOS_GRCh37 | 11 | 85159102 | G | A | 5.60103228086639e-6 | YES |
| PGS000364_hmPOS_GRCh37 | 11 | 85227439 | G | A | 0.004777737 | YES |
| PGS000364_hmPOS_GRCh37 | 11 | 85227751 | C | T | 0.000509368 | YES |
| PGS000364_hmPOS_GRCh37 | 11 | 85228358 | G | A | 0.003167696 | YES |
| PGS000364_hmPOS_GRCh37 | 11 | 85229456 | C | A | 0.003668395 | YES |
| PGS000364_hmPOS_GRCh37 | 11 | 85230491 | T | C | 0.00308908 | YES |
| PGS000364_hmPOS_GRCh37 | 11 | 85231475 | A | G | 0.004650775 | YES |
| PGS000364_hmPOS_GRCh37 | 11 | 85231961 | G | A | 0.004655878 | YES |
| PGS000364_hmPOS_GRCh37 | 11 | 85232508 | A | G | 0.000362739 | YES |
| PGS000364_hmPOS_GRCh37 | 11 | 85232636 | G | A | 0.004655734 | YES |
| PGS000364_hmPOS_GRCh37 | 11 | 85233493 | A | C | 0.003474199 | YES |
| PGS000364_hmPOS_GRCh37 | 11 | 85236332 | A | G | 0.004661021 | YES |
| PGS000364_hmPOS_GRCh37 | 11 | 85237627 | A | C | 0.00347731 | YES |
| PGS000364_hmPOS_GRCh37 | 11 | 85238368 | T | G | 0.001924128 | YES |
| PGS000364_hmPOS_GRCh37 | 11 | 85238677 | A | T | 0.001956568 | NO |
| PGS000364_hmPOS_GRCh37 | 11 | 85238886 | G | A | 0.000319446 | YES |
| PGS000364_hmPOS_GRCh37 | 11 | 85238901 | A | G | 0.003418018 | YES |
| PGS000364_hmPOS_GRCh37 | 11 | 85239189 | A | T | 0.001912512 | NO |
| PGS000364_hmPOS_GRCh37 | 11 | 85239290 | C | T | 0.001912242 | YES |
| PGS000364_hmPOS_GRCh37 | 11 | 85239562 | T | A | 0.003487947 | NO |
| PGS000364_hmPOS_GRCh37 | 11 | 85239988 | A | C | 0.003488329 | YES |
| PGS000364_hmPOS_GRCh37 | 11 | 85240145 | A | G | 0.003509249 | YES |
| PGS000364_hmPOS_GRCh37 | 11 | 85241143 | T | C | 0.002047627 | YES |
| PGS000364_hmPOS_GRCh37 | 11 | 85243213 | G | A | 0.004288071 | YES |
| PGS000364_hmPOS_GRCh37 | 11 | 85244314 | T | C | 0.00137661 | YES |
| PGS000364_hmPOS_GRCh37 | 11 | 85245151 | C | T | 0.002447225 | YES |
| PGS000364_hmPOS_GRCh37 | 11 | 85245772 | A | G | 0.000969751 | YES |
| PGS000364_hmPOS_GRCh37 | 11 | 85247973 | A | G | 1.72006E-05 | YES |
| PGS000364_hmPOS_GRCh37 | 11 | 85248141 | T | C | 6.15994217040283e-6 | YES |
| PGS000364_hmPOS_GRCh37 | 11 | 85255186 | T | G | 0.001393923 | YES |
| PGS000364_hmPOS_GRCh37 | 11 | 85255613 | T | A | 8.88635E-05 | NO |
| PGS000364_hmPOS_GRCh37 | 11 | 85255842 | T | C | 0.000953997 | YES |
| PGS000364_hmPOS_GRCh37 | 11 | 85260059 | A | G | 0.001523136 | YES |
| PGS000364_hmPOS_GRCh37 | 11 | 85260220 | C | T | 0.003088338 | YES |
| PGS000364_hmPOS_GRCh37 | 11 | 85267366 | G | A | 0.001194719 | YES |
| PGS000364_hmPOS_GRCh37 | 11 | 85284406 | A | T | 0.005242753 | NO |
| PGS000364_hmPOS_GRCh37 | 11 | 85287944 | G | A | 0.006307931 | YES |
| PGS000364_hmPOS_GRCh37 | 11 | 85295227 | G | C | 0.001628896 | NO |
| PGS000364_hmPOS_GRCh37 | 11 | 92558024 | T | C | -0.001335069 | YES |
| PGS000364_hmPOS_GRCh37 | 11 | 92558342 | A | G | -0.000765637 | YES |
| PGS000364_hmPOS_GRCh37 | 11 | 93080928 | T | C | 0.011643446 | YES |
| PGS000364_hmPOS_GRCh37 | 11 | 93087365 | A | G | 0.006197936 | YES |
| PGS000364_hmPOS_GRCh37 | 11 | 93087780 | C | A | 0.005963016 | YES |
| PGS000364_hmPOS_GRCh37 | 11 | 93090608 | A | T | 0.010868575 | NO |
| PGS000364_hmPOS_GRCh37 | 11 | 93096055 | C | T | 0.011254422 | YES |
| PGS000364_hmPOS_GRCh37 | 11 | 93097210 | G | C | 0.002483969 | NO |
| PGS000364_hmPOS_GRCh37 | 11 | 93105965 | C | T | 0.002488212 | YES |
| PGS000364_hmPOS_GRCh37 | 11 | 93106593 | C | A | 0.002498845 | YES |
| PGS000364_hmPOS_GRCh37 | 11 | 93107458 | G | A | 0.002441484 | YES |
| PGS000364_hmPOS_GRCh37 | 11 | 93108546 | A | G | 0.002406397 | YES |
| PGS000364_hmPOS_GRCh37 | 11 | 93112727 | C | G | 0.002568126 | NO |
| PGS000364_hmPOS_GRCh37 | 11 | 93114521 | A | G | 0.002382307 | YES |
| PGS000364_hmPOS_GRCh37 | 11 | 93131667 | A | C | 0.014184779 | YES |
| PGS000364_hmPOS_GRCh37 | 11 | 93134216 | G | A | 0.014348322 | YES |
| PGS000364_hmPOS_GRCh37 | 11 | 93140251 | A | T | 0.003486006 | NO |
| PGS000364_hmPOS_GRCh37 | 11 | 93140593 | A | G | 0.003423164 | YES |
| PGS000364_hmPOS_GRCh37 | 11 | 93140986 | G | T | 0.003506593 | YES |
| PGS000364_hmPOS_GRCh37 | 11 | 102586758 | T | C | 0.002423902 | YES |
| PGS000364_hmPOS_GRCh37 | 11 | 102588331 | G | A | 0.002322265 | YES |
| PGS000364_hmPOS_GRCh37 | 11 | 102588479 | A | C | 0.002359942 | YES |
| PGS000364_hmPOS_GRCh37 | 11 | 102588552 | C | T | 0.002360646 | YES |
| PGS000364_hmPOS_GRCh37 | 11 | 102588607 | A | G | 0.002357368 | YES |
| PGS000364_hmPOS_GRCh37 | 11 | 102589642 | T | C | 0.002397834 | YES |

| | | | | | | |
|---|---|---|---|---|---|---|
| PGS000364_hmPOS_GRCh37 | 11 | 102592854 | A | G | 0.002391824 | YES |
| PGS000364_hmPOS_GRCh37 | 11 | 102595258 | A | G | 0.002383429 | YES |
| PGS000364_hmPOS_GRCh37 | 11 | 102595388 | C | A | 0.0029483 | YES |
| PGS000364_hmPOS_GRCh37 | 11 | 102595408 | G | A | 0.0029483 | YES |
| PGS000364_hmPOS_GRCh37 | 11 | 102596583 | C | G | 0.00238043 | NO |
| PGS000364_hmPOS_GRCh37 | 11 | 102600377 | T | C | 0.003042633 | YES |
| PGS000364_hmPOS_GRCh37 | 11 | 102605880 | C | T | 0.024584442 | YES |
| PGS000364_hmPOS_GRCh37 | 11 | 102619072 | G | A | 0.005238336 | YES |
| PGS000364_hmPOS_GRCh37 | 11 | 102626401 | G | A | 0.024230101 | YES |
| PGS000364_hmPOS_GRCh37 | 11 | 112749621 | C | A | 0.009431873 | YES |
| PGS000364_hmPOS_GRCh37 | 11 | 112754441 | G | A | 0.008564432 | YES |
| PGS000364_hmPOS_GRCh37 | 11 | 123171353 | C | T | -0.002336281 | YES |
| PGS000364_hmPOS_GRCh37 | 11 | 125246545 | G | A | 9.4715E-05 | YES |
| PGS000364_hmPOS_GRCh37 | 11 | 131449365 | C | A | 0.022795356 | YES |
| PGS000364_hmPOS_GRCh37 | 11 | 131449439 | G | A | 0.023445733 | YES |
| PGS000364_hmPOS_GRCh37 | 11 | 131451947 | T | C | 0.023393511 | YES |
| PGS000364_hmPOS_GRCh37 | 11 | 131452418 | T | C | 0.023735005 | YES |
| PGS000364_hmPOS_GRCh37 | 11 | 131452511 | T | C | 0.023660025 | YES |
| PGS000364_hmPOS_GRCh37 | 11 | 131452912 | C | T | 0.023399741 | YES |
| PGS000364_hmPOS_GRCh37 | 11 | 131453013 | G | A | 0.023246312 | YES |
| PGS000364_hmPOS_GRCh37 | 11 | 131454686 | C | G | 0.023503076 | NO |
| PGS000364_hmPOS_GRCh37 | 11 | 131459065 | T | G | 0.011192467 | YES |
| PGS000364_hmPOS_GRCh37 | 11 | 131459900 | G | A | 0.020142264 | YES |
| PGS000364_hmPOS_GRCh37 | 11 | 131461173 | A | G | 0.021830435 | YES |
| PGS000364_hmPOS_GRCh37 | 11 | 131461473 | A | C | 0.01875157 | YES |
| PGS000364_hmPOS_GRCh37 | 11 | 131766987 | T | C | -0.007348433 | YES |
| PGS000364_hmPOS_GRCh37 | 12 | 1917188 | C | T | 0.003153511 | YES |
| PGS000364_hmPOS_GRCh37 | 12 | 3060224 | C | A | 0.005457662 | YES |
| PGS000364_hmPOS_GRCh37 | 12 | 4318723 | T | C | -0.008041023 | YES |
| PGS000364_hmPOS_GRCh37 | 12 | 6545611 | A | G | 0.004912894 | YES |
| PGS000364_hmPOS_GRCh37 | 12 | 7412101 | G | A | 0.014091314 | YES |
| PGS000364_hmPOS_GRCh37 | 12 | 9782811 | T | C | 0.010709846 | YES |
| PGS000364_hmPOS_GRCh37 | 12 | 16192611 | G | A | 0.018841117 | YES |
| PGS000364_hmPOS_GRCh37 | 12 | 19801876 | G | T | -0.000400377 | YES |
| PGS000364_hmPOS_GRCh37 | 12 | 19802337 | A | G | -0.000194244 | YES |
| PGS000364_hmPOS_GRCh37 | 12 | 19803880 | G | T | -0.00077434 | YES |
| PGS000364_hmPOS_GRCh37 | 12 | 19805071 | G | C | -0.000745929 | NO |
| PGS000364_hmPOS_GRCh37 | 12 | 19807016 | C | T | -0.000691067 | YES |
| PGS000364_hmPOS_GRCh37 | 12 | 19810161 | G | T | -0.001768408 | YES |
| PGS000364_hmPOS_GRCh37 | 12 | 27015040 | A | G | 0.000460151 | YES |
| PGS000364_hmPOS_GRCh37 | 12 | 27015992 | T | C | 0.002665198 | YES |
| PGS000364_hmPOS_GRCh37 | 12 | 27020704 | T | C | 0.004567045 | YES |
| PGS000364_hmPOS_GRCh37 | 12 | 27023804 | C | T | 0.003165469 | YES |
| PGS000364_hmPOS_GRCh37 | 12 | 56871314 | A | G | -0.001139632 | YES |
| PGS000364_hmPOS_GRCh37 | 12 | 56874522 | A | C | -0.001706044 | YES |
| PGS000364_hmPOS_GRCh37 | 12 | 56878120 | A | G | -0.001701671 | YES |
| PGS000364_hmPOS_GRCh37 | 12 | 56879548 | T | C | -0.001667292 | YES |
| PGS000364_hmPOS_GRCh37 | 12 | 64367622 | G | A | 0.026860676 | YES |
| PGS000364_hmPOS_GRCh37 | 12 | 64392815 | T | C | 0.002440499 | YES |
| PGS000364_hmPOS_GRCh37 | 12 | 102871356 | C | T | 0.013377739 | YES |
| PGS000364_hmPOS_GRCh37 | 12 | 118288488 | A | G | -0.002089097 | YES |
| PGS000364_hmPOS_GRCh37 | 12 | 118290238 | A | T | -0.005679245 | NO |
| PGS000364_hmPOS_GRCh37 | 12 | 118426505 | A | G | 0.003741739 | YES |
| PGS000364_hmPOS_GRCh37 | 12 | 119472411 | T | C | 0.004994363 | YES |
| PGS000364_hmPOS_GRCh37 | 12 | 119496871 | G | A | 0.008272297 | YES |
| PGS000364_hmPOS_GRCh37 | 12 | 119498993 | A | C | 0.008714137 | YES |
| PGS000364_hmPOS_GRCh37 | 12 | 119507217 | A | G | 0.012495199 | YES |
| PGS000364_hmPOS_GRCh37 | 12 | 119507266 | T | A | 0.008799403 | NO |
| PGS000364_hmPOS_GRCh37 | 12 | 119508185 | C | T | 0.012587822 | YES |
| PGS000364_hmPOS_GRCh37 | 12 | 119508430 | C | T | 0.012597576 | YES |
| PGS000364_hmPOS_GRCh37 | 12 | 119509633 | G | C | 0.00873157 | NO |
| PGS000364_hmPOS_GRCh37 | 12 | 119510036 | A | G | 0.00881446 | YES |
| PGS000364_hmPOS_GRCh37 | 12 | 119516437 | G | C | 0.009231229 | NO |

| | | | | | | |
|---|---|---|---|---|---|---|
| PGS000364_hmPOS_GRCh37 | 12 | 119516507 | C | T | 0.00925714 | YES |
| PGS000364_hmPOS_GRCh37 | 12 | 119516574 | T | A | 0.012446491 | NO |
| PGS000364_hmPOS_GRCh37 | 12 | 119520723 | T | C | 0.0095281 | YES |
| PGS000364_hmPOS_GRCh37 | 12 | 119521009 | C | A | 0.009462947 | YES |
| PGS000364_hmPOS_GRCh37 | 12 | 119521863 | A | G | 0.008084306 | YES |
| PGS000364_hmPOS_GRCh37 | 12 | 120195082 | G | C | -0.000151183 | NO |
| PGS000364_hmPOS_GRCh37 | 12 | 123437129 | T | C | 0.000779713 | YES |
| PGS000364_hmPOS_GRCh37 | 12 | 123832946 | A | G | 0.008960039 | YES |
| PGS000364_hmPOS_GRCh37 | 12 | 123919299 | A | C | 0.016930218 | YES |
| PGS000364_hmPOS_GRCh37 | 12 | 125808945 | G | A | 0.010513493 | YES |
| PGS000364_hmPOS_GRCh37 | 12 | 125811518 | G | A | 0.009394413 | YES |
| PGS000364_hmPOS_GRCh37 | 12 | 125812398 | G | A | 0.008375983 | YES |
| PGS000364_hmPOS_GRCh37 | 12 | 125813882 | T | C | 0.000472525 | YES |
| PGS000364_hmPOS_GRCh37 | 12 | 125813939 | G | C | 0.000866323 | NO |
| PGS000364_hmPOS_GRCh37 | 12 | 132127109 | T | C | 0.016649605 | YES |
| PGS000364_hmPOS_GRCh37 | 13 | 23460115 | T | G | 0.001085299 | YES |
| PGS000364_hmPOS_GRCh37 | 13 | 23464296 | A | G | 0.010456423 | YES |
| PGS000364_hmPOS_GRCh37 | 13 | 40339158 | T | C | 0.006410078 | YES |
| PGS000364_hmPOS_GRCh37 | 13 | 40512443 | T | C | 0.002394144 | YES |
| PGS000364_hmPOS_GRCh37 | 13 | 45277921 | T | C | 0.001155501 | YES |
| PGS000364_hmPOS_GRCh37 | 13 | 45279711 | A | G | 7.52455425617749e-6 | YES |
| PGS000364_hmPOS_GRCh37 | 13 | 45281435 | G | T | 0.001421014 | YES |
| PGS000364_hmPOS_GRCh37 | 13 | 45282032 | T | G | 0.001334659 | YES |
| PGS000364_hmPOS_GRCh37 | 13 | 45282108 | C | T | 0.000219097 | YES |
| PGS000364_hmPOS_GRCh37 | 13 | 45282123 | T | G | 0.001533343 | YES |
| PGS000364_hmPOS_GRCh37 | 13 | 45282165 | C | T | 0.000156926 | YES |
| PGS000364_hmPOS_GRCh37 | 13 | 45283021 | A | C | 0.001323191 | YES |
| PGS000364_hmPOS_GRCh37 | 13 | 45283387 | C | T | 0.001563176 | YES |
| PGS000364_hmPOS_GRCh37 | 13 | 45290219 | A | G | 0.000673085 | YES |
| PGS000364_hmPOS_GRCh37 | 13 | 45290220 | A | C | 0.000673085 | YES |
| PGS000364_hmPOS_GRCh37 | 13 | 45290248 | C | A | 0.000458859 | YES |
| PGS000364_hmPOS_GRCh37 | 13 | 47358540 | C | T | -0.000106101 | YES |
| PGS000364_hmPOS_GRCh37 | 13 | 47366033 | A | G | -0.000126135 | YES |
| PGS000364_hmPOS_GRCh37 | 13 | 47388527 | T | C | -0.000387992 | YES |
| PGS000364_hmPOS_GRCh37 | 13 | 50945705 | C | T | 0.004599067 | YES |
| PGS000364_hmPOS_GRCh37 | 13 | 51036012 | G | A | 0.003793568 | YES |
| PGS000364_hmPOS_GRCh37 | 13 | 54862440 | C | T | 0.002974922 | YES |
| PGS000364_hmPOS_GRCh37 | 13 | 66230992 | C | T | 0.005802564 | YES |
| PGS000364_hmPOS_GRCh37 | 13 | 66271716 | A | G | 0.009659613 | YES |
| PGS000364_hmPOS_GRCh37 | 13 | 66278609 | G | A | 0.009839628 | YES |
| PGS000364_hmPOS_GRCh37 | 13 | 66282880 | G | C | 0.005561777 | NO |
| PGS000364_hmPOS_GRCh37 | 13 | 66303531 | T | C | 0.005742591 | YES |
| PGS000364_hmPOS_GRCh37 | 13 | 66326352 | G | A | 0.005567126 | YES |
| PGS000364_hmPOS_GRCh37 | 13 | 66338384 | G | A | 0.000482008 | YES |
| PGS000364_hmPOS_GRCh37 | 13 | 67189769 | A | C | 0.00379078 | YES |
| PGS000364_hmPOS_GRCh37 | 13 | 74438910 | G | A | 0.006694225 | YES |
| PGS000364_hmPOS_GRCh37 | 13 | 74440131 | A | G | 0.008780631 | YES |
| PGS000364_hmPOS_GRCh37 | 13 | 74441736 | G | A | 0.004806782 | YES |
| PGS000364_hmPOS_GRCh37 | 13 | 84597229 | A | C | 0.001761835 | YES |
| PGS000364_hmPOS_GRCh37 | 13 | 85604370 | C | T | 0.002920811 | YES |
| PGS000364_hmPOS_GRCh37 | 13 | 96741468 | A | G | 0.005566999 | YES |
| PGS000364_hmPOS_GRCh37 | 13 | 96812343 | G | A | 0.005885899 | YES |
| PGS000364_hmPOS_GRCh37 | 13 | 97229113 | A | G | 0.01157628 | YES |
| PGS000364_hmPOS_GRCh37 | 13 | 104428829 | C | T | 0.004844436 | YES |
| PGS000364_hmPOS_GRCh37 | 13 | 104428900 | G | T | 0.004884695 | YES |
| PGS000364_hmPOS_GRCh37 | 13 | 104430136 | G | A | 0.004928515 | YES |
| PGS000364_hmPOS_GRCh37 | 13 | 104436201 | G | A | 0.004999741 | YES |
| PGS000364_hmPOS_GRCh37 | 13 | 104519202 | C | G | 0.009026399 | NO |
| PGS000364_hmPOS_GRCh37 | 13 | 109721968 | G | A | -0.000573096 | YES |
| PGS000364_hmPOS_GRCh37 | 14 | 21050409 | G | T | 0.006676162 | YES |
| PGS000364_hmPOS_GRCh37 | 14 | 21051027 | T | A | 0.000257945 | NO |
| PGS000364_hmPOS_GRCh37 | 14 | 23774161 | T | C | 0.001603206 | YES |
| PGS000364_hmPOS_GRCh37 | 14 | 23775111 | A | G | 0.000692392 | YES |

| | | | | | | |
|---|---|---|---|---|---|---|
| PGS000364_hmPOS_GRCh37 | 14 | 26184001 | A | G | 0.004801433 | YES |
| PGS000364_hmPOS_GRCh37 | 14 | 26185349 | A | T | 0.002120163 | NO |
| PGS000364_hmPOS_GRCh37 | 14 | 26186599 | C | T | 0.001424939 | YES |
| PGS000364_hmPOS_GRCh37 | 14 | 26189124 | A | G | 0.000992735 | YES |
| PGS000364_hmPOS_GRCh37 | 14 | 26189729 | C | A | 0.000847757 | YES |
| PGS000364_hmPOS_GRCh37 | 14 | 26190548 | T | G | 0.000723751 | YES |
| PGS000364_hmPOS_GRCh37 | 14 | 26191258 | G | A | 0.000446213 | YES |
| PGS000364_hmPOS_GRCh37 | 14 | 26191931 | T | C | 0.000500199 | YES |
| PGS000364_hmPOS_GRCh37 | 14 | 29386736 | G | C | 0.006770499 | NO |
| PGS000364_hmPOS_GRCh37 | 14 | 29405932 | C | T | 0.028113662 | YES |
| PGS000364_hmPOS_GRCh37 | 14 | 29429110 | G | A | 0.026892601 | YES |
| PGS000364_hmPOS_GRCh37 | 14 | 29450699 | A | G | 0.029154439 | YES |
| PGS000364_hmPOS_GRCh37 | 14 | 33499208 | C | G | 0.005105269 | NO |
| PGS000364_hmPOS_GRCh37 | 14 | 33501778 | G | A | 0.004067879 | YES |
| PGS000364_hmPOS_GRCh37 | 14 | 33503533 | G | C | 0.00387316 | NO |
| PGS000364_hmPOS_GRCh37 | 14 | 33505728 | G | T | 0.00084138 | YES |
| PGS000364_hmPOS_GRCh37 | 14 | 33506256 | T | A | 0.000912719 | NO |
| PGS000364_hmPOS_GRCh37 | 14 | 33507902 | T | A | 0.001286114 | NO |
| PGS000364_hmPOS_GRCh37 | 14 | 33513041 | G | A | 0.002329511 | YES |
| PGS000364_hmPOS_GRCh37 | 14 | 34262647 | A | G | 0.000235989 | YES |
| PGS000364_hmPOS_GRCh37 | 14 | 62081087 | G | A | 0.023296441 | YES |
| PGS000364_hmPOS_GRCh37 | 14 | 65356268 | A | G | 0.001006381 | YES |
| PGS000364_hmPOS_GRCh37 | 14 | 65369329 | T | C | 0.000983133 | YES |
| PGS000364_hmPOS_GRCh37 | 14 | 65370321 | G | T | 0.000972449 | YES |
| PGS000364_hmPOS_GRCh37 | 14 | 65372829 | C | T | 0.00107651 | YES |
| PGS000364_hmPOS_GRCh37 | 14 | 65378856 | C | T | 0.000957917 | YES |
| PGS000364_hmPOS_GRCh37 | 14 | 65380579 | T | C | 0.001092056 | YES |
| PGS000364_hmPOS_GRCh37 | 14 | 65381464 | G | C | 0.001129062 | NO |
| PGS000364_hmPOS_GRCh37 | 14 | 65388641 | C | T | 0.000628845 | YES |
| PGS000364_hmPOS_GRCh37 | 14 | 65396648 | G | C | 0.00144435 | NO |
| PGS000364_hmPOS_GRCh37 | 14 | 65403525 | A | T | 0.001440913 | NO |
| PGS000364_hmPOS_GRCh37 | 14 | 65403891 | G | C | 0.000623985 | NO |
| PGS000364_hmPOS_GRCh37 | 14 | 65404059 | A | G | 0.000627079 | YES |
| PGS000364_hmPOS_GRCh37 | 14 | 66292820 | T | C | 0.003802127 | YES |
| PGS000364_hmPOS_GRCh37 | 14 | 72799119 | G | T | 0.011985692 | YES |
| PGS000364_hmPOS_GRCh37 | 14 | 75676634 | A | G | 0.00184262 | YES |
| PGS000364_hmPOS_GRCh37 | 14 | 75677105 | G | T | 0.001306487 | YES |
| PGS000364_hmPOS_GRCh37 | 14 | 75679642 | G | C | 0.000541175 | NO |
| PGS000364_hmPOS_GRCh37 | 14 | 76031506 | A | G | -0.001932329 | YES |
| PGS000364_hmPOS_GRCh37 | 14 | 76031623 | A | G | -0.000132426 | YES |
| PGS000364_hmPOS_GRCh37 | 14 | 77788838 | A | G | -0.009537091 | YES |
| PGS000364_hmPOS_GRCh37 | 14 | 77792824 | C | G | -0.011818284 | NO |
| PGS000364_hmPOS_GRCh37 | 14 | 77799841 | A | G | -0.015068169 | YES |
| PGS000364_hmPOS_GRCh37 | 14 | 77801282 | G | T | -0.012968818 | YES |
| PGS000364_hmPOS_GRCh37 | 14 | 77801431 | C | T | -0.013833465 | YES |
| PGS000364_hmPOS_GRCh37 | 14 | 77801654 | C | G | -0.0142032 | NO |
| PGS000364_hmPOS_GRCh37 | 14 | 77809939 | T | C | -0.002638762 | YES |
| PGS000364_hmPOS_GRCh37 | 14 | 77810713 | C | G | -0.002675257 | NO |
| PGS000364_hmPOS_GRCh37 | 14 | 77836113 | A | G | -0.001735854 | YES |
| PGS000364_hmPOS_GRCh37 | 14 | 77841167 | A | G | -0.001130308 | YES |
| PGS000364_hmPOS_GRCh37 | 14 | 78369285 | T | G | 0.004377864 | YES |
| PGS000364_hmPOS_GRCh37 | 14 | 78371403 | G | A | 0.002156225 | YES |
| PGS000364_hmPOS_GRCh37 | 14 | 88978364 | A | T | 0.000632943 | NO |
| PGS000364_hmPOS_GRCh37 | 14 | 97955062 | G | A | 0.000861893 | YES |
| PGS000364_hmPOS_GRCh37 | 14 | 97958182 | A | G | 0.001120308 | YES |
| PGS000364_hmPOS_GRCh37 | 14 | 97962078 | A | G | 0.000800805 | YES |
| PGS000364_hmPOS_GRCh37 | 15 | 33978034 | G | T | 0.000389445 | YES |
| PGS000364_hmPOS_GRCh37 | 15 | 57971825 | G | A | 0.000348204 | YES |
| PGS000364_hmPOS_GRCh37 | 15 | 57973554 | G | A | 0.000351456 | YES |
| PGS000364_hmPOS_GRCh37 | 15 | 57975031 | T | C | 0.000486862 | YES |
| PGS000364_hmPOS_GRCh37 | 15 | 61614338 | T | C | -0.001485146 | YES |
| PGS000364_hmPOS_GRCh37 | 15 | 63219735 | A | G | -0.000268233 | YES |
| PGS000364_hmPOS_GRCh37 | 15 | 63220061 | T | C | -0.000317525 | YES |

| | | | | | | |
|---|---|---|---|---|---|---|
| PGS000364_hmPOS_GRCh37 | 15 | 72081280 | T | G | 0.002565888 | YES |
| PGS000364_hmPOS_GRCh37 | 15 | 76474749 | A | G | 0.003008002 | YES |
| PGS000364_hmPOS_GRCh37 | 15 | 76478925 | T | C | 0.003255966 | YES |
| PGS000364_hmPOS_GRCh37 | 15 | 76484947 | T | A | 0.006168636 | NO |
| PGS000364_hmPOS_GRCh37 | 15 | 76485536 | G | A | 0.006223414 | YES |
| PGS000364_hmPOS_GRCh37 | 15 | 76487507 | C | G | 0.00624991 | NO |
| PGS000364_hmPOS_GRCh37 | 15 | 76503295 | C | A | 0.002803993 | YES |
| PGS000364_hmPOS_GRCh37 | 15 | 76507206 | A | G | 0.006183455 | YES |
| PGS000364_hmPOS_GRCh37 | 15 | 76545844 | G | A | 0.004765596 | YES |
| PGS000364_hmPOS_GRCh37 | 15 | 76546933 | A | G | 0.007282979 | YES |
| PGS000364_hmPOS_GRCh37 | 15 | 76554755 | T | C | 0.004998787 | YES |
| PGS000364_hmPOS_GRCh37 | 15 | 76555986 | T | C | 0.005482475 | YES |
| PGS000364_hmPOS_GRCh37 | 15 | 76558870 | C | T | 0.004639579 | YES |
| PGS000364_hmPOS_GRCh37 | 15 | 76568209 | A | G | 0.002494346 | YES |
| PGS000364_hmPOS_GRCh37 | 15 | 80657732 | T | C | 0.000558608 | YES |
| PGS000364_hmPOS_GRCh37 | 15 | 80710748 | T | C | 0.009977249 | YES |
| PGS000364_hmPOS_GRCh37 | 15 | 89147342 | A | G | -0.007683361 | YES |
| PGS000364_hmPOS_GRCh37 | 15 | 89637418 | A | C | 0.000861799 | YES |
| PGS000364_hmPOS_GRCh37 | 15 | 89638091 | T | C | 0.00094344 | YES |
| PGS000364_hmPOS_GRCh37 | 15 | 89638440 | A | G | 0.000621548 | YES |
| PGS000364_hmPOS_GRCh37 | 15 | 89644938 | A | G | 0.004235712 | YES |
| PGS000364_hmPOS_GRCh37 | 15 | 89645230 | G | A | 0.006150634 | YES |
| PGS000364_hmPOS_GRCh37 | 15 | 89645942 | T | C | 0.004586053 | YES |
| PGS000364_hmPOS_GRCh37 | 15 | 89648943 | T | C | 0.017339192 | YES |
| PGS000364_hmPOS_GRCh37 | 15 | 89649276 | G | A | 0.016957928 | YES |
| PGS000364_hmPOS_GRCh37 | 15 | 89649508 | A | G | 0.017053926 | YES |
| PGS000364_hmPOS_GRCh37 | 15 | 89649704 | T | C | 0.00789207 | YES |
| PGS000364_hmPOS_GRCh37 | 15 | 89650795 | T | C | 0.009217704 | YES |
| PGS000364_hmPOS_GRCh37 | 15 | 89651254 | C | G | 0.017424969 | NO |
| PGS000364_hmPOS_GRCh37 | 15 | 89652138 | A | G | 0.017411686 | YES |
| PGS000364_hmPOS_GRCh37 | 15 | 89652552 | A | G | 0.017721359 | YES |
| PGS000364_hmPOS_GRCh37 | 15 | 89652572 | G | A | 0.007940088 | YES |
| PGS000364_hmPOS_GRCh37 | 15 | 89653262 | G | A | 0.00672086 | YES |
| PGS000364_hmPOS_GRCh37 | 15 | 89653764 | G | A | 0.006309179 | YES |
| PGS000364_hmPOS_GRCh37 | 15 | 89654197 | A | T | 0.017114707 | NO |
| PGS000364_hmPOS_GRCh37 | 15 | 89654764 | G | A | 0.006425936 | YES |
| PGS000364_hmPOS_GRCh37 | 15 | 89654976 | C | T | 0.016364508 | YES |
| PGS000364_hmPOS_GRCh37 | 15 | 89655405 | G | C | 0.00801344 | NO |
| PGS000364_hmPOS_GRCh37 | 15 | 89658421 | A | G | 0.004789794 | YES |
| PGS000364_hmPOS_GRCh37 | 15 | 89659425 | T | C | 0.014714998 | YES |
| PGS000364_hmPOS_GRCh37 | 15 | 89659451 | A | G | 0.005553364 | YES |
| PGS000364_hmPOS_GRCh37 | 15 | 89659467 | G | C | 0.005562703 | NO |
| PGS000364_hmPOS_GRCh37 | 15 | 89660319 | C | T | 0.005607125 | YES |
| PGS000364_hmPOS_GRCh37 | 15 | 89660476 | C | T | 0.0050143 | YES |
| PGS000364_hmPOS_GRCh37 | 15 | 89660915 | A | G | 0.014541967 | YES |
| PGS000364_hmPOS_GRCh37 | 15 | 89660987 | A | G | 0.004815387 | YES |
| PGS000364_hmPOS_GRCh37 | 15 | 89661039 | T | C | 0.003879761 | YES |
| PGS000364_hmPOS_GRCh37 | 15 | 89661746 | G | T | 0.015082482 | YES |
| PGS000364_hmPOS_GRCh37 | 15 | 89663644 | A | G | 0.01460769 | YES |
| PGS000364_hmPOS_GRCh37 | 15 | 89665802 | T | G | 0.007519449 | YES |
| PGS000364_hmPOS_GRCh37 | 15 | 89665823 | T | G | 0.007390938 | YES |
| PGS000364_hmPOS_GRCh37 | 15 | 89666056 | C | G | 0.008115838 | NO |
| PGS000364_hmPOS_GRCh37 | 15 | 89666734 | A | G | 0.017027519 | YES |
| PGS000364_hmPOS_GRCh37 | 15 | 89670504 | T | C | 0.005521341 | YES |
| PGS000364_hmPOS_GRCh37 | 15 | 90325061 | T | C | 0.005700167 | YES |
| PGS000364_hmPOS_GRCh37 | 15 | 90326422 | C | A | 0.006171353 | YES |
| PGS000364_hmPOS_GRCh37 | 15 | 90367960 | A | C | -0.00473111 | YES |
| PGS000364_hmPOS_GRCh37 | 15 | 93758050 | T | C | 0.009044824 | YES |
| PGS000364_hmPOS_GRCh37 | 15 | 95100958 | T | A | 0.000256387 | NO |
| PGS000364_hmPOS_GRCh37 | 15 | 95110896 | A | C | -0.011793801 | YES |
| PGS000364_hmPOS_GRCh37 | 15 | 95125867 | A | G | 0.001682646 | YES |
| PGS000364_hmPOS_GRCh37 | 15 | 96092442 | A | C | 0.002863858 | YES |
| PGS000364_hmPOS_GRCh37 | 15 | 96092607 | A | G | 0.003351325 | YES |

| | | | | | | |
|---|---|---|---|---|---|---|
| PGS000364_hmPOS_GRCh37 | 15 | 96093765 | A | G | 0.003352274 | YES |
| PGS000364_hmPOS_GRCh37 | 15 | 96094552 | T | G | 0.002365142 | YES |
| PGS000364_hmPOS_GRCh37 | 15 | 96094553 | A | T | 0.002342478 | NO |
| PGS000364_hmPOS_GRCh37 | 15 | 96095240 | G | A | 0.002307945 | YES |
| PGS000364_hmPOS_GRCh37 | 15 | 98020603 | T | C | 0.006992336 | YES |
| PGS000364_hmPOS_GRCh37 | 15 | 98202210 | A | G | 0.003721747 | YES |
| PGS000364_hmPOS_GRCh37 | 15 | 98215184 | A | G | 0.000601976 | YES |
| PGS000364_hmPOS_GRCh37 | 16 | 6451325 | A | G | -0.001292349 | YES |
| PGS000364_hmPOS_GRCh37 | 16 | 6452267 | A | G | -0.000435956 | YES |
| PGS000364_hmPOS_GRCh37 | 16 | 6452536 | G | A | -0.000445806 | YES |
| PGS000364_hmPOS_GRCh37 | 16 | 6452861 | C | G | -0.005798569 | NO |
| PGS000364_hmPOS_GRCh37 | 16 | 6453057 | C | G | -0.001631798 | NO |
| PGS000364_hmPOS_GRCh37 | 16 | 6456970 | G | C | -0.001386881 | NO |
| PGS000364_hmPOS_GRCh37 | 16 | 6457020 | C | T | -0.001777345 | YES |
| PGS000364_hmPOS_GRCh37 | 16 | 6457312 | G | A | -0.000482719 | YES |
| PGS000364_hmPOS_GRCh37 | 16 | 6457453 | G | A | -0.000528196 | YES |
| PGS000364_hmPOS_GRCh37 | 16 | 6460948 | G | C | -0.005361136 | NO |
| PGS000364_hmPOS_GRCh37 | 16 | 6462964 | T | C | -0.004707048 | YES |
| PGS000364_hmPOS_GRCh37 | 16 | 6462970 | T | C | -0.004486955 | YES |
| PGS000364_hmPOS_GRCh37 | 16 | 6463298 | G | C | -0.003126431 | NO |
| PGS000364_hmPOS_GRCh37 | 16 | 6463599 | A | G | -0.003295335 | YES |
| PGS000364_hmPOS_GRCh37 | 16 | 6464203 | C | A | -0.002040302 | YES |
| PGS000364_hmPOS_GRCh37 | 16 | 6464448 | G | A | -0.003367841 | YES |
| PGS000364_hmPOS_GRCh37 | 16 | 6464894 | G | C | -0.002737087 | NO |
| PGS000364_hmPOS_GRCh37 | 16 | 8857096 | A | G | 0.000709914 | YES |
| PGS000364_hmPOS_GRCh37 | 16 | 8901081 | C | A | 0.000734821 | YES |
| PGS000364_hmPOS_GRCh37 | 16 | 10856535 | A | G | 0.000161359 | YES |
| PGS000364_hmPOS_GRCh37 | 16 | 11225876 | T | C | 0.002546454 | YES |
| PGS000364_hmPOS_GRCh37 | 16 | 19413295 | G | A | -0.002624826 | YES |
| PGS000364_hmPOS_GRCh37 | 16 | 24953913 | T | C | 0.004243541 | YES |
| PGS000364_hmPOS_GRCh37 | 16 | 48654334 | C | T | 0.000349387 | YES |
| PGS000364_hmPOS_GRCh37 | 16 | 49253077 | T | C | 0.006364565 | YES |
| PGS000364_hmPOS_GRCh37 | 16 | 49268473 | A | G | 0.006336659 | YES |
| PGS000364_hmPOS_GRCh37 | 16 | 50813997 | A | G | -0.007009303 | YES |
| PGS000364_hmPOS_GRCh37 | 16 | 50815432 | T | C | -0.00555307 | YES |
| PGS000364_hmPOS_GRCh37 | 16 | 50817489 | A | G | -0.005603944 | YES |
| PGS000364_hmPOS_GRCh37 | 16 | 66933184 | G | A | 0.002988007 | YES |
| PGS000364_hmPOS_GRCh37 | 16 | 67489313 | A | G | 0.012071283 | YES |
| PGS000364_hmPOS_GRCh37 | 16 | 84656502 | C | T | 0.001488734 | YES |
| PGS000364_hmPOS_GRCh37 | 16 | 84662704 | G | T | 0.014877544 | YES |
| PGS000364_hmPOS_GRCh37 | 16 | 84664322 | A | G | -0.003115408 | YES |
| PGS000364_hmPOS_GRCh37 | 16 | 84668807 | G | C | -0.002974868 | NO |
| PGS000364_hmPOS_GRCh37 | 16 | 84671009 | A | G | 0.0005347 | YES |
| PGS000364_hmPOS_GRCh37 | 16 | 84683281 | T | G | -0.004551913 | YES |
| PGS000364_hmPOS_GRCh37 | 16 | 84683484 | A | G | -0.010740539 | YES |
| PGS000364_hmPOS_GRCh37 | 16 | 84685509 | T | A | -0.005102433 | NO |
| PGS000364_hmPOS_GRCh37 | 16 | 85617849 | C | T | 0.006897432 | YES |
| PGS000364_hmPOS_GRCh37 | 17 | 4706356 | T | C | 0.01035995 | YES |
| PGS000364_hmPOS_GRCh37 | 17 | 4710591 | A | G | 0.006405486 | YES |
| PGS000364_hmPOS_GRCh37 | 17 | 4710733 | C | T | 0.006476545 | YES |
| PGS000364_hmPOS_GRCh37 | 17 | 4712230 | T | C | 0.001904046 | YES |
| PGS000364_hmPOS_GRCh37 | 17 | 4712617 | T | C | 0.00211839 | YES |
| PGS000364_hmPOS_GRCh37 | 17 | 4725211 | C | T | 0.008264815 | YES |
| PGS000364_hmPOS_GRCh37 | 17 | 4732302 | G | C | 0.000239096 | NO |
| PGS000364_hmPOS_GRCh37 | 17 | 4732535 | C | T | 0.001302191 | YES |
| PGS000364_hmPOS_GRCh37 | 17 | 4943176 | G | A | 0.001751144 | YES |
| PGS000364_hmPOS_GRCh37 | 17 | 4949078 | A | T | 0.003675891 | NO |
| PGS000364_hmPOS_GRCh37 | 17 | 5765025 | A | G | -0.000476163 | YES |
| PGS000364_hmPOS_GRCh37 | 17 | 6327771 | C | T | -0.002855718 | YES |
| PGS000364_hmPOS_GRCh37 | 17 | 25597712 | A | G | -0.000771711 | YES |
| PGS000364_hmPOS_GRCh37 | 17 | 25599090 | G | C | -0.000439573 | NO |
| PGS000364_hmPOS_GRCh37 | 17 | 43972176 | G | C | -0.007500319 | NO |
| PGS000364_hmPOS_GRCh37 | 17 | 48124582 | G | T | 0.00212754 | YES |

| | | | | | | |
|---|---|---|---|---|---|---|
| PGS000364_hmPOS_GRCh37 | 17 | 53667178 | G | A | 0.001559372 | YES |
| PGS000364_hmPOS_GRCh37 | 17 | 53713610 | A | G | 0.010924765 | YES |
| PGS000364_hmPOS_GRCh37 | 17 | 53727616 | T | C | 0.004508573 | YES |
| PGS000364_hmPOS_GRCh37 | 17 | 53789045 | C | T | 0.00543526 | YES |
| PGS000364_hmPOS_GRCh37 | 17 | 53795175 | A | T | 0.003421244 | NO |
| PGS000364_hmPOS_GRCh37 | 17 | 76699587 | T | C | 0.013842488 | YES |
| PGS000364_hmPOS_GRCh37 | 17 | 76802098 | T | C | 0.006593668 | YES |
| PGS000364_hmPOS_GRCh37 | 17 | 76836414 | A | G | 0.001497243 | YES |
| PGS000364_hmPOS_GRCh37 | 17 | 77310798 | C | T | 0.012236998 | YES |
| PGS000364_hmPOS_GRCh37 | 17 | 78879613 | C | G | 0.008939302 | NO |
| PGS000364_hmPOS_GRCh37 | 17 | 78881927 | A | G | 0.000555891 | YES |
| PGS000364_hmPOS_GRCh37 | 17 | 78882840 | G | A | 0.004159515 | YES |
| PGS000364_hmPOS_GRCh37 | 17 | 78890238 | T | C | 0.004056748 | YES |
| PGS000364_hmPOS_GRCh37 | 17 | 78890955 | A | C | 0.003651071 | YES |
| PGS000364_hmPOS_GRCh37 | 17 | 78890975 | T | C | 0.00367978 | YES |
| PGS000364_hmPOS_GRCh37 | 17 | 78891030 | C | G | 0.003304991 | NO |
| PGS000364_hmPOS_GRCh37 | 17 | 78891741 | T | C | 0.006444394 | YES |
| PGS000364_hmPOS_GRCh37 | 17 | 78895301 | T | G | 0.000261784 | YES |
| PGS000364_hmPOS_GRCh37 | 17 | 78896488 | T | C | 0.000475737 | YES |
| PGS000364_hmPOS_GRCh37 | 17 | 78897056 | G | C | 0.000763458 | NO |
| PGS000364_hmPOS_GRCh37 | 17 | 78901443 | G | T | 0.000973817 | YES |
| PGS000364_hmPOS_GRCh37 | 17 | 78903681 | C | T | 0.000754083 | YES |
| PGS000364_hmPOS_GRCh37 | 17 | 78907809 | A | C | 0.000815714 | YES |
| PGS000364_hmPOS_GRCh37 | 17 | 78914245 | T | G | 0.001434127 | YES |
| PGS000364_hmPOS_GRCh37 | 17 | 78915387 | T | A | 0.003762539 | NO |
| PGS000364_hmPOS_GRCh37 | 17 | 78916379 | A | G | 0.00128652 | YES |
| PGS000364_hmPOS_GRCh37 | 17 | 78919162 | A | G | 0.000382464 | YES |
| PGS000364_hmPOS_GRCh37 | 18 | 1330559 | A | G | 0.007291319 | YES |
| PGS000364_hmPOS_GRCh37 | 18 | 1378362 | A | G | 0.006318356 | YES |
| PGS000364_hmPOS_GRCh37 | 18 | 9262101 | C | T | 0.001785656 | YES |
| PGS000364_hmPOS_GRCh37 | 18 | 9278178 | G | T | 0.001941084 | YES |
| PGS000364_hmPOS_GRCh37 | 18 | 9289676 | G | T | 0.000737118 | YES |
| PGS000364_hmPOS_GRCh37 | 18 | 9338344 | T | C | 0.001993433 | YES |
| PGS000364_hmPOS_GRCh37 | 18 | 9339276 | G | A | 0.002088517 | YES |
| PGS000364_hmPOS_GRCh37 | 18 | 9376108 | A | T | 0.003726923 | NO |
| PGS000364_hmPOS_GRCh37 | 18 | 9379877 | G | A | 0.002850622 | YES |
| PGS000364_hmPOS_GRCh37 | 18 | 9390227 | T | C | 0.003390706 | YES |
| PGS000364_hmPOS_GRCh37 | 18 | 9390878 | A | G | 0.003384866 | YES |
| PGS000364_hmPOS_GRCh37 | 18 | 9396597 | G | A | 0.003331276 | YES |
| PGS000364_hmPOS_GRCh37 | 18 | 21743391 | T | A | 0.001212498 | NO |
| PGS000364_hmPOS_GRCh37 | 18 | 34010470 | A | G | 0.000412753 | YES |
| PGS000364_hmPOS_GRCh37 | 18 | 57171281 | T | C | -0.000114562 | YES |
| PGS000364_hmPOS_GRCh37 | 18 | 57318902 | A | G | 0.005743808 | YES |
| PGS000364_hmPOS_GRCh37 | 18 | 63657268 | C | G | 0.001616723 | NO |
| PGS000364_hmPOS_GRCh37 | 18 | 63658175 | T | C | 0.001412633 | YES |
| PGS000364_hmPOS_GRCh37 | 18 | 66637488 | A | G | 0.009559737 | YES |
| PGS000364_hmPOS_GRCh37 | 18 | 67471154 | A | C | -0.001742552 | YES |
| PGS000364_hmPOS_GRCh37 | 18 | 67471483 | A | G | -0.014341836 | YES |
| PGS000364_hmPOS_GRCh37 | 18 | 71654492 | C | T | -0.005625996 | YES |
| PGS000364_hmPOS_GRCh37 | 19 | 3753874 | A | G | -0.001093354 | YES |
| PGS000364_hmPOS_GRCh37 | 19 | 3813013 | G | A | -0.000481795 | YES |
| PGS000364_hmPOS_GRCh37 | 19 | 3813684 | G | A | 0.008954201 | YES |
| PGS000364_hmPOS_GRCh37 | 19 | 3843896 | T | C | 0.000563974 | YES |
| PGS000364_hmPOS_GRCh37 | 19 | 6732852 | C | T | -0.01307661 | YES |
| PGS000364_hmPOS_GRCh37 | 19 | 6733545 | C | G | -0.013395931 | NO |
| PGS000364_hmPOS_GRCh37 | 19 | 6734712 | C | T | -0.012856357 | YES |
| PGS000364_hmPOS_GRCh37 | 19 | 6737644 | A | G | -0.009808265 | YES |
| PGS000364_hmPOS_GRCh37 | 19 | 11124470 | G | C | 0.00010834 | NO |
| PGS000364_hmPOS_GRCh37 | 19 | 11148934 | G | C | 0.000695539 | NO |
| PGS000364_hmPOS_GRCh37 | 19 | 11148948 | G | A | 0.000690626 | YES |
| PGS000364_hmPOS_GRCh37 | 19 | 11153543 | C | T | 0.002836705 | YES |
| PGS000364_hmPOS_GRCh37 | 19 | 11153574 | T | C | 0.002867483 | YES |
| PGS000364_hmPOS_GRCh37 | 19 | 12423818 | C | G | 7.65609920956589e-6 | NO |

| | | | | | | |
|---|---|---|---|---|---|---|
| PGS000364_hmPOS_GRCh37 | 19 | 22710737 | T | C | 0.001630508 | YES |
| PGS000364_hmPOS_GRCh37 | 19 | 29904906 | G | A | 0.005344496 | YES |
| PGS000364_hmPOS_GRCh37 | 19 | 32477283 | T | C | -0.00284337 | YES |
| PGS000364_hmPOS_GRCh37 | 19 | 32481907 | A | G | -0.001915735 | YES |
| PGS000364_hmPOS_GRCh37 | 19 | 32482727 | C | T | -0.002088053 | YES |
| PGS000364_hmPOS_GRCh37 | 19 | 32483917 | A | C | -0.002117038 | YES |
| PGS000364_hmPOS_GRCh37 | 19 | 32485263 | A | C | -0.002115336 | YES |
| PGS000364_hmPOS_GRCh37 | 19 | 32489168 | A | G | -0.002123407 | YES |
| PGS000364_hmPOS_GRCh37 | 19 | 32489272 | G | C | -0.002150055 | NO |
| PGS000364_hmPOS_GRCh37 | 19 | 32489923 | T | C | -0.002150618 | YES |
| PGS000364_hmPOS_GRCh37 | 19 | 32490507 | C | T | -0.002152072 | YES |
| PGS000364_hmPOS_GRCh37 | 19 | 32497463 | T | G | -0.002237191 | YES |
| PGS000364_hmPOS_GRCh37 | 19 | 34362298 | A | G | 0.015087517 | YES |
| PGS000364_hmPOS_GRCh37 | 19 | 34398808 | T | C | 0.011877746 | YES |
| PGS000364_hmPOS_GRCh37 | 19 | 34431555 | G | T | 0.012256034 | YES |
| PGS000364_hmPOS_GRCh37 | 19 | 34446444 | T | C | 0.008505154 | YES |
| PGS000364_hmPOS_GRCh37 | 19 | 44750883 | G | T | 0.015870341 | YES |
| PGS000364_hmPOS_GRCh37 | 19 | 44789598 | T | A | 0.015325737 | NO |
| PGS000364_hmPOS_GRCh37 | 19 | 44820995 | A | G | 0.013272492 | YES |
| PGS000364_hmPOS_GRCh37 | 19 | 48170545 | G | T | -0.001286459 | YES |
| PGS000364_hmPOS_GRCh37 | 19 | 48200803 | T | C | -0.001397768 | YES |
| PGS000364_hmPOS_GRCh37 | 19 | 48201652 | C | G | -0.001097385 | NO |
| PGS000364_hmPOS_GRCh37 | 19 | 51250181 | C | G | -0.002243428 | NO |
| PGS000364_hmPOS_GRCh37 | 19 | 51420756 | T | G | 0.006278393 | YES |
| PGS000364_hmPOS_GRCh37 | 19 | 51588884 | C | T | 0.001419665 | YES |
| PGS000364_hmPOS_GRCh37 | 19 | 51590173 | T | C | 0.005042874 | YES |
| PGS000364_hmPOS_GRCh37 | 19 | 53124451 | A | G | 0.001999831 | YES |
| PGS000364_hmPOS_GRCh37 | 19 | 53153196 | A | C | 0.009408568 | YES |
| PGS000364_hmPOS_GRCh37 | 20 | 1997672 | C | T | 0.000352152 | YES |
| PGS000364_hmPOS_GRCh37 | 20 | 2000819 | C | T | 0.00087301 | YES |
| PGS000364_hmPOS_GRCh37 | 20 | 2001015 | G | A | 0.000265127 | YES |
| PGS000364_hmPOS_GRCh37 | 20 | 6727734 | G | C | -0.001083611 | NO |
| PGS000364_hmPOS_GRCh37 | 20 | 6728586 | C | T | 0.006199537 | YES |
| PGS000364_hmPOS_GRCh37 | 20 | 6728704 | G | T | 0.006328897 | YES |
| PGS000364_hmPOS_GRCh37 | 20 | 6728762 | C | T | 0.006491301 | YES |
| PGS000364_hmPOS_GRCh37 | 20 | 6728807 | A | G | 0.005306528 | YES |
| PGS000364_hmPOS_GRCh37 | 20 | 6728966 | T | G | 0.002314402 | YES |
| PGS000364_hmPOS_GRCh37 | 20 | 6729461 | C | T | 0.001854681 | YES |
| PGS000364_hmPOS_GRCh37 | 20 | 6729499 | A | G | 0.001813161 | YES |
| PGS000364_hmPOS_GRCh37 | 20 | 6729560 | C | T | 0.001771664 | YES |
| PGS000364_hmPOS_GRCh37 | 20 | 6729571 | A | G | 0.001771356 | YES |
| PGS000364_hmPOS_GRCh37 | 20 | 6729734 | G | C | 0.001525671 | NO |
| PGS000364_hmPOS_GRCh37 | 20 | 6729789 | C | T | 0.001532815 | YES |
| PGS000364_hmPOS_GRCh37 | 20 | 6730079 | T | C | 0.000872157 | YES |
| PGS000364_hmPOS_GRCh37 | 20 | 6730405 | A | G | 0.000859983 | YES |
| PGS000364_hmPOS_GRCh37 | 20 | 6730911 | T | A | 0.000702243 | NO |
| PGS000364_hmPOS_GRCh37 | 20 | 18096932 | A | G | 0.002051616 | YES |
| PGS000364_hmPOS_GRCh37 | 20 | 18111708 | C | T | 0.003300859 | YES |
| PGS000364_hmPOS_GRCh37 | 20 | 20593272 | T | G | 0.000460807 | YES |
| PGS000364_hmPOS_GRCh37 | 20 | 24878525 | T | C | 0.017264221 | NO |
| PGS000364_hmPOS_GRCh37 | 20 | 24892552 | A | G | 0.006817007 | YES |
| PGS000364_hmPOS_GRCh37 | 20 | 24893707 | G | A | 0.005649159 | YES |
| PGS000364_hmPOS_GRCh37 | 20 | 24894067 | T | C | 0.005656302 | YES |
| PGS000364_hmPOS_GRCh37 | 20 | 24894448 | G | C | 0.00566937 | NO |
| PGS000364_hmPOS_GRCh37 | 20 | 24896365 | A | G | 0.00696608 | YES |
| PGS000364_hmPOS_GRCh37 | 20 | 24897406 | C | T | 0.004323774 | YES |
| PGS000364_hmPOS_GRCh37 | 20 | 44223600 | C | G | 0.007082033 | NO |
| PGS000364_hmPOS_GRCh37 | 20 | 44224978 | A | G | 0.00832963 | YES |
| PGS000364_hmPOS_GRCh37 | 20 | 44231598 | T | G | 0.008114658 | YES |
| PGS000364_hmPOS_GRCh37 | 20 | 44232288 | T | C | 0.008114784 | YES |
| PGS000364_hmPOS_GRCh37 | 20 | 44234781 | T | G | 0.00811491 | YES |
| PGS000364_hmPOS_GRCh37 | 20 | 44235043 | T | C | 0.008062202 | YES |
| PGS000364_hmPOS_GRCh37 | 20 | 44235865 | G | T | 0.008067378 | YES |

| | | | | | | |
|---|---|---|---|---|---|---|
| PGS000364_hmPOS_GRCh37 | 20 | 44238429 | T | G | 0.008044962 | YES |
| PGS000364_hmPOS_GRCh37 | 20 | 44238741 | G | T | 0.00480752 | YES |
| PGS000364_hmPOS_GRCh37 | 20 | 44242565 | G | T | 0.007854946 | YES |
| PGS000364_hmPOS_GRCh37 | 20 | 44242581 | C | T | 0.007855174 | YES |
| PGS000364_hmPOS_GRCh37 | 20 | 44242789 | C | T | 0.004981452 | YES |
| PGS000364_hmPOS_GRCh37 | 20 | 44243429 | A | G | 0.008035931 | YES |
| PGS000364_hmPOS_GRCh37 | 20 | 44244511 | C | T | 0.008059091 | YES |
| PGS000364_hmPOS_GRCh37 | 20 | 44245927 | G | C | 0.008059249 | NO |
| PGS000364_hmPOS_GRCh37 | 20 | 44247546 | G | A | 0.006953514 | YES |
| PGS000364_hmPOS_GRCh37 | 20 | 44247693 | G | T | 0.003979487 | YES |
| PGS000364_hmPOS_GRCh37 | 20 | 44248435 | A | G | 0.006872379 | YES |
| PGS000364_hmPOS_GRCh37 | 20 | 44248596 | T | C | 0.006813593 | YES |
| PGS000364_hmPOS_GRCh37 | 20 | 44248711 | A | G | 0.006840063 | YES |
| PGS000364_hmPOS_GRCh37 | 20 | 44249253 | A | T | 0.006822946 | NO |
| PGS000364_hmPOS_GRCh37 | 20 | 44250199 | G | A | 0.00678427 | YES |
| PGS000364_hmPOS_GRCh37 | 20 | 44251629 | A | G | 0.006863902 | YES |
| PGS000364_hmPOS_GRCh37 | 20 | 44253195 | A | C | 0.006885423 | YES |
| PGS000364_hmPOS_GRCh37 | 20 | 44256591 | C | T | 0.003905448 | YES |
| PGS000364_hmPOS_GRCh37 | 20 | 44257650 | C | T | 0.004699073 | YES |
| PGS000364_hmPOS_GRCh37 | 20 | 44257652 | C | T | 0.004639149 | YES |
| PGS000364_hmPOS_GRCh37 | 20 | 44258743 | G | A | 0.003896516 | YES |
| PGS000364_hmPOS_GRCh37 | 20 | 44258932 | C | T | 0.003896547 | YES |
| PGS000364_hmPOS_GRCh37 | 20 | 44259490 | A | G | 0.003910094 | YES |
| PGS000364_hmPOS_GRCh37 | 20 | 44259673 | T | G | 0.006790646 | YES |
| PGS000364_hmPOS_GRCh37 | 20 | 44261397 | T | C | 0.003916072 | YES |
| PGS000364_hmPOS_GRCh37 | 20 | 44261643 | T | C | 0.003917591 | YES |
| PGS000364_hmPOS_GRCh37 | 20 | 44262911 | C | A | 0.006805043 | YES |
| PGS000364_hmPOS_GRCh37 | 20 | 44268065 | G | A | 0.003700878 | YES |
| PGS000364_hmPOS_GRCh37 | 20 | 44268779 | C | G | 0.006648614 | NO |
| PGS000364_hmPOS_GRCh37 | 20 | 44269042 | G | A | 0.003712633 | YES |
| PGS000364_hmPOS_GRCh37 | 20 | 44269982 | C | T | 0.003746501 | YES |
| PGS000364_hmPOS_GRCh37 | 20 | 44270415 | G | A | 0.003773862 | YES |
| PGS000364_hmPOS_GRCh37 | 20 | 44271254 | A | G | 0.003741217 | YES |
| PGS000364_hmPOS_GRCh37 | 20 | 44272936 | A | C | 0.003993645 | YES |
| PGS000364_hmPOS_GRCh37 | 20 | 44272944 | T | C | 0.004009091 | YES |
| PGS000364_hmPOS_GRCh37 | 20 | 44277111 | A | G | 0.003799149 | YES |
| PGS000364_hmPOS_GRCh37 | 20 | 44278346 | C | T | 0.006427847 | YES |
| PGS000364_hmPOS_GRCh37 | 20 | 44279768 | G | A | 0.007119451 | YES |
| PGS000364_hmPOS_GRCh37 | 20 | 44288068 | C | T | 0.007116125 | YES |
| PGS000364_hmPOS_GRCh37 | 20 | 44311598 | G | A | 0.00705187 | YES |
| PGS000364_hmPOS_GRCh37 | 20 | 44312991 | T | C | 0.007027614 | YES |
| PGS000364_hmPOS_GRCh37 | 20 | 44322500 | T | C | 0.007346221 | YES |
| PGS000364_hmPOS_GRCh37 | 20 | 44324215 | G | A | 0.007107301 | YES |
| PGS000364_hmPOS_GRCh37 | 20 | 44326104 | G | A | 0.007143468 | YES |
| PGS000364_hmPOS_GRCh37 | 20 | 44327802 | A | G | 0.002645131 | YES |
| PGS000364_hmPOS_GRCh37 | 20 | 44330128 | T | C | 0.002754404 | YES |
| PGS000364_hmPOS_GRCh37 | 20 | 54547988 | T | C | 0.002778417 | YES |
| PGS000364_hmPOS_GRCh37 | 21 | 19772457 | G | A | 0.002866212 | YES |
| PGS000364_hmPOS_GRCh37 | 21 | 27414936 | A | G | -0.000345527 | YES |
| PGS000364_hmPOS_GRCh37 | 21 | 27417001 | C | A | -0.000305238 | YES |
| PGS000364_hmPOS_GRCh37 | 21 | 27422395 | T | A | -0.002106274 | NO |
| PGS000364_hmPOS_GRCh37 | 21 | 27425859 | C | G | -0.004306725 | NO |
| PGS000364_hmPOS_GRCh37 | 21 | 32217621 | C | T | 0.000801404 | YES |
| PGS000364_hmPOS_GRCh37 | 21 | 32219380 | C | T | 0.000362774 | YES |
| PGS000364_hmPOS_GRCh37 | 21 | 32219570 | G | A | 0.000364953 | YES |
| PGS000364_hmPOS_GRCh37 | 21 | 32219923 | C | T | 0.000239045 | YES |
| PGS000364_hmPOS_GRCh37 | 21 | 32221534 | A | T | 0.000313859 | NO |
| PGS000364_hmPOS_GRCh37 | 21 | 32223082 | C | T | 0.000241642 | YES |
| PGS000364_hmPOS_GRCh37 | 21 | 32225713 | T | A | 0.000308485 | NO |
| PGS000364_hmPOS_GRCh37 | 21 | 32225824 | C | A | 0.000224154 | YES |
| PGS000364_hmPOS_GRCh37 | 21 | 32227016 | C | G | 0.00030567 | NO |
| PGS000364_hmPOS_GRCh37 | 21 | 32228253 | A | G | 0.000342035 | YES |
| PGS000364_hmPOS_GRCh37 | 21 | 32228556 | A | G | 0.000311819 | YES |

| | | | | | | |
|---|---|---|---|---|---|---|
| PGS000364_hmPOS_GRCh37 | 21 | 32229736 | T | G | 0.000293355 | YES |
| PGS000364_hmPOS_GRCh37 | 21 | 32233838 | T | A | 0.000487662 | NO |
| PGS000364_hmPOS_GRCh37 | 21 | 32235779 | T | C | 0.000570481 | YES |
| PGS000364_hmPOS_GRCh37 | 21 | 32237062 | T | C | 0.000545466 | YES |
| PGS000364_hmPOS_GRCh37 | 21 | 32237708 | T | C | 0.000491435 | YES |
| PGS000364_hmPOS_GRCh37 | 21 | 32237833 | A | T | 0.000567019 | NO |
| PGS000364_hmPOS_GRCh37 | 21 | 32239665 | C | A | 0.002027585 | YES |
| PGS000364_hmPOS_GRCh37 | 21 | 32242221 | A | C | 0.000392369 | YES |
| PGS000364_hmPOS_GRCh37 | 21 | 32242431 | T | A | 0.00025601 | NO |
| PGS000364_hmPOS_GRCh37 | 21 | 32242774 | G | A | 0.000409485 | YES |
| PGS000364_hmPOS_GRCh37 | 21 | 32243319 | C | T | 0.000268644 | YES |
| PGS000364_hmPOS_GRCh37 | 21 | 32243997 | A | G | 0.000487342 | YES |
| PGS000364_hmPOS_GRCh37 | 21 | 32244060 | A | T | 0.000502017 | NO |
| PGS000364_hmPOS_GRCh37 | 21 | 32245247 | T | C | 0.000762118 | YES |
| PGS000364_hmPOS_GRCh37 | 21 | 32245923 | C | T | 0.000579143 | YES |
| PGS000364_hmPOS_GRCh37 | 21 | 32246233 | C | G | 0.000473927 | NO |
| PGS000364_hmPOS_GRCh37 | 21 | 32246912 | C | T | 0.000589246 | YES |
| PGS000364_hmPOS_GRCh37 | 21 | 32248452 | A | G | 0.001058911 | YES |
| PGS000364_hmPOS_GRCh37 | 21 | 32248498 | G | A | 0.000852894 | YES |
| PGS000364_hmPOS_GRCh37 | 21 | 32250952 | C | T | 0.000886218 | YES |
| PGS000364_hmPOS_GRCh37 | 21 | 32252110 | A | T | 0.001007583 | NO |
| PGS000364_hmPOS_GRCh37 | 21 | 32252137 | T | G | 0.001275042 | YES |
| PGS000364_hmPOS_GRCh37 | 21 | 32252565 | T | G | 0.001166356 | YES |
| PGS000364_hmPOS_GRCh37 | 21 | 32253629 | T | C | 0.001884524 | YES |
| PGS000364_hmPOS_GRCh37 | 21 | 32253966 | C | T | 0.001434148 | YES |
| PGS000364_hmPOS_GRCh37 | 21 | 35042415 | C | A | -0.00151282 | YES |
| PGS000364_hmPOS_GRCh37 | 21 | 37601762 | G | C | 0.003605164 | NO |
| PGS000364_hmPOS_GRCh37 | 21 | 39955226 | A | C | 0.001809284 | YES |
| PGS000364_hmPOS_GRCh37 | 21 | 40318829 | A | G | 0.009711648 | YES |
| PGS000364_hmPOS_GRCh37 | 21 | 40324223 | C | T | -0.001837455 | YES |
| PGS000364_hmPOS_GRCh37 | 21 | 40325715 | A | T | -0.004114253 | NO |
| PGS000364_hmPOS_GRCh37 | 21 | 40328320 | G | T | -0.001677184 | YES |
| PGS000364_hmPOS_GRCh37 | 21 | 40330208 | T | C | -0.006960639 | YES |
| PGS000364_hmPOS_GRCh37 | 21 | 42027045 | T | C | 0.0057356 | YES |
| PGS000364_hmPOS_GRCh37 | 21 | 47973610 | A | G | 0.005652494 | YES |
| PGS000364_hmPOS_GRCh37 | 21 | 47974388 | G | C | 0.006734096 | NO |
| PGS000364_hmPOS_GRCh37 | 21 | 47980913 | T | G | -0.005407249 | YES |
| PGS000364_hmPOS_GRCh37 | 21 | 47980948 | C | G | 0.009179307 | NO |
| PGS000364_hmPOS_GRCh37 | 21 | 47982040 | T | C | 0.007310141 | YES |
| PGS000364_hmPOS_GRCh37 | 21 | 47982363 | A | G | 6.61274E-05 | YES |
| PGS000364_hmPOS_GRCh37 | 21 | 47982738 | A | G | 0.005016136 | YES |
| PGS000364_hmPOS_GRCh37 | 21 | 47983543 | T | C | 0.006936028 | YES |
| PGS000364_hmPOS_GRCh37 | 21 | 47988364 | T | C | 0.007507896 | YES |
| PGS000364_hmPOS_GRCh37 | 22 | 17675324 | T | C | -0.013531696 | YES |
| PGS000364_hmPOS_GRCh37 | 22 | 25217890 | T | G | 0.004517201 | YES |
| PGS000364_hmPOS_GRCh37 | 22 | 27297757 | T | C | 0.00093769 | YES |
| PGS000364_hmPOS_GRCh37 | 22 | 36065885 | A | G | 0.004887987 | YES |
| PGS000364_hmPOS_GRCh37 | 22 | 39575221 | G | A | -0.008383493 | YES |
| PGS000364_hmPOS_GRCh37 | 22 | 39629609 | A | G | -0.005544208 | YES |
| PGS000364_hmPOS_GRCh37 | 22 | 47221990 | G | T | 0.004763435 | YES |
| PGS000364_hmPOS_GRCh37 | 22 | 49599953 | T | G | 0.003587219 | YES |

Abbreviations: SNP, Single nucleotide polymorphism; PRS, polygenic risk score.

Total number of variants: 2001
Matched, n (%): 1682 (84.06%)
Exclusions due to ambiguously matched, n (%): 312 (15.59%)
Exclusions due to unmatched, n (%): 7 (0.35%)
